# Biallelic variants in *ARHGAP19* cause a motor-predominant neuropathy with asymmetry and conduction slowing

**DOI:** 10.1101/2024.05.10.24306768

**Authors:** Natalia Dominik, Stephanie Efthymiou, Christopher J. Record, Xinyu Miao, Renee Lin, Jevin Parmar, Annarita Scardamaglia, Reza Maroofian, Gabriel Aughey, Abigail Wilson, Simon Lowe, Riccardo Curro, Ricardo P. Schnekenberg, Shahryar Alavi, Leif Leclaire, Yi He, Kristina Zhelchenska, Yohanns Bellaiche, Isabelle Gaugué, Mariola Skorupinska, Liedewei Van de Vondel, Sahar I. Da’as, Valentina Turchetti, Serdal Güngör, Ehsan Ghayoor Karimiani, Camila Armirola Ricaurte, Haluk Topaloglu, Albena Jordanova, Mashaya Zaman, Selina H. Banu, Wilson Marques, Pedro José Tomaselli, Busra Aynekin, Ali Cansu, Huseyin Per, Ayten Güleç, Javeria Raza Alvi, Tipu Sultan, Arif Khan, Giovanni Zifarelli, Shahnaz Ibrahim, Grazia M.S. Mancini, M. Mahdi Motazacker, Esther Brusse, Vincenzo Lupo, Teresa Sevilla, A Nazlı Başak, Seyma Tekgul, Robin Palvadeau, Jonathan Baets, Yesim Parman, Arman Çakar, Rita Horvath, Tobias B. Haack, Jan-Hendrik Stahl, Kathrin Grundmann-Hauser, Joohyun Park, Stephan Züchner, Nigel G. Laing, Lindsay Wilson, Alexander M. Rossor, James Polke, Fernanda Barbosa Figueiredo, André Luiz Pessoa, Fernando Kok, Antônio Rodrigues Coimbra-Neto, Marcondes C. França, Yalda Jamshidi, Gianina Ravenscroft, Sherifa Ahmed Hamed, Wendy K. Chung, Daniel P. Osborn, Michael Hanna, Andrea Cortese, Mary M. Reilly, James E. C. Jepson, Nathalie Lamarche-Vane, Henry Houlden

**Author notes:** Correspondence to Dr. Stephanie Efthymiou, Department of Neuromuscular Disorders, UCL Queen Square Institute of Neurology, UCL, London, WC1N 3BG, UK. Contributed equally (Natalia Dominik, Stephanie Efthymiou, Mary M. Reilly, James E. C. Jepson, Nathalie Lamarche-Vane, Henry Houlden).

## Abstract

Charcot-Marie-Tooth Disease is a clinically and genetically heterogeneous group of hereditary neuropathies, with over 100 causative genes identified to date. Despite progress in genetic sequencing, around a quarter of patients remain unsolved. Through international collaborations, we identified 16 recessive variants in Rho GTPase activating protein 19 (*ARHGAP19*) causing motor-predominant neuropathy with conduction slowing in 25 individuals from 20 unrelated multi-ancestry families. ARHGAP19 is a GTPase-activating protein with activity towards RhoA. *In vitro* biochemical assays revealed that variants located within the GAP domain cause loss of GAP activity. iPSc-derived motor neurons exhibited 50% knockdown of ARHGAP19 protein. *In vivo* genetic perturbations of the *Drosophila melanogaster ARHGAP19* ortholog *RhoGAP54D* reduced self-driven locomotor activity and startle responses to visual stimuli. Zebrafish loss-of-function models similarly exhibited movement deficits, coupled with increased motor neuron axonal branching but shorter caudal primary motor neurons. Together, these findings establish *ARHGAP19* as a novel cause of early-onset neuropathy through a loss-of-function mechanism.

## Introduction

Charcot-Marie-Tooth disease (CMT), also called hereditary motor and sensory neuropathy (HMSN) is the most prevalent Mendelian inherited neuropathy. Notably, inherited peripheral neuropathies are amongst the most frequently inherited neurologic diseases. The prevalence of CMT varies amongst populations but is estimated at around 1 in 2,500 individuals (1). Patients with CMT can range from mildly affected to severely disabled and the disease presents with progressive weakness and atrophy of muscles, especially in distal limbs. Often foot abnormalities such as *pes cavus* or hammer toes may be associated with the disease. Symptoms of CMT overlap between neuropathies and subtypes of CMT. Despite many genes being associated to inherited CMT, there remains a large proportion of genetically unexplained cases. However, family history, nerve conduction studies and thorough clinical evaluation can aid differential diagnosis (2).

The Rho family of small GTPases is composed of 20 members that include RhoA, Rac1, and CDC42. Rho GTPases act as molecular switches by cycling between inactive guanine nucleotide diphosphate (GDP) bound state and active, triphosphate (GTP) bound state and are involved in signalling pathways that control actin cytoskeleton reorganisation, cell adhesion, migration, and cell division. The activity of Rho GTPases is tightly regulated by three classes of proteins, Guanine nucleotide Exchange Factors (GEFs) which facilitate the exchange from GDP to GTP; GTPase-Activating Proteins (GAPs) which stimulate the intrinsic GTPase activity, resulting in hydrolysis of GTP and protein inactivation; and Guanine Nucleotide Dissociation Inhibitors (GDIs) which sequester and maintain Rho GTPases inactive in the cytoplasm(3) (4). In humans, over 66 RhoGAPs and 80 RhoGEFs have been identified for 20 members of Rho family that they can act on, and many are implicated in neurological diseases, including CMT (4). For example, *PLEKHG5*, a GEF gene that regulates autophagy of synaptic vesicles in axonal terminals, is associated with CMT (5). Furthermore, variants in *MYO9B*, a RhoGAP gene, have recently been shown to cause CMT type 2 (CMT2) and optic atrophy. Whether CMT can be caused by mutations in other regulators of Rho GTPases, however, remains an open question.

Here, we report the clinical phenotypes associated with biallelic, autosomal recessive variants in *ARHGAP19* in 25 individuals with CMT from 20 families. ARHGAP19 is a small, 494 amino acid (AA) RhoGAP protein, displaying GAP activity towards RhoA but not Rac1 and Cdc42, thus acting as a negative regulator of the RhoA/ Rho-associated kinase (ROCK) pathway. ARHGAP19 has previously been shown to have an essential role in T-lymphocyte cytokinesis and phosphorylation by kinases such as ROCK at Ser422 and CDK1 at Thr404 and Thr476 is essential for its role in the control of cell division (3, 6). We provide mechanistic evidence for the contribution of variants to CMT suggesting a loss of function (LOF) mechanism through the use of individual patient cell lines, *in vitro* GAP assay, *in vivo Drosophila melanogaster* and *Danio rerio* knockdown models as well as *in-silico* molecular modelling. Both *Drosophila* and zebrafish LOF models exhibit significant motor defects, while zebrafish model indicates an impact of ARHGAP19 LOF on motoneuron length and branching.

## Methods

### Study Participants

Individuals were recruited via an international collaborative network of research and diagnostic sequencing laboratories. Samples and clinical information were obtained, with informed consent, from each institution using local institutional review board (IRB) ethics for functional analysis of human DNA, fibroblasts and biomaterial. Clinical data collection involved a detailed review of medical records, photographs, videos, and phone interviews, as well as a clinical re-evaluation of nerve conduction studies by a neurologist. GeneMatcher and RD-Connect (GPAP) platform facilitated the identification of additional patients. Table 1 and Supplementary table 1 summarize the clinical details of the included cases. Study subject IDs used are for the purpose of this study and are not known to anyone outside the research group.

**Table 1.** Clinical features of *ARHGAP19* individuals. MP motor predominant * neurophysiology data not seen +/− report available only ** very limited study, Clinical sensory involvement refers to symptoms and/or signs. Δ assessed approximately 2 years aBer onset of acute leB hand weakness. CS conducFon slowing LL lower limb UL upper limb KJ knee jerk, AJ ankle jerk, SG increased sandal gap, MRC grade ADF/APF = medical research council power grading of power in ankle dorsiflexion/ankle plantar flexion. ∼= approximate (where known). If there is discrepancy between limbs, a comma separates right and leB. > greater than, LD length-dependent, M male, F female, Unk unknown, dist distal, prox proximal.

| Family | 1 | 2 | 3 | 4 | 5 | 6 | 7 | 8 | 9 | 10 | 11 | 12 | 13 | 14 | 15 | 16 | 17 | 18 | 19 | 20 |
| --- | --- | --- | --- | --- | --- | --- | --- | --- | --- | --- | --- | --- | --- | --- | --- | --- | --- | --- | --- | --- |
| Case | PT1<br>(F1-II:1) | PT2<br>(F2-II:5) | PT3<br>(F3-II:1) | PT4<br>(F4-II:1) | PT5<br>(F5-II:2) | PT6<br>(F6-II:7) | PT7<br>(F7-II:1) | PT8<br>(F8-II:2) | PT9<br>(F9-II:3) | PT11<br>(F10-II:1) | PT12<br>(F11-II:2) | PT13<br>(F12-II:1) | PT14<br>(F13-II:3) | PT15<br>(F14-II:2) | PT17<br>(F15-II:2) | PT18<br>(F16-II:1) | PT19<br>(F17-II:1) | PT20<br>(F18-II:1) | PT21<br>(F19-II:1) | PT25<br>(F21-IV:1) |
| Nucleotide<br>change | c.261d<br>up | c.585d<br>upA | c.1243<br>C>T | c.1219<br>C>T | c.419G<br>>A | c.203T<br>>C | c.451C<br>>A | c.1A>G | c.452d<br>el | c.85A><br>G | c.932C<br>>G | c.683T<br>>A | c.585d<br>upA | c.451C<br>>A | c.203T<br>>C | c.422T<br>>G | c.717T<br>>A | c.563d<br>el | c.683T<br>>A | c.818C>T |
| Amino acid<br>change | p.Pro8<br>8Alafs*<br>43 | p.His19<br>6Glnfs*<br>9 | p.Gln4<br>15* | p.Arg4<br>07* | p.Gly14<br>0Asp | p.Leu6<br>8Pro | p.Gln1<br>51Lys | p.Met1<br>? | p.Asn1<br>60Metf<br>s*21 | p.Asn2<br>9Asp | p.Pro3<br>11Arg | p.Leu2<br>28His | p.His19<br>6Glnfs*<br>9 | p.Gln1<br>51Lys | p.Leu6<br>8Pro | p.Leu1<br>41Trp | p.Asn2<br>39Lys | p.Pro1<br>88Argfs<br>*5 | p.Leu2<br>28His | p.Pro273<br>Leu |
| Sex | M | M | F | F | M | F | F | F | M | M | M | M | F | M | F | F | F | F | M | F |
| Phenotype | CMT-MP** | CMT<br>with<br>CS* | dHMN<br>with CS | CMT2-MP | dHMN | dHMN<br>with CS | CMT<br>with<br>CS* | CMT2-MP | CMT2w<br>ith<br>CS** | CMT2-MP | CMT2-MP<br>with CS | Unk* | CMT** | dHMN<br>* | CMTi-MP | dHMN<br>** | CMT<br>with<br>CS* | HMN<br>with<br>CS** | CMT2-MP | dHMN* |
| Consangui<br>nity | Yes | No | Yes | No | No | Yes | No | Yes | Yes | Yes | Yes | Yes | Yes | Yes | No | No | Yes | Yes | Yes | Yes |
| Age of<br>onset<br>(years) | 6-10 | 11-15 | 6-10 | 6-10 | 10-20 | infancy | 11-15 | 11-15 | 11-15 | 26-30 | 11-15 | 0-5 | n/a | 11-15 | 11-15<sup>Δ</sup> | 11-15 | 6-10 | 6-10 | 11-15 | 0-5 |
| Symptom<br>at onset | Falls | Walkin<br>g<br>difficult<br>y | Falls | Dist LL<br>weakn<br>ess | Dist LL<br>weakn<br>ess | Walkin<br>g<br>difficult<br>y | Dist LL<br>weakn<br>ess | Walkin<br>g<br>difficult<br>y | Dist<br>right LL<br>weakn<br>ess | Leg<br>cramps | Dist LL<br>weakn<br>ess | Acute<br>UL<br>weakn<br>ess | Unk | Falls | Acute<br>left<br>hand<br>weakn<br>ess | Dist LL<br>weakn<br>ess | Right<br>leg<br>weakn<br>ess | Dist<br>right LL<br>weakn<br>ess | Dist left<br>LL<br>weakn<br>ess | Dist LL<br>weaknes<br>s |
| Asymmetr<br>y | Yes | Yes | Yes | No | No | Yes | No | Yes | Yes | No | No | No | Unk | No | Yes | Yes | Yes | Yes | Yes | Yes |
| Clinical<br>sensory<br>involveme<br>nt | No | Yes | No | Yes | No | Yes | No | Yes | Yes | Yes | Yes | Yes | Unk | No | Yes | Yes | Unk | No | No | Yes |
| LL<br>Areflexia | No | Brisk<br>KJ, AJ | Yes | Yes | Yes | KJ<br>present | Yes | KJ<br>present | Yes | KJ<br>brisk,<br>AJ<br>present<br>, L<br>Babins<br>ki | KJ<br>present | Unk | Unk | Yes | Brisk<br>KJ, AJ<br>present | Yes | Left AJ<br>present | Yes | No | KJ<br>present |
| **Pattern of muscle weakness( MRC grade ADF/APF)** | Dist LL L>R (4,3/4, 4) | Distal LL L>R (1,2) | LD (~3,0/4 ) | LD ('severe') | LL predominant dist > prox (0/0) | LD (~2,3/4 ) | LD (4/4) | LD (0/4) | LD (~1,3/5 ) | LL predominant dist > prox (4/5) | LD (0/3) | UL > LL (Unk) | Unk | ?LD (Unk) | Dist UL> LL (5/5) | LD (0,1/1) | Right dist LL only (Unk) | LL predominant dist > prox (~1/?) | Dist LL L>R, (4,0/5) | LD (2/4) |
| **Foot deformity** | Pes cavus | Pes planus, hammer toe | Pes cavus | Pes cavus | Nil | Pes cavus, SG | Pes cavus | Pes planus | Pes planus | Nil | Pes cavus | Nil | Unk | Pes cavus | Hammer toe, SG | Pes cavus | Unk | Unk | No | Yes |
| **Gait** | Left foot drop | Unk | Steppage | Unk | Steppage | Steppage | Steppage | Steppage | Steppage | Steppage and waddle | Steppage | Unclear | Unk | Steppage | Normal | Steppage | Unk | Steppage | Steppage | Steppage |
| **Muscle atrophy** | Dist LL L>R | Distal LL (calf hypertrophy) | Calves down | Dist LL | Calves down | Feet | Feet | Calves down | None | Calves down | Dist LL | Unk | Unk | Dist LL | Hands and dist LL | Dist UL and LL | Unk | Dist LL R>L | Dist LL | Dist LL > UL |
| **Neurophysiology** | Limited study: LL motor only. Asymmetric LD motor neuropathy | *Axonal and demyelinating neuropathy (mainly axonal) bilaterally, L>R | LD motor neuropathy with significant conduction slowing and block | LD MP axonal neuropathy | MP LL predominant axonal neuropathy | MP LL predominant axonal neuropathy with some conduction slowing | *Mixed axonal and demyelinating neuropathy (mainly axonal) UL and LL | LD MP axonal neuropathy with upper limb asymmetry | Very limited study; LD sensory and motor neuropathy | LD axonal neuropathy, some acute EMG changes | LD MP axonal neuropathy with conduction slowing | Not performed | Very limited only single nerve studied | *motor axonal neuropathy | LD MP demyelinating neuropathy | Limited study; MPLD neuropathy | *Mixed motor> sensory neuropathy (axonal >demyelinating) involving both LL, R>L | **Motor neuropathy with conduction slowing | LD MP axonal neuropathy with minimal conduction slowing | *Axonal motor neuropathy |

### Next generation sequencing

Genomic DNA was extracted from peripheral blood samples of subjects and parents according to standard procedures. Exome sequencing on DNA of subjects P2, P3, P12, P15, P18 was performed as described elsewhere (7) in Macrogen, Korea. Briefly, target enrichment was performed with 2 μg genomic DNA using the SureSelectXT Human All Exon Kit version 6 (Agilent) to generate barcoded whole-exome sequencing libraries. Libraries were sequenced on the HiSeqX platform (Illumina) with 50x coverage. Quality assessment of the sequence reads was performed by generating QC statistics with FastQC. The bioinformatics filtering strategy included screening for only exonic and donor/acceptor splicing variants. In accordance with the pedigree and phenotype, priority was given to rare variants (<0.01% in public databases, including 1000 Genomes project, NHLBI Exome Variant Server, Complete Genomics 69, and Exome Aggregation Consortium [ExAC v0.2]) that were fitting a recessive (homozygous or compound heterozygous) or a *de novo* model and/or variants in genes previously linked to neuropathy, developmental delay, and other neurological disorders. Next generation sequencing for other samples was carried out using a number of methods with different analysis platforms and pipelines used depending on the centre where sequencing was performed (Supplementary Table 2). Recommendations of the Human Genome Variation Society were used to describe the cDNA and protein sequence variants using NM_032900.6 and NP_116289.4 as the reference. All the candidate variants were further verified, and family segregation was performed via Sanger sequencing in-house or in the referring centres. Homozygosity mapping was performed on Automap(8) and haplotype analysis was performed by plotting a colour banding of the variants flanking the pathogenic ARHGAP19 variants for affected individuals and control samples. By comparing the banding patterns of patients and controls, the status of being a founder/recurrent variant was investigated. In the case of founder variants, we estimated the age of the most recent common ancestor (MRCA) using the length of shared haplotypes between patients(9).

### Multiple sequence alignment

To examine the conservation of substituted amino acid positions, we performed multiple sequence alignments of ARHGAP19 across multiple different species. ARHGAP19 protein sequences for each species were retrieved from UniProt using their respective accession codes (Supplementary Table 3). Alignments were performed using the MAFFT algorithm in Jalview (v2.11.2).

### Protein structure modelling and in silico mutagenesis

The predicted wild-type ARHGAP19 protein structure was retrieved from the AlphaFold Protein Structure Database (10) using its UniProt accession code (Q14CB8). Three-dimensional protein structures were visualised using PyMol (v.2.5.2). To examine variant effects on three-dimensional protein structure, *in silico* mutagenesis for identified missense substitutions was performed through the PyMol ‘mutagenesis’ function. For nonsense and frameshift variants, mutant protein structures were generated using the open-source AlphaFold v2.0 (AlphaFold2) pipeline (10) with the input being FASTA files of the mutant amino acid sequences. The resultant protein structures were then aligned to the wild-type ARHGAP19 protein structure within PyMol. AlphaMissense substitution scores for identified *ARHGAP19* missense variants were downloaded and extracted from resources generated by Cheng *et al* (11).

### Calculation of free energy changes

To investigate the impact of *ARHGAP19* variants on protein stability, free energy changes between the wild-type and mutant proteins were calculated using FoldX v5.0.(12) Free energy change predictions were only performed on missense substitutions within high confidence regions (pLDDT > 90) of the AlphaFold2-predicted protein structure.(13) The AlphaFold2 derived wild-type ARHGAP19 protein structure was energy minimised and prepared for mutagenesis with the ‘RepairPDB’ and ‘BuildModel’ commands in FoldX, respectively. Subsequent free energy calculations were performed with the ‘Stability’ command, and differences in free energy were determined by subtracting the free energy value of the wild-type protein monomer from that of the mutant protein (ΔG_MUT_ - ΔG_WT_). Calculations were replicated 10 times with default parameters for each substitution. Substitutions were classified as having a destabilising effect on protein folding if the free energy difference (ΔΔG) was >1.6 kcal/mol (12, 14, 15).

### Cell culture of primary dermal fibroblasts

Primary dermal fibroblasts were obtained from skin biopsies of subjects in family 5 (P5 (F5-II:2) (c.419G>A), family 6 (P6 (F6-II:7) (c.203T>C) and family 10 (P11 (F10-II:1) (c.85A>G) and age/ gender matched control individuals. Fibroblasts were cultured in Dulbecco’s modified Eagle medium (DMEM; Thermo Fisher Scientific) supplemented with 10% fetal bovine serum (FBS; GE Healthcare) and penicillin-streptomycin (100 U/mL and 100 mg/mL, respectively; Thermo Fisher Scientific). For all experiments, the same passage number of subject and control fibroblasts was used. Primary fibroblasts were regularly tested for mycoplasma contamination and confirmed to be mycoplasma free.

### Generation and culture of iPSCs

Healthy control iPSCs were obtained through the *StemBANCC Consortium*. Patients-derived fibroblasts (P5 (F5-II:2) and P6 (F6-II:7)) were reprogrammed by non-integrating Sendai viral vectors at Oxford StemTech (Oxford, United Kingdom). All iPSC lines were subject to quality control checks, including flow cytometry for pluripotency markers, global screening array karyotyping, and mycoplasma test. iPSCs were plated in Matrigel-coated 6-well plates and maintained in mTesR1 (StemCell Technologies).

### Differentiation of iPSCs to spinal motor neurons

iPSCs were differentiated into MNs using a protocol previously published (16). Briefly, iPSCs were dissociated with accutase and resuspended in a 10mm2 Petri dish to form embryoid bodies (EBs). Differentiation medium consisted of (1:1 DMEM/F12-Neurobasal media, supplemented with N2, B27, 2 mM L-glutamine, 1% Pen-Strep, 0.1 mM β-ME; all from ThermoFisher Scientific), with 10 μM Y-27632 (Tocris), 0.1 μM LDN 193189, 20 μM SB431542, and 3 μM CHIR-99021 (Cambridge Bioscience). Media was replaced every 2-3 days, with the addition of the following small molecules: 100 nM retinoic acid (RA, Sigma Aldrich) from day 2; 500 nM Smoothened Agonist (SAG, Sigma Aldrich) from day 4; 10 μM DAPT (Cambridge Bioscience) from day 9. LDN 193189, SB431542 and CHIR-99201 were discontinued on day 7. The neurotrophic factors BDNF, CNTF and GDNF (Peprotech) were added to the differentiation medium from day 11, at a concentration of 10 ng/mL. On day 14, EBs were dissociated, and post-mitotic neurons were seeded on poly-L-ornithine (Sigma Aldrich) and laminin (Biotechne) coated plates. Eleven days after seeding cells were fixed for immunocytochemistry and harvested for RNA and protein extraction.

### Migration assay

Migration assays were performed and analysed as previously described(17). 100,000 cells were resuspended in serum-free medium and plated on the top chamber (24-well transwell insert, Falcon: 353097). Cells were incubated at 37°C for 24h, which allowed migration towards the bottom chamber containing complete medium with 10% FBS. Cells on the bottom surface of the insert were fixed in 10% formalin (BioShop: FOR201) and stained with a crystal violet solution. Ten images were taken for each transwell insert using a Nikon inverted microscope camera with a 10X objective lens (Nikon Eclipse TE300 Inverted microscope). Quantitative analysis was assessed using Image J software. Data represent the fold change relative to wild type cells obtained from at least three independent experiments.

### Wound healing scratch assay

15,000 cells were seeded in 100 µL of culture medium into sterile transparent 96-well plates and incubated for 24h. A scratch wound was made in the confluent cell monolayer of each well using the IncuCyte 96-well WoundMaker from Essen Bioscience as described in manufacturer’s manual. After carefully removing the cellular debris, 100 μL of culture medium was added to each well. Cell images were captured every two hours using IncuCyte Live-Cell Imaging Systems (Essen BioScience, USA). Images were analysed using the IncuCyte S3 software (2019A) to calculate cell confluency over time.

### RNA isolation, cDNA synthesis, and quantitative real-time PCR (RT-qPCR)

Total RNA was extracted from cultured primary fibroblasts of subjects P5 (F5-II:2), P6 (F6-II:7), and P11 (F10-II:1) and controls and from iPSC motor neuron of subjects P5 (F5-II:2) and P6 (F6-II:7) and controls (using QIAGEN RNeasy extraction kit). gDNA purification was performed with DNA-*free*™ DNase (Thermofisher). RNA concentration and purity of the samples were assessed using NanoDrop equipment (NanoDrop Technologies Inc., Wilmington, DE). 0.5 µg total RNA was reverse transcribed (Superscript III, Thermofisher). Technical triplicates of RT-qPCR samples were prepared as a 10 µL approach with the Fast SYBR™ Green Master Mix (Thermofisher), 500 nM of each primer, and 1 µl of the reverse transcription reaction. Primer sequences for RT-qPCR are described in Supplementary Table 6. RT-qPCR was performed using the QuantStudio 3 Real-Time PCR System (Thermo Fisher Scientific) equipped with QuantStudio Design&Analysis Software v1.4.3 (Thermo Fisher Scientific). The PCR conditions included a pre-run at 95°C for 5 min, followed by 40 cycles of 30 s at 95°C, 30 s at 58°C and 45 s at 72°C. PCR amplification specificity was determined by melting curve analysis with a range from 60°C to 95°C. The values of the cycle threshold (CT) of the target mRNAs were normalized to the mRNA of *RPL18*. For relative gene expression, the comparative cycle threshold (ΔΔCT) values were calculated with the QuantStudio Design&Analysis Software (Thermo Fisher Scientific) with *RPL18* as housekeeping gene and expressed as x-fold change to controls.

### RNA sequencing and analysis

RNA sequencing was performed on RNA from subjects P5 (F5-II:2), P6 (F6-II:7) and P11 (F10-II:1) as well as 3 controls using the Illumina TruSeq Stranded mRNA Library Prep kit. Pools were sequenced using the Illumina NovaSeq 6000 Sequencing system to generate 100 bp paired-end reads with an average read depth of ∼100 million reads per sample. RNA sequencing raw data was mapped to human reference genome assembly hg38 using STAR (18). Gene count tables were imported into R environment to analyse differential gene expression patterns using DESeq2 package (19). For gene enrichment analysis, pathfindR package was employed (20). Briefly, Biogrid database was used for mapping the genes to protein-protein interaction network, with their associated p-values. The resulting active subnetworks were filtered based on the significant genes they contain. The filtered subnetworks were then enriched by the related cellular pathways. This was followed by hierarchical clustering to group biologically relevant pathways. Major pathways with enrichment fold greater than 2 were selected for plotting per sample expression scores.

### Western blotting

Fibroblasts of subjects P5 (F5-II:2), P6 (F6-II:7) and P11 (F10-II:1) and controls and fibroblast-derived iPSc motor neurons of patients subjects P5 (F5-II:2) and P6 (F6-II:7) and controls were harvested in ice-cold RIPA buffer (50 mM Tris-HCl, pH 8.0; 150 mM NaCl; 1% NP-40; 0.5% DOC [sodium deoxycholate]; 0.1% SDS [sodium dodecyl sulfate]) supplemented with Mini Protease Inhibitor (Roche) and lysed for 60 min on ice. Cell debris was removed by centrifugation for 10 min. Protein extracts were separated on 4-12 % SDS polyacrylamide gel under denaturing conditions and transferred to polyvinylidene fluoride membranes. Membranes were blocked followed by incubation with a ARHGAP19 primary antibody (Santa Cruz Biotechnology; 1:500) overnight at 4°C. After washing, membranes were incubated HRP-linked anti-mouse secondary antibody (#G-21040 Invitrogen; 1:10,000) at room temperature for 1 h. After washing, immunoblots were digitally imaged using a Machine iBright 750 (invitrogen). Exposure time was optimized to avoid saturation. Bands were automatically defined, and intensities were determined using the build-in band detection tool of the Image Lab v6.0 software (Bio-Rad). GAPDH (ab9484, Abcam; 1:10000) was used as a loading control. Blots were repeated in duplicates and statistics were performed using Graph Pad Prism. The significance between the variables was shown based on the p value obtained (ns indicates p > 0.05, *p < 0.05, **p < 0.005, ***p <0.0005, ****p < 0.00005).

### Protein expression and purification

Recombinant wild type and mutant GST-tagged ARHGAP19 GAP domain proteins were expressed in BL21 (DE3) bacteria and purified using glutathione-sepharose beads as previously described (21). PreScission protease (GenScript) was used for the cleavage of the GST tag and elution of the purified ARHGAP19 GAP proteins. Eluted proteins were concentrated using Amicon Ultra-4 centrifugal filters (MilliporeSigma), resolved by SDS-PAGE followed by Coomassie blue staining. Proteins were quantified using bovine serum albumin (BSA) as standard.

### *In vitro* GAP assay

The GAP activity of ARHGAP19 wild type and protein mutants was assessed using the RhoGAP assay biochem kit (BK105, Cytoskeleton) according to manufacturer’s instructions and recommendations. Briefly, 1.5 μg of purified ARHGAP19 protein was mixed with His-RhoA protein and GTP for 20 min at 37 °C. CytoPhos reagent was added to the reaction mixture for 10 min at room temperature before measuring the absorbance at 650nm with Infinite M200 Pro Microplate reader (TECAN).

### *Drosophila* stocks and husbandry

Flies were raised on standard fly food at 25°C in 12 h light: 12 h dark cycles. All experiments and crosses were conducted in these conditions unless otherwise specified. The following *Drosophila* stocks were obtained from the Bloomington Drosophila stock center: *y[1] w[*]; P{Act5C-GAL4-w}E1/CyO* (actin-Gal4; BDSC #25374), [1] *v[1]; P{y[+t7.7] v[+t1.8]=TRiP.HMS03522}attP40* (UAS-RhoGAP54D shRNA; BDSC #54051), *y[1] w[*]; TI{GFP[3xP3.cLa]=CRIMIC.TG4.1}RhoGAP54D[CR02433-TG4.1]/SM6a* (RhoGAP54D-Gal4; BDSC #92267), *y[1] w[*]; P{w[+mC]=tubP-GAL4}LL7/TM3, Sb[1] Ser[1]* (tubulin-Gal4; BDSC #5138), and w[*]; P{w[+mC]=UAS-Nslmb-vhhGFP4}3 (BDSC #38421). The RhoGAP54D::GFP fusion allele (w[*]; *RhoGAP54d:GFP:2Ma*; MKRS/TM6B) was generated via CRISPR-Cas9 gene editing, as described previously (22). The w[*]; P{Act5C-GAL4-w}E1/CyO insertion was outcrossed into an isogenized background (iso31) for five generations, with the X-linked *y*[1] marker removed in the process (Supplementary Table 7).

### Generation of a *Drosophila RhoGAP54D* null allele

The *RhoGAP54D*^KO^ loss of function allele was generated using CRISPR/Cas9 mediated homologous recombination. We generated homologous recombination donor and guide plasmids to replace the *RhoGAP54D* reading frame from amino-acids (AA) T4 to L1004 (numbered according to the isoform RA, 1004 AA in length) by a *mw* gene flanked by two attP Phi31 recombination sites using the pCRISPR-del (23) and the pCFD5 (24) plasmids. pCFD5 was a gift from Simon Bullock (Addgene plasmid # 73914; http://n2t.net/addgene:73914; RRID:Addgene_73914). To generate the pCRISPR-del-RhoGAP54D targeting the *RhoGAP54D* locus, the genomic 5’ HR1 (∼1kb) and 3’ HR2 (∼1kb) regions were PCR amplified using the following primers: HR1 5’- <u>CCGGGCTAATTATGGGGTGTCGCCCTTCGC</u>GGATCTCCGTAGACGCCGTTC and 5’- <u>ACTCAAAGGTTACCCCAGTTGGGGCACTAC</u>TGCTTCCATGCAATCTGTGTGGTTTATCC, HR2 5’- <u>ACTCAAAGGTTACCCCAGTTGGGGCACTAC</u>GACACGTTTAGCTTGGCCGCG and 5’- <u>GCCCTTGAACTCGATTGACGCTCTTCTGTA</u>CAAGCCACCCCACACACTGAG to clone them in pCRISPR-del (underlined nucleotides matching the plasmid sequence used for cloning). We used 4 distinct gRNA, two in Nter and two in Cter to the *RhoGAP54D* locus, cloned in tandem in gRNA pCFD5 (Addgene #73914) plasmid to generate 2 pCFD5-RhoGAP54D guide plasmids using the following primer: 5- <u>GCGGCCCGGGTTCGATTCCCGGCCGATGCA</u>AGATTGCATGGAAGCAACGA<u>GTTTTAGAGCTAGAAATAGCAAG</u> and 5’- <u>ATTTTAACTTGCTATTTCTAGCTCTAAAAC</u>TTTCGCCCGGATACTTCTCGTGC<u>ACCAGCCGGGAATCGAAC</u> as well as <u>5’- GCGGCCCGGGTTCGATTCCCGGCCGATGCA</u>AGATTGCATGGAAGCAACGA<u>GTTTTAGAGCTAGAAATAGCAAG</u> and <u>5’</u>- <u>ATTTTAACTTGCTATTTCTAGCTCTAAAAC</u>TATCCATCGTTGCTTCCATG<u>TGCACCAGCCGGGAATCGAAC-</u>. Cloning was performed by primer annealing and ligation, or by SLIC. All regions amplified by PCR as well as junctions between tag and genomic sequences were checked by sequencing. *RhoGAP54D*^KO^ allele was then generated by co-injection of the pCRISPR-del-RhoGAP54D (300ng/ml) and each of the guide plasmid (25ng/ml) in embryos of the PBac{y+-attP-9A}VK00027 (BDSC #51324) line. Embryos injection was performed by BestGene transgenesis services. The RhoGAP54D^KO^ genomic deletion and locus organization were confirmed by PCR and sequencing.

### *Drosophila* activity assays

To quantify locomotion in adult male flies, the *Drosophila* Activity Monitor (DAM; Trikinetics inc., MA, USA) was used as described previously(25). Individual flies were housed in glass tubes containing 4% sucrose and 2% agar, bisected by an infrared beam, allowing activity to be quantified as the number of beam breaks made by each fly over time. Fly movement was measured at 25°C across 24 h comprised of 12 h light and 12 h dark period, from which total activity and activity during ZT (zeitgeber time) 12-13 were quantified. Experiments were performed on 3-7 day old male flies. All flies were acclimatised to the DAM conditions for 48 h prior to the experimental period.

### *Drosophila* immuno-histochemistry

Adult brains were dissected and immuno-stained as described previously(26). Briefly, brains were fixed for 20 min at room temperature via incubation in 4% paraformaldehyde (MP Biomedicals), and blocked for 1 h in 5% normal goat serum in Phosphate Buffered Saline containing 0.3% Triton-X (Sigma-Aldrich) (0.3% PBT). Primary antibodies were as follows: mouse anti-Bruchpilot (BRP) (nc82, Developmental Studies Hybridoma Bank), 1:50; rabbit anti-dsRed (Clontech, Cat. no. #632496), 1:1000. Secondary antibodies were: goat anti-rabbit AlexaFluor-555 (ThermoFisher, Cat. no. #A32732), 1:1000; and goat anti-mouse AlexaFluor-647 (ThermoFisher Cat. no. #A21236), 1:500. Brains were incubated in primary and secondary antibodies overnight at 4°C. Brains were washed then mounted and imaged in SlowFade Gold anti-fade mounting solution (ThermoFisher Scientific). Images were taken using a Zeiss LSM 710 confocal microscope with an EC ‘Plan-Neoflar’ 20x air objective. Images were analysed using ImageJ.

### Quantitative PCR

Total RNA was extracted from dissected brain tissue of *actin* > UAS-*RhoGAP54D* shRNA and *actin* > UAS-*mCherry* shRNA (control) adult male flies using standard phenol chloroform extraction. RNA concentration and purity were assessed using NanoDrop equipment (NanoDrop Technologies Inc., Wilmington, DE). 0.5 µg of total RNA was reverse transcribed to generate cDNA (Superscript III, Thermofisher). Technical duplicates of the qPCR samples were prepared as a 10 µL approach with the Fast SYBR™ Green Master Mix (Thermofisher), using 500 nM of each primer (forward: ATGGAAGCAACGATGGATACG; reverse: CTCGTGACAGGGGAGATCGAA), and 1 µl of the reverse transcription reaction. qPCR was performed using the QuantStudio 3 Real-Time PCR System (Thermo Fisher Scientific) equipped with QuantStudio Design&Analysis Software v1.4.3 (Thermo Fisher Scientific). The PCR conditions included a pre-run at 95°C for 5 min, followed by 40 cycles of 30 s at 95°C, 30 s at 58°C and 45 s at 72°C. PCR amplification specificity was determined by melting curve analysis with a range from 60°C to 95°C. The values of the cycle threshold (CT) of the target mRNAs were normalized to the mRNA of RpL4 (forward primer: TCCACCTTGAAGAAGGGCTA; reverse primer: TTGCGGATCTCCTCAGACTT). For relative gene expression, the comparative cycle threshold (ΔCT) values were calculated with the QuantStudio Design&Analysis Software (Thermo Fisher Scientific) with RpL4 used as a control gene. *RhoGAP54D* expression was normalised to *RpL4* and expressed as a fold change relative to *actin* > UAS-*mCherry* shRNA controls.

### *Danio rerio* functional analyses

Wild-type zebrafish, *Danio rerio*, were housed and bred within UCL Fish Facility at 28.5°C on a 14h day/10h dark cycle. All experiments were conducted under licences awarded by the UK Animal (Scientific Procedures) Act 1986 implemented by the Home Office in England. For whole-mount *in situ* hybridization, zebrafish embryos were first fixed in 4% paraformaldehyde (PFA/PBS) overnight at 4°C, dechorionated, and dehydrated in methanol at −20°C. A T7 bacteriophage promoter containing ampliconof *arhgap19* was synthesised from cDNA libraries at three different developmental stages: 24 hours post-fertilization (hpf), 48 hpf, and 5 days post-fertilization (dpf). A primer set was designed with the T7 bacteriophage promoter sequence incorporated at the 5’ end of the reverse primer for antisense probe production (Forward primer: 5’-<u>GGCCGAATTCTCACAGCTAC</u>-3’; Revers primer: 5’- **<u>TAATACGACTCACTATAGG</u>**TCTTACACGCGCTGATGAAC-3’). A Digoxigenin-labeled antisense probe was synthesized using a DIG-RNA Labeling Kit (T7 polymerases, Roche). Whole-mount in situ hybridization (WISH) was carried out as previously described (27) on 48 hpf, 72 hpf, and 5 dpf larvae. For the color reaction, NBT/BCIP Stock Solution (Roche) was used in the staining buffer. The stain was fixed in methanol and embryos mounted in 3% methylcellulose for imaging.

The zebrafish arhgap19 gene (ZDB-GENE-100922-157) is an ortholog of *Homo sapiens* ARHGAP19; there is 59.95 % nucleotides similarity and 55 % amino acid similarity between the zebrafish and human loci. No zebrafish paralogs corresponding to ARHGAP19 exist. CRISPR/Cas9-mediated F0 biallelic knockout was performed as described previously (28). Six crRNAs were designed according to their on-target and off-target scores. The six sgRNAs sequences were: 1) 5’- <u>ACGCTCCTCAAGAGTTTCTT</u>-3’, 2) 5’-<u>CAAGATGTCTGCTCACAACC</u>-3’, 3) 5’- <u>CAAGAGTTTCTTGGGAGAAT</u>-3’, 4) 5’-<u>GAACCCAAGACTCCCAACGC</u>-3’, 5) 5’-<u>TGACTTTCATCCCAATGACG</u>-3’, 6) 5’- <u>AGGCAACAAGACGAGTTTTC</u>-3’. Each cRNA was annealed to tracrRNA and complexed to Alt-R S.p. HiFi Cas9 nuclease to form a ribonucleoprotein (RNP). A pool of three RNPs were co-injected into one to two-cell stage zebrafish embryos. For each RNP pool, 0.5nL and 1nL volumes were injected into the yolk of batches of embryos. At 5 dpf, three zebrafish larvae were randomly selected for genomic DNA extraction to determine the targeting efficiency by Illumina Miseq (Eurofins). PCR amplification targeting RNP cut sites were performed for each crRNA and products purified, pooled and sent for next generation sequencing. Primers included universal adaptors for MiSeq (Supplementary Table 5). Cut efficiency was calculated for each RNP using CRISPResso2 (29). An antisense translational blocking Morpholino (MO, GeneTools.LLC) against the AUG-containing mRNA sequence (5ʹ-GGCCATCTTTCATCTTCCGTTTGAA-3ʹ) and a splice MO targeting the Exon1-Intron1 (E1I1) boundary (5ʹ-ATAAATCTTCGTTACCTTCTGTCTC-3ʹ) was designed to knockdown arhgap19 function. MOs were diluted to the desired working concentrations (3 ng, 4 ng, 6 ng, and 8ng per embryo) before use. Microinjection was performed by injecting 0.5-1nL morpholino solution into one to two-cell stage embryos in the yolk. Dose dependent phenotyping was used to identify an appropriate concentration that balanced survival with specific phenotypic changes.

### *Danio rerio* behavioural assays

Zebrafish larvae at 5 dpf were transferred into individual wells of a square flat-bottomed96-well plate. Baseline locomotor activity was recorded for a duration of 30 minutes and analysed using the DanioVision (Noldus, Netherlands) monitoring chamber, which was integrated with the EthoVision XT 14 video tracking software (Noldus, Netherlands). Plots were analysed for distance travelled (in millimeters), total time spent and velocity. For assessment of muscle integrity**, z**ebrafish larvae at 5 dpf were fixed and their skeletal muscle was analysed for total birefringence using polarized light microscopy on a Nikon SMZ1000 stereo microscope.

### *Danio rerio* Immuno-histochemistry

Zebrafish larvae at 5 dpf were permeabilized with 10ug/ml Proteinase in PBS Tween (0.2%, PBT)K, followed by 20 mins fixation in 4% PFA. Larvae were then blocked with goat serum (5% in PBT), and incubated with anti-tubulin mouse monoclonal antibody (1:500 dilution; T6793-2ML, Sigma) overnight at 4°C. After four washing steps (PBT), larvae were incubated with Alexa Fluor™ anti-mouse 568 secondary antibody (1:1000 dilution; A-11004, ThermoFisher) in the dark at 4°C. For nucleus detection, larvae were incubated with DAPI (1:1000 dilution; D1306, ThermoFisher) under dark conditions at 4°C. Embryos were washed again in PBT (four times) and imaged using a Nikon A1R confocal microscope to assess motor neuron morphology. Confocal fluorescent images were processed and code depth adjusted with FIJI/ImageJ while motor neuron quantification was conducted using the SNT plugin.

### Statistical Analysis

Statistical evaluations were carried out using IBM SPSS Statistics. One-way ANOVA and Welch’s t-test were used for statistical comparisons between groups with calculated standard deviations/errors of the mean. For *Drosophila* work, we run non-parametric tests for normal distribution using Shapiro-Wilk test for normality. Normally distributed datasets were subject to tests as stated. Non-normal were subject to Mann-Whitney U-test or Kruskal-Wallis test with Dunn’s post-hoc test (single or multiple comparisons respectively). Significant differences were determined at a threshold of p < 0.05. The corresponding p-values are provided in the figure legends.

### Data availability

The data that supports the findings of this study are available within the paper and in the supplementary material. Whole-exome sequencing data are not publicly available due to privacy or ethical restrictions. The identified variants reported in this manuscript were submitted to the LOVD database (https://databases.lovd.nl/shared/genes/ARHGAP19), with the LOVD variant IDs: #0000971425, #0000971426, #0000971427, #0000971440, #0000971442, #0000971444, #0000971446, #0000971450, #0000971466, #0000971468, #0000971470, #0000971471, #0000971473, #0000971474, #0000971475, #0000971478, #0000971479 and #0000971480.

## Results

### Genetic Findings

We identified 20 unrelated families with 25 affected individuals harbouring biallelic variants in *ARHGAP19* (Figure 1B for variant schematic within the protein and Figure 2A for pedigrees). All variant alleles identified in the families were either absent or observed only as heterozygous at extremely low frequencies in over 1.5 million alleles across multiple publicly available and private genetic variant databases (range 0–0.003). Four variants (4/16, 25%) were observed in more than one family. Notably, p.His196Gln*fs*\*9, p.Leu68Pro, p.Gln151Lys and p.Leu228His variants were found in 2 independent Arab, 4 independent Turkish, and 2 independent Bangladeshi/Afghani families, respectively.

**Figure 1.**
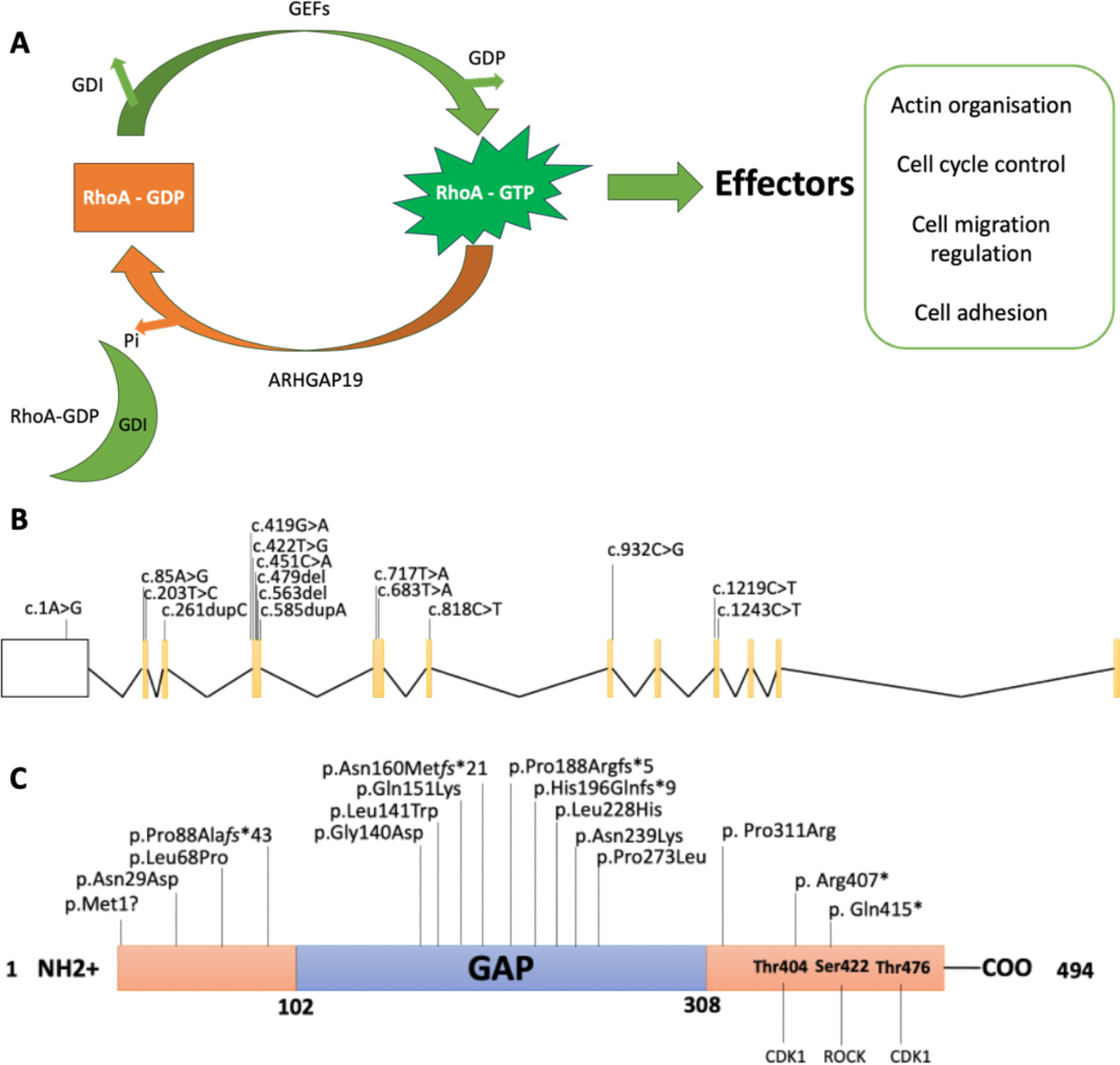
**A.** Pathway showing GTPase-activating proteins (GAPs) such as ARHGAP19 responsible for promoting cycling of Rho GTPases between the active GTP-bound and the inactive GDP-bound conformations. GEF: Guanine nucleotide Exchange Factor, GDP: Guanine nucleotide diphosphate, GDI: Guanine Nucleotide Dissociation Inhibitor, Pi: dihydrogen phosphate. **B-C.** Schematic diagrams of ARHGAP19 gene (B) and protein (C). Introns are not to scale. Exon numbers are according to the canonical transcript (NM_032900.6). Amino acid changes are according to the reference sequence NP_116289.4.

**Figure 2.**
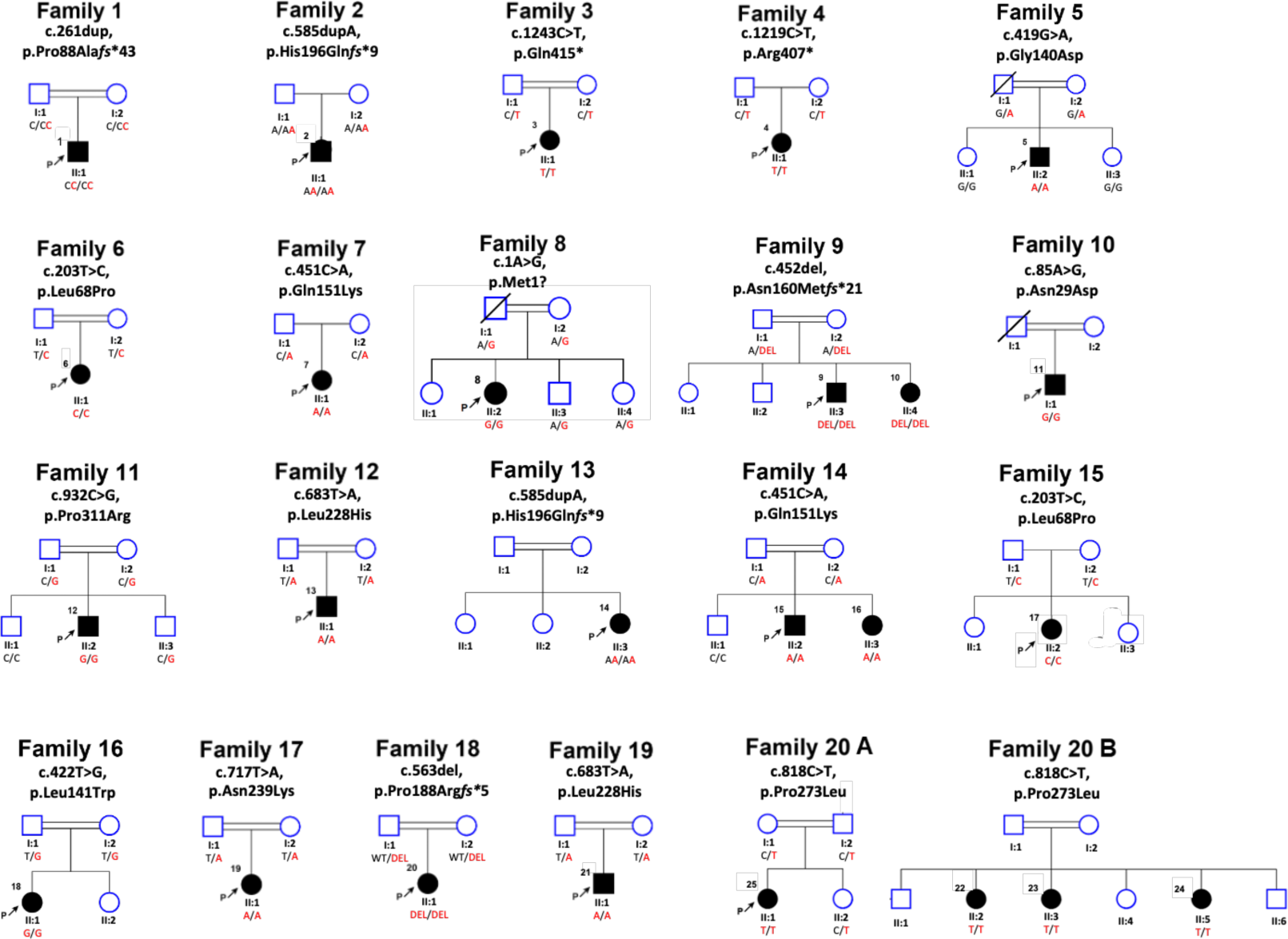
Genetic presentation of individuals harbouring *ARHGAP19* variants. Pedigrees of affected families showing segregation of the biallelic *ARHGAP19* variants identified.

Haplotype plots of homozygous regions encompassing *ARHGAP19* variants were compared to assess if the same causative variants are inherited from a common ancestor (Supplementary Figure 1). The chr10-97259559-A-T (p.Leu228His) and chr10:97263447-G-GT (p.His196Gln*fs*\*9) variants are recurrent since patients harbouring the shared variants have different haplotypes. Patients F14-II:2 and F7-II:1 harbour the chr10-97263582-G-T variant (p.Gln151Lys) and share a large 3.4 MB long haplotype. MRCA analysis shows that this founder variant has emerged about 26 generations ago (equivalent to 520 years). Variant chr10-97265979-A-G (p.Leu68Pro) is the other founder variant that we detected in patients F15-II:3 and F6-II:7 who harbour the same haplotype. The shared homozygous region is 3.0 MB in length, and they have inherited the variant from a common ancestor back to about 29 generations ago (equivalent to 580 years).

### Clinical Delineation

Table 1 summarises the core clinical features of affected probands with ARHGAP19 defects (see Supplementary Table 1 for Neurophysiology and for detailed clinical vignettes please contact the corresponding author to request access to these materials). A total of 25 individuals were identified from 20 families and 9 (36%) were male. They were from a wide range of ancestral backgrounds including Pakistan, Turkey, Egypt, Syria, Bangladesh, Spain, Australia, Brazil, Iran, Dubai and Afghanistan. 72% (18/25) were born from consanguineous parents. The mean (median) age at symptom onset (AAO) was 9.9 (10.0) years, and at assessment was 22.8 (16.0) years. The presenting symptom was a motor deficit of the lower limbs in 91% (21/23), and 64% (14/22) had some form of clinical sensory involvement (symptoms or signs), but as the disease progressed it typically remained either exclusively motor or motor predominant. Patients typically had a length-dependent pattern (17/23, 74%, which includes those with only distal lower limb involvement) of lower motor neuron signs of areflexia and muscle atrophy, with foot drop. Lower limb-predominant disease was seen (distal > proximal weakness, with normal upper limbs) in 17% (4/23) and upper limb predominant disease seen in 9% (2/23). Foot deformity was present in 80% (16/20), with a recurrent, but not ubiquitous finding of increased sandal gap (access to photos of deformities mentioned can be accessed by contacting the corresponding author). However, the presence brisk knee jerks, and preserved lower limb reflexes each in 9% (3/23) respectively suggests some mild UMN involvement in these individuals. There were no consistent features outside of the peripheral nervous system.

A prominent feature of the phenotype is its significant asymmetry in terms of limb involvement, seen at onset or at assessment in 61% (14/23). Two cases presented acutely with upper limb weakness on a background of mild or subclinical widespread neuropathy. Neurophysiology (Supplementary Table 1) was performed in 20 individuals. Detailed numerical study data was available in 15/20; five cases had a report only. All had a motor neuropathy, with variable sensory involvement. Evidence of motor conduction slowing was seen or described in 50% (10/20) and conduction block in 20% of the studies with numerical data (3/15). Combining clinical and neurophysiological data the following phenotypes were seen: CMT-intermediate (CMTi) (2/25), CMT2 (6/25), hereditary motor neuropathy (HMN, 7/25) and CMT (indeterminate from neurophysiology, or no neurophysiology available, 10/25). Within the cases of CMT2/HMN, 38% (5/13) demonstrate conduction slowing that does not meet criteria for CMTi or CMT1 (upper limb motor conduction velocities 25-45 m/s) either because the slowing was patchy or subtle, or the phenotype was HMN. The mean (median) ulnar motor velocity was 42.6 (45) m/s (range 25-58 m/s, n = 11).

It is difficult to comment on disease progression with only a single assessment in many cases, but in some individuals, from the history there is a more rapid progression than is typical for CMT, with mean (median) duration between AAO and assessment of 13.0 (5) years; the mean difference is skewed by some outlying cases of mildly affected relatives. Two cases presented with acute upper limb weakness. The first (P13 (F12-II:1)), a male infant carrying the recurrent variant p.Leu228His presented with bilateral upper limb weakness over fifteen days. Spinal imaging was normal, but neurophysiology was never performed. Upper limb function improved over time, but the patient sadly died before 5 years of age. The second (P17 (F15-II:2)), a teenage female carrying the recurrent p.Leu68Pro, presented with acute left-hand weakness, on a background of more widespread conduction slowing neuropathy. She was treated with intravenous immunoglobulin (Ig) for presumed chronic inflammatory demyelinating polyneuropathy (CIDP), with no clear response. Interestingly, P6 (F6-II:7), a female who carries the same homozygous variant, had walking difficulty from infancy but presented in her teenage years with a four-month deterioration in left lower limb weakness. She had a conduction slowing neuropathy in the lower limbs. She is currently being treated with subcutaneous Ig for presumed CIDP.

### In silico modelling

Multiple sequence alignments performed for ARHGAP19 orthologs across 11 animal species (Figure 2A), anchored to the human ARHGAP19 protein sequence, showed that most variants affect highly conserved amino acid residues. Of note, the variants affecting moderately conserved residues (Asn160, Arg407 and Gln415) are frameshift or nonsense. All variants segregated with disease within the families (Figure 2A). Computational variant prediction tools such as MutationTaster, SIFT and PolyPhen predict the functional impact of all but two variants as mainly damaging and deleterious. According to the American College of Medical Genetics and Genomics and the Association for Molecular Pathology (ACMG-AMP) system for variant classification, all the variants are classified as either likely pathogenic or pathogenic. The characteristics of all reported variants are summarized in Supplementary Table 2.

Three-dimensional visualisation of ARHGAP19 wild-type and mutant variants as predicted by AlphaFold2 (Figure 3B) depicts that mutant protein structures for frameshift variants show substantial deviation from the wild-type protein (Figure 3C), likely disrupting the RhoGAP domain structure. Nonsense substitutions modelled using AlphaFold2 (Figure 3D) display a similar structure to the WT protein, with greater variation only occurring in the final alpha-helix and the subsequent C-terminal end of the protein. Missense substitutions showed little or no change when modelled with the AlphaFold2 pipeline. As such, the ‘mutagenesis’ function on PyMol was used to predict changes in protein structure and/or folding upon substituted amino acid residue incorporation into the sequence (Figure 3E). All substitutions showed changes in steric hindrance with nearby amino acid residues. The calculated free energy changes for p.(Gly140Asp), p.(Leu141Trp), p.(Gln151Lys), p.(Leu228His), and p.(Pro311Arg) substitutions show a decrease in free energy >1.6 kcal/mol, indicating a protein destabilising effect (Figure 3F). Notably, three substitutions are predicted to result in protein instability with high confidence (p.(Gly140Asp), p.(Leu141Trp), and p.(Pro311Arg)). In addition, the p.(Gln151Lys) variant shows an increase in free energy and thus an increase in protein stability. Using further analysis with AlphaMissense, of the nine missense variants identified, seven (77.78%) are predicted to be likely pathogenic (Supplementary Table 4).

**Figure 3.**
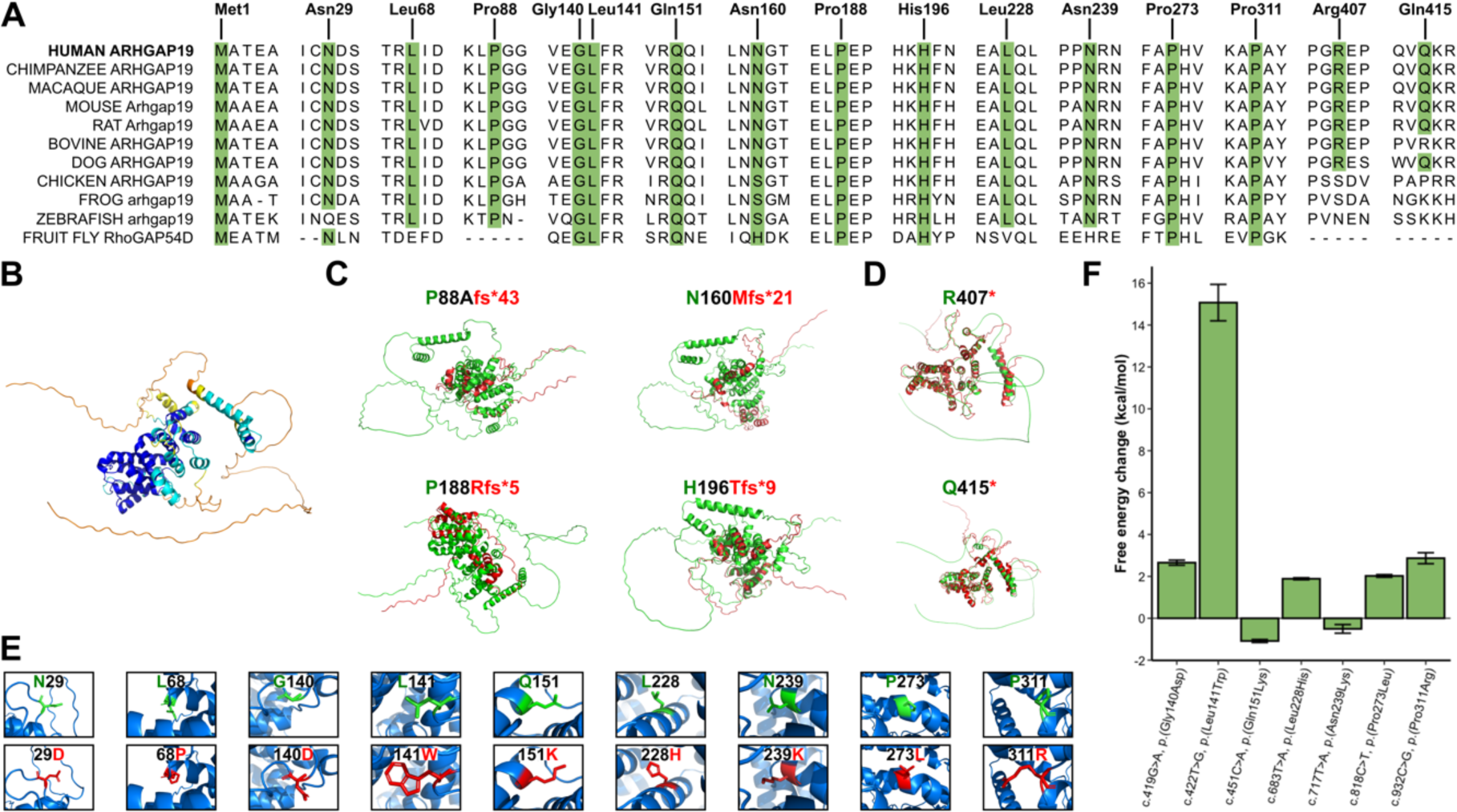
Investigating the effect of *ARHGAP19* variants *in silico*. **A.** Multiple sequence alignment of ARHGAP19 orthologs shows conservation of identified variants in different species. **B.** Predicted wild-type ARHGAP19 protein structure (UniProt: Q14CB8) using AlphaFold2, coloured with the per-residue confidence score (pLDDT). Dark blue, very high confidence (pLDDT > 90); light blue, confident (90 > pLDDT > 70); yellow, low confidence (70 > pLDDT > 50); orange, very low confidence (pLDDT < 50). AlphaFold-generated mutant protein structures (red) derived from the identified frameshift **C.** and nonsense **D.** variants and aligned to the wild-type ARHGAP19 protein (green). **E.** The effect of identified missense variants on three-dimensional ARHGAP19 structure. Wild-type residues are coloured green and mutant residues are coloured red. **F.** Predicted free energy changes (ΔGMUT – ΔGWT) for missense variants on protein stability. Calculations were performed on variants within the very high confidence regions (pLDDT >90) of the AlphaFold-generated ARHGAP19 protein. Data are given as mean ± s.d.

### Functional Characterisation

#### Western Blotting and qPCR

Given the potential loss of function in homozygous ARHGAP19 individuals, we investigated gene expression levels through the use of qPCR and protein levels through Western blotting of ARHGAP19 in fibroblasts from families harbouring the c.85A>G (p.Asn29Asp), c.419G>A (p.Gly140Asp), c.203T>C (p.Leu68Pro) variants and in fibroblast-derived iPSc of P5 (F5-II:2) and P6 (F6-II:7). Whist gene expression remained unchanged in both fibroblasts and motor neurones, western blotting assessment of ARHGAP19 protein levels revealed significant reduction of ARHGAP19 in iPSC motor neurons of patients as opposed to controls (Figure 4).

**Figure 4.**
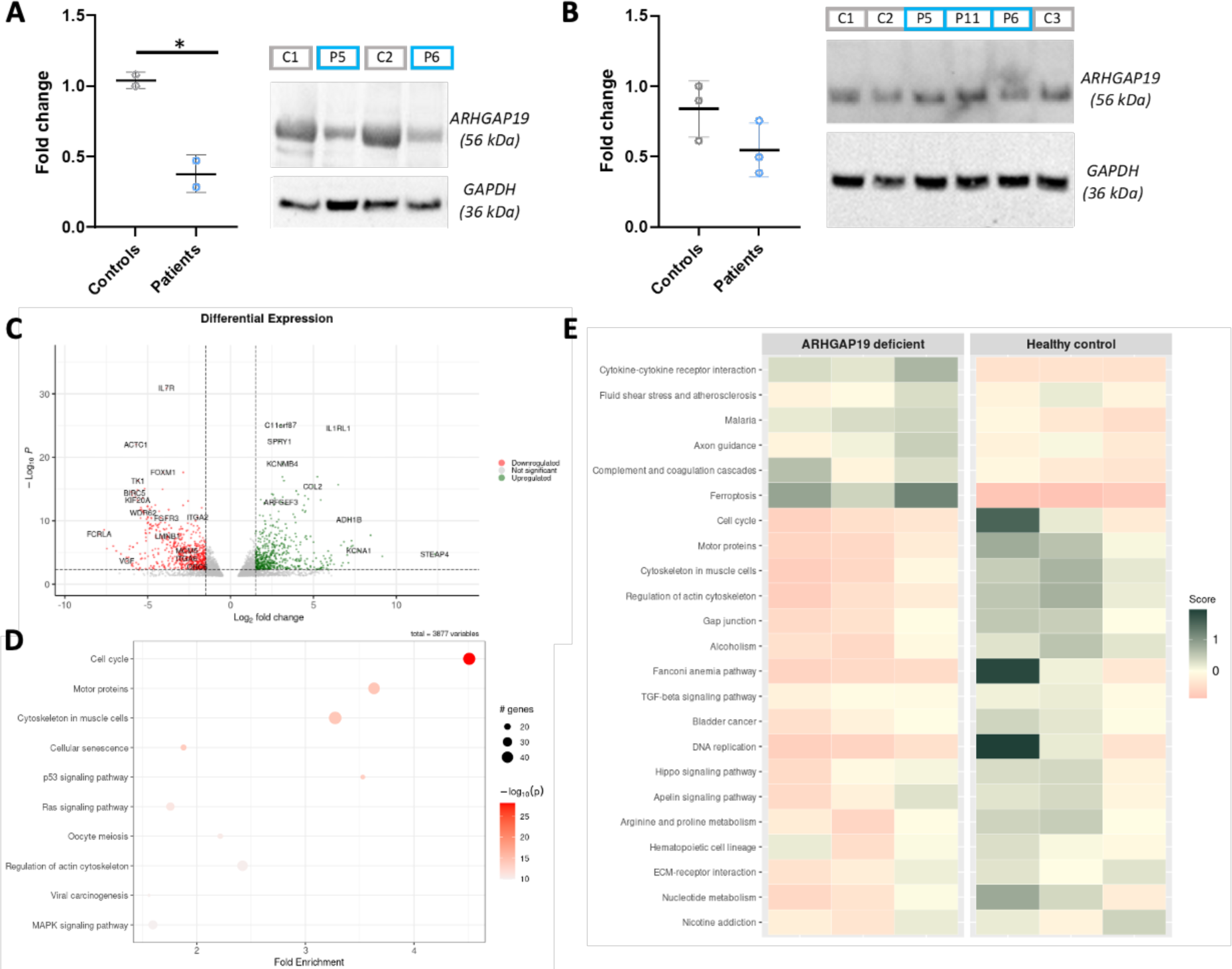
Western blot and RNA-Seq analyses confirm downregulation of ARHGAP19 as well as cell cycle, motor and muscular cytoskeleton pathways. **A-B.** Protein expression levels of ARHGAP19 in iPSc-derived motor neuronal lines (A) and patient-derived fibroblasts (B). Fibroblasts from P5 (F5-II:2), P6 (F6-II:7) and P11 (F10-II:1) harbouring the c.419G>A (p.Gly140Asp), c.203T>C (p.Leu68Pro) and c.85A>G (p.Asn29Asp) variants respectively were analysed by Western blot, as well as fibroblast-derived iPSc neurons from P5 (F5-II:2) and P6 (F6-II:7) patients. **C.** Volcano plot showing log2 of fold change in *ARHGAP19* mutants compared to controls and −log10 (adjusted p value). **D.** Pathway enrichment: differentially expressed genes with p-value < 0.005 were selected for performing pathways enrichment analysis. Top 10 pathways with the highest enrichment scores were plotted, showing 3 cellular pathways highly enriched with dozens of affected genes and a very low p-value: cell cycle, motor proteins, and cytoskeleton in muscle cells. **E.** Pathway scores per sample: enriched pathways were analysed to compare their expression levels in patients and controls. Some pathways are overexpressed in patients while most of them are downregulated.

### RNA Sequencing

Differential expression analysis showed that there is no difference in ARHGAP19 RNA levels between the ARHGAP19 deficient patient fibroblasts and healthy controls (log2 fold change = −0.29 and p-value = 0.43). Thus, we hypothesised that despite having the same expression level, mutant ARHGAP19 may affect downstream genes in its signalling pathway, causing the disease phenotype. Principle component analysis (PCA) showed a distinctive RNA expression pattern between the patients and controls (Supplementary Figure 2). Pathway enrichment analysis showed that the expression patterns are significantly different in 3 pathways: cell cycle, motor proteins, and muscle cells’ cytoskeleton (Figure 4D). Cellular pathways were further analysed to infer their expression levels in patients compared to controls, which showed that the 3 mentioned pathways are downregulated in ARHGAP19 patients (Figure 4E).

### *In vitro* GAP assay shows loss of GAP activity in *ARHGAP19* variants in the GAP domain

ARHGAP19 has been previously reported for its RhoGAP activity. Interestingly, several mutations in *ARHGAP19* are clustering around the region encoding the GAP domain (Figure 1C). To investigate the GAP activity of these *ARHGAP19* variants, the GAP domain of ARHGAP19 or the mutated GAP proteins were expressed as GST fusion proteins in *E. coli* for *in vitro* GAP assays. Specific GAP activity toward RhoA was measured as the rate of inorganic phosphate released by GTPase-mediated GTP hydrolysis. RhoA alone showed little intrinsic GTPase activity, while the addition of wild type ARHGAP19 significantly accelerated the rate of RhoA-mediated GTP hydrolysis (Figure 5). Two ARHGAP19 missense mutations, p.Gly140Asp, and p.Gln151Lys, abrogated the GAP activity of ARHGAP19, decreasing the GTPase hydrolysis rate to the basal level. Another ARHGAP19 mutation, p.His196Gln*fs*\*9, which led to a truncated GAP domain of ARHGAP19, completely abolished the GAP activity as evidenced by severely impaired phosphate release (Figure 5A). Altogether, these results demonstrate that these mutations in the GAP domain of ARHGAP19 are dominant loss-of-function alleles, abolishing its GAP activity.

**Figure 5.**
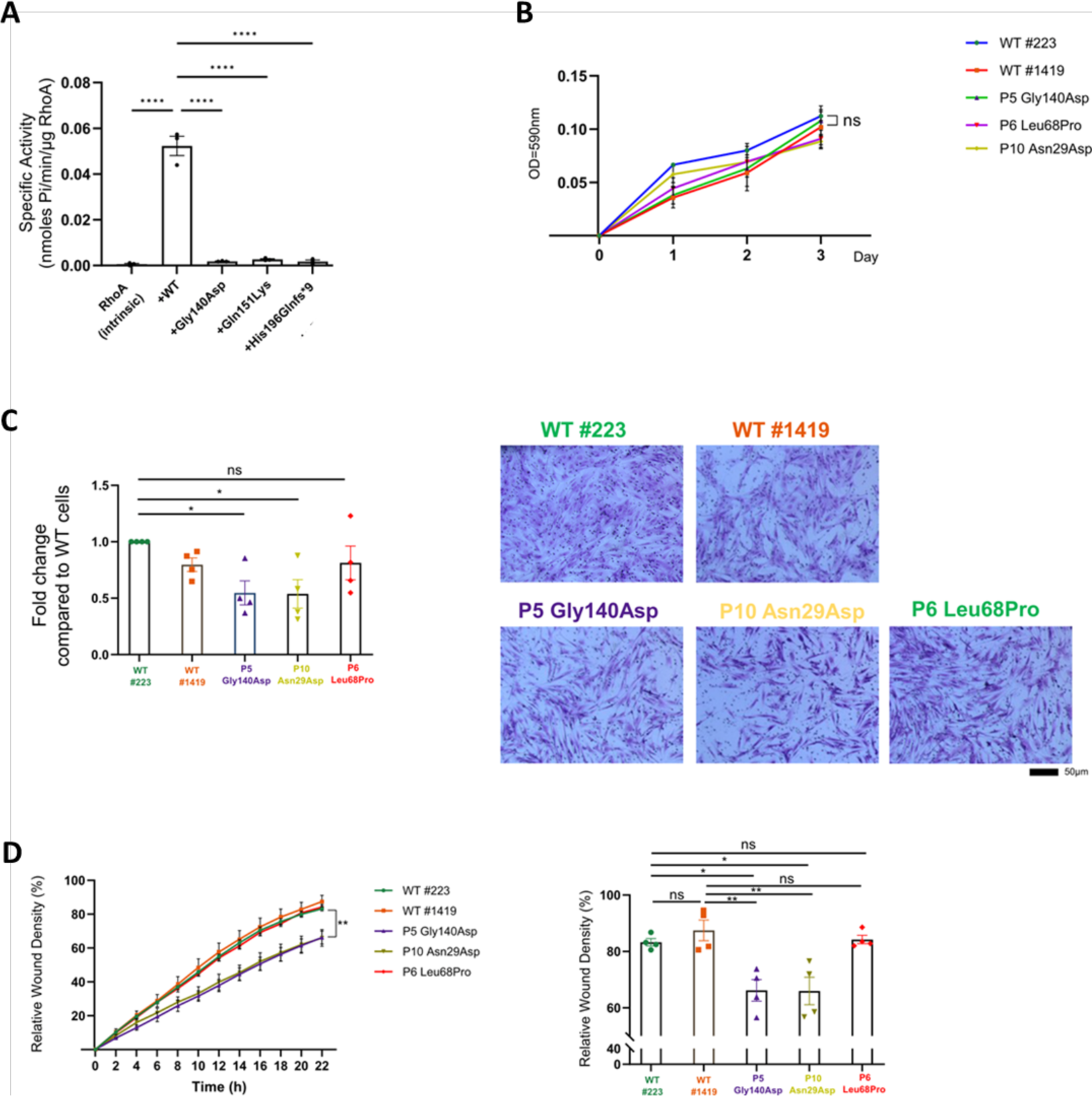
*ARHGAP19* variants have defective GAP-activity and cell migration. **A.** *In vitro* GAP activity assay measuring the GTPase rate of RhoA in the absence (intrinsic) or the presence of GAP domain from ARHGAP19 wild type (WT) or mutants. Data are presented as means ± SEM from three independent experiments (n=3; ****, p<0.0001; one-way ANOVA) **B.** MTT assays in fibroblasts from healthy controls (wt#223 and wt#1419) and ARHGAP19 mutants (n=3). **C and D.** Fibroblasts cells were subjected to the Boyden chamber migration assay (C) or wound healing scratch assay (D) as described in Materials and methods (n=4). Data are presented as means ± SEM (*p<0.05, **p<0.01, ns, not significant, one-way ANOVA)

### Patient-derived fibroblast cells demonstrate impaired cell migration

To further analyse the consequences of the *ARHGAP19* variants on cellular morphology, we derived fibroblasts from the patients of family 5 (c.419G>A), family 6 (c.203T>C) and family 10 (c.85A>G), paired with age/ gender matched health control (wild type). Since the *in vitro* GAP assay indicated that *ARHGAP19* c.419G>A (p.Gly140Asp) show defective GAP activity towards RhoA (Figure 5A), we hypothesized that variants in *ARHGAP19* may affect cell proliferation and migration. *ARHGAP19* variants and wild type fibroblasts showed no significant differences in cell proliferation (Figure 5B). However, patient-derived fibroblasts carrying the Gly140Asp and Asn29Asp mutations had a significant reduction in cell migration compared to control cells using both a Boyden chamber migration and wound healing assays (Figures 5C and D). In contrast, patient-derived fibroblasts harbouring the Leu68Pro showed no significant differences with control cells (Figures 5C and D). These results confirm the GAP defective function of the *ARHGAP19-* Gly140Asp mutant and suggest that the GAP activity of the *ARHGAP19*-Asn29Asp mutant may also be altered.

### The *ARHGAP19* ortholog *RhoGAP54D* promotes movement in *Drosophila*

To explore the consequences of ARHGAP19 loss of function *in vivo*, we first utilised the fruit fly, *Drosophila melanogaster*. The *Drosophila* genome contains a single *ARHGAP19* ortholog termed *RhoGAP54D*, which exhibits 51% amino-acid similarity and 31% identity to the human ARHGAP19 protein (Supplementary Figure 3). To examine the expression of *RhoGAP54D* in the *Drosophila* nervous system, we utilised a CRIMIC T2A-GAL4 enhancer trap in intron 3 of the RhoGAP54D locus (30), which we term *RhoGAP54D*^CRIMIC-Gal4^ and which results in expression of Gal4 in the pattern of the endogenous gene (Figure 6A). Driving expression of a UAS-*CD4::TdTomato* reporter via *RhoGAP54D*^CRIMIC-Gal4^ allowed us to label *RhoGAP54D*-positive cells in the adult fly brain and thoracic ganglion (Figure 6B). In agreement with previously published single-cell RNAseq data (31, 32) (Supplementary Figure 4A), this approach revealed sparse expression of *RhoGAP54D*, including projections to the antennal mechanosensory motor centre (AMMC) and isolated cell bodies within the brain and thoracic ganglion (Figure 6B). In addition, we noted RhoGAP54D-driven TdTomato signal surrounding the central and thoracic synaptic regions, suggesting that *RhoGAP54D* may be expressed in the perineural and/or subperineural glia (SPG) that form the blood-brain barrier covering the central and thoracic neuropil domains (Figure 6B) (33).

**Figure 6.**
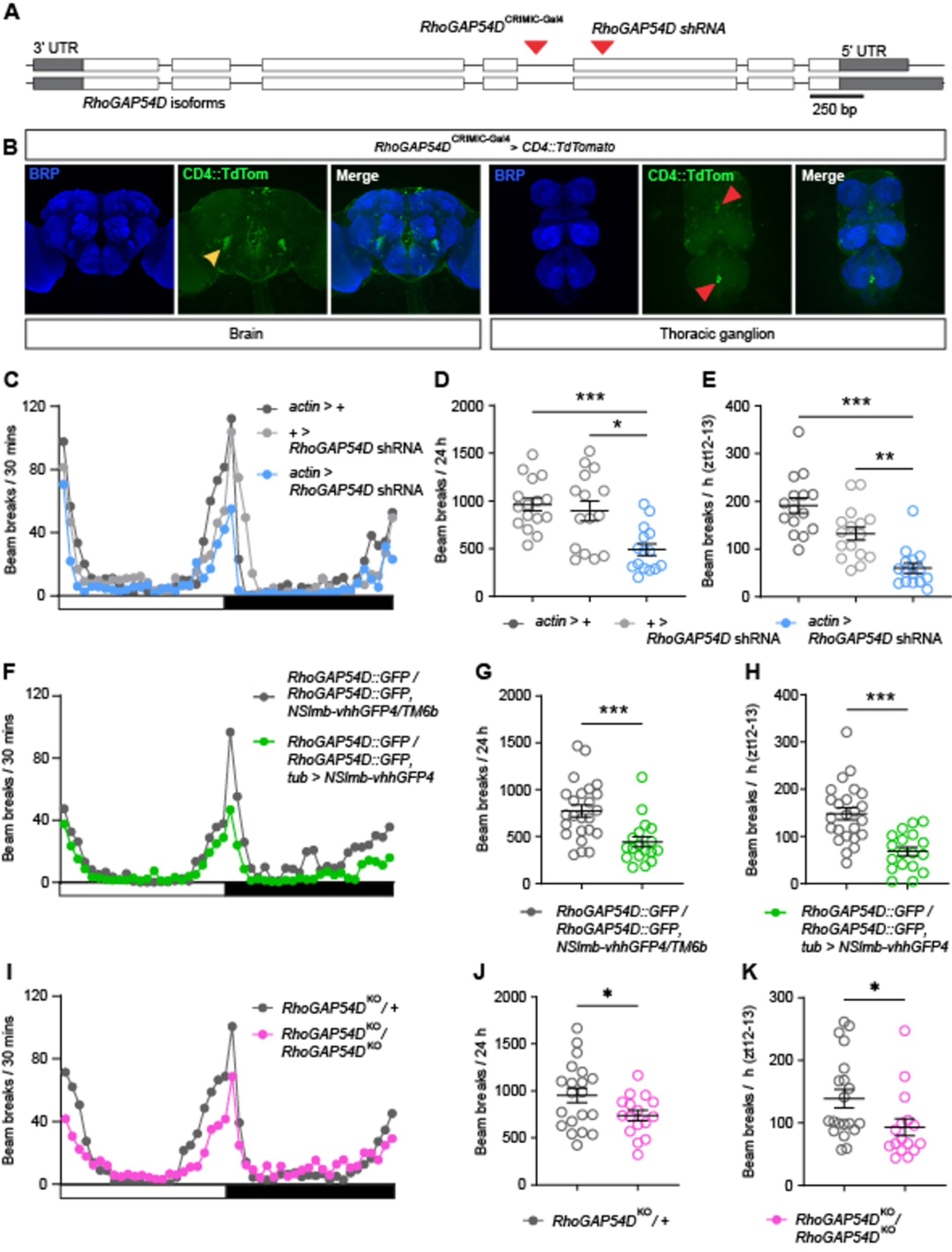
ARHGAP19 ortholog RhoGAP54D promotes movement in Drosophila melanogaster. **A.** Schematic of the *RhoGAP54D* locus. Exons – white blocks; untranslated regions (UTRs) – grey blocks; introns – line. Insertion site of the *RhoGAP54D*^CRIMIC-Gal4^ element and exonic region targeted by *RhoGAP54D* shRNA are shown (red arrows). **B.** Confocal images illustrating *RhoGAP54D*-driven membrane-tagged CD4::TdTomato expression in the adult male *Drosophila* brain and thoracic ganglion. Brain: yellow arrow points to projections close to the antennal mechanosensory motor center. Thoracic ganglion: red arrows point to isolated cell bodies. Note the CD4::TdTomato signal surrounding Bruchpilot (BRP)-labelled neuropil domains. **C.** Patterns of locomotor activity in flies globally expressing *RhoGAP54D* shRNA (*actin* > *RhoGAP54D* shRNA) and driver/shRNA alone controls. White bar: lights on; black bar: lights off. **D-E.** Number of beam breaks across 24 h (**D**) or during ZT12-13 (**E**), a period of peak activity. n = 15-16. **F-H.** Patterns of locomotor activity (**F**), total (**G**) and peak (ZT12-13; **H**) beam breaks in adult flies harbouring a *RhoGAP54D* GFP fusion allele and expressing deGradFP components, enabling degradation of the RhoGAP54D::GFP fusion protein, and a *RhoGAP54D*::GFP homozygote control. N = 18 and 24 respectively. **I-K.** Patterns of locomotor activity (**I**), total (**J**) and peak (ZT12-13; **K**) beam breaks in adult flies heterozygous or homozygous for the *RhoGAP54D*^KO^ null allele. N = 16 and 20 respectively. Central line in dot plots: mean. Error bars: standard error of the mean (SEM). * P < 0.05, **P < 0.005, ***P < 0.0005, one-way ANOVA with Dunnett’s post-hoc test (D), Kruskal-Wallis test with Dunn’s post-hoc test (E), Mann-Whitney U-test (G, K), or t-test with Welch’s correction (H, J)

Given that ARHGAP19 variants perturb movement in humans, we tested whether reducing RhoGAP54D expression similarly disrupted movement in *Drosophila*. Since RhoGAP54D is likely expressed in a variety of distinct cell-types, we used a global driver (*actin*-Gal4) in concert with an shRNA predicted to cleave *RhoGAP54D* mRNA in exon 3 of the primary transcript to induce ubiquitous RhoGAP54D knockdown (Figure 6A). Using quantitative PCR, we confirmed that expression of this shRNA caused an ∼ 50% reduction in *RhoGAP54D* mRNA expression (Supplementary Figure 4B). We then used the *Drosophila* Activity Monitor (DAM) system (25) to test how reducing RhoGAP54D expression affected locomotor activity in adult flies (Supplementary Figure 4C). In 12 h light: 12 h dark conditions, wild-type *Drosophila* exhibit crepuscular peaks of activity centred around lights-on and lights-off, interspersed by periods of low activity (Figure 6C). Global RhoGAP54D knockdown reduced both total activity occurring over 24 h and activity occurring in the hour following lights-off (zeitgeber time (ZT) 12-13), a measure of peak motor capacity (Figure 6C-E).

To complement the above data we utilised degradFP, a genetic system that promotes degradation of GFP-tagged fusion proteins via the ubiquitin pathway (34). Ubiquitous expression of degradGFP components in a background homozygote for a *RhoGAP54D::GFP* knock-in allele (22) similarly reduced overall and peak movement relative to *RhoGAP54D::GFP* homozygote controls (Figure 6F-H). As a final confirmation of the above results, we generated a *RhoGAP54D* null allele (*RhoGAP54D*^KO^) through CRISPR-Cas9 gene editing (Figure 6D). Comparison of *RhoGAP54D*^KO^ heterozygote and homozygote flies again revealed reduced overall and peak movement in *RhoGAP54D*^KO^ homozygotes relative to heterozygote controls (Figure 6I-K). Collectively, these data demonstrate that RhoGAP54D promotes robust locomotor activity in *Drosophila*, supporting the genetic link between variants in the human ortholog ARHGAP19 and disrupted movement.

### Danio rerio arhgap19 is important for motor neuron function

We next utilised zebrafish (*Danio rerio*) to examine the molecular, cellular, and developmental impact of ARHGAP19 loss of function in a vertebrate system. To analyse the endogenous expression pattern and subcellular localization of zebrafish *arhgap19* during development, we conducted whole-mount in situ hybridization (WISH) assays at three different embryonic stages, utilizing digoxigenin-labelled antisense RNA probes specific for *arhgap19*. ISH analyses revealed a ubiquitous expression pattern of arhgap19 across multiple brain regions, notably in the forebrain and hindbrain compartments. This expression was predominantly enriched within neural tissues at the 48 hpf. Specifically, heightened arhgap19 expression was observed in anatomically defined regions such as the cerebrum, thalamus, tuberculum, and tegmentum (Figure 7B). Intriguingly, a temporal downregulation of arhgap19 expression was evident as development progressed; by 5 dpf, the expression levels had substantially diminished (Figure 7B).

**Figure 7.**
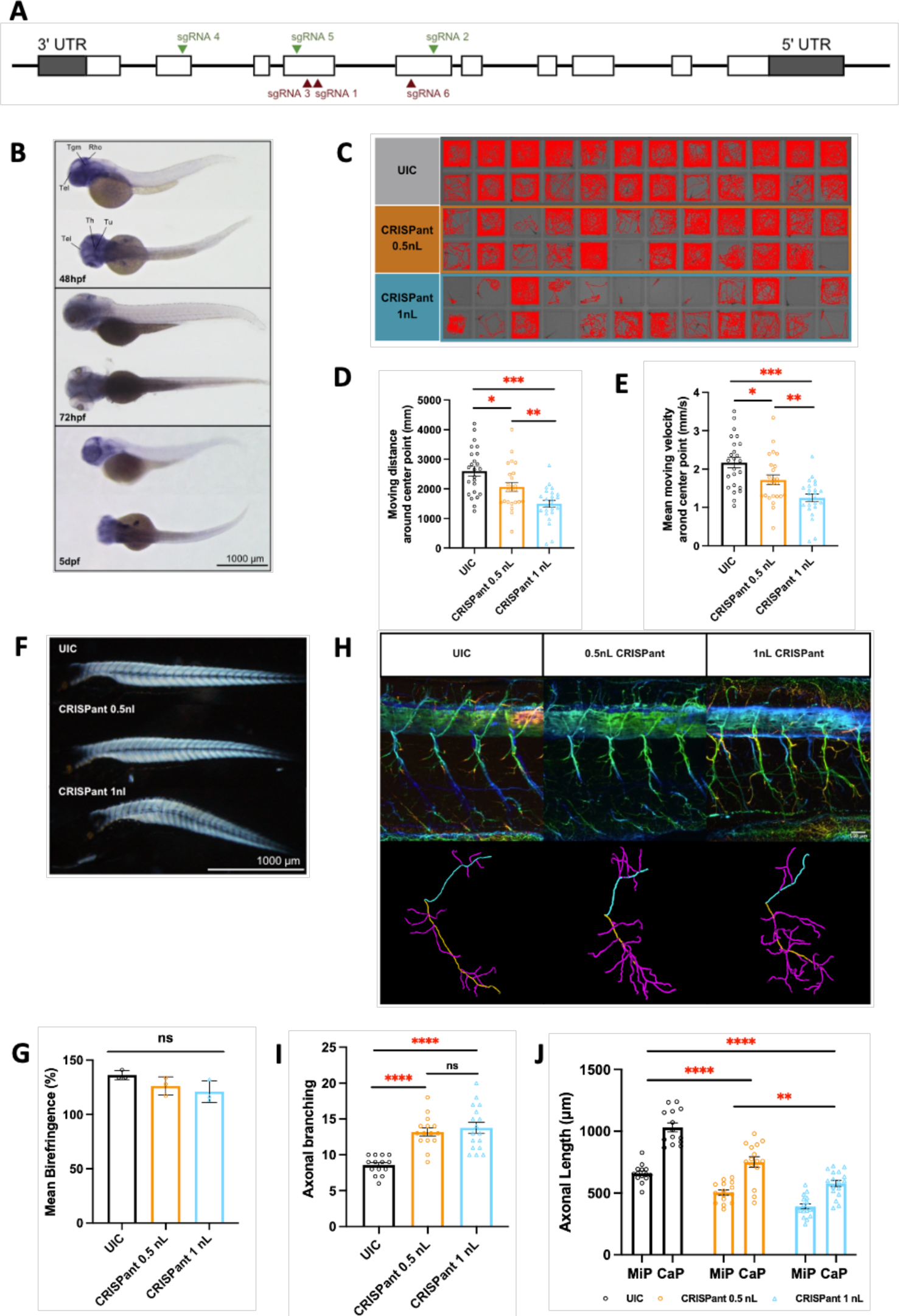
*Danio rerio arhgap19* is important for motor neuron function. **A.** Schematic representation of ten exons to cover the complete coding region. The position of the sgRNA targets are indicated (green illustrate the highest level of knockdown efficiency). **B.** *arhgap19* expression at three different embryonic stages in zebrafish. At 48 hpf, WISH signal of *arhag19* is localized in the forebrain and hindbrain regions; scale bar = 1000 μm. Rho: Rhombencephalon (hindbrain); Tel: Telencephalon; Th: Thalamus; Tu: Tuberculum; Tgm: Tegmentum. **C-E.** Behavior analysis of UIC and *arhgap19* mutant zebrafish larvae at 5-dpf. (C) reveals the swimming trajectories of each larva. Quantification of total travel distance (D) and travel velocity (E) of UIC and *arhgap19* mutant zebrafish larvae for 30 mins (UIC, CRISPant 0.5nL, CRISPant 1nL: n = 24). Each bar represents mean (± SEM). Asterisks above the bars indicate significant difference (*P ≤ 0.05, **P ≤ 0.01, ***P ≤ 0.001). **F-G.** UIC and *arhgap19* mutant zebrafish larvae at 5-dpf were evaluated for muscle integrity using birefringence. (F) A representative image of one larva from each treatment group; scale bar = 1000 μm. Each bar in plot (G) represents average birefringence (± SEM) for all zebrafish larvae (UIC, CRISPant 0.5nL, CRISPant 1nL : n = 3). **H-J.** Spinal motor neurons morphogenesis defects in *arhgap19* mutant zebrafish larvae. (H) Confocal imaging analysis and three-dimensional reconstruction of spinal motor neurons in UIC and *arhgap19* mutant groups at 5-dpf; scale bar = 100 μm. (I) Axonal branching number of Cap axons in UIC and *arhgap19* mutant zebrafish larvae. (J) Average axonal length of Cap (yellow) and Mip (blue) axons in UIC and *arhgap19* mutant zebrafish larvae. Each bar represents mean (± SEM). Asterisks above the bars indicate significant difference (*P ≤ 0.05, **P ≤ 0.01, ***P ≤ 0.001, ****P ≤ 0.0001).

To investigate the functional role of Arhgap19 in neuronal and motor development, we generated an F0 biallelic knockout mutant model utilizing CRISPR/Cas9 system. Genomic deletions were introduced at three selected loci within the zebrafish arhgap19 gene—exons 2, 4, and 5—informed by considerations of targeting efficiency and potential off-target effects (Figure 7A). Of the initial batch of 62 eggs, 32 (51.6%) were successfully fertilized. Embryos were monitored on a daily basis, with dead ones discarded. Those that survived were sacrificed at 5 dpf for genotyping and subsequent analysis. NGS sequencing showed that sgRNAs 2, 4, and 5 manifested high gene-editing efficiency, with respective average modification rates of 96.1%, 99.7% and 94.6%. Conversely, sgRNA 3 exhibited suboptimal performance, achieving a comparatively low modification rate of merely 11%.

To evaluate the changes in behaviour in the arhgap19 mutant model, we collected 24 zebrafish larvae at the 5 dpf and analysed their motor activity. Metrics such as the total duration of movement, aggregate distance traversed, and mean velocity were measured. Behavioural assays demonstrated that *arhgap19* knockout induced conspicuous motor deficits. Larvae in both mutant (CRISPants) groups exhibited decreased motor activity, alongside idiosyncratic and involuntary movements. In contrast, larvae from the Uninjected Control (UIC) group exhibited normal locomotor behaviour, exploring the well’s periphery (Figure 7C).

Statistical analysis revealed significant discrepancy in motor parameters among the groups. Specifically, larvae from both CRISPants groups were significantly different in the total travel distance (Figure 7D; one-way ANOVA, F(2, 69) = 13.954, and p < 0.0001, followed by Welch T test. CRISPant 0.5nL, t(45.82) = −2.376 and p = .022; CRISPant 1nL, t(41.939) = −5.418 and p < 0.0001). Likewise, the mean velocity also showed significant differences between mutant and control groups (Figure 5E; one-way ANOVA, F(2, 69) = 13.954, and p < 0.0001, followed by Welch T test. CRISPant 0.5nL, t(45.82) = −2.376 and p = .022; CRISPant 1nL, t(41.939) = −5.418 and p < 0.0001). Larvae from the CRISPants 1nL group swam approximately twice as slow compared to those from the UIC group.

To investigate the impact of arhgap19 knockout on muscular architecture, we quantitatively assessed the birefringence intensity of zebrafish skeletal muscle at the 5-dpf, employing a polarizing light stereomicroscope for imaging. Statistical analysis of the birefringence levels revealed no significant difference between the control group and the three knockout groups (Figure 7G; one-way ANOVA, F(2, 8) = 2.904, p=0.131). These findings strongly suggest that the skeletal muscle integrity remains largely intact in the absence of arhgap19. Consequently, the motor deficits observed in the behavioural analyses are more likely attributed to impairments in motor neuron function rather than muscular deficiencies.

To investigate the effects of *arhgap19* knockout on primary motor neurons, we employed immunostaining techniques complemented by confocal microscopy for visualization. Notably, a more robust axonal bundle was observed in both CRISPant groups (Figure 7H). Quantitative assessments were carried out to analyze both axonal length and branching complexity. In *arhgap19* CRISPant larvae, the branching density of the Caudal Primary (CaP) Motorneurons was higher than that of the control group (Figure 5I; one-way ANOVA, F(2, 43) = 21.341 and p < 0.0001, followed by Welch T test with Bonferroni’s adjustment. CRISPant 1nL, t(22.271) = 6.33 and ***p < 0.0001). Moreover, axonal length was markedly affected in *arhgap19* CRISPants. Statistical analysis revealed a notable reduction in the average length of CaP and Middle Primary (MiP) motorneurons in 1nL injected CRISPants, measuring 577.2 μm and 391.2 μm, respectively, in contrast to the control values of 1012.5 μm and 639.8 μm. (Figure 7J; one-way ANOVA, F(2, 43) = 41.936 and p < 0.0001, followed by Welch T test with Bonferroni’s adjustment. CRISPant 0.5nL, t(26.647) = −4.943 and ***p < 0.0001; CRISPant 1nL, t(24.101) = −10.092 and ***p < 0.0001) Particularly, the axons of CaPs in *arhgap19* CRISPants exhibited diminished length which failed to reach the ventral musculatures. Furthermore, embryos injected with *arhgap19* splice site and translation blocking morpholinos also exhibited branching abnormalities similar to those observed in CRISPants mutants (Supplementary Figure 5). Taken together, these results indicated that impaired motor neuron developmental were attributable to the loss of *arhgap19*.

## Discussion

We show that biallelic *ARHGAP19* variants are a novel cause of inherited early-onset neuropathy. We identified 25 individuals harbouring missense and nonsense variants both within the functional GAP domain as well as outside this structural domain of ARHGAP19. The patients had a motor-predominant neuropathy with AAO almost exclusively in the first two decades of life. The clinical phenotype was generally length-dependent, but there are some unusual features including frequency conduction slowing +/− conduction block, prominent asymmetry, in some individuals relatively rapid progression and upper limb onset. There was no observable genotype-phenotype correlation at the first examination; mean AAO of variants in GAP versus non-GAP domain 9.0 vs 11.6 years (*p* = 0.32, n = 15 and 8 respectively) and mean ulnar motor conduction velocity 41.0 vs 44.8 m/s (*p* = 0.62 n = 5 and 5). However, longitudinal studies would help better delineate the disease progression and any genotype-phenotype correlation.

Haplotype analysis using genetic data from our cohort as well as from control databases suggested that p.His196Gln*fs*\*9 is unlikely to be from a recent common ancestor and possibly suggests that two independent *ARHGAP19* mutational events within Arabian Middle Eastern populations. Moreover, p.Gln151Lys and p.Leu68Pro variants are founder effect variants, probably originating in Turkey given the ethnicity of patients. Variant p.Gln151Lys which affects a highly conserved residue within the GAP domain, could have a significant structural or functional role. Additionally, this variant affects the same N-terminal catalytic stretch where the arginine motif at codon 143 is found, and therefore might have a comparable mechanistic effect as do other *AHRGAP19* variants identified nearby, i.e. p.Gly140Asp, p.Leu141Trp. Together with p.Leu228His and p.Asn239Lys, they are predicted to disrupt the domain’s structure and subsequently its function as a GTPase-activating protein in a variety of cellular processes (3, 35). Similarly, previously reported variants in the Rho-GAP domain of Myo9b, abrogate GAP activity (36). Interestingly, the corresponding *MYO9B* gene has recently been associated with a subtype of CMT2 and isolated optic atrophy (37). Other variants which increase free energy, such as p.Gln151Lys, may cause a deleterious effect to protein folding or stability through other mechanism. We note that free energy changes were calculated using the AlphaFold2-derived ARHGAP19 protein. A recent study concluded that AlphaFold is not immediately applicable when calculating free energy changes for predicted protein structures (38). However, results were based on the first iteration of AlphaFold. Conversely, another study showed that free energy changes using AlphaFold2-predicted protein models consistently matched those of experimentally determined structures, particularly in high confidence regions (pLDDT > 90) (13). Thus, until an experimental ARHGAP19 protein structure is generated, it is important that *in silico* investigations of variant impact are complemented by corresponding functional studies.

The phenotype of conduction slowing in a motor(-predominant) neuropathy is unusual in CMT, limited to very few genes. Notably, recessive variants in the GEF gene *PLEKHG5* cause this phenotype and have been showed to cause both loss of large, myelinated fibres and thin myelination seen on nerve biopsy, and motor axonal degeneration in a knockout mouse model (39, 40). Combined with the conduction slowing seen with biallelic variants in *ARHGEF10,* mechanistically this would suggest that Rho GTPase activity, mediated by these GEF and GAP proteins, is implicated in a process that may involve both myelin and axonal pathologies.

Data derived from three global *in vivo* loss-of-function *Drosophila* models and three CRISPR-Cas9 mediated knockout zebrafish models demonstrate a conserved role for ARGHAP19 orthologs in promoting locomotor activity across metazoan species, supporting the above genotype-phenotype correlations in human patients harbouring *ARGHAP19* mutations. We observed more pronounced axonal bundles and a significantly increased number of axonal branches in all experimental mutant zebrafish groups, suggesting that ARGHAP19 LOF may cause CMT in part by perturbing the cytoskeleton of motoneurons. This hypothesis is consistent with previous reports of RhoA playing a critical role in modulating cytoskeletal dynamics, including the mediation of stress fibre formation and focal adhesions(41), and facilitating axonal outgrowth and branching (42). Further transcriptomics analysis provided evidence that pathways associated with motor proteins, cell cycle and muscular cytoskeleton are downregulated, further explaining that clinical features observed in our patients might be due to alteration in these physiological pathways. Of note, there are some upregulated cellular functions in the patients, including the ferroptosis and cytokine-cytokine receptor interactions.

How might *ARHGAP19* mutations perturb the cytoskeletal network in motor neurons? In this work, the pathogenicity of *ARHGAP19* variants p.Gly140Asp, p.Gln151Lys and p.His196Gln*fs*\*9 are supported by *in vitro* GAP activity assays which show that variants within GAP domain cause complete loss of GAP activity. ARHGAP19 stimulates the intrinsic low GTPase activity of RhoA thereby negatively regulating the RhoA/ROCK pathway. Variants causing loss of GAP activity of ARHGAP19 may therefore cause overactivity of RhoA with further consequences in downstream effectors such as ROCK activation which has important functions in actin organisation, cell migration, and axon outgrowth and guidance. Importantly, this is in line with *in vitro* data from patient-derived fibroblasts harbouring p.Asn29Asp and p. Gly140Asp variants, that show significant decrease in cell migration using cell migration assays. Together, these results reveal that GAP defective *ARHGAP19* variants lead to altered RhoA activity thereby altering cell migration and cytoskeletal dynamics. Interestingly, protein expression analysis by Western blot revealed significant reduction in ARHGAP19 in iPSC motor neurons of patients compared to controls. Taken together, our findings are consistent with the patients’ phenotype and functional assays, explaining that the muscular and motor defects observed in patients as well as animal models could be due to the dysregulation of the ARHGAP19 related signalling cascade. We provide evidence for loss-of-function of ARHGAP19 as the pathomechanism of the disease and suggest that the loss of function might be more robust in motor neurons.

Of note, variant p.Leu68Pro showed no change in the migration and wound healing assays as opposed to WT controls. Interestingly, the two patients with this variant in our cohort were first suspected to have an autoimmune cause of neuropathy and were put on intravenous Ig for treatment of presumed CIDP with no improvement. These patients also have an upper limb involvement, and it is noteworthy that even if the variant lies outside the GAP domain, it is predicted pathogenic using various *in silico* methods.

Our findings do not fully elucidate the mechanism of axonal damage caused by ARHGAP19 deficiency. Regarding the animal models created in this study, we were not able to investigate the expression of RhoGAP54d in the fly peripheral nervous system and the zebrafish model showed no expression of the protein ortholog in its periphery. On the basis of frequent conduction slowing and the early-onset phenotype observed, it will therefore be important to examine peripheral myelination in the human subjects, as well as use better techniques for spatial localization and quantitative expression in models of ARHGAP19 deficiency.

Despite the theoretically simple on/off switch model of Rho, the Rho GTPase signalling pathway has a more sophisticated picture. The high number of GAP and GEF proteins – 66 and 80 respectively, which outnumber Rho proteins, coupled with unclear specificity of the proteins, means that understating their signalling activities as well as role in disease, remain challenging. Nevertheless, overactive RhoA signalling in neurons, be it due to genetic variations or imbalance between signalling molecules, has been reported in Charcot-Marie-Tooth disease. It is increasingly emerging that this is an important pathway not only in neuronal health but also in disease. Hence, modulation of this pathway may represent a potential strategy for future therapeutic treatments.

## Data Availability

All data produced in the present study are available upon reasonable request to the authors

https://databases.lovd.nl/shared/variants/ARHGAP19?search_var_status=%3D%22Marked%22%7C%3D%22Public%22

## Acknowledgments

We thank the patient and relatives for consent to be part of the study as well as the clinicians for helping with patient phenotyping. The families were collected as part of the SYNaPS Study Group collaboration funded by The Wellcome Trust and strategic award (Synaptopathies) funding (WT093205 MA and WT104033AIA) and research was conducted as part of the Queen Square Genomics group at University College London, supported by the National Institute for Health Research University College London Hospitals Biomedical Research Centre. This work was partly supported by an MRC strategic award to establish an International Centre for Genomic Medicine in Neuromuscular Diseases (ICGNMD) MR/S005021/1 and ND, SE, CR, PT, MGH, MMR and HH received direct support from this award. JP is supported by Medical Research Future Fund (MRFF) Genomics Health Futures Mission (APP2007681) and by the Australian Government Research Training Program. LVdV is supported by a predoctoral fellowship of the Research Fund - Flanders (FWO) under grant agreement N°11F0921N. TS is member of the European Reference Network for Rare Neuromuscular Diseases (ERN EURO-NMD). JB is supported by a Senior Clinical Researcher mandate of the Research Fund - Flanders (FWO) under grant agreement N°1805021N. ANB gratefully acknowledges the support of SVIKV and the use of the services and facilities of the Koç University Research Center for Translational Medicine (KUTTAM), funded by the Presidency of Turkey, Head of Strategy and Budget. We thank collaborator Simon Bullock (University of Cambridge) for plasmids, and Florencia di Pietro for characterization of the RhoGAP54D loss of function allele. JECJ was funded by an MRC Senior Non-Clinical Fellowship (MR/V03118X/1). Work in the Bellaïche group is funded by the CNRS, the INSERM and the Institut Curie as well as by the ANR (TiMecaDiv 20CE13000801) grant. TBH was supported by the Deutsche Forschungsgemeinschaft (DFG, German Research Foundation – 418081722, 433158657), and the European Commission (Recon4IMD - GAP-101080997). This work was supported in part by the Fund for Scientific Research (FWO-Flanders) (research grants G048220N and G0A2122N to A.J.), the Research Fund of the University of Antwerp (doctoral grant to C.A.), the Association Belge contre les Maladies Neuromusculaires’ (ABMM-Telethon) (research grantsto A.J.), the French Muscular Dystrophy Association (AFM-Telethon, research grant 23708 to A.J.). JPark was supported by the Clinician Scientist program “PRECISE.net” funded by the Else Kröner-Fresenius-Stiftung. Work in the Lamarche-Vane group funded by Natural Sciences and Engineering Research Council of Canada grant RGPIN/04809-2017 and CIHR project grant PJT-180367. Leif Leclaire holds a CIHR master studentship. RH is supported by the Wellcome Discovery Award (226653/Z/22/Z), the Medical Research Council (UK) (MR/V009346/1), the Addenbrookes Charitable Trust (G100142), the Evelyn Trust, the Stoneygate Trust, the Lily Foundation, Ataxia UK, Action for AT, the Muscular Dystrophy UK. This research was supported by the NIHR Cambridge Biomedical Research Centre (BRC-1215-20014). The views expressed are those of the authors and not necessarily those of the NIHR or the Department of Health and Social Care.We are also grateful to Queen Square Genomics at the Institute of Neurology University College London, supported by the National Institute for Health Research University College London Hospitals Biomedical Research Centre, for the bioinformatics support.

## Online resources

CPDB web tool, http://cpdb.molgen.mpg.de/

Ensembl, http://www.ensembl.org/i

FastQC, http://www.bioinformatics.babraham.ac.uk/projects/fastqc/

GATK documentation, https://software.broadinstitute.org/gatk/

Human Genome Variation Society, http://www.hgvs.org

Mutation Taster version 2, http://www.mutationtaster.org/

PolyPhen-2, http://genetics.bwh.harvard.edu/pph2/

SIFT, http://sift.jcvi.org/

University of California—San Francisco (UCSC) Genome Browser, https://genome.ucsc.edu/

Jalview, https://www.jalview.org/

UniProt, https://www.uniprot.org/

PyMol, https://pymol.org/2/

AlphaFold Protein Structure Database, https://alphafold.ebi.ac.uk/

FoldX, https://foldxsuite.crg.eu/

DIOPT http://flybase.org/reports/FBgn0034249.html

