## Supplementary Figure 1 for "Biallelic variants in *ARHGAP19* cause a motor-predominant neuropathy with asymmetry and conduction slowing"

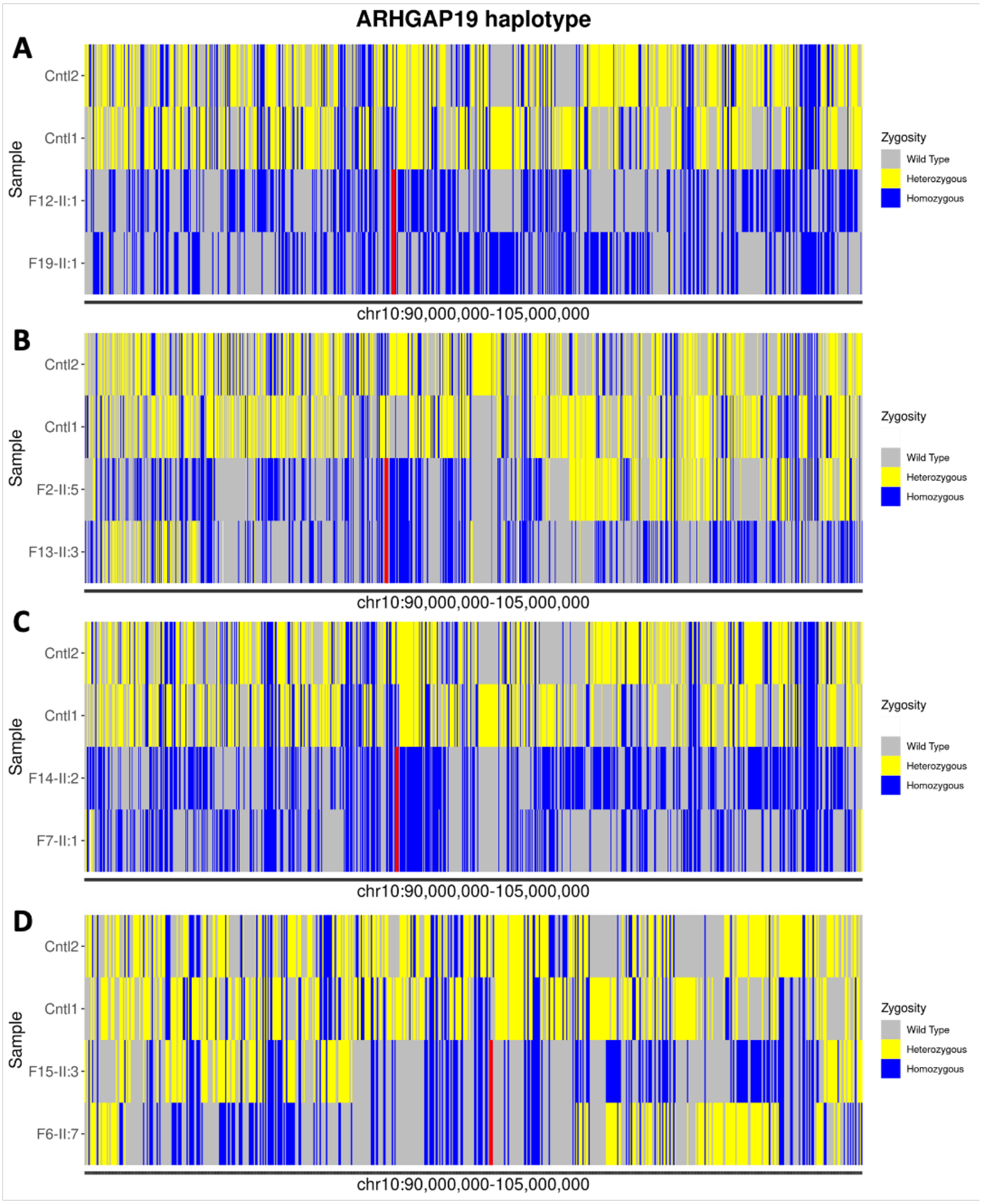


**Supplementary Figure 1. Haplotype analysis of *ARHGAP19* variants A.** chr10-97259559-A-T (p.Leu228His) **B.** chr10:97263447-G-GT (p.His196Gln*fs**9) **C.** chr10-97263582-G-T (p.Gln151Lys) and **D.** chr10-97265979-A-G (p.Leu68Pro). Variations pattern of flanking regions of shared pathogenic variants was coloured as follows: homozygous variants as blue, heterozygous variants as yellow, and WT locus as grey. The position of the pathogenic variant is highlighted in red.
