## Supplementary Figure 2 for "Biallelic variants in *ARHGAP19* cause a motor-predominant neuropathy with asymmetry and conduction slowing"

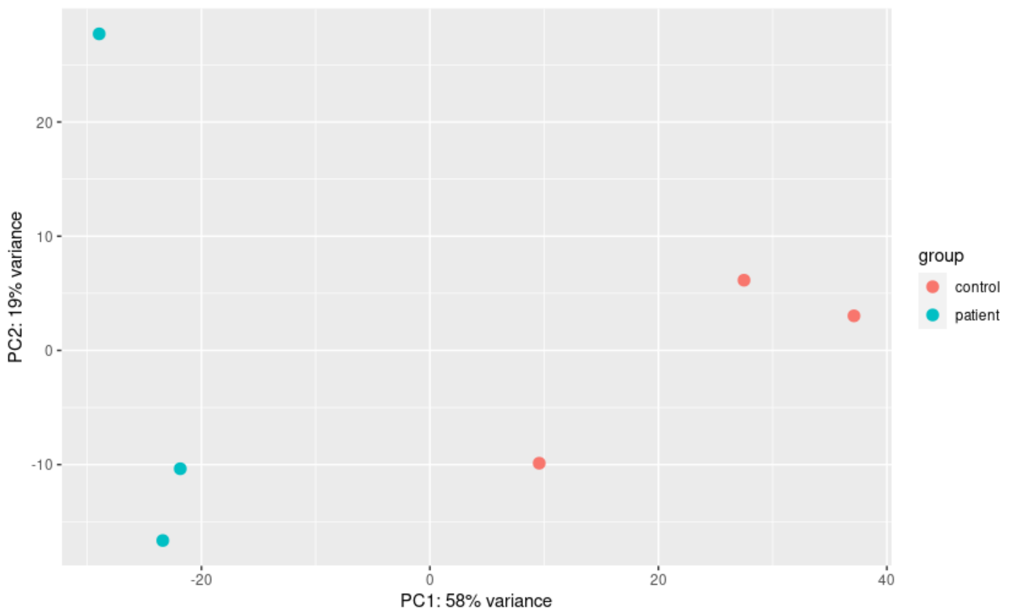


**Supplementary Figure 2. RNA-Seq principal component analysis (PCA) plot.** Differential expression analysis was done on 3 patients along with 3 controls, showing a separation between their RNA expression patterns.
