## Supplementary Figure 3 for "Biallelic variants in *ARHGAP19* cause a motor-predominant neuropathy with asymmetry and conduction slowing"

```
  Fly   113 LMHLQELKEFLMLEKNLTQEGLFRKAGAVSRQNELRMHIQHDKPLNLELAGFSAHDCATVFKGFL 177
            :..:.:|.|:  |.|||..|||||..|...||..||..:.:...::||...|.::|.||:.|.||
Human   122 IAQIYQLIEY--LHKNLRVEGLFRVPGNSVRQQILRDALNNGTDIDLESGEFHSNDVATLLKMFL 184

  Fly   178 SELPEPLLTDAHYPAHLQIAPLCQALN-GQTTATAERQQHLLNSVQLLLLLLPEEHRELLQHIIE 241
            .||||||||..|:.|||:||.|.|..: |..|...::.:. :.::|||.|:||..:|.||:.:::
Human   185 GELPEPLLTHKHFNAHLKIADLMQFDDKGNKTNIPDKDRQ-IEALQLLFLILPPPNRNLLKLLLD 248

  Fly   242 MLHAVAKHEKSNKMSADNLATLFTPHLICPRQLPPEVLHYQAKKMSSIVTYMIVRGLDIFEVPGK 306
            :|:..||.:..|||||.|||.:|.||::.|:.:....|.....|::|.:.:||.....:|:.|..
Human   249 LLYQTAKKQDKNKMSAYNLALMFAPHVLWPKNVTANDLQENITKLNSGMAFMIKHSQKLFKAPAY 313

  Fly   307 LSTDIRAYFLERKRKKTMSPEQTLDESISDVSTVNTVYTFVDRAATAAATN-----------TNN 360
            :....|.::|..:.:          .|..|:..:.:.:|...:.|.:...|           .::
Human   314 IRECARLHYLGSRTQ----------ASKDDLDLIASCHTKSFQLAKSQKRNRVDSCPHQEETQHH 368

  Fly   361 TDTELAQLYAHIQSMPESSKKRRLIKQFNKQNGQGTPLQLVVMNRLKNNEATRSAKSLGDSIKKH 425
            |:..|.:|:.|:..||||:||::||:|||||:...||.:....::::....:||...|   ||:.
Human   369 TEEALRELFQHVHDMPESAKKKQLIRQFNKQSLTQTPGREPSTSQVQKRARSRSFSGL---IKRK 430

  Fly   426 IFHKSLMSRTPKRVPPSFHLASGSETPNMSHVKPPKMRVLFQ-SP-TPPTPT 475
            :....:||...|:.|....:|.|    .:.........:||. || ...|||
Human   431 VLGNQMMSEKKKKNPTPESVAIG----ELKGTSKENRNLLFSGSPAVTMTPT 478
```

**Supplementary Figure 3**. Protein Alignment of orthologs RhoGAP54D (fly; NP_001261060.1) and ARHGAP19 (human; NP_NP_116289.4) using DIOPT ortholog prediction tool Version 9. *Drosophila* RhoGAP54D and human ARHGAP19 share 51% similarity and 31% amino-acid identity.
