## Supplementary Figure 4 for "Biallelic variants in *ARHGAP19* cause a motor-predominant neuropathy with asymmetry and conduction slowing"

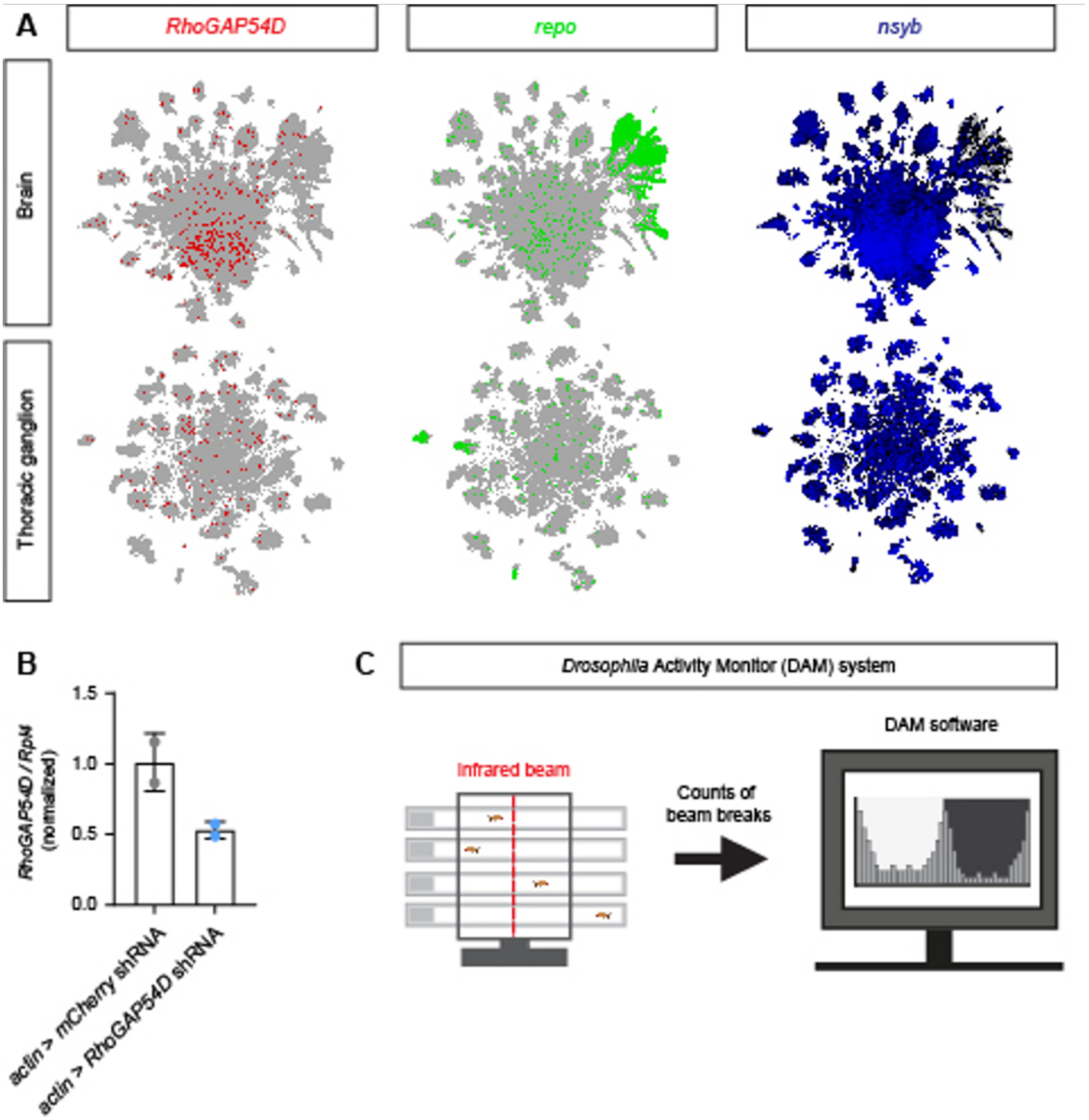


**Supplementary Figure 4. A.** Visualisation of single-cell RNAseq data from the adult *Drosophila* central nervous system (above) and thoracic ganglion (below) using SCOPE (28, 29). *RhoGAP54D* expression is compared to the post-mitotic glial and neuronal markers *repo* and neuronal synaptobrevin (*nsyb*). **B**. Validation of *RhoGAP54D* knockdown via quantitative PCR. *RhoGAP54D* expression was normalised to the *Rpl4* house-keeping gene. **C.** Schematic of the DAM system. Flies housed individually in glass tubes are shown crossing a central infrared beam, allowing the number of beams breaks over time to be used as a readout of temporal changes in locomotor activity.
