## Supplementary Figure 5 for "Biallelic variants in *ARHGAP19* cause a motor-predominant neuropathy with asymmetry and conduction slowing"

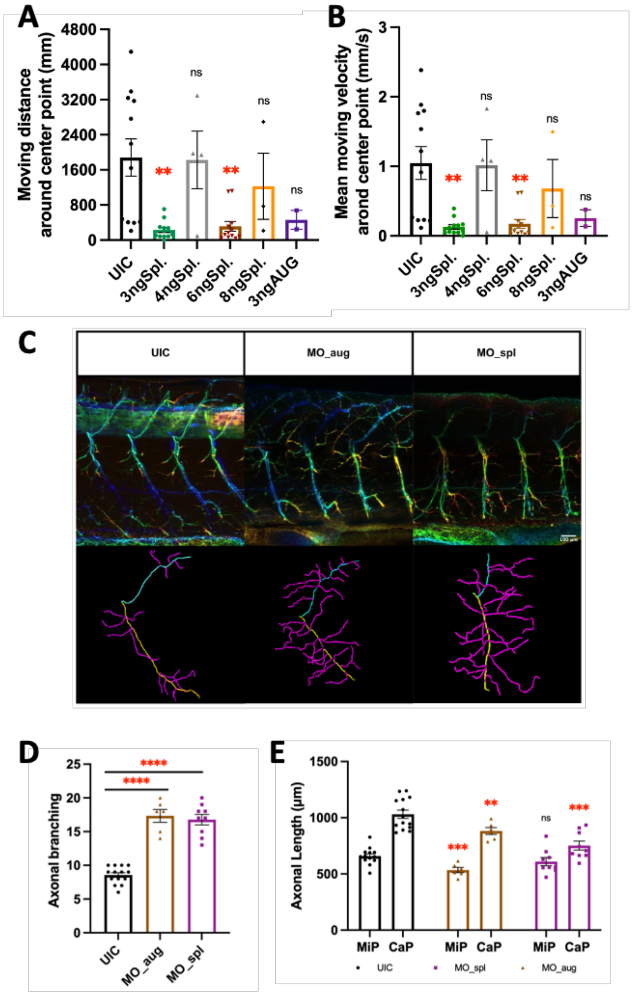


**Supplementary Figure 5. A-B. Behavior analysis of UIC and morpholino-injected mutants (MOs) zebrafish larvae at 5-dpf:** Quantification of total travel distance (A) and travel velocity (B) of UIC and MOs zebrafish larvae for 30 mins (3ngSpl_MO & 6ngSpl_MO: n = 12; 4ngSpl_MO n = 4; 8ngSpl_MO n = 3; 3ngAUG_MO n = 2). Each bar represents mean (± SEM). Asterisks above the bars indicate significant difference (*P ≤ 0.05, **P ≤ 0.01, ***P ≤ 0.001, ****P ≤ 0.0001). **C-E. Spinal motor neurons morphogenesis defects in MOs mutant zebrafish larvae.** (C) Confocal imaging analysis and three-dimensional reconstruction of spinal motor neurons in MOs mutant larvae at 5-dpf; scale bar = 100 µm. (D) Axonal branching (magenta) number of Cap axons (yellow) in UIC and MOs mutant zebrafish larvae. (E) Average axonal length of Cap (yellow) and Mip (blue) axons in UIC and MOs mutant zebrafish larvae. Each bar represents mean (± SEM). Asterisks above the bars indicate significant difference (*P ≤ 0.05, **P ≤ 0.01, ***P ≤ 0.001, ****P ≤ 0.0001).
