## Supplementary Table 1 for "Biallelic variants in *ARHGAP19* cause a motor-predominant neuropathy with asymmetry and conduction slowing"

| Family | 1 | 3 | 4 | 5 | 6 | 8 | 9a | 9b | 10 | 11 | 13 | 15 | 16 | 18 | 19 |
|---|---|---|---|---|---|---|---|---|---|---|---|---|---|---|---|
| **Case** | PT1 (F1-II:1) | PT3 (F3-II:1) | PT4 (F4-II:1) | PT5 (F5-II:2) | PT6 (F6-II:7) | PT8 (F8-II:2) | PT9 (F9-II:3) | PT9 (F9-II:4) | PT11 (F10-II:1) | PT12 (F11-II:2) | PT14 (F13-II:3) | PT17 (F15-II:2) | PT18 (F16-II:1) | PT20 (F18-II:1) | PT21 (F19-II:1) |
| **Phenotype** | CMT-MP | dHMN with CS | CMT2-MP | dHMN | dHMN with CS | CMT2-MP | CMT2 with CS | CMTi | CMT2-MP | CMT2-MP with CS | CMT | CMTi-MP | dHMN | HMN with CS | CMT2-MP |
| **Age at study (years)** | 6-10 | 16-20 | 11-15 | 46-50 | 16-20 | n/a | 16-20 | 11-15 | 46-60 | 11-15 | n/a | 11-15** | 11-15 | 6-10 | 16-20 |
| **Median CMAP mV** | ND | 12.3 | 8.2 | 4.4 | 8.3 | 0.2$^{\Delta}$ | ND | ND | 6.5 | 11.7 | ND | 3.7 | ND | 3.4 | 11.9 |
| **Median DML ms** | ND | 3.8 | 3.7 | 6.9 | 4.0 | 5.5$^{\Delta}$ | ND | ND | 3.6 | 6.6 | ND | 4.2 | ND | 3.6 | 2.8 |
| **Median NCV m/s** | ND | 40 | 56 | ND | 51 | 54$^{\Delta}$ | ND | ND | 56 | 42 | ND | 29 | ND | 33 | 54 |
| **Ulnar CMAP mV** | ND | 5.2 | ND | 7.6 | 6.0 | 1.7$^{\Delta}$ | 13.3 | 3.0 | 6.5 | 13.9 | 1.4 | 3.8 | ND | ND | 4.7 |
| **Ulnar DML ms** | ND | 3.2 | ND | 3.6 | 2.7 | 4.0$^{\Delta}$ | 2.5 | 3.0 | 2.7 | 4.3 | ND | 2.7 | ND | ND | 1.78 |
| **Ulnar NCV m/s** | ND | 40 | ND | 47 | 56 | 35$^{\Delta}$ | 49 | 25 | 58 | 47 | 39 | 28 | ND | ND | 45 |
| **Peronel (TA) CMAP mV** | 3.2 | 3.6 | ND | 0.2 | 2.5 | ND | ND | ND | ND | 1.3 | ND | ND | ND | ND | 0.37 |
| **Peronel (TA) DML ms** | 4.0 | 3 | ND | 5.2 | 5.8 | ND | ND | ND | ND | 6.6 | ND | ND | ND | ND | 3.8 |
| **Peronel (TA) NCV m/s** | 30 | 45 | ND | ND | 15 | ND | ND | ND | ND | 22 | ND | ND | ND | ND | 68 |
| **Peroneal (EDB) CMAP mV** | Absent | Absent | Absent | Absent | ND | Absent | Absent | Absent | 1.1 | Absent | ND | 0.7 | Absent | 0.87 | 0.04 |
| **Peroneal (EDB) DML ms** | Absent | Absent | Absent | Absent | ND | Absent | Absent | Absent | 5.4 | Absent | ND | ND | Absent | 9.24 | 5.2 |
| **Peroneal (EDB) NCV m/s** | Absent | Absent | Absent | Absent | ND | Absent | Absent | Absent | 33 | Absent | ND | 22 | Absent | ND | ND |
| **Tibial (AH) CMAP mV** | Absent | 1.7 | Absent | Absent | ND | 0.2 | 0.6 | Absent | ND | Absent | ND | 3.6 | Absent | ND | 0.38 |
| **Tibial (AH) DML ms** | Absent | 4.7 | Absent | Absent | ND | 4.1 | 6.6 | Absent | ND | Absent | ND | 6.6 | Absent | ND | 3.7 |
| **Tibial (AH) NCV m/s** | Absent | 19 | Absent | Absent | ND | 22 | 22 | Absent | ND | Absent | ND | 29 | Absent | ND | ND |
| **Median SNAP uV** | ND | 36 | 36 | NR | 44 | 2.7$^{\Delta}$ | ND | ND | 6 | 51 | ND | 16 | ND | 14.7 | 44 |

| | | | | | | | | | | | | | | | |
|---|---|---|---|---|---|---|---|---|---|---|---|---|---|---|---|
| **Median CV m/s** | ND | 43 | 49 | NR | 57 | 48$^{\Delta}$ | ND | ND | 60 | ND | ND | 53 | ND | 36.9 | 65 |
| **Ulnar SNAP uV** | ND | 16 | ND | 6.1 | 35 | 2.6$^{\Delta}$ | 6.7 | 4.3 | 5 | 24 | ND | 10.8 | ND | ND | 22 |
| **Ulnar CV m/s** | ND | 39 | ND | 47 | 52 | 46$^{\Delta}$ | 50 | 34 | 65 | 0 | ND | 37 | ND | ND | 54 |
| **Radial SNAP uV** | ND | 22 | ND | 11.7 | ND | ND | ND | ND | 28 | ND | ND | ND | ND | ND | 30 |
| **Radial CV m/s** | ND | 45 | ND | ND | ND | ND | ND | ND | 68 | ND | ND | ND | ND | ND | 69 |
| **Peroneal SNAP uV** | ND | ND | 3.5 | 10 | ND | ND | ND | ND | 1 | ND | ND | ND | 12 | 16.8 | Absent |
| **Peroneal CV m/s** | ND | ND | 36 | ND | ND | ND | ND | ND | 46 | ND | ND | ND | ND | 35.8 | Absent |
| **Sural SNAP uV** | ND | 11 | 1.8 | 8.7 | ND | 1.5 | Absent | Absent | Absent | Absent | ND | Absent | 14 | ND | 8 |
| **Sural CV m/s** | ND | 48 | 30 | 44 | ND | 30 | Absent | Absent | Absent | Absent | ND | Absent | ND | ND | 48 |
| **EMG** | ND | *Length dep den.* | *ND* | *Length dep den.* | ND | ND | ND | ND | Probably length-dependent den., some acute | *Length dep den.* | ND | Acute den. left ADM/FDIO on BG chronic den. | Length dep den. | Den. including paraspinal muscles | Lower limb den. with some acute changes |

**Supplementary Table 1. Neurophysiology data of *ARHGAP19* individuals.** Left upper limb unless other very severely affected, and right lower limb (unless very severely affected) unless only one limb available. Distal amplitudes reported EMG electromyography, CMAP compound motor action potential, CV conduction velocity, SNAP sensory nerve action potential, ND not done NA study not available CB conduction block , CS conduction slowing, R right L left UL upper limb FDIO first dorsal interosseous, ADM abductor digiti minimi, den. denervation, Δ right UL used * data not seen but report available only ** post IVIG study 18 months after onset.
