## Supplementary Table 2 for "Biallelic variants in *ARHGAP19* cause a motor-predominant neuropathy with asymmetry and conduction slowing"

| FAMILY ID | Family 1 | Family 2 | Family 3 | Family 4 | Family 5 | Family 6 & Family 15 | Family 7 & Family 14 | Family 8 | Family 9 | Family 10 | Family 11 | Family 12 & Family 19 | Family 13 | Family 16 | Family 17 | Family 18 | Family 20 |
|---|---|---|---|---|---|---|---|---|---|---|---|---|---|---|---|---|---|
| **Number of Affected** | 1 | 1 | 1 | 1 | 1 | 2 | 3 | 1 | 2 | 1 | 1 | 2 | 1 | 1 | 1 | 1 | 4 |
| **TESTING/METHODOLOGY INFORMATION** | | | | | | | | | | | | | | | | | |
| **Center Location** | Brazil | UK | UK | Netherland | Spain | Turkey | UK | UK | Germany | UK | Australia | UK | UK | UK | Germany | UK | Brazil |
| **Method of Identifying Variant** | WES | WES | WES | WES | WES | WES | WGS | WES | WGS | WGS | WGS | WES/WGS | WES | WES | WES | WES | WES |
| **Method of Validating Variant** | WES | Sanger sequencing | Sanger sequencing | Sanger sequencing | Sanger sequencing | Sanger sequencing | WGS | Sanger sequencing | WGS | N/A | WGS | Sanger sequencing | Sanger sequencing | Sanger sequencing | Sanger sequencing | Sanger sequencing | WES |
| **VARIANT DESCRIPTION** | | | | | | | | | | | | | | | | | |
| **Genomic Position Change (GRCh38/hg38)** | chr10:97265921 | chr10:97263448-97263450 | chr10:97235258-97235258 | hr10:97235282-97235282 | hr10:97263614-97263614 | chr10:97265979 | hr10:97263582-97263582 | chr10:97292627 | chr10:97263554 | chr10:97266097 | chr10:97246333 | chr10:97259559 | hr10:97263448-97263450 | chr10:97263611 | chr10:97259525 | chr10:97263470 | chr10:97259424 |
| **Genomic Position Change (GRCh37/hg19)** | chr10:99025678 | chr10:99023205-99023207 | chr10:98995015-98995015 | hr10:98995039-98995039 | hr10:99023371-99023371 | chr10:99025736 | hr10:99023339-99023339 | chr10:99052384 | chr10:99023311 | chr10:99025854 | chr10:99006090 | chr10:99019316 | hr10:99023205-99023207 | chr10:99023368 | chr10:99019282 | chr10:99023227 | chr10:99019181 |
| **Coding Sequence Change (NM_001136035)** | c.261dup | c.585dupA | c.1243C>T | c.1219C>T | c.419G>A | c.203T>C | c.451C>A | c.1A>G | c.479del | c.85A>G | c.932C>G | c.683T>A | c.585dupA | c.422T>G | c.717T>A | c.563del | c.818C>T |
| **Protein Sequence Change (NP_001129507)** | p.Pro88Alafs*43 | p.His196 Thrfs*10 | p.Gln415* | p.Arg407* | p.Gly140Asp | p.Leu68Pro | p.Gln151Lys | p.Met1? | p.Asn160Metfs*21 | p.Asn29Asp | p.Pro311Arg | p.Leu228His | p.His196 Thrfs*10 | p.Leu141Trp | p.Asn239Lys | p.Pro188Argfs*5 | p.Pro273Leu |
| **Exon Number Position** | 2 of 12 | 4 of 12 | 9 of 12 | 9 of 12 | 4 of 12 | 2 of 12 | 4 of 12 | 1 of 12 | 4 of 12 | 2 of 12 | 7 of 12 | 5 of 12 | 4 of 12 | 4 of 12 | Exon 5 of 12 | 4 of 12 | 5 of 12 |
| **Codon Change** | C/CC | _/A | CAA/TAA | CGA/TGA | GGT/GAT | CTC/CCC | CAG/AAG | ATG/GTG | AAT/AT | AAT/GAT | CCT/CGT | CTC/CAC | _/A | TTG/TGG | AAT/AAA | CCT/CT | CCC/CTC |
| **Consequence** | Insertion | frameshift | stop gained | stop gained | missense | missense/NMD | missense | start lost | frameshift | missense | missense | missense | frameshift | missense | missense | missense | missense |
| **Zygosity** | Hom | Hom | Hom | Hom | Hom | Hom | Hom | Hom | Hom | Hom | Hom | Hom | Hom | Hom | Hom | Hom | Hom |
| **dbSNP ID** | rs767899310 | rs772718801 | rs757781028 | rs751754099 | - | rs1026767404 | rs1842859321 | - | - | rs754312797 | - | rs562006800 | rs772718801 | rs755015441 | - | rs760101189 | rs1162465048 |
| **ALLELE FREQUENCY [Allele count/Total Allele Nu** | | | | | | | | | | | | | | | | | |
| **gnomAD v3.1.2 (Highest subpopulation if applicable) (76,156 genomes)** | 0.00001315 (0.0000294 Non-Finnish European) [2/152076] | 0 | 0.00001315 (0.00003267 South Asian) [1/251376], (0.000008795 Non-Finnish European) [1/251376] | 0.00003287 (0.0002621 Latino/Admixed American) [4/152091], (0.0000147 Non-Finnish European) [1/152091] | 0 | 0.000006572 (0.00002414 African/African American) [1/152165] | 0 | 0 | 0 | 0 | 0 | 0.00002628 (0.001 South Asian) [4/152192] | 0 | 0 | 0 | 0.000006576 (0.0002076 South Asian) [1/152065] | 0 |
| **gnomAD v2.1.1 (Highest subpopulation if applicable) (15,708 genomes and 125,748 exomes)** | 0.00003181 (0.00006154 Non-Finnish European) [7/251464] | 0.00002784 (0.0001848 Finnish) [4/251439], (0.00002638 Non-Finnish European) [3/251439] | 0.00001591 (0.0004139 South Asian) [2/152140] | 0.00003978 (0.0000868 Latino) [3/251344], (0.00006533 South Asian) [5/251344], (0.00004399 Non-Finnish European) [2/251344] | 0 | 0.000003976 (0.00002891 Latino) [1/251491] | 0 | 0 | 0 | 0 | 0 | 0.00009543 (0.001 South Asian) [24/251464] | 0.00002784 (0.0001848 Finnish) [4/251439], (0.00002638 Non-Finnish European) [3/251439] | 0.000003979 (0.00003267 South Asian) [1/251345] | 0 | 0.00005568 (0.0004573 South Asian) [14/251438] | 0 |
| **Ensembl (Highest frequency observed)** | < 0.01 | < 0.01 | < 0.01 | < 0.01 | N/A | < 0.01 | < 0.01 | N/A | N/A | < 0.01 | N/A | < 0.01 | < 0.01 | < 0.01 | N/A | < 0.01 | < 0.01 |
| **Iranome (~800 exomes)** | 0 | 0 | 0 | 0 | 0 | 0 | 0 | 0 | 0 | 0 | 0 | 0 | 0 | 0 | 0 | 0 | 0 |
| **GME Variome (~2,500 exomes)** | 0 | 0 | 0 | 0 | 0 | 0 | 0 | 0 | 0 | 0 | 0 | 0 | 0 | 0 | 0 | 0 | 0 |
| **UK Biobank (394,841 exomes)** | 0 | 0 | 0.00000271782 | 0 | 0 | 0 | 0 | 0.0000298948 | 0 | 0.00002174138 | 0 | 0 | 0 | 0 | 0 | 0 | 0.0000013589 |
| **TOPMed (132,345 genomes)** | 0.000023891 | 0.000007964 | 0 | 0.000015928 | 0 | 0.000015928 | 0 | 0 | 0 | 0 | 0 | 0 | 0.000007964 | 0 | 0 | 0.000007964 | 0.000007964 |
| **54KJPN (~54,000 genomes)** | 0 | 0 | 0 | 0 | 0 | 0 | 0 | 0 | 0 | 0 | 0 | 0 | 0 | 0 | 0 | 0 | 0 |
| **QSG Database (23741 exomes)** | 0 | 0 | 1 hom | 0 | 0 | 0 | 0 | 0 | 0 | 0 | 0 | 2 hom | 0 | 0 | 0 | 0 | 0 |
| ***IN SILICO PREDICTIONS AND CLASSIFICATION*** | | | | | | | | | | | | | | | | | |
| **GERP** | 3.11 | 2.64 | 1.57 | -1.56 | 5.87 | 2.63 | 3.94 | 2 | - | 2.63 | 5.66 | 2.46 | 2.64 | 3.94 | 2.97 | 3.94 | 5.11 |
| **CADD_Phred** | - | 34 | 39 | 35 | 26.3 | 28 | 26.2 | 22.6 | | 23.7 | 27.5 | 27.6 | 34 | 29.2 | - | 27 | 27.1 |
| **Polyphen-2** | - | - | - | - | PD 1 | PD 0.987 | PD 0.996 | B 0 | - | PD 0.57 | PD 1 | PD 0.998 | - | PD 1 | - | PD 0.998 | - |
| **SIFT** | - | - | - | - | D 0 | D 0.01 | D 0 | D 0 | - | D 0.21 | D 0 | D 0 | - | D 0 | 0.004 | D 0 | 0.003 |
| **PROVEAN** | - | D-14.45 | N -1.66 | N -2.25 | D -6.72 | D -2.75 | D -3.58 | N -0.68 | N -1.15 | N -0.28 | D -7.95 | D -6.07 | D -14.45 | D -5.33 | D -4.93 | - | D -9.23 |
| **MutationTaster** | - | DC 1 NMD | DC 0.99 NMD | DC 0.99 NMD | DC 0.99 | DC 0.99 | DC 0.99 | DC 1 | DC 1 NMD | DC 0.503 | DC 0.99 | DC 0.99 | DC 1 NMD | DC 0.99 | DC 0.99 | DC 0.99 | DC 1 |
| **ACMG Clinical Significance Classification** | Pathogenic (PVS1, PM2, PM4, PP1, PP3) | Pathogenic (PS3, PM1, PM2, PM4, PP1, PP3) | Pathogenic (PVS1, PM2, PM4, PP1, PP3) | Pathogenic (PVS1, PM2, PP3) | Pathogenic (PS3, PM1, PM2, PP1, PP3) | Likely Pathogenic (PS4, PM2, PP1, PP3) | Pathogenic (PS3, PS4, PM1, PM2, PP1, PP3) | VUS (PM2, PP1, PP3) | Pathogenic (PM1, PM2, PM4, PP1, PP3) | Likely Pathogenic (PS3, PM2, PP3) | Likely Pathogenic (PS4, PM2, PP1, PP3) | Pathogenic (PS4, PM1, PM2, PP1, PP3) | Likely Pathogenic (PS3, PM1, PM2, PP3) | Likely Pathogenic (PM1, PM2, PP1, PP3) | Likely Pathogenic (PM1, PM2, PP1, PP3) | Pathogenic (PM1, PM2, PM4, PP1, PP3) | VUS (PM1, PM2, PP3) |

**Supplementary Table 2.** *ARHGAP19* variants table. Abbreviations: ACMG - American College of Medical Genetics and Genomics, CADD - Combined Annotation Dependent Depletion, Comp Het - Compound Heterozygous, D – Damaging, DC - Disease causing, Del – Deleterious, GERP - Genomic Evolutionary Rate Profiling, GME - The Greater Middle East, Hom – Homozygous, N – Natural, N/A - Not available, Not applicable, PD - Probably Damaging, PM/_M- Pathogenic Moderate, PP/_P - Pathogenic Supporting, PS/_S - Pathogenic Strong, PSD - Possibly Damaging, PVS/_VS - Pathogenic Very Strong, QSG - Queen Square Genomics, RoH - Region of Homozygosity, SNP - Single Nucleotide Polymorphism, Tol – Tolerated, WES - Whole Exome Sequencing.
