## Supplementary Table 3 for "Biallelic variants in *ARHGAP19* cause a motor-predominant neuropathy with asymmetry and conduction slowing"

**Supplementary Table 3.** UniProt accession codes used for Multiple Sequence Alignment of ARHGAP19 orthologs.

| **Sequence** | **Animal species** | **Uniprot accession code** |
|---|---|---|
| HUMAN ARHGAP19 | *Homo sapiens* | Q14CB8 |
| CHINPANZEE ARHGAP19 | *Pan troglodytes* | K7DQC3 |
| MACAQUE ARHGAP19 | *Macaca mulatta* | H9G104 |
| MOUSE Arhgap19 | *Mus musculus* | Q8BRH3 |
| RAT Arhgap19 | *Rattus norvegicus* | M0R5V4 |
| BOVINE ARHGAP19 | *Bos taurus* | F1MGB9 |
| DOG ARHGAP19 | *Canis lupus familiaris* | F6UNS4 |
| CHICKEN ARHGAP19 | *Gallus gallus* | Q5F3G0 |
| FROG arhgap19 | *Xenopus laevis* | Q6INE5 |
| ZEBRAFISH arhgap19 | *Danio rerio* | F6NW06 |
| FRUIT FLY RhoGAP54D | *Drosophila melanogaster* | Q8IGY7 |
