## Supplementary Table 4 for "Biallelic variants in *ARHGAP19* cause a motor-predominant neuropathy with asymmetry and conduction slowing"

**Supplementary Table 4. Extracted AlphaMissense scores and associated predicted variant impact for identified *ARHGAP19* missense variants** (in yellow shade are variants located within the GAP domain).

| **Variant (c.)** | **AA change (p.)** | **AlphaMissense pathogenicity**** | **AlphaMissense Class** |
|---|---|---|---|
| c.85A>G | p. Asn29Asp | 0.0882 | likely_benign |
| c.203T>C | p.Leu68Pro | 0.9953 | likely_pathogenic |
| c.419G>A | p.Gly140Asp | 0.9985 | likely_pathogenic |
| c.422T>G | p.Leu141Trp | 0.9915 | likely_pathogenic |
| c.451C>A | p.Gln151Lys | 0.7953 | likely_pathogenic |
| c.683T>A | p.Leu228His | 0.8643 | likely_pathogenic |
| c.717T>A | p.Asn239Lys | 0.9058 | likely_pathogenic |
| c.818C>T | p.Pro273Leu | 0.9793 | likely_pathogenic |
| c.932C>G | p.Pro311Arg | 0.9945 | likely_pathogenic |
