## Supplementary Table 5 for "Biallelic variants in *ARHGAP19* cause a motor-predominant neuropathy with asymmetry and conduction slowing"

**Supplementary Table 5.** Primers for NGS sequencing (MiSeq).

| crRNA | Forward primer | Reverse primer |
|---|---|---|
| 1 | ACACTCTTTCCCTACACGACGCTCTTCCGATC Tggttgctgtttcccctactat | GACTGGAGTTCAGACGTGTGCTCTTCCGATC TagtgtggatgatgaacCTGTGA |
| 2 | ACACTCTTTCCCTACACGACGCTCTTCCGATC Tccggaaatatgttaggaccattg | GACTGGAGTTCAGACGTGTGCTCTTCCGATC TAGGTCACTGGCATTCATctga |
| 3 | ACACTCTTTCCCTACACGACGCTCTTCCGATC Tggttgctgtttcccctactat | GACTGGAGTTCAGACGTGTGCTCTTCCGATC TgtgtggatgatgaacCTGTGA |
| 4 | ACACTCTTTCCCTACACGACGCTCTTCCGATC TGGCCGCCCATTATCTTCAAC | GACTGGAGTTCAGACGTGTGCTCTTCCGATC Tgtctgcaattttggacgca |
| 5 | ACACTCTTTCCCTACACGACGCTCTTCCGATC Taattgtgggttgctgtttcc | GACTGGAGTTCAGACGTGTGCTCTTCCGATC Tcagctaatgatgtagaaggcaca |
| 6 | ACACTCTTTCCCTACACGACGCTCTTCCGATC Ttgtttacaccctgacacagtt | GACTGGAGTTCAGACGTGTGCTCTTCCGATC TTGTTTGGCGGTGTGGTAGA |
