## Supplementary Table 6 for "Biallelic variants in *ARHGAP19* cause a motor-predominant neuropathy with asymmetry and conduction slowing"

**Supplementary Table 6.** qPCR primers

| **Primer name** | **Primer sequence** |
|---|---|
| *ARHGAP19* qPCR forward | GGCATCAAAGGATGACCTTG |
| *ARHGAP19* qPCR reverse | CTTTGCTGACTCTGGCATATC |
| *RPL18A* qPCR forward | CCCACAACATGTACCGGGAA |
| *RPL18A* qPCR reverse | TCTTGGAGTCGTGGAACTGC |
