## Supplementary Table 7 for "Biallelic variants in *ARHGAP19* cause a motor-predominant neuropathy with asymmetry and conduction slowing"

**Supplementary Table 7.** Drosophila stocks.

| **Stock name** | **Genotype** | **Source** |
|---|---|---|
| actin-Gal4 | *y[1] w[*]; P{Act5C-GAL4-w}E1/CyO* | BDSC #25374 |
| UAS-RhoGAP54D shRNA | [1] *v[1]; P{y[+t7.7] v[+t1.8]=TRiP.HMS03522}attP40* | BDSC #54051 |
| RhoGAP54D-Gal4 | y[1] w[*]; TI{GFP[3xP3.cLa]=CRIMIC.TG4.1}RhoGAP54D[CR02433-TG4.1]/SM6a | BDSC #92267 |
| deGradFP | w[*]; P{w[+mC]=UAS-Nslmb-vhhGFP4}3 | BDSC #38421 |
| GFP:RhoGAP54D | w*; GFP:RhoGAP54D / CyO | (19) |
| | yw; UAS-CD4::TdTom | unknown |
| | UAS-mCherry shRNA | unknown |
| RhoGAP54D null allele | w*; RhoGAP54D$^{KO}$ / CyO | Gift from Bellaïche lab |
| Tubulin-Gal4 | Tub-Gal4 / Tm6b | Gift from Southall lab |
| | w*; GFP:RhoGAP54D / CyO ; Tub-Gal4 / Tm6b | This study |
| | w*; GFP:RhoGAP54D / CyO ; UAS-Nslmb-vhhGFP4 / TM6b | This study |
