## Supplementary Table 8 for "Biallelic variants in *ARHGAP19* cause a motor-predominant neuropathy with asymmetry and conduction slowing"

**Supplementary Table 8.** List of differentially expressed genes containing log2-fold change values with their corresponding p values.

| hgnc_symbol | Log2FC | Pvalue |
|---|---|---|
| TSPAN6 | 0.639570257 | 0.019933541 |
| TNMD | 4.553646945 | 0.041557808 |
| CFH | 1.1605223 | 0.03832081 |
| FUCA2 | -0.82273497 | 0.019694874 |
| GCLC | 0.91613137 | 0.004091995 |
| STPG1 | -0.889011171 | 0.021474172 |
| NIPAL3 | -0.89531352 | 0.02836666 |
| ENPP4 | 1.120612538 | 0.010399131 |
| SEMA3F | 1.413306495 | 0.001822619 |
| LASP1 | -1.003030389 | 0.00137536 |
| SNX11 | -0.611960111 | 0.036273953 |
| TMEM176A | 5.482464221 | 0.002656089 |
| CYP26B1 | 1.608948126 | 0.000197585 |
| CFLAR | 0.692584861 | 0.018705819 |
| TFPI | 1.504175898 | 0.000955 |
| POLDIP2 | -0.562364787 | 0.037797603 |
| CAMKK1 | -1.358356449 | 0.001852072 |
| HSPB6 | -1.887008515 | 3.17E-05 |
| ARHGAP33 | -1.09467204 | 0.016481325 |
| PDK4 | 2.795909188 | 1.47E-10 |
| ATOSB | -1.178128921 | 3.91E-06 |
| ITGA3 | -1.353700966 | 0.019206802 |
| ZFX | 0.766930576 | 0.03948758 |
| CRLF1 | -2.173646998 | 2.07E-05 |
| TMEM132A | 0.90423349 | 0.032896232 |
| AP2B1 | -0.499081092 | 0.043099649 |
| TAC1 | 2.696361109 | 0.031588449 |
| CX3CL1 | 4.346055178 | 0.035416116 |
| TNFRSF12A | -1.517040977 | 4.53E-06 |
| CCL26 | -2.514854259 | 6.17E-05 |
| DBF4 | -1.59623422 | 0.005380249 |
| TBXA2R | -1.20937122 | 0.015398714 |
| ELAC2 | -0.614486392 | 0.024783894 |
| MYH13 | 2.903619459 | 0.041381242 |
| CCDC124 | -0.655457161 | 0.022197718 |
| RPUSD1 | -0.633156798 | 0.019845337 |
| TSR3 | -0.640890211 | 0.036072127 |
| FMO3 | 3.467515376 | 0.003283028 |
| MYLIP | 1.369249552 | 0.005034653 |

| | | |
|---|---|---|
| E2F2 | -3.667857981 | 0.042985388 |
| CYTH3 | -0.677633096 | 0.006571254 |
| CYB561 | -0.949245166 | 0.012175941 |
| IL32 | 1.898604732 | 0.001655311 |
| RHOBTB2 | -0.667738209 | 0.034019893 |
| MLXIPL | 7.106759081 | 0.000413842 |
| ETV7 | 1.29016718 | 0.030925459 |
| UQCRC1 | -0.809633293 | 0.015302457 |
| STARD3NL | -0.644940962 | 0.015726725 |
| CD9 | -1.777581614 | 0.003145227 |
| NCAPD2 | -1.028541924 | 0.030733583 |
| IDS | 0.709279807 | 0.044028491 |
| CD4 | -2.757359929 | 0.040554576 |
| FYN | 0.886305338 | 0.016892132 |
| FMO1 | 6.658266352 | 7.37E-07 |
| LYPLA2 | -0.897748802 | 0.007858991 |
| TSPAN9 | 1.356984141 | 0.000309403 |
| MBTD1 | 0.701626369 | 0.035863527 |
| UTP18 | -0.725741055 | 0.007168265 |
| PTBP1 | -0.543067834 | 0.022059087 |
| DPF1 | -2.894354671 | 1.90E-09 |
| SYT7 | 3.130453921 | 2.35E-06 |
| LARS2 | -0.650410153 | 0.021138958 |
| PLAUR | -0.835387409 | 0.047442943 |
| ANLN | -4.441160012 | 5.79E-10 |
| PPP5C | -0.698646897 | 0.005771363 |
| CEP68 | 0.611546036 | 0.022499971 |
| LDAF1 | 0.709890298 | 0.0089436 |
| EHD3 | -0.927811695 | 0.007749429 |
| PSMC4 | -0.543143894 | 0.043570765 |
| SLC25A39 | -0.756513628 | 0.013764806 |
| MVP | -0.837416802 | 0.039261962 |
| RWDD2A | 0.699208163 | 0.021787997 |
| ANGEL1 | 0.573194785 | 0.028488756 |
| DNASE1L1 | -1.127192918 | 0.040583486 |
| DDX11 | -0.616249426 | 0.049182777 |
| TACC3 | -3.202084749 | 7.40E-07 |
| POLA2 | -1.148817671 | 0.03666799 |
| CAPN1 | -0.964460154 | 0.004817342 |
| MTMR11 | -1.215194787 | 0.000785444 |
| SLC38A5 | -1.392907222 | 0.003819583 |
| WWTR1 | 0.732217936 | 0.044583559 |

| | | |
|---|---|---|
| ATP1A2 | 3.176460748 | 0.003144709 |
| PHLDB1 | -0.719766888 | 0.009850571 |
| CD74 | -1.067911567 | 0.025694533 |
| HGF | 4.553404972 | 2.57E-08 |
| NRXN3 | 3.347028828 | 0.003505523 |
| FHL1 | 0.735939459 | 0.02745138 |
| SLC45A4 | 0.56958837 | 0.043081405 |
| GRAMD1B | -1.511111781 | 0.00027725 |
| RNH1 | -0.851754218 | 0.008997279 |
| BIRC3 | 1.512388518 | 0.005798191 |
| PLEKHO1 | -1.325131359 | 6.76E-06 |
| GCLM | 0.65638233 | 0.039125359 |
| DEPDC1 | -4.658953154 | 6.31E-10 |
| CCDC28A | 0.74946828 | 0.003345607 |
| RRAGD | 1.164539844 | 0.027362434 |
| HSD17B6 | 2.006752117 | 0.000236498 |
| NCAPH2 | -0.902255385 | 0.015618655 |
| TOMM34 | -1.638966309 | 6.92E-05 |
| SEC63 | 0.809938926 | 0.012645056 |
| VIM | -2.071791149 | 4.41E-07 |
| RNASET2 | 0.933158524 | 0.015444891 |
| BTN3A1 | 1.131199177 | 9.41E-05 |
| PRKCH | 1.458747769 | 0.017575641 |
| INSRR | 3.502074306 | 1.23E-05 |
| IFNGR1 | 1.389040748 | 1.84E-07 |
| BMAL2 | -1.838339021 | 0.000136361 |
| ANK1 | -2.292259445 | 0.000241268 |
| HMGB3 | -1.180671931 | 0.014545011 |
| BAK1 | -0.634460011 | 0.023010955 |
| IKZF2 | 1.229005091 | 0.004564648 |
| ARHGAP31 | -0.90959396 | 0.044976068 |
| EIPR1 | -0.81421773 | 0.006266258 |
| PNPLA6 | -0.78024344 | 0.011366288 |
| CHPF2 | -0.689279394 | 0.028439449 |
| LRRC7 | 3.294050211 | 0.00235308 |
| FUT8 | 0.580494302 | 0.020579734 |
| MAP2K3 | -1.856050151 | 7.54E-07 |
| TMSB10 | -1.068932927 | 0.001049778 |
| RNF19A | 0.801270383 | 0.00641626 |
| MYOC | 3.158818966 | 0.001685824 |
| SH3YL1 | 0.643874118 | 0.010072928 |
| VCL | -1.032885171 | 0.000477111 |

| | | |
|---|---|---|
| DEPDC1B | -3.851215423 | 2.10E-06 |
| RPL26L1 | -1.085998381 | 0.002119332 |
| PI4K2B | -0.872307901 | 0.000354736 |
| EDC4 | -0.513701862 | 0.041615923 |
| TRIO | -0.989244608 | 0.001155496 |
| RIPOR1 | -0.614590888 | 0.01464506 |
| PHLPP2 | -0.87546425 | 0.006829276 |
| SPDL1 | -1.953878214 | 0.000293193 |
| CTNS | 0.537451476 | 0.048842414 |
| PHF23 | -0.628466484 | 0.01958838 |
| CDH10 | 1.698255382 | 0.000608117 |
| RIPOR3 | 4.34904898 | 2.17E-12 |
| TDP1 | -0.945001079 | 0.013040686 |
| AIFM2 | 0.832939975 | 0.001546908 |
| SPATA7 | 0.605874416 | 0.03695941 |
| CAPG | -2.3638383 | 0.000113066 |
| AP2S1 | -0.827290971 | 0.008818886 |
| TG | 1.456738037 | 0.00341532 |
| ADAM28 | 2.137639732 | 0.000119527 |
| OFD1 | 0.627952596 | 0.022247705 |
| GPM6B | 1.684699245 | 0.048470168 |
| PREX2 | 2.82031422 | 0.021770475 |
| ATOSA | 0.655687603 | 0.037385061 |
| ARAP2 | 5.388458873 | 0.003815644 |
| SCML1 | 1.018002451 | 0.001538843 |
| MAP4 | -0.534307968 | 0.029604682 |
| TSPAN17 | -0.547564717 | 0.032033573 |
| NOP16 | -0.602006865 | 0.042524565 |
| CC2D2A | -1.043290648 | 0.023171639 |
| RRM2B | 1.183643231 | 9.71E-06 |
| CELF2 | 2.226256381 | 1.01E-05 |
| COL9A2 | -1.882730983 | 0.005709941 |
| H6PD | 0.787146069 | 0.033297567 |
| PER3 | 0.563063525 | 0.032050949 |
| RFC2 | -0.893856077 | 0.041283792 |
| NEDD4L | 1.551380542 | 1.08E-05 |
| DKK3 | -1.433332312 | 0.000266067 |
| LETMD1 | 0.52531476 | 0.041256346 |
| SLC4A8 | -2.403747862 | 2.17E-06 |
| PTGER3 | -1.051668194 | 0.025902315 |
| BCAR1 | -1.181271831 | 8.52E-06 |
| HERPUD1 | 0.943558983 | 0.00247147 |

| | | |
|---|---|---|
| HOMER3 | -0.648306413 | 0.048857885 |
| RAD51 | -2.803553631 | 3.15E-06 |
| THOC3 | -1.15803758 | 0.02676223 |
| PLEKHA5 | 0.589562448 | 0.044924616 |
| TTC17 | 0.650177821 | 0.023194381 |
| FOXN3 | 0.802913955 | 0.009258155 |
| METTL24 | 1.91861885 | 0.001070186 |
| OPN3 | 1.066234477 | 0.026824993 |
| PTPRN | -1.969033266 | 2.88E-05 |
| HHAT | 1.116565405 | 0.001956604 |
| SYNE2 | -1.485957394 | 0.023948282 |
| PLEKHH1 | 2.20038196 | 0.009263967 |
| RELT | -1.62372289 | 3.93E-06 |
| GALC | 0.791253975 | 0.012245393 |
| SZRD1 | -0.826039497 | 0.006482609 |
| GINM1 | 1.020936586 | 0.004420897 |
| TRAF1 | 1.322837856 | 3.89E-05 |
| DCBLD2 | -0.798509019 | 0.008563355 |
| F7 | 2.008185859 | 0.047669085 |
| USP13 | -0.886515596 | 0.028809146 |
| PPP1R12A | -0.719231991 | 0.026279313 |
| ATP2B4 | -0.522879479 | 0.045330049 |
| UNKL | 1.034426489 | 0.000108781 |
| TBXAS1 | -1.582026632 | 0.037369912 |
| MXD1 | 0.94536319 | 0.007309364 |
| SLC2A3 | 1.084869333 | 0.013779981 |
| PTPRU | 1.13104339 | 0.002114338 |
| LZTS1 | -1.575366966 | 0.021706818 |
| GUCY1B1 | 1.901138084 | 0.00123916 |
| GPBP1 | 0.609493229 | 0.018228768 |
| CS | -0.724592355 | 0.008018693 |
| LTK | -2.010064036 | 0.032478645 |
| POLD1 | -0.863127471 | 0.039804064 |
| SLC6A16 | 2.363056319 | 7.44E-05 |
| SPHK2 | -0.681391828 | 0.038649792 |
| ISOC2 | -0.810330003 | 0.044012729 |
| U2AF2 | -0.554455221 | 0.030672254 |
| EPN1 | -0.56423582 | 0.0359043 |
| GPC1 | -0.894138679 | 0.018051194 |
| DLX3 | -2.592819732 | 0.002855556 |
| CCN5 | 2.574906476 | 0.000461132 |
| CDON | 2.571716878 | 7.49E-15 |

| | | |
|---|---|---|
| BCAS1 | 1.626206008 | 0.040735983 |
| PMS1 | 0.635709574 | 0.024866385 |
| HMG20B | -0.823448939 | 0.006374641 |
| CALCRL | 1.218521322 | 0.025411731 |
| AP3D1 | -0.467047641 | 0.048067621 |
| NHERF2 | -1.519221234 | 0.013539346 |
| NTHL1 | -0.721405504 | 0.048542654 |
| BLTP3A | 0.727684179 | 0.013617762 |
| GNAI3 | -0.484277436 | 0.044258168 |
| WDR18 | -0.848435164 | 0.016708009 |
| TRAM2 | -0.634804334 | 0.017334182 |
| MCM10 | -3.943867589 | 4.27E-06 |
| ERBB3 | -2.032553764 | 0.004609064 |
| PDIA5 | -0.784811683 | 0.03506371 |
| GSTO2 | -2.230318776 | 0.001246888 |
| TLE2 | 0.894640557 | 0.005820736 |
| TBC1D1 | -0.986596101 | 0.010139732 |
| ASPM | -4.456912471 | 0.000323471 |
| ELOVL1 | -1.103313687 | 0.000157033 |
| MPPED2 | 4.827978002 | 0.002049048 |
| TRMT11 | 0.710761761 | 0.022454822 |
| PFKP | -1.127558972 | 0.000327258 |
| KLF6 | -0.797347114 | 0.011680579 |
| PKM | -1.052056683 | 0.002195547 |
| RHOA | -0.823150035 | 0.0057803 |
| IDH3G | -0.704943184 | 0.033349 |
| ROGDI | -1.063438868 | 0.009485902 |
| PDZD4 | 1.912910335 | 0.001239883 |
| PDK3 | 1.091763363 | 0.013260474 |
| HYAL2 | -0.714881587 | 0.035919934 |
| RASSF1 | -1.329479811 | 4.65E-06 |
| FGFR3 | -3.858134893 | 3.89E-11 |
| PLEKHH3 | -0.60676769 | 0.04465157 |
| FTSJ1 | -0.850722244 | 0.001965873 |
| PRR11 | -3.980528818 | 2.64E-09 |
| REEP1 | -3.727673197 | 0.021974057 |
| LAPTM4A | 0.629661492 | 0.025034412 |
| PITX1 | -3.725824044 | 0.000738277 |
| GAL | -5.881000584 | 1.38E-06 |
| CLEC2D | 1.134943383 | 0.007272986 |
| MAOB | 3.94808256 | 0.000142805 |
| RORA | 1.424470416 | 0.01910354 |

| | | |
|---|---|---|
| TGFBR3 | 1.033207629 | 0.016987274 |
| ATP1B3 | -1.020417252 | 0.001371868 |
| SPTB | -3.98561724 | 0.00206431 |
| FSTL3 | -1.294236919 | 5.68E-06 |
| RNF126 | -0.564162441 | 0.036546391 |
| JMJD6 | -0.582686872 | 0.032664783 |
| AP3M2 | 0.719099009 | 0.01415678 |
| ST6GALNAC2 | -2.830423891 | 0.000232878 |
| CAMK2A | -1.563646253 | 0.022080855 |
| RAD18 | -0.857654972 | 0.021529139 |
| ATP2B1 | 1.064165701 | 0.003172176 |
| MGAT4A | 3.527178666 | 0.011504618 |
| WDR1 | -1.268347108 | 1.43E-05 |
| SEL1L | 0.932887237 | 0.01047228 |
| TRIP13 | -3.144361093 | 3.67E-07 |
| TRIB2 | 1.07724001 | 0.001447412 |
| DAZAP1 | -0.898729652 | 0.002239121 |
| CYBRD1 | 1.037183169 | 0.001391066 |
| SLC6A15 | -1.207025951 | 0.044409425 |
| ADGRL1 | 0.945148446 | 0.000991142 |
| ACTN1 | -0.971435579 | 0.005545441 |
| PTPN18 | -0.552103567 | 0.032279155 |
| LIMS2 | -1.035502844 | 0.002705004 |
| SPEG | -1.200184914 | 0.001350817 |
| ALDH3A2 | 1.634120274 | 0.002557615 |
| TFRC | 0.863547163 | 0.028390563 |
| UBE2D1 | 0.705104706 | 0.018571276 |
| HSD17B10 | -0.787073085 | 0.014025263 |
| HMMR | -4.745634414 | 6.79E-10 |
| STK10 | -1.022391222 | 0.000500488 |
| EVC | 1.250410047 | 8.45E-05 |
| IRAG1 | -1.764619375 | 0.002114425 |
| AP1M1 | -0.785824547 | 0.007887672 |
| MCM2 | -2.180580197 | 9.82E-05 |
| ALPK1 | 0.867808936 | 0.001136882 |
| GSDMB | 1.17565836 | 0.029672137 |
| FRY | 2.44014645 | 7.05E-09 |
| PPP2R2C | -2.737229 | 0.007936208 |
| NTN4 | 1.251297205 | 0.026276241 |
| MYDGF | -1.12638319 | 0.000278114 |
| TUBE1 | 1.282993981 | 0.000731263 |
| GTSE1 | -5.018050692 | 5.28E-13 |

| | | |
|---|---|---|
| FOSL2 | 0.925229336 | 0.004560399 |
| FSCN1 | -1.423064761 | 1.50E-05 |
| ACTB | -1.504006552 | 3.24E-06 |
| MOCOS | 1.152785362 | 0.011274948 |
| PLD1 | 1.730045948 | 3.21E-05 |
| WDR62 | -5.257948788 | 5.23E-12 |
| FMO4 | 1.082988803 | 0.009520914 |
| RGS11 | 1.329208446 | 0.034851244 |
| SPAG5 | -3.6472863 | 2.59E-07 |
| ACACB | 0.815741159 | 0.015288615 |
| ARHGEF1 | -0.552639151 | 0.020262662 |
| RARB | -3.207148153 | 0.000196402 |
| UBE2T | -2.524934097 | 1.03E-05 |
| IL4R | -0.720500782 | 0.018528336 |
| CAPN6 | 3.842806804 | 0.005719533 |
| SNRPA | -1.247796533 | 8.95E-05 |
| CAPZB | -0.93489672 | 0.003420175 |
| GPR137B | 0.626897031 | 0.047634999 |
| NAALAD2 | 2.084834394 | 0.00520031 |
| FGFR1 | 0.560013919 | 0.034927433 |
| FBLN1 | 1.33868981 | 7.93E-05 |
| AMPH | -1.575658381 | 6.35E-05 |
| SYNJ2 | -1.669894491 | 6.05E-06 |
| ADCY2 | 1.740746606 | 0.004578599 |
| GNB1 | -0.52782454 | 0.022892979 |
| EDN1 | -1.99503771 | 6.53E-10 |
| ZCWPW1 | 1.279212727 | 0.001566375 |
| BRINP1 | 1.93103248 | 3.11E-08 |
| MYH7B | 1.643927911 | 0.027405896 |
| RUNX1T1 | 0.991206963 | 0.000166697 |
| SLC1A3 | 2.687346704 | 1.41E-08 |
| TNS1 | -0.803577205 | 0.021601621 |
| REXO1 | -0.563873772 | 0.020128516 |
| RAPGEF3 | -1.078879636 | 0.031094499 |
| FDFT1 | -0.850672112 | 0.009575703 |
| PAFAH1B3 | -2.236725626 | 5.37E-09 |
| KIF22 | -2.445469199 | 9.29E-06 |
| DNM2 | -0.526226568 | 0.031812626 |
| RABL2B | 0.544392861 | 0.034858592 |
| SLC4A4 | -2.065242621 | 5.02E-05 |
| COL5A3 | 1.969350575 | 5.63E-05 |
| PSEN1 | 0.611364313 | 0.008672804 |

| | | |
|---|---|---|
| NDC80 | -3.777451986 | 9.73E-08 |
| COL4A4 | 1.714433108 | 0.010718533 |
| TCF7 | -3.441123031 | 6.73E-07 |
| MEF2C | -1.280656777 | 0.001254982 |
| ZNF510 | 0.685531583 | 0.029848096 |
| AACS | -0.826823396 | 0.010696037 |
| PCDHB4 | 1.26538928 | 0.000496922 |
| ATP8B1 | -0.700339007 | 0.040332181 |
| MPP4 | -2.35857215 | 0.000127083 |
| STRADB | 0.568966337 | 0.018135815 |
| C1QTNF3 | -1.178768641 | 0.004322235 |
| DLG3 | 0.63280282 | 0.026768093 |
| KCNK2 | 1.115866313 | 0.00096467 |
| SERTAD4 | 2.157076924 | 2.47E-05 |
| GSK3B | 0.727223505 | 0.008689076 |
| RNF13 | 0.742769156 | 0.001511514 |
| TRPM3 | 5.98773456 | 7.12E-05 |
| OXCT1 | -1.469407289 | 2.45E-05 |
| FAM234B | 0.956079952 | 0.016135178 |
| NCOA1 | 0.758141066 | 0.013631786 |
| EFR3B | -2.454351596 | 0.001167368 |
| KIF3C | -0.541218174 | 0.048204193 |
| MAPRE3 | 0.663494974 | 0.020483433 |
| CD59 | -0.872101027 | 0.032667139 |
| CD82 | 1.841360528 | 0.000119646 |
| OVGP1 | 1.163563042 | 0.033391663 |
| CTTN | -0.599180369 | 0.019591621 |
| WNT11 | 1.202506153 | 0.032047229 |
| ORC1 | -2.854989462 | 0.000101309 |
| RAD54L | -4.112418462 | 7.71E-08 |
| MRPL28 | -0.701336086 | 0.039473949 |
| FAT2 | 2.053046796 | 0.007511868 |
| ERO1B | 1.262460209 | 0.000310656 |
| TXLNG | 0.794272274 | 0.010032072 |
| NOX4 | 2.409040801 | 0.000144497 |
| ACOX3 | -0.845087305 | 0.001673686 |
| TRIP6 | -0.765025178 | 0.023842517 |
| ACHE | -1.864571446 | 0.036834032 |
| FTL | 1.009617444 | 0.003124133 |
| ATXN7L3 | -0.50346937 | 0.044600275 |
| LPCAT2 | -1.247572208 | 0.00012933 |
| NID2 | 1.560129209 | 0.002590909 |

| | | |
|---|---|---|
| GNAS | -0.736260223 | 0.004499369 |
| AURKA | -4.369728834 | 1.13E-09 |
| CASS4 | 2.449790231 | 0.000547626 |
| PIR | 1.731379032 | 2.21E-05 |
| SULT2B1 | 2.060312583 | 0.046309417 |
| ASAP3 | 0.843713759 | 0.004739194 |
| TPX2 | -4.509956755 | 3.61E-10 |
| FKBP1A | -1.089261157 | 0.002221884 |
| EBF4 | 2.172995749 | 1.07E-05 |
| KIZ | 0.918685727 | 0.001605972 |
| DYNLL1 | -0.768723841 | 0.015713932 |
| P2RX7 | 1.801262002 | 0.000138669 |
| SLC23A2 | 0.721806784 | 0.004433399 |
| SLC8B1 | 0.662525427 | 0.028596412 |
| OAS1 | 2.379611387 | 0.025335555 |
| GCN1 | -0.723419033 | 0.00804386 |
| PXN | -0.584203165 | 0.03233889 |
| FXYD5 | -1.578646556 | 9.07E-05 |
| ZNF302 | 0.690842679 | 0.036219624 |
| GMIP | -0.583442691 | 0.042038106 |
| BIRC5 | -5.778652281 | 4.57E-15 |
| LAG3 | -1.490104022 | 0.012441289 |
| ARHGAP4 | 1.443017002 | 0.025727948 |
| ANKRD24 | 1.891610939 | 0.00030045 |
| DHX32 | 0.560285366 | 0.03332647 |
| PCBP4 | -1.272460336 | 0.00027831 |
| NDUFB2 | -0.806123664 | 0.014901469 |
| ICAM1 | 1.81978739 | 0.001972228 |
| THPO | 1.903785316 | 0.040913456 |
| CHRD | 1.658068429 | 0.003331413 |
| CERS4 | 2.952241664 | 0.001264482 |
| EFNB1 | 1.042281417 | 0.001561618 |
| KIF4A | -4.962088909 | 1.72E-10 |
| NAT14 | -0.739794735 | 0.007026414 |
| ITGA6 | -2.658193587 | 8.81E-05 |
| RAPGEF4 | 1.988013325 | 0.025971325 |
| FH | -0.562897188 | 0.037233713 |
| TF | 1.755349787 | 0.034604883 |
| ORC6 | -2.000934176 | 0.001893409 |
| ZC3HC1 | -0.675514063 | 0.02496763 |
| ESR1 | 2.391656484 | 0.000620971 |
| TMEM101 | -0.611814245 | 0.043264865 |

| | | |
|---|---|---|
| SEMA6A | 2.07106006 | 0.005865916 |
| TYRO3 | -1.735528241 | 0.000227362 |
| WDR76 | -1.767891398 | 0.000628466 |
| TBX15 | 0.588559069 | 0.022035386 |
| EZR | -2.467693973 | 1.04E-06 |
| MYL6 | -1.327166757 | 0.00019997 |
| CLSPN | -4.467230901 | 3.61E-09 |
| DPYSL2 | 0.513340785 | 0.047305368 |
| TGFB2 | 1.869612808 | 0.004531594 |
| CDC45 | -4.99051002 | 2.04E-10 |
| CDC6 | -2.352418342 | 0.000151062 |
| FMO2 | 4.24589054 | 0.000155138 |
| NANS | -0.837522322 | 0.007114676 |
| TBC1D2 | -1.114625671 | 0.002352117 |
| TLL2 | -2.858715114 | 3.12E-05 |
| IL11 | -1.652277771 | 0.00014201 |
| FKBP5 | -1.390931759 | 0.01726667 |
| EFHC1 | 0.70940313 | 0.006894407 |
| DSP | -1.628555747 | 0.001844452 |
| ACOT7 | -1.983624251 | 6.24E-06 |
| PCSK5 | 3.102707887 | 3.93E-09 |
| SCD | -0.728789741 | 0.034242853 |
| MYO9B | -0.786305727 | 0.004148964 |
| KCNK6 | -1.573452385 | 0.004648075 |
| FBXL19 | -0.643621168 | 0.009255221 |
| BCL7C | -1.164458695 | 0.000483889 |
| HNRNPM | -1.233833942 | 1.74E-05 |
| POLR2E | -0.769164081 | 0.013338475 |
| IGF2-AS | 1.976826569 | 0.042390621 |
| ARVCF | 1.430236999 | 0.000430995 |
| RANBP1 | -0.8758742 | 0.009800053 |
| SMARCB1 | -0.576703563 | 0.038962131 |
| P2RX6 | 1.615578469 | 0.007993929 |
| DERL3 | -3.94612317 | 0.00311865 |
| CABIN1 | -0.524694815 | 0.033707897 |
| SUSD2 | 1.286411117 | 0.035081894 |
| GGT5 | 2.299713286 | 0.002047493 |
| SEC14L2 | -1.020706902 | 0.002627684 |
| YPEL1 | 1.648437486 | 0.000244414 |
| CARD10 | -1.975892335 | 0.00386553 |
| SLC25A1 | -0.811014118 | 0.037621371 |
| SEZ6L | 3.800570619 | 0.00979352 |

| | | |
|---|---|---|
| LGALS1 | -1.415655118 | 0.000529802 |
| PIK3IP1 | 0.847111001 | 0.003906439 |
| GCAT | -1.088642804 | 0.003169791 |
| MICALL1 | -0.800796794 | 0.012940938 |
| CENPM | -4.987683333 | 3.01E-11 |
| SEPTIN3 | 1.633121638 | 0.024461534 |
| KDELR3 | -1.164084225 | 0.000634969 |
| DMC1 | -2.326296181 | 0.000694827 |
| RAB36 | 1.198797145 | 0.000122071 |
| TIMP3 | -0.86144152 | 0.029007102 |
| SUN2 | -0.912968669 | 0.002753227 |
| CYB5R3 | -0.746802397 | 0.023225964 |
| LMF2 | -0.707392244 | 0.008003248 |
| PACSIN2 | -0.84056079 | 0.017320759 |
| AP1B1 | -0.915195484 | 0.001915309 |
| HMOX1 | -0.895409249 | 0.014743938 |
| MCM5 | -2.627176541 | 5.43E-06 |
| TSPO | -1.460582307 | 7.69E-05 |
| RASD2 | -2.219568944 | 0.012892296 |
| TTLL12 | -0.738138684 | 0.030045806 |
| CBX7 | 1.031763097 | 0.000588219 |
| SYNGR1 | 0.96427827 | 0.028437194 |
| APOL1 | 2.078705792 | 0.002576257 |
| PNPLA3 | -1.652587733 | 0.000109143 |
| MYH9 | -1.276015171 | 0.000664975 |
| KIAA0930 | 0.792381108 | 0.00780026 |
| IL2RB | 2.41202239 | 0.003159212 |
| RBX1 | -0.715233362 | 0.024657669 |
| RANGAP1 | -1.198248417 | 0.000291511 |
| PMM1 | -0.831620531 | 0.002945303 |
| POLE2 | -2.083654615 | 0.004078232 |
| TRIM9 | 2.658455735 | 0.003071338 |
| DDHD1 | 0.645630001 | 0.044523151 |
| CDKN3 | -3.372531327 | 8.22E-08 |
| CGRRF1 | 0.502985866 | 0.044958318 |
| ATP6V1D | -0.642886877 | 0.019802794 |
| PLEK2 | -2.923866006 | 3.72E-07 |
| PIGH | 0.71886797 | 0.010068418 |
| SPTLC2 | -0.954608556 | 0.001232231 |
| RIN3 | -0.67550465 | 0.009055771 |
| SUSD6 | 0.608466778 | 0.017394238 |
| MTHFD1 | -1.077230759 | 0.008709708 |

| | | |
|---|---|---|
| TELO2 | -0.586283499 | 0.0462802 |
| BDKRB1 | -1.852902848 | 0.001040995 |
| GSKIP | -0.553225917 | 0.025087966 |
| CCNB1IP1 | 0.528463237 | 0.033662563 |
| SEC23A | -0.603080125 | 0.034014186 |
| GSS | -0.673068473 | 0.021679875 |
| ABHD12 | -0.966026408 | 0.006520949 |
| GINS1 | -2.63070525 | 7.04E-05 |
| CD40 | 1.700928916 | 0.00050658 |
| SGK2 | 1.965913968 | 0.048499912 |
| MYBL2 | -5.270013186 | 3.63E-12 |
| RAB5IF | -0.55584069 | 0.048937384 |
| TPD52L2 | -0.610159086 | 0.033678319 |
| PSMA7 | -0.675371244 | 0.010489441 |
| NTSR1 | -5.119972296 | 0.003835305 |
| SLC17A9 | -1.750455494 | 1.29E-07 |
| ARFGAP1 | -0.555352174 | 0.03341272 |
| EEF1A2 | -3.730488781 | 7.85E-05 |
| GMEB2 | -0.610987534 | 0.019832901 |
| ADISSP | -1.422999689 | 0.000160246 |
| CDC25B | -1.172511672 | 0.027759496 |
| ISM1 | 1.83884155 | 0.042198981 |
| RNF24 | 0.497827902 | 0.034536599 |
| SEL1L2 | 2.568129879 | 0.028900273 |
| TRIB3 | 1.455480146 | 0.000572668 |
| RASSF2 | 0.876276384 | 0.047133061 |
| MYLK2 | -5.695208357 | 0.003246392 |
| FERMT1 | -1.810503106 | 0.010023891 |
| MYL9 | -1.071685549 | 0.001343761 |
| TLDC2 | -2.058306347 | 0.006783179 |
| SAMHD1 | 0.679156033 | 0.024847493 |
| MROH8 | 1.216198888 | 0.006778255 |
| JAG1 | 2.663458565 | 2.95E-14 |
| SNTA1 | -1.328793356 | 0.00062214 |
| E2F1 | -3.28002045 | 6.37E-08 |
| FAM83D | -5.408858646 | 7.82E-13 |
| SYNDIG1 | -1.103479598 | 0.037934582 |
| TNNC2 | 2.317156138 | 0.00035696 |
| SMAD7 | -1.756747868 | 1.68E-06 |
| CSTF2 | -0.735671453 | 0.0153773 |
| MXRA5 | 0.897322901 | 0.03063472 |
| STS | 0.700981791 | 0.039460109 |

| | | |
|---|---|---|
| SUV39H1 | -1.568863865 | 0.005190009 |
| PLP2 | -1.946689629 | 2.66E-05 |
| ASB9 | -2.400939588 | 0.049301618 |
| KCND1 | -2.001927772 | 0.000246158 |
| SLC35A2 | -0.699491276 | 0.022329691 |
| SMS | -1.957732376 | 4.96E-07 |
| PHEX | 2.415021527 | 0.001018713 |
| MAGED2 | -1.028192815 | 0.001845829 |
| RBM3 | -1.333409262 | 0.000117462 |
| CENPI | -3.152241797 | 6.79E-06 |
| DRP2 | -2.651635556 | 4.76E-05 |
| GLA | -0.614359183 | 0.048907777 |
| BEX4 | 1.554608628 | 2.43E-05 |
| HTR2A | 1.428610415 | 0.014904917 |
| TNFSF13B | 1.717541641 | 0.004151498 |
| CDADC1 | 0.54446635 | 0.041136224 |
| CAB39L | 0.980299076 | 0.000235262 |
| KLF5 | 1.129331028 | 0.025118181 |
| ACP5 | -5.631585215 | 3.44E-07 |
| ARHGEF7 | 0.645578648 | 0.008411926 |
| FGF9 | 2.518114935 | 1.50E-06 |
| SGCG | -1.031510099 | 0.026411861 |
| DGKH | -1.366896723 | 0.006711714 |
| KATNAL1 | -0.672736114 | 0.021664797 |
| MEDAG | 0.911860054 | 0.02153744 |
| CLN5 | 0.787112234 | 0.00333891 |
| OLFM4 | 4.840101215 | 0.024998129 |
| CORO1A | -3.055025704 | 0.000666784 |
| NUTF2 | -0.568315926 | 0.034514875 |
| TSNAXIP1 | 1.021358618 | 0.006510782 |
| PARD6A | 2.213287444 | 0.000393112 |
| MMP15 | -0.970589409 | 0.020783607 |
| NDRG4 | -2.890545883 | 0.00053647 |
| SLC7A6 | -0.625904462 | 0.015549366 |
| PLA2G15 | -0.832652589 | 0.008493927 |
| SEC14L5 | 3.993948461 | 0.000232199 |
| COTL1 | -1.846379958 | 7.50E-09 |
| METRN | -0.640600463 | 0.031493719 |
| PIEZO1 | -0.758688341 | 0.004803426 |
| GSPT1 | -0.672094144 | 0.016386199 |
| ELOB | -0.642121587 | 0.047470981 |
| RRN3P2 | 4.287375335 | 0.016846834 |

| | | |
|---|---|---|
| QPRT | -1.441606539 | 0.047379877 |
| PYCARD | -1.544669422 | 0.003901627 |
| MAZ | -0.848383284 | 0.013137908 |
| IL21R | -2.190285835 | 0.003945482 |
| AQP9 | 3.435253865 | 0.008223259 |
| IQCH | 0.889505399 | 0.043713067 |
| LACTB | -0.92027313 | 0.019086593 |
| CORO2B | 1.404405802 | 1.46E-06 |
| FAH | -0.954918013 | 0.023503379 |
| CEMIP | -1.096746482 | 0.047235794 |
| EHD4 | -1.251889042 | 7.72E-05 |
| TGM5 | 2.387811945 | 0.004252423 |
| ENTREP2 | -4.110160795 | 0.008775401 |
| OIP5 | -4.128341198 | 2.78E-07 |
| PDGFRL | 2.263651716 | 0.04253027 |
| ZDHHC2 | -0.702407181 | 0.022844796 |
| FZD3 | 2.957580875 | 9.04E-07 |
| TRPA1 | 1.438003419 | 0.049032198 |
| POP1 | -1.168484808 | 0.000242783 |
| CCN4 | 1.233834443 | 0.038626312 |
| IL7 | 3.287532505 | 4.72E-08 |
| STMN2 | 0.845459092 | 0.008665607 |
| SNX16 | 0.678598474 | 0.045849374 |
| GFUS | -0.807685811 | 0.00628246 |
| PYCR3 | -1.191656738 | 0.001673668 |
| SQLE | -0.961328131 | 0.022542117 |
| SH2D4A | -1.125812146 | 0.012526902 |
| NEFM | -1.63955822 | 0.013985185 |
| MCM4 | -1.751332228 | 0.000220162 |
| ASAH1 | 0.551549022 | 0.038219294 |
| BNIP3L | 0.754437027 | 0.023684008 |
| MAN2B1 | 0.627761715 | 0.01410918 |
| KCNN4 | -4.540117524 | 2.63E-06 |
| NFKBIB | -0.630460003 | 0.020735864 |
| LIN7B | -1.336535229 | 0.00014601 |
| ERCC2 | -1.115511689 | 0.000387862 |
| DOT1L | -0.706828498 | 0.025903862 |
| PLEKHJ1 | -0.765116249 | 0.004859545 |
| DMPK | -0.897726767 | 0.048501376 |
| SNAPC2 | -0.661288678 | 0.016575412 |
| CCDC61 | -0.621303974 | 0.039482981 |
| ASF1B | -4.207927292 | 3.60E-11 |

| | | |
|---|---|---|
| TNNT1 | -2.257463496 | 0.006646655 |
| OLFM2 | 3.128870867 | 1.26E-17 |
| ILVBL | -0.608088071 | 0.037226526 |
| TBCB | -0.987401921 | 0.001287436 |
| FSD1 | -1.100191186 | 0.006265689 |
| APLP1 | -1.920030865 | 8.65E-05 |
| CCDC9 | -0.603162992 | 0.026815586 |
| FZR1 | -0.673230436 | 0.01923234 |
| TGFB1 | -1.253296674 | 0.000284932 |
| DENND3 | -1.619047505 | 2.73E-05 |
| PLIN3 | -1.1204022 | 0.002254605 |
| ICAM5 | 1.388278915 | 0.012840669 |
| ETFB | -0.973250851 | 0.014079088 |
| CDC37 | -0.703506319 | 0.006308204 |
| RABAC1 | -0.723783003 | 0.03974075 |
| PTPRS | 0.587350998 | 0.016984431 |
| MEGF8 | 0.758532973 | 0.011427866 |
| GRWD1 | -0.707886054 | 0.013272138 |
| GRIN2D | -1.166774302 | 0.003124152 |
| CLEC11A | -1.307680046 | 0.001149065 |
| CARD8 | 0.587891129 | 0.035625597 |
| LIG1 | -1.890279072 | 1.51E-05 |
| DBP | 1.038760805 | 0.003658649 |
| BCAT2 | -1.002644107 | 0.004678457 |
| PPP2R1A | -1.066868088 | 0.001365264 |
| GCDH | -0.68508253 | 0.029047026 |
| UPK1A | 2.427603508 | 0.001514879 |
| COPE | -0.926608096 | 0.005957885 |
| ARMC6 | -0.991474504 | 0.001400716 |
| LSR | -1.082728196 | 0.034840121 |
| ERF | -1.197910483 | 1.43E-05 |
| CADM4 | 1.148697727 | 0.009375677 |
| SMG9 | -0.556151086 | 0.036865391 |
| CFAP69 | 1.209992758 | 5.75E-05 |
| TFPI2 | -3.725530527 | 8.12E-09 |
| HBP1 | 0.572483522 | 0.018426301 |
| PTN | -2.027653459 | 0.002151228 |
| OGDH | -0.78768052 | 0.016261983 |
| CAV1 | -1.607724377 | 0.000372379 |
| MET | -2.449192871 | 0.000104417 |
| HOXA3 | 1.628557695 | 0.010736623 |
| HOXA13 | 2.212419939 | 0.001329698 |

| | | |
|---|---|---|
| STX1A | -0.880882841 | 0.012959887 |
| EPHB6 | 2.730617546 | 8.59E-05 |
| MINDY4 | 1.886647308 | 0.024111552 |
| HSPB1 | -1.450969151 | 6.88E-05 |
| SNX8 | -0.888948141 | 0.000611381 |
| NUDT1 | -0.887619972 | 0.042535522 |
| IMPDH1 | -0.888020505 | 0.001072547 |
| AGFG2 | -0.881736923 | 0.016119114 |
| AP1S1 | -0.7731937 | 0.022028167 |
| PLOD3 | -0.565030352 | 0.045163914 |
| GLCCI1 | 1.359025627 | 2.23E-05 |
| TMEM106B | 0.789528171 | 0.010923928 |
| MEST | 2.450838387 | 2.06E-05 |
| ACTR3C | -1.463707807 | 0.012748718 |
| TSPAN13 | -1.885310896 | 0.00126337 |
| GIMAP2 | 2.581564082 | 1.24E-05 |
| TMEM176B | 5.615792664 | 0.002674821 |
| POLD2 | -0.804763066 | 0.022641474 |
| YKT6 | -0.787125576 | 0.003316715 |
| LIMK1 | -0.768467788 | 0.003724647 |
| NMRK1 | 0.887180845 | 0.005986568 |
| PRUNE2 | -1.485753445 | 0.022983298 |
| TRIM14 | -1.755288635 | 1.32E-05 |
| CORO2A | -3.082476542 | 0.000108427 |
| TGFBR1 | 0.666689648 | 0.033694933 |
| ECM2 | 2.296335167 | 5.30E-06 |
| AKNA | 0.951575282 | 0.000635026 |
| KANK1 | -0.579550573 | 0.049970983 |
| NCS1 | -0.97099802 | 0.00082154 |
| CREB3 | -0.649384811 | 0.006773446 |
| RIGI | 1.726820882 | 0.039895669 |
| PDLIM1 | -1.032417855 | 0.039229392 |
| RASSF4 | 1.601613984 | 2.22E-05 |
| DNMBP | -0.729661945 | 0.01687843 |
| RAB11FIP2 | 0.823437156 | 0.005800821 |
| TRDMT1 | -0.886369132 | 0.002211782 |
| UNC5B | 1.174133544 | 0.000834927 |
| LZTS2 | -0.540972569 | 0.042439611 |
| KAZALD1 | -1.45906152 | 0.000158201 |
| NPM3 | -0.758234951 | 0.032717947 |
| GBF1 | -0.533608718 | 0.017824388 |
| STN1 | 1.682338388 | 1.75E-07 |

| | | |
|---|---|---|
| DKK1 | -4.102822972 | 1.16E-15 |
| TFAM | 0.587057385 | 0.033580884 |
| UBE2S | -1.988965852 | 6.14E-05 |
| DNAJC12 | 1.838005991 | 0.025648055 |
| PPIF | -0.535483054 | 0.036311272 |
| FBXL20 | 0.536979815 | 0.028302051 |
| WNT3 | 0.977793289 | 0.000919324 |
| ASPA | 2.116142167 | 0.000129111 |
| KPNB1 | -0.590877801 | 0.024907929 |
| PNPO | -0.799125104 | 0.019631913 |
| PFN1 | -1.352028622 | 3.56E-05 |
| RASD1 | 1.170767116 | 0.024371882 |
| CHRNE | -1.864088146 | 0.004077075 |
| B9D1 | 0.867878869 | 0.017449409 |
| ASIC2 | 4.088689551 | 0.001491775 |
| CCL7 | 4.563631073 | 0.000755674 |
| CCL2 | 4.95638662 | 3.88E-16 |
| CCL8 | 4.819585428 | 0.001369021 |
| DHX58 | 0.734081575 | 0.037736554 |
| KAT2A | 0.810660816 | 0.010147257 |
| DLX4 | 1.748662131 | 0.018248678 |
| SGCA | 2.244433695 | 0.001041365 |
| MRPL27 | -0.690334764 | 0.037063168 |
| VAT1 | -0.841051911 | 0.004232123 |
| LRRC59 | -1.242388023 | 3.77E-05 |
| HDAC5 | -1.190432957 | 5.86E-05 |
| EFTUD2 | -0.596589789 | 0.029797733 |
| FAM20A | 3.32552678 | 1.77E-08 |
| MAP2K6 | 1.054940086 | 0.008351844 |
| MYH1 | 4.58608634 | 0.006378333 |
| NHERF1 | -1.028471999 | 0.032776934 |
| TMEM104 | -1.042241484 | 0.000664375 |
| CDR2L | -0.938362778 | 0.000112965 |
| CRACD | 3.420294232 | 0.012443402 |
| ELF2 | 0.563568462 | 0.029739658 |
| GAB1 | 1.022291664 | 0.003407146 |
| CPE | 2.062987967 | 0.000792449 |
| SEPSECS | 0.738804093 | 0.01706021 |
| CPZ | 2.486608728 | 2.51E-05 |
| SLC2A9 | 1.98439933 | 1.35E-05 |
| NEIL3 | -2.632185281 | 0.000498629 |
| NSD2 | -0.98377532 | 0.027005079 |

| | | |
|---|---|---|
| GLRB | 1.552185225 | 5.04E-06 |
| LRP2BP | 1.574059491 | 0.008087735 |
| KLF3 | 0.784796105 | 0.026842839 |
| KLHL5 | 0.836568493 | 0.007730144 |
| NCAPG | -4.166057487 | 2.48E-08 |
| PPARGC1A | 3.451172712 | 1.99E-06 |
| CRYAB | -1.069627282 | 0.009302249 |
| CCDC34 | -1.080659058 | 0.01942006 |
| HSPA8 | -0.489607123 | 0.039844841 |
| VWA5A | 1.473666507 | 2.28E-05 |
| SIAE | 0.678416294 | 0.049486242 |
| LPXN | -1.795031162 | 2.87E-05 |
| DTX4 | 1.482147007 | 0.022595851 |
| EHD1 | -0.821962812 | 0.01175233 |
| ST3GAL4 | -0.820997682 | 0.010772706 |
| CPT1A | -0.96641739 | 0.000145181 |
| CCND1 | -1.236932593 | 0.000295922 |
| HPX | -1.993163487 | 0.002194983 |
| ANAPC15 | -1.486999694 | 3.57E-06 |
| FOLR3 | -5.964481882 | 0.00111961 |
| ARHGEF17 | -0.644226626 | 0.033586924 |
| RNF141 | 0.628693852 | 0.028095664 |
| CEP126 | 1.092358504 | 4.07E-05 |
| KIAA1549L | -1.071055559 | 0.028774442 |
| COMMD9 | -0.652024645 | 0.033443757 |
| SLC15A3 | 1.115242159 | 0.00790349 |
| SLC35F2 | -1.489272738 | 0.000242608 |
| PITPNM1 | -0.666598525 | 0.00446827 |
| TCIRG1 | -0.877775499 | 0.003035275 |
| EXPH5 | 2.650199283 | 0.000557974 |
| P3H3 | -1.204755885 | 0.000485908 |
| CLEC2B | 1.689412755 | 0.000275959 |
| SELPLG | -0.943169716 | 0.04422409 |
| CORO1C | -1.070660128 | 1.25E-05 |
| TSPAN11 | -1.430206887 | 0.003130484 |
| MVK | -1.544653729 | 0.000156742 |
| ATP5F1B | -0.797724526 | 0.007629155 |
| LIN7A | 0.978240035 | 0.005458873 |
| ACSS3 | 0.977565176 | 0.014957269 |
| GLI1 | -2.365043507 | 3.45E-06 |
| FOXM1 | -4.06727907 | 2.33E-18 |
| ARPC3 | -0.805004135 | 0.009249387 |

| | | |
|---|---|---|
| RAD51AP1 | -2.619139084 | 5.04E-05 |
| AKAP3 | 1.727700174 | 0.024027607 |
| KCNA1 | 7.748057947 | 5.33E-06 |
| GSG1 | 1.641279807 | 0.006617406 |
| MGP | 1.474787426 | 0.002048701 |
| ARHGDIB | -2.800970388 | 3.32E-06 |
| MDM1 | 0.717410624 | 0.017059571 |
| CNOT2 | 0.589827271 | 0.022403269 |
| TIMELESS | -1.404805941 | 0.007420393 |
| KRR1 | 0.533630352 | 0.046047816 |
| CDCA3 | -4.035342706 | 1.96E-09 |
| USP5 | -0.734763085 | 0.009096706 |
| SPSB2 | -0.940798068 | 0.011671798 |
| ENO2 | 1.078937309 | 0.01198438 |
| NT5DC3 | -1.722765941 | 7.59E-07 |
| LDHB | -0.927120524 | 0.02248991 |
| ST8SIA1 | 1.912890856 | 4.36E-05 |
| DDX12B | -1.622954833 | 0.031402805 |
| BTN3A3 | 1.105177835 | 0.000893082 |
| NEDD9 | 1.917678395 | 9.79E-05 |
| MAN1A1 | 1.178312823 | 0.000620531 |
| HINT3 | 0.948373323 | 0.001328651 |
| NCOA7 | 1.357144321 | 0.001869788 |
| RIPOR2 | -1.714224649 | 0.000467322 |
| ULBP1 | 0.771793098 | 0.048951674 |
| MCM3 | -1.067220565 | 0.029905387 |
| SIM1 | 1.532922949 | 0.016853112 |
| WASF1 | 0.643540893 | 0.044865219 |
| GPLD1 | 2.233268188 | 0.00946699 |
| ALDH5A1 | 1.321071329 | 0.004595212 |
| GMNN | -1.10817045 | 0.019796817 |
| ARFGEF3 | 3.034042002 | 1.15E-13 |
| CUTA | -1.00854137 | 0.005689598 |
| SMOC2 | 3.388170936 | 1.47E-09 |
| CCND3 | -1.368165327 | 6.44E-06 |
| DUSP22 | 0.69414854 | 0.018820262 |
| COX7A2 | -0.685606698 | 0.048123052 |
| VEGFA | 2.039391998 | 1.64E-07 |
| TTK | -4.974294973 | 3.80E-05 |
| LAMA4 | 1.244204198 | 0.000188821 |
| ENPP5 | 1.970235563 | 0.003309197 |
| PCDHB2 | 1.281969221 | 0.000135498 |

| | | |
|---|---|---|
| GHR | 1.541932572 | 0.000464655 |
| DAP | -0.783105784 | 0.030605553 |
| KIF20A | -5.599062973 | 5.78E-14 |
| PCDHB6 | 1.453518063 | 0.044138373 |
| PCDHB15 | 1.577562329 | 0.000533376 |
| RNF130 | 1.133129039 | 2.08E-05 |
| RASGRF2 | 2.332492487 | 0.037678497 |
| LMNB1 | -3.778515453 | 2.78E-08 |
| ARRDC3 | 1.501459858 | 9.63E-09 |
| RAD1 | 0.697880557 | 0.012186256 |
| PRLR | -3.008477767 | 3.19E-05 |
| SLC12A7 | 0.927899178 | 0.020194962 |
| RARS1 | -0.709788928 | 0.008748095 |
| WWC1 | -1.956544304 | 0.00051461 |
| PDGFRB | 0.591206482 | 0.016288986 |
| CDX1 | 2.934202939 | 0.001159875 |
| DBN1 | -0.841705508 | 0.008392246 |
| SMC4 | -1.160972149 | 0.036111427 |
| CRBN | 0.487186378 | 0.04480667 |
| BCL6 | 1.53877112 | 4.10E-09 |
| ARL6 | 0.768868555 | 0.009917463 |
| FAM162A | 0.899390835 | 0.002767261 |
| RBP1 | 2.653801453 | 0.001377811 |
| SLC25A36 | 0.841333036 | 0.029411098 |
| RNF7 | -0.681615341 | 0.03535486 |
| TFDP2 | 0.806571661 | 0.004252139 |
| WNT5A | 1.074989161 | 0.00800593 |
| COL7A1 | -1.450027388 | 0.010996172 |
| PRKAR2A | -0.58572447 | 0.013358993 |
| ECT2 | -1.824031447 | 0.002026296 |
| GNAI2 | -0.863531565 | 0.003050304 |
| HYAL1 | -1.70457016 | 0.027164399 |
| CYB561D2 | -0.687689514 | 0.036112314 |
| C3orf52 | -2.198490592 | 4.49E-05 |
| PLSCR4 | 1.785700835 | 9.87E-06 |
| ACVR2B | 0.867954858 | 0.038194755 |
| EIF1B | 0.761874951 | 0.001827236 |
| KLHL24 | 0.88993829 | 0.03252355 |
| SSR3 | -0.690924783 | 0.013059882 |
| EIF4G1 | -0.599372123 | 0.017954002 |
| IL1A | 4.55961911 | 0.039866661 |
| ACTR3 | -1.08261811 | 9.04E-06 |

| | | |
|---|---|---|
| STEAP3 | -1.170973612 | 0.023944116 |
| EPB41L5 | 1.004311195 | 0.000385665 |
| TP53I3 | -1.606837402 | 4.86E-05 |
| DNAJC27 | 0.75359026 | 0.013777007 |
| POMC | -1.552541936 | 0.022197121 |
| OTOF | -5.457238235 | 0.002721826 |
| CENPA | -5.015293581 | 2.65E-10 |
| ACVR1 | 0.551196387 | 0.022466245 |
| SNX17 | -0.640060466 | 0.036699854 |
| PDE1A | 1.272628672 | 0.007501687 |
| PCSK4 | 1.025416374 | 0.018418068 |
| IFIH1 | 1.458337542 | 0.002219123 |
| GCA | 1.300366587 | 0.021946498 |
| NDUFS7 | -0.687715025 | 0.035336478 |
| GRB14 | -2.35167866 | 0.034109133 |
| TLX2 | -1.637202983 | 0.030766201 |
| DOK1 | -0.784773506 | 0.002032531 |
| POLE4 | -1.127661053 | 0.012953781 |
| TACR1 | -4.195719901 | 0.010695829 |
| CCDC88A | -0.697649098 | 0.042413858 |
| EFEMP1 | 1.272619562 | 0.0162507 |
| STAT1 | 0.941007827 | 0.005530661 |
| GLS | -0.895097135 | 0.025079208 |
| IGFBP2 | 2.902802601 | 6.69E-11 |
| ELMOD3 | 0.597370502 | 0.016510213 |
| IGFBP5 | 4.526198792 | 2.82E-12 |
| CCT4 | -0.601351739 | 0.031413633 |
| PDCL3 | -0.56259397 | 0.044994777 |
| IL1R1 | 1.233891057 | 0.004148527 |
| IL1RL1 | 6.495000178 | 2.92E-25 |
| IL18R1 | 4.685396736 | 3.77E-14 |
| FHL2 | -0.983418461 | 0.001476709 |
| MLPH | -3.81209475 | 2.95E-08 |
| STK25 | -0.489988899 | 0.036985063 |
| PROC | -4.014544781 | 0.001954619 |
| DLX2 | -1.768127441 | 0.015548272 |
| SDC1 | -0.886383889 | 0.004721856 |
| PLCL1 | 1.413421767 | 0.006343996 |
| SOS1 | 0.899166138 | 0.00507416 |
| EPAS1 | 1.184584984 | 0.00033743 |
| ARID3A | -0.608046932 | 0.043497188 |
| NFE2L2 | 0.753679761 | 0.002443102 |

| | | |
|---|---|---|
| PARD3B | 1.770146524 | 5.59E-09 |
| MARK1 | 1.765561885 | 0.001323803 |
| PAPPA2 | 1.706358276 | 0.012779454 |
| RALGPS2 | -1.298985914 | 6.97E-05 |
| ANGPTL1 | 2.609770212 | 3.13E-13 |
| ERRFI1 | 1.089607067 | 0.005646603 |
| AMPD2 | -0.67474245 | 0.014441611 |
| KCNC4 | -1.165172294 | 0.014207132 |
| WDR77 | -0.786876634 | 0.005130921 |
| HDAC1 | -0.730966294 | 0.04522154 |
| ARHGEF2 | 0.820345399 | 0.007670441 |
| SRM | -0.637050344 | 0.03025381 |
| LEPR | 1.297833115 | 0.002523535 |
| PRG4 | -3.401400683 | 0.000843725 |
| NCF2 | -2.160844648 | 0.004537815 |
| GADD45A | 1.019151973 | 0.006526804 |
| WLS | 0.701072337 | 0.022687412 |
| AGMAT | -2.124213802 | 2.61E-06 |
| OLFML3 | -1.044602776 | 0.004557376 |
| MAP7D1 | -0.66960825 | 0.004597458 |
| NID1 | 0.883387926 | 0.012436674 |
| SIPA1L2 | 2.330488888 | 2.68E-07 |
| ZNF684 | 0.797655292 | 0.020535369 |
| RIMS3 | 4.487688644 | 1.16E-14 |
| ST6GALNAC5 | -2.052228431 | 7.92E-06 |
| ADGRL2 | 0.482707767 | 0.03933255 |
| PADI2 | 3.565594996 | 0.006151741 |
| UAP1 | -0.752885735 | 0.021877418 |
| CTBS | 0.558385421 | 0.036349377 |
| RGS4 | -1.561553049 | 0.010322398 |
| SSX2IP | -1.876428685 | 4.37E-06 |
| GBP3 | -1.021578678 | 0.030886011 |
| KIF17 | -2.461401952 | 0.007259688 |
| GPR89A | -0.567479918 | 0.046817059 |
| CDK18 | 2.513946093 | 1.35E-05 |
| GALE | -1.367251398 | 0.000130339 |
| ID3 | -2.431103725 | 7.79E-06 |
| EBNA1BP2 | -0.717511989 | 0.030432931 |
| CDC20 | -5.98354852 | 2.85E-15 |
| ATP6V0B | -0.704287535 | 0.022930343 |
| PIK3R3 | -1.395418073 | 0.022443736 |
| VAMP4 | 0.934865416 | 0.000553855 |

| | | |
|---|---|---|
| TNFSF4 | 2.011850086 | 2.52E-05 |
| DARS2 | -0.919316114 | 0.00521871 |
| HSD11B1 | 2.317900245 | 0.041555474 |
| UTP25 | -0.668170117 | 0.023386638 |
| RCAN3 | -1.611339146 | 1.27E-05 |
| STMN1 | -2.981486759 | 1.85E-09 |
| MAN1C1 | 1.65648191 | 0.000394985 |
| NEK2 | -6.072539618 | 1.01E-06 |
| DHDDS | -0.695648256 | 0.015751434 |
| CENPF | -3.63054495 | 2.39E-07 |
| USP35 | -0.679597426 | 0.020209499 |
| SGK1 | -0.764387517 | 0.004039707 |
| MYL12B | -0.817450981 | 0.011286442 |
| SPP1 | -2.864450072 | 0.000702974 |
| CCNI | 0.529926803 | 0.041123945 |
| PPL | 4.428049338 | 3.76E-13 |
| ELL2 | -0.949901929 | 0.002242012 |
| DNAH7 | 1.419039344 | 0.022584301 |
| NDUFB3 | -0.741469771 | 0.035530758 |
| UBE2B | 0.531160093 | 0.034834188 |
| KLF9 | 1.233976824 | 0.001223889 |
| PIGZ | 0.815812395 | 0.003259305 |
| DYNC2I2 | -0.958034391 | 0.016142866 |
| PTPA | -1.011922844 | 0.002711828 |
| PHF19 | -1.067699372 | 0.017130915 |
| DCAF4 | -0.596718736 | 0.019564824 |
| IFI27L2 | -0.978111947 | 0.033572924 |
| LTBP2 | -0.985399127 | 0.008826709 |
| TMEM214 | -0.583961915 | 0.041870123 |
| FKBP1B | -2.359604063 | 5.51E-06 |
| FAM98A | -0.856633168 | 0.001544197 |
| BCL11A | 1.913144349 | 9.21E-05 |
| OGFRL1 | 1.028883157 | 0.003117074 |
| TECTB | -7.458081511 | 2.42E-06 |
| IFIT2 | 1.014019146 | 0.015160103 |
| CUTC | 0.959588395 | 0.006071816 |
| MXI1 | 1.760633789 | 1.32E-11 |
| HELLS | -2.564681122 | 5.24E-06 |
| HOXB5 | -0.793619445 | 0.033111028 |
| MSX2 | 1.428471116 | 0.032186454 |
| CCDC170 | 1.48219043 | 0.001523274 |
| PCMT1 | -0.751957294 | 0.004788801 |

| | | |
|---|---|---|
| PLEKHG1 | 3.291791571 | 2.09E-13 |
| PCDHB10 | 1.361283707 | 0.001234162 |
| PCDHB14 | 1.205513374 | 0.00160874 |
| PCDHB12 | 1.453974798 | 0.027316709 |
| TNFSF18 | 3.901307467 | 0.000165199 |
| ACAT2 | -2.039140518 | 8.68E-07 |
| TCP1 | -0.49926336 | 0.040048891 |
| KIAA1217 | -1.283301922 | 0.009349135 |
| EPC1 | 0.548899399 | 0.044541911 |
| ENOX1 | 1.207763333 | 0.00712723 |
| TGFBI | -1.310546909 | 0.003757445 |
| EGR1 | 2.25021991 | 0.047127974 |
| SOCS2 | -0.794968777 | 0.013413747 |
| TNFRSF10B | 0.962332204 | 0.000227816 |
| RNF170 | 0.682764386 | 0.012616335 |
| ZNF706 | 0.569095747 | 0.025172544 |
| RDH10 | 1.704461641 | 0.002063765 |
| NCAPH | -4.485571472 | 2.48E-09 |
| KCNJ8 | -0.858132659 | 0.039837163 |
| POPDC2 | 1.958888327 | 0.00087797 |
| KIF18A | -3.877973599 | 7.62E-08 |
| CRY2 | 0.729114892 | 0.037308787 |
| PILRB | -1.977780575 | 0.00284907 |
| ADGRB2 | 0.845917974 | 0.003882835 |
| TNFSF10 | 5.442129616 | 5.32E-14 |
| CSMD2 | -2.587150549 | 2.40E-10 |
| CLCC1 | 0.741319022 | 0.005138133 |
| GTDC1 | 1.088072795 | 0.000222596 |
| ACVR2A | 1.432118605 | 0.00338823 |
| XPNPEP2 | 1.987843776 | 1.46E-05 |
| FMOD | 1.650658749 | 0.000115645 |
| COPA | -0.500273808 | 0.035303781 |
| HS3ST2 | -2.860145919 | 0.048365604 |
| ANXA11 | -0.797965587 | 0.021874255 |
| LDB3 | 2.622944757 | 0.000296732 |
| SHLD2 | -0.533266264 | 0.030103269 |
| PTGFR | 1.661700257 | 0.000109598 |
| CCDC18 | -1.438911087 | 0.019052796 |
| HNRNPA2B1 | -0.757979713 | 0.023560627 |
| ARL4A | -1.00687037 | 0.006843837 |
| GLIPR2 | -0.878601634 | 0.000630587 |
| CLTA | -0.818928162 | 0.005280467 |

| | | |
|---|---|---|
| PLAU | 2.084754546 | 7.42E-05 |
| SRGN | -2.241582625 | 0.004742861 |
| EGR2 | 4.767269181 | 0.00544485 |
| ECD | -0.519100794 | 0.040352118 |
| P4HA1 | 1.071567915 | 0.002596248 |
| ZWINT | -3.576029211 | 1.44E-09 |
| CIT | -3.396817794 | 4.84E-07 |
| CDKN2C | -0.85832301 | 0.044227198 |
| ACOT9 | -0.769273779 | 0.002744012 |
| DDX39A | -1.605787291 | 9.50E-05 |
| GIPC1 | -1.027476771 | 0.000452797 |
| NLN | -0.652557728 | 0.009832399 |
| CENPK | -2.918128997 | 3.10E-05 |
| ITIH5 | 2.487327394 | 0.000174705 |
| ORMDL2 | -1.448173488 | 3.77E-06 |
| PDE1B | -3.065582818 | 0.044136979 |
| CDK2 | -1.204011685 | 0.002744935 |
| TUBA1B | -2.223308627 | 6.25E-10 |
| SARDH | -1.435538696 | 0.045971308 |
| STIL | -2.135263831 | 0.00022603 |
| HJURP | -4.854075169 | 4.31E-10 |
| IL13RA2 | -2.948522495 | 8.53E-05 |
| AMD1 | -0.615366212 | 0.029434985 |
| MORF4L2 | -0.50961552 | 0.032319866 |
| NMI | 0.875753629 | 0.021541411 |
| TNFAIP6 | 1.406998883 | 0.043428069 |
| EXOSC9 | -1.155163948 | 0.00040751 |
| PLA2G12A | 0.636828866 | 0.017368629 |
| CKS2 | -2.840034328 | 1.96E-06 |
| INHA | 1.378987518 | 0.049374019 |
| PARD6B | 3.229857653 | 0.0094344 |
| SNAI1 | -1.919206779 | 1.73E-05 |
| KCNK15 | 1.610057602 | 0.00343317 |
| PEPD | -0.707541515 | 0.040444948 |
| STAMBP | 0.640481558 | 0.023311096 |
| NAGK | -0.661792158 | 0.011470009 |
| F13A1 | 5.161651782 | 7.78E-05 |
| TRERF1 | -0.891026182 | 0.018294742 |
| RRP36 | -0.773858807 | 0.00468165 |
| SNRPC | -0.748908814 | 0.012175366 |
| ZNF391 | 1.160750378 | 0.035468156 |
| COL21A1 | 3.489720081 | 9.73E-07 |

| | | |
|---|---|---|
| SOX4 | 1.26792268 | 0.023151173 |
| CPNE5 | -2.313959496 | 6.45E-05 |
| NRN1 | 1.009562768 | 0.021421357 |
| RPP40 | -0.76030089 | 0.043007088 |
| AHNAK | -0.750752473 | 0.008571513 |
| SH3TC1 | 2.708066832 | 0.037292479 |
| BBS2 | 0.591516174 | 0.038379321 |
| GOT2 | -1.053291745 | 0.000362674 |
| DOK4 | -0.714570413 | 0.011947797 |
| CLYBL | 1.245445757 | 0.013850378 |
| HROB | -1.67917358 | 0.001480816 |
| IRF1 | 0.877665501 | 0.032624544 |
| BMP4 | 3.23884 | 8.87E-06 |
| PTGER2 | -2.711604714 | 7.67E-06 |
| SLC25A35 | -0.809477835 | 0.012237373 |
| MIF4GD | 1.085183394 | 0.00044321 |
| MBOAT7 | -0.64306397 | 0.038500924 |
| INSIG2 | 1.147853722 | 0.000771065 |
| PSD4 | -1.863498705 | 2.40E-06 |
| C3 | 2.592649845 | 2.25E-07 |
| EML2 | -0.555266886 | 0.045980893 |
| VASP | -1.480492171 | 1.08E-06 |
| NAPB | 0.879522309 | 0.00316926 |
| CENPB | -0.756901095 | 0.009595456 |
| DTD1 | -0.684932495 | 0.013395005 |
| RBCK1 | 0.894279496 | 0.030721348 |
| TMX4 | 0.750844595 | 0.005663529 |
| SNRPB | -1.07534046 | 0.000655207 |
| FLRT3 | 1.298410631 | 0.042806953 |
| PCSK2 | 2.354494046 | 0.015919231 |
| MKKS | 0.704858739 | 0.020826913 |
| ITPA | -1.042335765 | 0.003195055 |
| MCM8 | -1.289849145 | 0.034447978 |
| FAM110A | -0.889383403 | 0.044088542 |
| NCLN | -0.913439824 | 0.008822109 |
| ZNF436 | 1.481659329 | 4.21E-06 |
| ID1 | -3.57172646 | 1.67E-06 |
| RALY | -0.573031182 | 0.039780858 |
| ROMO1 | -0.867421609 | 0.027501693 |
| KLC1 | -0.72472459 | 0.007053202 |
| XRCC3 | -1.212793636 | 0.018167713 |
| CAPNS1 | -1.002472062 | 0.006618625 |

| | | |
|---|---|---|
| RBM42 | -0.670367132 | 0.01063168 |
| FRMD8 | -1.203999561 | 1.70E-05 |
| BCL2L12 | -1.054040168 | 0.013294838 |
| SCAF1 | -0.499027881 | 0.042422335 |
| ASL | -1.708093536 | 0.00027495 |
| WNK4 | -3.15726637 | 1.47E-08 |
| GLIS2 | 0.602356759 | 0.038409413 |
| TIMM17B | -0.578615776 | 0.041427494 |
| L3HYPDH | 1.02710477 | 0.011410708 |
| OMD | 1.623273766 | 0.006908957 |
| FGD3 | 4.532495195 | 0.019975026 |
| BCL11B | 1.520561536 | 0.010479792 |
| TRAF2 | -0.924494346 | 0.006399791 |
| ABHD8 | 0.853447367 | 0.003871914 |
| TSPAN8 | 2.776243216 | 1.75E-08 |
| RAB3IP | 1.197502529 | 0.0029789 |
| LRRC61 | -1.685997151 | 0.004364032 |
| IDUA | 0.935274773 | 0.002758813 |
| FGFRL1 | -1.55432083 | 6.12E-05 |
| AUNIP | -3.341412709 | 0.000477506 |
| PIN1 | -0.849730718 | 0.007388136 |
| PKMYT1 | -4.878423746 | 1.53E-12 |
| CHTF18 | -1.687832157 | 0.005959151 |
| TUBBP1 | -1.074398536 | 0.045571502 |
| SMARCA4 | -0.731688947 | 0.010680722 |
| IL17B | -2.617554366 | 0.012675804 |
| TUBA4A | -1.468473317 | 0.040473662 |
| ZNF835 | 2.852733922 | 0.020226884 |
| GNG11 | -1.321290581 | 4.47E-05 |
| HIP1 | -1.08287497 | 0.000165906 |
| FGL2 | -2.839423544 | 3.57E-06 |
| STEAP4 | 12.32752715 | 1.80E-05 |
| CASD1 | 0.673577782 | 0.034706432 |
| RASL11B | -4.137226798 | 0.001116218 |
| KDR | -2.340144571 | 0.015996154 |
| ADM2 | 0.949555105 | 0.009771655 |
| ASPHD2 | 1.031006059 | 0.003577818 |
| SDF2L1 | -1.126531151 | 0.004104115 |
| YWHAH | -1.44123974 | 2.16E-06 |
| MGAT3 | 3.382030643 | 0.0477556 |
| CDC42EP1 | -0.780005317 | 0.012617696 |
| TPST2 | -1.606167957 | 0.000596643 |

| | | |
|---|---|---|
| BAIAP2L2 | -2.133145662 | 3.69E-07 |
| RAC2 | -2.039314615 | 1.06E-06 |
| LIF | 3.990520166 | 0.000100866 |
| RIBC2 | -3.634226309 | 0.002867394 |
| SPECC1 | -1.866373522 | 2.86E-08 |
| CPA4 | 1.382160199 | 0.035181882 |
| DOCK4 | 1.226945207 | 0.02811776 |
| VGF | -6.254420855 | 0.000228017 |
| PODXL | -4.784796227 | 3.18E-11 |
| FOXP2 | 1.574270294 | 0.001645417 |
| DNAJB9 | 1.587699969 | 8.49E-06 |
| FLNC | -1.277974771 | 4.86E-05 |
| LRRC4 | -3.315382819 | 2.15E-07 |
| SMO | 1.184254095 | 5.50E-06 |
| MRPS12 | -0.862848191 | 0.010054759 |
| HOXD1 | -1.624705143 | 0.011631228 |
| HERC2 | -0.615900223 | 0.033925581 |
| CGNL1 | 2.051974409 | 0.00027608 |
| KNSTRN | -1.388210247 | 0.011133618 |
| CLN6 | -0.774277608 | 0.025164194 |
| ISLR | -1.17953949 | 0.000235539 |
| MBD4 | 0.632877299 | 0.011184144 |
| E2F8 | -5.106578813 | 3.97E-08 |
| PIMREG | -5.462871726 | 1.97E-12 |
| TXNDC17 | -0.625990388 | 0.035363075 |
| FXR2 | -0.492251301 | 0.031321127 |
| KIF1C | -0.74475151 | 0.004448442 |
| MPDU1 | -0.709359209 | 0.038660889 |
| DNAAF11 | 1.123614774 | 0.00072945 |
| CDKN2D | -1.399456811 | 0.000708736 |
| EGLN3 | 3.720100052 | 0.019065092 |
| EPB41L4A | 1.90702221 | 0.0078197 |
| QRICH2 | 1.072691279 | 0.010872052 |
| ARHGEF6 | 1.08785426 | 0.005977848 |
| SGO1 | -5.13113807 | 8.52E-09 |
| KLF16 | -0.975335254 | 0.000514572 |
| PGAP6 | -0.896554862 | 0.004188012 |
| DOHH | -0.784563524 | 0.008952413 |
| SHC2 | 5.001281124 | 2.36E-07 |
| PHF10 | 1.217685076 | 0.000274833 |
| PRRG3 | 3.214600978 | 0.000114666 |
| GALNT8 | 4.092916832 | 0.030225104 |

| | | |
|---|---|---|
| CRACR2A | -1.051493952 | 0.000687261 |
| APOE | -1.467973558 | 0.016936728 |
| TOMM40 | -0.857744116 | 0.004664383 |
| APOC1 | -2.877905299 | 0.044894977 |
| GADD45G | 1.210576444 | 0.046719852 |
| COLGALT1 | -0.711975193 | 0.014070222 |
| LSM7 | -0.856248431 | 0.008324566 |
| ACSBG2 | 4.777127768 | 0.020548351 |
| ACTN4 | -1.465475455 | 6.44E-08 |
| ARPC1B | -1.442017294 | 0.000138333 |
| KLHDC7B | 3.286249006 | 1.02E-07 |
| GDF15 | 2.164902966 | 2.31E-08 |
| LSM4 | -1.081258775 | 0.002698486 |
| TRPM4 | -0.911374862 | 0.011315274 |
| PAK4 | -0.848425954 | 0.010622907 |
| ASS1 | 1.558749255 | 0.000698466 |
| FIBCD1 | -3.528189871 | 0.008812973 |
| TRIM28 | -0.766225163 | 0.006115597 |
| YIPF2 | -0.76315817 | 0.021722559 |
| TMEM160 | -1.403266705 | 0.001383144 |
| NPAS1 | -1.672805339 | 0.001863134 |
| SESN2 | 0.952424594 | 0.001945036 |
| ATP5IF1 | -0.713911064 | 0.014580307 |
| DNMT1 | -1.227137283 | 0.002161769 |
| SLC6A8 | 0.472387251 | 0.049690207 |
| PNCK | 2.493552049 | 0.015135806 |
| PLXNA3 | -0.671238788 | 0.015539172 |
| MPP1 | -0.704336198 | 0.025516904 |
| CASZ1 | -2.738125982 | 0.001420935 |
| HABP4 | -1.172684925 | 0.000699674 |
| UBA1 | -0.734527743 | 0.007507231 |
| POLN | 1.291557454 | 0.039443792 |
| ULBP2 | -0.728809011 | 0.02113198 |
| AKAP12 | -1.186936125 | 0.012486696 |
| SYNE1 | -0.797794426 | 0.016391133 |
| ULBP3 | -1.591034265 | 0.001237182 |
| EPS8L1 | -2.208420679 | 0.004917953 |
| EDA2R | 0.798126478 | 0.001687696 |
| C1QL1 | -2.495837493 | 8.24E-07 |
| GINS2 | -3.539881721 | 2.29E-06 |
| CHMP1A | -0.61418277 | 0.026956955 |
| CAP1 | -1.193899495 | 9.47E-06 |

| | | |
|---|---|---|
| PPT1 | -1.27610516 | 0.000110536 |
| RAB11FIP4 | 1.305887284 | 0.047235553 |
| SLC6A6 | 1.346985826 | 0.000766296 |
| KCNC3 | 2.833811036 | 0.007956006 |
| PDLIM4 | -0.613848771 | 0.022325971 |
| TUBG1 | -0.884650533 | 0.006952655 |
| PSMC3IP | -1.2565682 | 0.025885436 |
| AOC3 | 4.591312414 | 7.95E-08 |
| ACLY | -0.874179193 | 0.003329637 |
| RAMP2 | -1.415671002 | 0.040594261 |
| AOC2 | 1.059162625 | 0.049377579 |
| DIAPH1 | -1.590351203 | 1.17E-07 |
| ACAP3 | -0.705367834 | 0.026006022 |
| NINJ1 | 0.959026737 | 0.000755147 |
| MAP1B | -1.051306492 | 0.000931664 |
| KRT34 | -2.263430569 | 0.002125039 |
| KRT33B | -3.187059652 | 0.032845328 |
| TOP2A | -4.661284433 | 2.61E-05 |
| PIAS3 | -0.606332992 | 0.03329052 |
| PDHA1 | -0.627778053 | 0.018471261 |
| SELENOS | -0.586512038 | 0.033522815 |
| RHPN2 | 0.915213182 | 0.037737822 |
| ZSWIM4 | 1.267059504 | 0.004479014 |
| SPATA6 | 1.124058767 | 0.00015065 |
| PPARG | 2.307773471 | 0.002322784 |
| FCRLA | -7.913346835 | 1.22E-08 |
| TRIM22 | 0.790973223 | 0.016737869 |
| PER2 | 1.420204773 | 3.44E-07 |
| PTPRE | 2.548427605 | 0.000195264 |
| RAN | -0.642994241 | 0.03636776 |
| MYBBP1A | -0.497821635 | 0.037677835 |
| TMEM128 | 0.52833842 | 0.033397399 |
| COQ3 | -0.918843902 | 0.010320101 |
| PNISR | 0.911250744 | 0.011703236 |
| EIF5A | -1.338945886 | 0.000107533 |
| KDM6B | 1.32466112 | 0.000164496 |
| SLC52A1 | 1.745916689 | 0.040273118 |
| XAF1 | 1.165356911 | 1.28E-05 |
| RGS22 | 5.890459431 | 5.95E-06 |
| MATN2 | -1.605263473 | 0.028282233 |
| MTSS2 | 0.717386522 | 0.032280851 |
| PCED1A | 0.77651463 | 0.011861128 |

| | | |
|---|---|---|
| SNAP25 | -4.070365879 | 0.023402566 |
| NES | -2.565270361 | 6.97E-07 |
| CRP | 4.743051796 | 0.020968214 |
| DPH2 | -0.540469553 | 0.035739132 |
| DMGDH | 1.935287659 | 7.76E-09 |
| ZBED3 | 1.347487465 | 0.000137427 |
| KANK4 | -3.713562548 | 0.031818831 |
| FBXO44 | 0.770418825 | 0.023380516 |
| MTUS2 | -1.828660252 | 0.001331003 |
| USPL1 | -0.611832643 | 0.035559739 |
| HMGB1P5 | -1.320595328 | 0.000337224 |
| RNF17 | 4.853174722 | 0.018301662 |
| MYH8 | 5.174761925 | 0.004286203 |
| MYBPH | -4.343591859 | 0.00715082 |
| PIK3C2B | 0.99349323 | 0.014107987 |
| DCLK1 | 1.701342218 | 0.018974559 |
| EPSTI1 | 1.460891151 | 0.024925989 |
| POSTN | -1.827899159 | 0.012117614 |
| TPT1 | 0.645663288 | 0.017922791 |
| STARD13 | 0.802401627 | 0.004888049 |
| RNF128 | -2.315818574 | 0.039058986 |
| BEX1 | -6.699846516 | 0.019452025 |
| HSPBP1 | -0.840775047 | 0.007639328 |
| CNDP2 | -0.691454139 | 0.006536179 |
| PLAAT4 | 3.211305763 | 1.24E-06 |
| LARGE1 | -1.226415357 | 0.026192423 |
| GSTT2B | -1.900377594 | 0.001530057 |
| C1QTNF6 | 0.901264216 | 0.000113334 |
| GIMAP6 | 5.519671918 | 0.002217439 |
| KRBA1 | -0.75050496 | 0.019617808 |
| BTG1 | 0.623968867 | 0.027786626 |
| TMEM254 | 0.572767052 | 0.038194409 |
| TMTC1 | 2.682516586 | 5.99E-08 |
| E2F5 | 1.043845516 | 0.006796558 |
| MICAL2 | -0.998602879 | 0.030789544 |
| SARAF | 0.651498962 | 0.012181141 |
| GSC | 2.197806428 | 5.54E-05 |
| CCNB1 | -4.306525321 | 3.16E-10 |
| CDK7 | -0.940411293 | 0.011476733 |
| CNTN6 | 4.275687036 | 0.01450643 |
| CHL1 | 2.537911074 | 0.013762912 |
| PSRC1 | -2.296535685 | 5.65E-06 |

| | | |
|---|---|---|
| PTPN22 | -2.130672894 | 0.001110179 |
| PTGFRN | 1.356010864 | 0.000759341 |
| CD101 | 2.765484742 | 0.007138747 |
| FKBP11 | -0.879318644 | 0.024652788 |
| PLEKHA8P1 | -0.890879188 | 0.030796159 |
| YWHAQ | -0.872708578 | 0.012533062 |
| GRHL1 | 3.957390397 | 2.09E-15 |
| SAA2 | 4.202632304 | 0.045387301 |
| ANO3 | -2.441414433 | 0.001337614 |
| IL6ST | 1.361572733 | 4.67E-06 |
| TIMM17A | -0.764098976 | 0.007003584 |
| ANKRD16 | 0.666347693 | 0.020526149 |
| IL15RA | 2.206923882 | 0.000234563 |
| CABLES1 | -1.112552397 | 0.002538157 |
| DOCK2 | -3.855149623 | 5.21E-07 |
| KLRD1 | 2.446908974 | 0.003436264 |
| LRP4 | -1.064271772 | 0.010110728 |
| DDB2 | 0.902329829 | 0.005342784 |
| RTL8C | -1.090991547 | 0.002941221 |
| SPOCD1 | -2.831200765 | 1.54E-08 |
| CDCA8 | -4.771711441 | 4.27E-11 |
| BTF3L4 | 0.584776489 | 0.028188762 |
| DTNA | 2.275293405 | 2.12E-06 |
| APLNR | -3.509986794 | 0.000234861 |
| C5AR2 | -2.234472291 | 0.013394698 |
| UBAC2 | 0.818243253 | 0.00333809 |
| CARS2 | -0.770149918 | 0.017758694 |
| ADAMTS8 | 4.477037542 | 0.000208001 |
| SLC37A2 | -3.178504147 | 0.00016177 |
| NREP | -0.908396115 | 0.024185161 |
| OSTF1 | -0.728520494 | 0.022723424 |
| ISCA1 | 0.507751349 | 0.026817608 |
| ADAM19 | -1.684069699 | 0.000138421 |
| USP30 | 0.519339647 | 0.044492263 |
| TBX3 | -1.730214509 | 0.000111332 |
| CCDC146 | 1.514271907 | 4.93E-05 |
| MRAP2 | -4.356709303 | 1.85E-05 |
| LCA5 | 0.775023268 | 0.010058479 |
| PRR5L | -1.16623674 | 0.015711063 |
| LMO2 | 1.225409427 | 0.029378564 |
| PHF21A | 0.96064826 | 0.000411083 |
| GDF11 | -0.59050373 | 0.021608498 |

| | | |
|---|---|---|
| TROAP | -5.942962915 | 9.50E-15 |
| FAIM2 | 1.142572872 | 0.004095917 |
| ESPL1 | -4.266094601 | 4.80E-09 |
| KRT7 | -1.571586516 | 0.002127323 |
| HNRNPA1 | -0.871567671 | 0.001851402 |
| PKIB | 2.629417537 | 0.023368129 |
| MICAL1 | -1.130114178 | 0.000795598 |
| PRADC1 | -1.152030091 | 0.000971439 |
| CCT7 | -0.698204327 | 0.019268869 |
| RAB11FIP5 | -0.773883345 | 0.00211496 |
| DYSF | -0.903533702 | 0.021411262 |
| KCNMB4 | 3.136620647 | 1.05E-19 |
| MDM2 | 0.718310489 | 0.002377846 |
| FHOD1 | -0.816493005 | 0.011888758 |
| STX6 | 0.695669328 | 0.019718679 |
| RNASEL | 0.490255428 | 0.043903406 |
| NIBAN1 | 1.109347508 | 0.018959438 |
| LAMC1 | 1.049836139 | 0.003629749 |
| DOCK10 | -1.543102889 | 8.24E-05 |
| ARHGEF4 | 0.852615486 | 0.02882984 |
| USP44 | 3.268897434 | 1.60E-06 |
| CKAP4 | -0.827045565 | 0.000550477 |
| PWP1 | -0.548096273 | 0.031167669 |
| CKAP2 | -2.286152261 | 4.56E-05 |
| BORA | -1.300667176 | 0.013329564 |
| SPRY2 | 1.151353546 | 0.000140229 |
| NUDT15 | -0.633296872 | 0.036849144 |
| RAPGEF5 | 2.828955361 | 0.002894967 |
| RAC1 | -0.550771938 | 0.020308343 |
| IL6 | 2.700824442 | 0.019069287 |
| ZDHHC4 | 0.571988021 | 0.044784813 |
| BZW2 | -1.127684759 | 0.002026214 |
| TBRG4 | -0.742926849 | 0.003471455 |
| NACAD | -0.744512658 | 0.014634856 |
| DBNL | -0.762341434 | 0.008041007 |
| SRSF1 | -0.672419454 | 0.00688326 |
| LIMD2 | -0.779927749 | 0.014552013 |
| BRIP1 | -3.071231622 | 1.18E-08 |
| GALNT5 | -2.353638865 | 1.41E-08 |
| HLX | 1.367201104 | 0.001050227 |
| SMPD4 | -0.518358237 | 0.048046307 |
| HS6ST1 | 0.590488164 | 0.012553061 |

| | | |
|---|---|---|
| GYPC | 0.879410716 | 0.018535076 |
| NIPSNAP3A | -0.897371203 | 0.011486577 |
| RALGPS1 | 0.922508692 | 0.008998337 |
| NIBAN2 | -0.960689138 | 0.003648279 |
| ANGPTL2 | 1.344716094 | 0.000618854 |
| TLR4 | 1.886244653 | 0.000124576 |
| STX17 | 0.856210498 | 0.00036151 |
| FPGS | -0.77826464 | 0.012038196 |
| GARNL3 | 2.345423917 | 1.34E-07 |
| XPA | 0.633751951 | 0.034842119 |
| CCN3 | -1.250931356 | 0.035219242 |
| UBAP2 | -0.588704779 | 0.024945188 |
| RNF38 | 0.600557418 | 0.04203439 |
| TLN1 | -1.014468057 | 0.000242388 |
| DCTN3 | -0.9199522 | 0.008312263 |
| ALDH1B1 | -1.323591793 | 0.010597036 |
| ARHGEF39 | -3.268633601 | 3.13E-06 |
| IGFBPL1 | -1.553047401 | 0.045081718 |
| CNPY3 | -0.647791935 | 0.030496456 |
| PPIL1 | -0.702085223 | 0.025638692 |
| PIM1 | 1.385357071 | 0.002094512 |
| SLC22A23 | 0.727025895 | 0.018449661 |
| TUBB2A | -1.567227045 | 3.67E-06 |
| HMGA1 | -1.441750092 | 0.000123808 |
| TCF19 | -2.667476747 | 1.45E-05 |
| TPMT | -0.657432828 | 0.014848945 |
| NRM | -1.229493705 | 0.006273333 |
| MTCH1 | -0.873048059 | 0.001556523 |
| FAM8A1 | 0.599757424 | 0.025134487 |
| FHDC1 | -1.70020033 | 0.025995761 |
| ANKRD42 | 0.592979863 | 0.035081562 |
| SYTL2 | 1.00677058 | 0.01395615 |
| RAB30 | 0.555942971 | 0.042742236 |
| PI15 | 5.901396999 | 0.008154622 |
| GGH | -2.855188355 | 2.38E-18 |
| SULF1 | -1.499740441 | 0.00693216 |
| SORL1 | -1.621272998 | 0.038458952 |
| TRPC6 | 1.581999686 | 0.040778319 |
| MMP27 | 3.123928951 | 0.000298714 |
| FDX1 | 0.644814044 | 0.037547508 |
| FXYD6 | -3.315259357 | 0.039807604 |
| ARHGAP20 | 1.626506454 | 0.008039674 |

| | | |
|---|---|---|
| CASP1 | 1.504546606 | 0.015105639 |
| MAP2K5 | 1.057946467 | 9.19E-06 |
| KIF23 | -3.067401159 | 8.25E-08 |
| KNL1 | -4.988299773 | 2.71E-11 |
| PARP6 | 0.588418673 | 0.049384149 |
| PAQR5 | -3.414761006 | 8.89E-07 |
| LRRC49 | 0.874748283 | 0.005075415 |
| SMAD6 | -1.097194247 | 0.007403234 |
| PLCB2 | -2.076193215 | 0.007458571 |
| DUOX1 | 2.947406244 | 0.011342619 |
| CYP19A1 | 1.777789194 | 0.018954781 |
| SEMA6D | 2.348991184 | 1.31E-08 |
| SPTBN5 | -2.390330601 | 0.00065959 |
| BCAR3 | -0.636108562 | 0.03413163 |
| IFI44L | 1.639977854 | 0.003848092 |
| SELENOI | -0.572903975 | 0.026475806 |
| DYNC2LI1 | 1.021609785 | 5.77E-05 |
| PREPL | 0.694134901 | 0.027543836 |
| EMILIN1 | -1.030725246 | 0.004310093 |
| CENPO | -1.833737006 | 0.000259318 |
| ACTR1A | -0.752426314 | 0.005763474 |
| MYOF | -0.950976316 | 0.001495654 |
| LOXL4 | 1.837779588 | 5.62E-05 |
| ATAD1 | 0.654385881 | 0.030380174 |
| KIF11 | -2.94919338 | 4.75E-06 |
| CEP55 | -5.275987495 | 3.69E-12 |
| PLCE1 | -1.072107059 | 0.014523578 |
| ADAMTS14 | -1.495924448 | 0.008392193 |
| MYPN | -4.636277461 | 0.049996401 |
| BARD1 | -1.3486699 | 0.014518434 |
| CARF | 1.329932771 | 0.000496385 |
| ASNSD1 | 0.465012824 | 0.048928398 |
| CDK15 | -2.689526269 | 5.10E-06 |
| MDH1B | 1.358366191 | 0.043560958 |
| SLC40A1 | 2.29597121 | 0.000448685 |
| SLC49A4 | 0.907984099 | 0.004251243 |
| PARP9 | 1.188747471 | 0.002118674 |
| GLCE | -0.60089536 | 0.041377011 |
| CILP | 5.031772611 | 1.15E-05 |
| SEMA7A | -2.384220252 | 5.09E-08 |
| FAM13A | 0.789994082 | 0.009497539 |
| HERC5 | 1.816585879 | 0.010490345 |

| | | |
|---|---|---|
| FGF5 | -1.2649914 | 0.002399062 |
| GPAT3 | -2.767078203 | 1.24E-06 |
| NAAA | -0.863309304 | 0.02815526 |
| CCNG2 | 0.791100342 | 0.029616687 |
| SHROOM3 | -2.034542762 | 8.81E-05 |
| CENPE | -3.030740725 | 1.37E-05 |
| LEF1 | 1.969478906 | 0.001750175 |
| EGF | -2.668104452 | 0.022337763 |
| SLC39A8 | 2.702252177 | 1.08E-09 |
| FBN2 | -1.817521292 | 0.001759819 |
| RGS3 | 0.671478756 | 0.035566979 |
| GABARAPL1 | 0.900278628 | 0.001381741 |
| PRICKLE1 | 1.037282052 | 0.048349531 |
| C1RL | 1.269592124 | 0.003051321 |
| CLSTN3 | 0.794595734 | 0.04666977 |
| LRIG3 | 1.516546089 | 1.69E-05 |
| INHBE | 2.038510453 | 0.009783419 |
| GLIPR1 | -1.322828082 | 0.002039605 |
| PHLDA1 | -0.992422118 | 0.003079703 |
| LGR5 | 2.542679278 | 5.20E-07 |
| PTPRQ | 1.517953956 | 0.013667838 |
| DUSP6 | 1.479435831 | 0.000338032 |
| GAS2L3 | -2.450939219 | 0.000242007 |
| SLC46A3 | 2.054727111 | 1.89E-13 |
| SLC7A1 | -0.678981764 | 0.014525854 |
| LNX2 | 1.129991427 | 0.010950211 |
| TARBP2 | -0.90666522 | 0.002800004 |
| N4BP2L1 | 2.229654868 | 9.84E-07 |
| BRCA2 | -1.274555911 | 0.042070321 |
| ITGB7 | -1.503989786 | 0.007230196 |
| GALNT6 | -2.944235488 | 7.87E-08 |
| ESYT1 | -0.660108246 | 0.018891309 |
| ANKRD52 | -0.723649503 | 0.011155187 |
| LPAR6 | 1.081412378 | 0.01023937 |
| MORN3 | 2.285516055 | 0.000152706 |
| DIAPH3 | -2.258178424 | 3.73E-06 |
| ABHD13 | 0.835047931 | 0.007348819 |
| GRTP1 | 1.567135207 | 0.003930696 |
| CUL4A | -0.473740368 | 0.039483311 |
| CDH24 | -1.163074912 | 0.002303507 |
| ARMH4 | -1.309153778 | 3.34E-06 |
| STON2 | 4.822800546 | 2.85E-08 |

| | | |
|---|---|---|
| PTGR2 | 0.912556027 | 0.005916787 |
| WARS1 | 0.784502592 | 0.037135831 |
| SLC12A6 | 0.711829856 | 0.024918923 |
| BAHD1 | -0.577102664 | 0.02178939 |
| DISP2 | -1.655819386 | 0.002434174 |
| COMMD4 | -0.777915672 | 0.017572615 |
| TSPAN3 | 1.349146169 | 6.72E-06 |
| TPM1 | -1.61111337 | 0.000161844 |
| ARRDC4 | 1.128774107 | 0.029054371 |
| CYP11A1 | 1.378702804 | 0.025251752 |
| ADAMTS17 | 1.847273162 | 0.026661267 |
| PCSK6 | 3.045785398 | 8.18E-05 |
| HAPLN3 | -1.747728941 | 0.009687408 |
| FANCI | -2.029941432 | 0.000833636 |
| TICRR | -3.235102995 | 7.05E-05 |
| FURIN | -0.593976397 | 0.015006192 |
| IQGAP1 | -0.63725267 | 0.018598512 |
| TGFB1I1 | -1.439111185 | 6.09E-06 |
| ARMC5 | -0.787709569 | 0.003356125 |
| DHX38 | -0.480768448 | 0.034435004 |
| KIFC3 | -1.069994148 | 7.77E-06 |
| ADAMTS18 | 4.08677021 | 0.0076932 |
| CDH11 | 1.453375911 | 0.001552202 |
| MEAK7 | -0.904856488 | 0.000300961 |
| TCF25 | -0.540295892 | 0.030527172 |
| VPS53 | -0.580832425 | 0.016942434 |
| SLC14A1 | -5.556040523 | 0.000714455 |
| ZMYND15 | 2.053208634 | 0.009272289 |
| CCDC40 | 1.211500913 | 0.004375002 |
| ARHGDIA | -1.047239246 | 0.000514211 |
| TTYH2 | 1.410803141 | 0.003630603 |
| ANAPC11 | -0.690816875 | 0.043490961 |
| TBCD | -0.803902151 | 0.008982275 |
| SECTM1 | 2.447377046 | 0.03221539 |
| ARK2C | 5.085506289 | 0.037068387 |
| FBXO15 | 2.981456977 | 0.001732712 |
| STAC2 | -3.140237326 | 0.026674465 |
| TPGS1 | -1.247075446 | 0.001744505 |
| PLPP2 | -2.434251176 | 4.27E-05 |
| MVB12A | -0.778970774 | 0.040580995 |
| SH3GL1 | -0.792483925 | 0.005927363 |
| DPP9 | -0.731837984 | 0.003596598 |

| | | |
|---|---|---|
| IFNAR1 | 0.48455543 | 0.049332892 |
| APP | 0.66320081 | 0.032775798 |
| EMP3 | -1.399907292 | 0.001436085 |
| SAE1 | -1.070492802 | 0.000365037 |
| NTN5 | 1.696132429 | 0.048850301 |
| GEMIN7 | -0.738233943 | 0.034515101 |
| WTIP | -0.521661229 | 0.038421626 |
| ADAMTS10 | 0.799488164 | 0.024064903 |
| MYO1F | -1.64525056 | 0.001809708 |
| PSMB6 | -0.799925926 | 0.025224728 |
| CTU1 | -0.698380366 | 0.015001782 |
| RCN3 | -1.056023881 | 0.000285941 |
| SLC2A5 | 3.419937276 | 0.000201078 |
| EFHD2 | -1.274542029 | 2.92E-05 |
| MYOM3 | -2.212876784 | 0.014509592 |
| SH3BGRL3 | -1.616803399 | 1.66E-05 |
| KIAA0319L | 0.507448934 | 0.044100501 |
| PLK4 | -3.121347895 | 1.26E-05 |
| NBPF3 | 0.927784639 | 0.015403172 |
| PRKACB | 0.511538083 | 0.040419883 |
| KIF2C | -3.128030528 | 2.11E-05 |
| LMO4 | 0.72058671 | 0.01949019 |
| SLC44A3 | 1.076182346 | 0.025896629 |
| IGSF3 | 1.334047192 | 0.033045079 |
| PSMA5 | -0.639229391 | 0.038971215 |
| ITGA10 | -1.589075051 | 0.001381832 |
| CREG1 | 1.399988039 | 2.73E-05 |
| TBX19 | 1.215397976 | 0.000441727 |
| DPT | 2.63827147 | 5.77E-08 |
| NUF2 | -4.378848459 | 4.47E-10 |
| SDHC | -0.769599411 | 0.022121189 |
| MRPL24 | -0.602164815 | 0.047781594 |
| CRABP2 | -1.333393948 | 0.013664606 |
| HDGF | -0.631959925 | 0.048071468 |
| ABL2 | -0.708109142 | 0.013158019 |
| RGS16 | 2.359290176 | 0.000157086 |
| PRUNE1 | 0.569893518 | 0.020194725 |
| ANP32E | -1.130767174 | 0.005547103 |
| MINDY1 | 0.714451649 | 0.023537301 |
| SYT14 | -3.082614076 | 2.21E-09 |
| DTL | -3.799330425 | 5.47E-07 |
| SUSD4 | -4.589592212 | 0.007970157 |

| | | |
|---|---|---|
| DUSP10 | -0.6862225 | 0.029340919 |
| HHIPL2 | -3.295146023 | 1.44E-06 |
| ATP8B2 | -0.613851799 | 0.038109656 |
| TPM3 | -1.22832785 | 1.65E-06 |
| EFNA3 | 1.862946962 | 0.0034371 |
| ILF2 | -0.5804093 | 0.048321855 |
| ACTA1 | -2.386593745 | 0.00430544 |
| TTC13 | 0.767053035 | 0.02727717 |
| CNIH4 | 0.677031253 | 0.005351872 |
| ITPKB | 0.995737841 | 0.000560753 |
| GUK1 | -0.947079181 | 0.006495847 |
| CNIH3 | -1.892020552 | 0.000254838 |
| C1orf35 | -0.620633424 | 0.032479807 |
| WNT9A | -1.412159684 | 0.021564174 |
| EPHX1 | 1.226975651 | 0.004730356 |
| PDIA6 | -0.739853111 | 0.007869943 |
| RHOB | -1.261120263 | 0.008905089 |
| CHAC2 | -1.568592461 | 0.007691116 |
| MALL | -5.72600629 | 2.08E-14 |
| RABL2A | 1.336956013 | 4.05E-05 |
| ZC3H8 | 0.730437796 | 0.010695961 |
| LIPT1 | 0.837073188 | 0.019241567 |
| AFF3 | 0.857856428 | 0.028968419 |
| SPOPL | 0.828364537 | 0.035222921 |
| LNPK | 0.798740681 | 0.005320836 |
| TMEFF2 | -2.444112185 | 0.028702686 |
| CDCA7 | -3.111944684 | 1.16E-05 |
| GULP1 | 1.022413954 | 0.041534663 |
| CCDC150 | -1.587935749 | 0.035854349 |
| METTL21A | 1.200085509 | 0.000158914 |
| KANSL1L | 0.74569465 | 0.016535347 |
| ACKR3 | 1.679848217 | 0.001151726 |
| FANCD2 | -2.753713294 | 8.01E-06 |
| MARCHF4 | -1.711454458 | 1.04E-05 |
| GMPPA | -0.642382358 | 0.033164977 |
| CNTN4 | 3.457236156 | 0.016547392 |
| POMGNT2 | 0.643897025 | 0.03525918 |
| ACKR2 | 2.57553882 | 0.019520337 |
| ITGA9 | -1.065366795 | 0.014295033 |
| PTPRG | 0.729431149 | 0.016833141 |
| IL17RD | 1.329051634 | 0.000965529 |
| LRIG1 | 0.725096867 | 0.031248149 |

| | | |
|---|---|---|
| NFKBIZ | 1.560984043 | 0.006507055 |
| COL8A1 | 1.004230725 | 0.037107541 |
| AGTR1 | 1.571461816 | 0.001629281 |
| MED12L | 2.66964997 | 2.95E-05 |
| NCEH1 | -1.164553387 | 0.005012469 |
| RUBCN | -0.778500099 | 0.005588975 |
| TCTA | -1.343845092 | 0.000961007 |
| MANF | -0.7770011 | 0.027772786 |
| SLIT2 | 1.615811616 | 0.002952192 |
| OCIAD2 | -1.471037079 | 0.008820372 |
| KLHL8 | 0.63922963 | 0.025038074 |
| SNCA | 4.394140422 | 0.046517729 |
| DDIT4L | -1.480753976 | 0.00847638 |
| ANK2 | 0.911780648 | 0.007688468 |
| CCNA2 | -3.871700779 | 1.75E-09 |
| MARCHF1 | 3.346913786 | 0.002939948 |
| RNF175 | 2.33285965 | 0.002795079 |
| CYP4V2 | 1.410482306 | 9.75E-05 |
| CDH18 | -1.585487561 | 0.011637906 |
| MYO10 | -1.465354792 | 1.96E-05 |
| OTULINL | -2.444703501 | 0.008659329 |
| SKP2 | 0.886266908 | 0.003866103 |
| OSMR | 1.461477057 | 5.48E-05 |
| SSBP2 | 0.685302173 | 0.007028338 |
| ANKRD31 | 2.793701665 | 0.003283528 |
| GIN1 | 0.55608958 | 0.046399593 |
| TSLP | 3.87207907 | 0.001568736 |
| TNFAIP8 | 1.412554968 | 0.000149665 |
| ARHGAP26 | 1.35200297 | 0.019840448 |
| C1QTNF2 | -2.926945734 | 0.000884737 |
| NHP2 | -0.875071395 | 0.009323149 |
| MYLK4 | 1.514870846 | 0.000206532 |
| GFOD1 | -1.551486718 | 0.007348561 |
| KLHL3 | 1.275574321 | 0.008754256 |
| TRIM7 | -3.020133912 | 1.02E-07 |
| RNF44 | 0.718795053 | 0.021421754 |
| PPP1R18 | -0.677071982 | 0.017390368 |
| PRIM2 | -0.896513787 | 0.020608005 |
| ANO7 | 1.207097329 | 0.014084767 |
| TCTE1 | 2.822450621 | 0.015010481 |
| CYP39A1 | 3.637423217 | 0.027346656 |
| PNRC1 | 1.032122777 | 0.000498663 |

| | | |
|---|---|---|
| RARS2 | 0.491459633 | 0.040423104 |
| RNF217 | 0.788146835 | 0.044891853 |
| ARHGAP18 | -1.139440175 | 6.92E-05 |
| ABRACL | -1.336296557 | 0.000172163 |
| MTFR2 | -3.498032634 | 9.88E-05 |
| SLC2A12 | 1.581530252 | 0.001975321 |
| SHPRH | 0.797584849 | 0.033729209 |
| ARMT1 | 0.488070335 | 0.045601472 |
| CREB5 | 1.825325875 | 8.91E-05 |
| CDCA5 | -3.81146219 | 3.45E-08 |
| MDH2 | -0.625442683 | 0.033011622 |
| CCT6A | -0.729297866 | 0.005128418 |
| TRAPPC14 | -0.626606381 | 0.015205742 |
| SLC12A9 | -0.620858268 | 0.038782578 |
| TMEM209 | 0.883504362 | 0.015595185 |
| TMEM140 | 1.809801081 | 0.001945503 |
| NCAPG2 | -1.684189253 | 0.006049663 |
| NLGN4X | 4.096690354 | 8.26E-06 |
| DENND2A | 2.018350938 | 0.001872038 |
| MSN | -1.267793725 | 1.31E-06 |
| SLC16A2 | -1.169555907 | 0.001306685 |
| CHST7 | -1.113400298 | 0.040530768 |
| LPAR4 | 2.166225686 | 0.000836704 |
| EBP | -1.349586904 | 0.001517932 |
| OGT | 0.822418997 | 0.036252549 |
| PRPS1 | -2.566771727 | 7.47E-14 |
| FRMPD3 | -4.216534203 | 1.82E-10 |
| NSDHL | -0.759490054 | 0.021773107 |
| CSGALNACT1 | 2.044429858 | 0.00052399 |
| SLC25A37 | 0.901499826 | 0.048403239 |
| DOCK5 | -1.515697118 | 2.61E-07 |
| GINS4 | -2.745955001 | 1.79E-06 |
| ADHFE1 | 1.677632244 | 0.008257609 |
| LACTB2 | 0.604558052 | 0.038369727 |
| SYBU | 1.137465895 | 0.000583536 |
| DPYS | 7.282370145 | 1.16E-06 |
| EBAG9 | 1.085574104 | 0.000136618 |
| MAL2 | 4.926677517 | 0.010826617 |
| UTP23 | 0.633107947 | 0.047681408 |
| VLDLR | 1.396660927 | 0.031920255 |
| AK3 | 0.84100358 | 0.001547868 |
| ZCCHC7 | 0.554998266 | 0.021721914 |

| | | |
|---|---|---|
| NTRK2 | 3.318867219 | 3.27E-10 |
| IDNK | 1.027176248 | 0.017253319 |
| AUH | 0.655592344 | 0.017462962 |
| UGCG | 0.848348764 | 0.035332047 |
| WDR31 | 0.918832948 | 0.001166492 |
| SURF4 | -0.51521295 | 0.045071354 |
| ASB6 | -0.803698883 | 0.000891402 |
| NTMT1 | -0.988676568 | 0.001216475 |
| MIGA2 | -0.809240186 | 0.000946136 |
| PTGES | -1.15677113 | 0.000202022 |
| NACC2 | -0.70365547 | 0.011964518 |
| RSU1 | -1.046421182 | 0.0004959 |
| ST8SIA6 | -4.698467306 | 0.002488684 |
| ZEB1 | 0.98272605 | 0.001597479 |
| FAM13C | 1.31671537 | 0.001502248 |
| NRBF2 | 0.623637565 | 0.046859008 |
| POLR3A | -0.552521796 | 0.034477228 |
| ADIRF | -3.371807871 | 0.018161497 |
| FRA10AC1 | 0.566055562 | 0.045146502 |
| TCF7L2 | 1.212630425 | 7.99E-05 |
| MKI67 | -5.450885 | 1.66E-05 |
| LRRC27 | 0.532163259 | 0.046911189 |
| GSTO1 | -0.965112404 | 0.020157744 |
| PPRC1 | -0.825324048 | 0.000572818 |
| ADAM12 | -1.103191082 | 0.017166105 |
| SLC5A12 | 2.529875638 | 0.021053153 |
| ZNF214 | 0.965672725 | 0.011055475 |
| DGKZ | -0.513644516 | 0.044935496 |
| SERPING1 | 1.443368357 | 0.001541399 |
| PTPRJ | -0.80409986 | 0.016081754 |
| SESN3 | 2.032189869 | 0.000147788 |
| CAPN5 | -0.750138619 | 0.033679018 |
| AASDHPPT | 0.538550772 | 0.047229453 |
| P4HA3 | -1.497852651 | 0.00107315 |
| KAT14 | 0.577551684 | 0.047036949 |
| INCENP | -1.595207793 | 0.004889035 |
| B3GAT3 | -0.79733213 | 0.016721905 |
| CHEK1 | -1.62116515 | 1.40E-06 |
| KIRREL3 | -2.016616071 | 2.01E-05 |
| TAGLN | -2.024314641 | 0.000232425 |
| JPH2 | -1.336266939 | 0.042055743 |
| C20orf144 | -4.681930971 | 0.002937571 |

| | | |
|---|---|---|
| KIAA1755 | -1.775767398 | 0.00230341 |
| CABLES2 | 0.626789581 | 0.044514843 |
| PLCB3 | -1.204936726 | 1.18E-05 |
| TM7SF2 | 0.878357817 | 0.03278924 |
| PPP4C | -0.786124438 | 0.005548362 |
| HMGA2 | -1.168902081 | 0.027773822 |
| MMP3 | -5.610590294 | 1.40E-15 |
| MKX | 1.939500954 | 8.76E-07 |
| MPP7 | 1.51163044 | 0.010144425 |
| DCUN1D2 | 0.658073531 | 0.026447577 |
| HNMT | 2.036007288 | 1.44E-13 |
| LYPD1 | -3.619074608 | 3.98E-05 |
| ADRA2A | 1.402358426 | 0.00014939 |
| CCDC102B | 1.404138061 | 0.003275789 |
| PRSS23 | 1.615360165 | 0.006456649 |
| CCT5 | -0.726932568 | 0.008231271 |
| PIP4K2A | -0.590688451 | 0.026082122 |
| CRIM1 | -0.860471428 | 0.005597396 |
| DHX37 | -0.595964213 | 0.031805173 |
| GPR158 | 2.0178608 | 0.007552996 |
| CACNA2D4 | 1.306620057 | 0.018588536 |
| THRB | 0.937417058 | 0.02892559 |
| ABTB3 | -1.963706687 | 0.00155002 |
| IPMK | 0.990394627 | 0.024601151 |
| PLBD2 | 0.817969268 | 0.006885012 |
| NPAS3 | 1.573484355 | 0.000122338 |
| MBIP | 0.504885345 | 0.037409238 |
| ME3 | 0.883725045 | 0.046240891 |
| ADAMTS12 | 1.059850392 | 0.02936345 |
| FRMD4A | -0.807958606 | 0.014077389 |
| MMAA | 0.58730265 | 0.023694066 |
| AKR1C2 | 1.629056399 | 0.002223185 |
| ADAM17 | 0.904388936 | 0.01282335 |
| CENPU | -3.406330604 | 3.77E-07 |
| ACSL1 | 0.61283914 | 0.041221292 |
| CCDC122 | 1.397584757 | 0.004468978 |
| SACS | -1.498417263 | 3.22E-07 |
| PARP8 | 0.873018768 | 0.004754435 |
| DST | -0.695621979 | 0.025441702 |
| BAG3 | -0.743807388 | 0.001696944 |
| MZT2B | -0.884537659 | 0.005407698 |
| ASTN1 | 1.752453836 | 0.010242899 |

| | | |
|---|---|---|
| TMEM178A | 2.325014485 | 0.002059901 |
| SETBP1 | 1.855547168 | 2.42E-05 |
| EPG5 | -0.864837333 | 0.000217917 |
| ATP5F1A | -0.746930103 | 0.014416946 |
| SPC25 | -5.413150017 | 0.000107411 |
| SPOCK1 | 0.888076673 | 0.020843681 |
| JMY | 0.817621929 | 0.028282334 |
| NMT2 | -0.705811045 | 0.010339051 |
| CCDC50 | 0.726416235 | 0.027116954 |
| PLEKHH2 | 1.389322251 | 0.004980844 |
| IGSF10 | 3.956351153 | 3.25E-08 |
| SPARCL1 | 4.850300649 | 0.023502084 |
| GJA1 | 1.911975157 | 9.30E-09 |
| DYNLT5 | -2.683374793 | 7.00E-05 |
| DNAI4 | 0.782604373 | 0.032697448 |
| ANKRD22 | 4.899427951 | 0.007420598 |
| PANK1 | -0.92371882 | 0.009848553 |
| GRM1 | 1.874240023 | 0.036676417 |
| PTPRK | 0.847390371 | 0.018970323 |
| MARVELD2 | 2.628880912 | 3.82E-05 |
| PLOD2 | 1.117817212 | 0.035297703 |
| STK32B | -0.853129727 | 0.021737147 |
| CPB1 | 5.230217145 | 1.14E-17 |
| MR1 | 0.574509169 | 0.046728657 |
| CENPH | -2.473584866 | 0.000101315 |
| BCL2L11 | 2.178870088 | 4.12E-07 |
| CLGN | 2.300599589 | 4.50E-05 |
| SYCP2L | 5.102103719 | 0.00754698 |
| CCDC148 | 1.558895431 | 0.007061434 |
| RBMS1 | 0.661003565 | 0.030441461 |
| SLC25A27 | 1.992964407 | 2.45E-05 |
| ASAP1 | -0.516543826 | 0.024672299 |
| TEX29 | 4.241686634 | 8.22E-05 |
| LURAP1L | 0.69968265 | 0.039089189 |
| SPMIP4 | 0.776432934 | 0.039479766 |
| PID1 | 0.790063476 | 0.021910161 |
| CEBPG | 0.796623321 | 0.042523388 |
| HS2ST1 | 0.749289038 | 0.003455105 |
| GDPD1 | 2.1396446 | 4.52E-05 |
| AK5 | -1.935579628 | 1.84E-05 |
| TBCEL | 0.562027877 | 0.044979041 |
| UBASH3B | -2.755335249 | 2.80E-07 |

| | | |
|---|---|---|
| TBRG1 | 0.765193536 | 0.003335228 |
| NRGN | -4.36120619 | 2.63E-06 |
| CC2D1B | -0.656333103 | 0.007415961 |
| PRKCA | -0.983682421 | 0.008710303 |
| ABCA9 | 3.683468539 | 0.001019579 |
| ABCA6 | 2.552863376 | 6.92E-10 |
| ABCA10 | 3.288486759 | 8.66E-08 |
| DISP1 | 0.730657653 | 0.025093663 |
| TNIK | -1.342268136 | 0.007761519 |
| ENAH | -0.974523861 | 0.000102045 |
| GBP5 | -2.34019645 | 0.006018044 |
| L3MBTL4 | 2.548083789 | 0.020414696 |
| JAM2 | 2.447768999 | 1.13E-05 |
| ADAMTS5 | 2.092529302 | 2.08E-06 |
| XPC | 0.751483407 | 0.00928538 |
| SKA1 | -4.741915996 | 7.32E-10 |
| APCDD1 | 2.722695348 | 5.24E-05 |
| PIEZO2 | 1.587772522 | 0.029891599 |
| EME1 | -2.484544087 | 0.000570104 |
| EPHB1 | -1.602296817 | 3.55E-05 |
| VOPP1 | -0.774825054 | 0.001332091 |
| DKK2 | 2.507109144 | 8.95E-06 |
| CYP2U1 | 1.118862963 | 0.000902626 |
| FBXL18 | -0.550211279 | 0.046859778 |
| AK9 | 0.931093357 | 0.015581965 |
| MMS19 | -0.508293512 | 0.039005111 |
| GPR78 | 4.197744726 | 5.70E-07 |
| USP25 | 0.600907281 | 0.023424089 |
| RHOC | -1.184177674 | 0.000429848 |
| DBI | -0.894967238 | 0.035877223 |
| SLC7A7 | 1.08350869 | 0.01907392 |
| GRIA1 | 6.492193181 | 2.09E-16 |
| ZKSCAN2 | 0.565868214 | 0.048935662 |
| OTOA | -3.193857375 | 0.017089313 |
| KCTD18 | 0.5723438 | 0.023643763 |
| C2CD6 | -1.195799188 | 0.009290865 |
| TMEM237 | -0.672982341 | 0.012706911 |
| DEPTOR | 1.468282058 | 0.010295867 |
| FMN2 | -2.112846686 | 9.93E-11 |
| PPARGC1B | -3.095566609 | 0.020421257 |
| SLC24A2 | 3.261997292 | 0.040868635 |
| RMND1 | 0.518785527 | 0.049730381 |

| | | |
|---|---|---|
| SLA | 2.929042964 | 0.010324768 |
| CLIC2 | 1.832317479 | 0.001209891 |
| KIF5A | -1.528431962 | 0.007662505 |
| MIDEAS | 0.600593533 | 0.025716723 |
| MMP16 | -1.293540342 | 0.037393391 |
| ADAMTS3 | 1.905596931 | 0.004908313 |
| ALX3 | -2.882174323 | 0.021130321 |
| DRAM2 | 0.517671701 | 0.030127431 |
| BACH1 | 0.94270986 | 0.003408844 |
| ANKRD9 | -0.876172973 | 0.008432325 |
| SFXN2 | -1.427021103 | 0.000181607 |
| PCDH1 | 3.084089502 | 3.43E-07 |
| GDF6 | 1.621926352 | 0.025338883 |
| PPP2R2B | -2.416271854 | 0.008300242 |
| FBXO43 | -2.618447759 | 0.009535506 |
| HK1 | -0.540381126 | 0.026387779 |
| CD109 | -1.060297682 | 5.07E-05 |
| RAB11FIP1 | 1.258969399 | 0.002478988 |
| ATAD2 | -1.30647277 | 0.023067896 |
| BUB1B | -4.852389022 | 1.42E-09 |
| SST | -4.62579195 | 0.001428991 |
| SEC13 | -0.718476617 | 0.021810971 |
| ODR4 | 0.686854668 | 0.014902587 |
| NECAP2 | -0.808734163 | 0.00621431 |
| LRP8 | -1.662190072 | 0.000442252 |
| STEAP2 | 1.067277 | 0.001115154 |
| FZD1 | 0.84169365 | 0.022599125 |
| ZFHX2-AS1 | 2.508630075 | 0.000554944 |
| CLEC18A | 4.630463837 | 0.027663628 |
| CFAP107 | 2.302423231 | 0.041148028 |
| ARMC12 | 3.585806482 | 0.005949813 |
| KIT | 1.763731003 | 2.18E-05 |
| AASDH | 0.574537903 | 0.039351387 |
| CACNA2D3 | 2.215814797 | 0.00652111 |
| CCNB2 | -5.180161842 | 8.57E-16 |
| ETS2 | -1.85027401 | 2.15E-07 |
| TMEM164 | -0.742117715 | 0.009928113 |
| MX1 | 1.320654621 | 0.004655429 |
| DGKI | -1.822822251 | 0.003646731 |
| ACAN | -3.105325634 | 0.014774948 |
| WDR19 | 0.659067998 | 0.013393825 |
| RAB28 | 0.671217382 | 0.013521272 |

| | | |
|---|---|---|
| PRXL2B | -1.026372503 | 0.004897702 |
| EXTL1 | -1.612851471 | 0.020322857 |
| DUSP2 | -2.230496639 | 0.034341184 |
| HPD | -2.504589884 | 0.017486427 |
| TMSB15A | -2.474512676 | 0.002488859 |
| MRAS | -0.702887168 | 0.009148236 |
| TENT5B | -2.060558466 | 0.0046519 |
| CLSTN2 | 3.779985308 | 9.22E-12 |
| GPR153 | 1.690442805 | 5.68E-08 |
| RHBDL2 | -2.092249349 | 0.005117157 |
| H2BC5 | 0.996124429 | 0.00992168 |
| CDC25C | -5.096517779 | 9.62E-09 |
| H4C8 | 2.108614033 | 8.86E-06 |
| AHCYL2 | 0.589216499 | 0.018668454 |
| CPA5 | 5.090784485 | 0.015661761 |
| PPP1R9A | 2.796436507 | 6.94E-07 |
| DYNC1I1 | -1.90113724 | 0.003337014 |
| TAGLN2 | -1.262641421 | 0.001724109 |
| NBL1 | -1.346959056 | 0.019909349 |
| EDA | 1.480924261 | 0.028912869 |
| FGF17 | 5.703545468 | 9.33E-05 |
| CDA | -2.992127376 | 8.58E-10 |
| PINK1 | -0.699689017 | 0.013450002 |
| MPZ | 2.18782105 | 0.001194376 |
| WNT9B | 4.663661073 | 0.004464514 |
| CACHD1 | 1.009537491 | 0.02386468 |
| RAPGEF6 | 0.799708183 | 0.026134958 |
| IFNAR2 | 0.846465495 | 0.020179333 |
| SV2A | -0.874091873 | 0.035998453 |
| STC1 | 1.553653289 | 0.0002463 |
| CSRP1 | -1.468423703 | 0.000878574 |
| HOXB13 | -4.714608467 | 0.006185553 |
| ATP5MC1 | -0.794076415 | 0.014307682 |
| CIART | 0.741007855 | 0.012340794 |
| SNF8 | -0.709500552 | 0.015377372 |
| IGF2BP1 | -1.585666346 | 0.009387195 |
| GIP | -6.617602988 | 1.66E-05 |
| CBR3 | -1.31748745 | 0.005700168 |
| GJD2 | -3.793599659 | 4.73E-08 |
| ACTC1 | -5.700158496 | 9.80E-23 |
| CHAF1B | -1.082093539 | 0.032816955 |
| HLCS | 0.811459438 | 0.01841323 |

| | | |
|---|---|---|
| PTMS | -1.226308995 | 0.000116484 |
| C1R | 1.942894684 | 1.70E-06 |
| ALDH4A1 | -1.564611704 | 0.000175254 |
| SPON2 | -3.744266549 | 0.0012675 |
| ZDHHC1 | 0.771212784 | 0.022694786 |
| CARMIL2 | -4.543045807 | 3.38E-13 |
| ZYX | -1.195981719 | 0.000252679 |
| LYPD5 | 1.668366189 | 0.004920214 |
| ZNF221 | 1.232458268 | 0.039304395 |
| PTGIR | -1.078013705 | 0.006260107 |
| CALM3 | -0.861546556 | 0.003630833 |
| ATAD3B | -0.892434612 | 0.02277691 |
| SLC37A1 | 1.210521375 | 0.038881474 |
| GATD3 | 1.775007179 | 0.00714934 |
| SLX9 | -0.604855636 | 0.025420201 |
| LSS | -0.849258065 | 0.018723225 |
| SLC2A6 | -1.489383628 | 7.23E-05 |
| SHKBP1 | -1.106324954 | 0.003197904 |
| RDH13 | 0.902299284 | 0.008664325 |
| ZDHHC12 | -1.400985181 | 0.000350895 |
| PKN3 | -1.524803771 | 0.00481695 |
| PLPP7 | 1.095610909 | 0.01861234 |
| TLCD1 | -1.064377778 | 0.003535443 |
| PCSK7 | -0.985029657 | 0.000130501 |
| ANO10 | -0.857084052 | 0.009123656 |
| FDPS | -0.801204508 | 0.011628921 |
| RUSC1 | -0.767129344 | 0.009405412 |
| PAQR6 | 1.358424065 | 0.011350788 |
| LMNA | -1.513426564 | 0.000100645 |
| PTH1R | 1.529667399 | 1.49E-05 |
| MYL3 | 1.078769218 | 0.04117599 |
| PPP1R35 | -0.854865764 | 0.011643693 |
| GPATCH4 | -0.795085657 | 0.019825563 |
| LRRC71 | -2.943166144 | 0.002460628 |
| NACC1 | -0.69748298 | 0.008150567 |
| LY6K | -2.65923112 | 3.91E-09 |
| CPSF4 | 0.674668135 | 0.014385635 |
| TONSL | -1.636382512 | 0.004479549 |
| PTGER1 | -4.134750224 | 0.000281619 |
| RECQL4 | -2.285954241 | 0.000300002 |
| COL26A1 | 2.879257171 | 0.007275952 |
| ORAI2 | -0.597579136 | 0.018596462 |

| | | |
|---|---|---|
| SH2B2 | 1.324630865 | 0.001967844 |
| MFSD12 | -0.812318821 | 0.009935702 |
| YDJC | -0.942679121 | 0.00378522 |
| AP2M1 | -0.754905478 | 0.002138569 |
| BDH1 | -1.764736241 | 0.000357354 |
| LRRC56 | 0.899093726 | 0.049058685 |
| PLXDC1 | 1.478304569 | 0.003628945 |
| SRSF2 | -0.61438435 | 0.017900514 |
| ZNF577 | 0.910122898 | 0.015110587 |
| MPP3 | -1.135163675 | 0.004768215 |
| NAGS | -1.731859559 | 0.000550315 |
| JOSD2 | -0.778234948 | 0.036706471 |
| DBF4B | -1.859900272 | 0.001456906 |
| FMNL3 | -0.707329617 | 0.032892092 |
| RACGAP1 | -2.771879952 | 6.36E-06 |
| RAVER1 | -0.670344223 | 0.005886129 |
| SPC24 | -3.95092049 | 1.95E-10 |
| CXCL16 | 2.901581555 | 3.31E-12 |
| BCL6B | -2.488201087 | 0.020481633 |
| TEDC2 | -1.554973999 | 0.007324338 |
| CCNF | -2.982581275 | 5.50E-06 |
| PAQR4 | -2.217417962 | 7.23E-06 |
| SAXO4 | 1.407959853 | 0.016208098 |
| ASRGL1 | -1.913275781 | 0.027531682 |
| RPS6KA4 | -1.074404308 | 0.000862638 |
| LRP5 | 0.782276496 | 0.025479395 |
| PDZK1IP1 | 4.511997409 | 0.015815087 |
| BEND5 | 2.112199459 | 6.62E-05 |
| SLC1A7 | 2.129872618 | 0.003459627 |
| ACOT11 | -2.441207925 | 1.35E-05 |
| PRKAA2 | 1.555082995 | 0.003619813 |
| AK4 | 1.492875068 | 0.004238205 |
| FBLIM1 | -0.650020633 | 0.032822431 |
| PDPN | 1.760514493 | 0.000593776 |
| SYNC | -1.08840056 | 0.005626507 |
| TMCO4 | -0.966938802 | 0.026246491 |
| ALPL | 3.196421017 | 4.83E-07 |
| WNT4 | 2.557030957 | 5.03E-10 |
| SCNN1D | 1.274168661 | 0.002532287 |
| MXRA8 | -0.873556353 | 0.018867012 |
| ADGRL4 | -1.172947392 | 0.010283775 |
| NTNG1 | -2.998710926 | 1.80E-08 |

| | | |
|---|---|---|
| C1orf52 | 0.773395897 | 0.016197257 |
| GBP2 | 1.06002849 | 0.004512098 |
| GBP4 | -1.319736336 | 0.014509456 |
| VCAM1 | 4.486604859 | 4.44E-12 |
| SLC30A7 | 0.594296152 | 0.0456734 |
| ARPC5 | -0.784398086 | 0.002675289 |
| CADM3 | 5.614909132 | 3.45E-05 |
| SLAMF9 | -3.426902236 | 0.023351108 |
| VANGL2 | 2.994087081 | 0.043992345 |
| FCRLB | -3.001303883 | 3.84E-06 |
| ATF3 | 1.414386834 | 0.005079771 |
| SNED1 | 1.265398901 | 0.014264952 |
| C1orf115 | 1.569224359 | 0.046283489 |
| HAAO | -2.458248484 | 0.008051314 |
| B3GALNT2 | 0.724422912 | 0.010666457 |
| CAPN2 | -0.876505648 | 0.000878774 |
| SANBR | -0.816314231 | 0.016833869 |
| RFTN2 | 1.424004226 | 1.10E-06 |
| DISC1 | 0.943476259 | 0.045010078 |
| LRATD1 | 1.251406025 | 0.000108951 |
| TEKT4 | 2.696264024 | 0.022042876 |
| SPATA18 | 0.713559156 | 0.035029379 |
| NOSTRIN | -1.99406267 | 0.017629642 |
| CFAP221 | 2.446437512 | 2.74E-05 |
| INHBB | 4.975087874 | 0.000235792 |
| PDLIM5 | -1.148588553 | 0.03426965 |
| RPRD2 | 0.477004698 | 0.049984287 |
| MSX1 | -1.028570418 | 0.007814701 |
| PACRGL | 0.534165901 | 0.048965688 |
| BOLA3 | -0.737990935 | 0.045761329 |
| ARHGAP25 | 3.853316822 | 0.021013621 |
| TGFA | -2.217377524 | 0.04353426 |
| FZD5 | 0.769841979 | 0.016722095 |
| ANTXR2 | 0.998305553 | 0.001893693 |
| GPR155 | 1.296643142 | 0.000445839 |
| CLDN1 | 2.635606568 | 0.046192033 |
| LINC01116 | -1.014115779 | 0.010389484 |
| KBTBD8 | -1.476920741 | 0.021358344 |
| EIF4E3 | 0.782882064 | 0.02081106 |
| LMOD1 | -1.409604354 | 0.010501855 |
| TRIM46 | -1.315657972 | 4.25E-06 |
| ARPC2 | -0.811031068 | 0.002245091 |

| | | |
|---|---|---|
| CCT3 | -0.794798455 | 0.004196077 |
| TMEM79 | 0.786602731 | 0.016093468 |
| RNF25 | -0.497608324 | 0.048808401 |
| ADORA1 | -1.281819786 | 0.003776242 |
| CIP2A | -2.239327399 | 0.000342791 |
| AZI2 | 0.678510227 | 0.027978608 |
| TGFBR2 | 0.591626326 | 0.02149086 |
| FBLN2 | 0.905636184 | 0.00090988 |
| NFASC | 0.584582835 | 0.023148246 |
| SGO2 | -2.275963095 | 0.000659795 |
| SPTA1 | 2.389979587 | 0.041051498 |
| RPL22L1 | -2.085501648 | 7.03E-09 |
| PPM1L | 2.569827818 | 4.84E-06 |
| RYBP | 0.68270425 | 0.02100538 |
| ADAMTS9 | 2.525079878 | 5.30E-07 |
| TIPARP | -0.821489075 | 0.005437991 |
| CCNL1 | 0.811562872 | 0.020178025 |
| PTX3 | 2.201772943 | 8.54E-08 |
| HESX1 | 1.48447284 | 0.036719364 |
| SMIM14 | 0.634760121 | 0.016548566 |
| DNASE1L3 | -7.627205938 | 2.70E-09 |
| RBM47 | -1.472900192 | 0.021430258 |
| IL17RE | -3.011892196 | 0.000198212 |
| PCOLCE2 | -1.542691339 | 0.02701304 |
| CCDC158 | 1.730816182 | 0.017676389 |
| CPA3 | 3.021941808 | 0.002851577 |
| UCN | 1.75064348 | 0.007099693 |
| PLB1 | -2.981455053 | 0.000210192 |
| KIF15 | -2.75223788 | 0.000159578 |
| LZTFL1 | 0.590884881 | 0.019326008 |
| DTX3L | 0.869225237 | 0.018767379 |
| NMNAT3 | 3.588107007 | 7.47E-05 |
| SMIM12 | -0.644244861 | 0.027716235 |
| PRKCD | -0.712894727 | 0.024935558 |
| ARHGEF3 | 1.089768116 | 0.03498981 |
| METAP1 | -0.581314287 | 0.020411834 |
| BDH2 | 0.729038326 | 0.020459219 |
| CDC25A | -2.38167584 | 2.83E-05 |
| ZNF589 | 1.292189623 | 0.000122245 |
| PLXNB1 | 0.762243985 | 0.048394873 |
| ATRIP | -0.679716874 | 0.034908008 |
| SPRY1 | 2.955452591 | 3.19E-23 |

| | | |
|---|---|---|
| HSPA4L | 2.100893505 | 3.43E-10 |
| MST1R | 2.185434359 | 0.005666418 |
| DUSP7 | -0.715652849 | 0.01171887 |
| POC1A | -2.369887625 | 0.000209927 |
| HMGB2 | -1.01460513 | 0.006741893 |
| MAD2L1 | -2.842981966 | 1.29E-05 |
| ANXA5 | -0.902714368 | 0.006071532 |
| GUCY1A1 | 3.976390454 | 3.18E-06 |
| CEP44 | 0.739670587 | 0.029903149 |
| GASK1B | 1.41122679 | 0.002060208 |
| IL15 | 1.438048222 | 0.004376501 |
| HHIP | -4.117358733 | 0.021522367 |
| ITGA2 | -1.990494925 | 2.85E-11 |
| LMBRD2 | 0.890542338 | 0.02651234 |
| RANBP3L | 4.497178947 | 4.10E-08 |
| STARD4 | -1.035654469 | 0.028547197 |
| F2RL2 | 2.579378123 | 0.001307578 |
| ANKRD33B | 0.869008448 | 0.018120929 |
| RHOBTB3 | 1.107714854 | 0.003965362 |
| ENPP6 | 1.857195558 | 0.023872634 |
| ERAP1 | 1.304997834 | 2.06E-05 |
| EBF1 | 0.895000712 | 0.007230162 |
| ANKRA2 | 0.758602155 | 0.004109478 |
| TLR3 | 1.103049897 | 0.018330271 |
| GRIK2 | -1.325260832 | 0.008564533 |
| CREBRF | 0.912352336 | 0.005243576 |
| IL31RA | 2.069619979 | 0.005013037 |
| GALNT10 | -0.956750978 | 0.005200852 |
| SAP30L | 0.609199174 | 0.007923023 |
| MYOZ3 | -1.982482445 | 0.00284516 |
| GPR85 | -1.359929515 | 0.011528659 |
| PTTG1 | -4.303660387 | 3.09E-12 |
| BMPER | 1.072231775 | 0.034041129 |
| RELL2 | -0.970586119 | 0.020069278 |
| SLC29A4 | 2.408238413 | 5.49E-10 |
| ELAPOR2 | 2.610337019 | 8.71E-07 |
| SYTL3 | -1.332972529 | 0.013289902 |
| IQUB | 2.284699915 | 0.004171336 |
| ZNF704 | 1.310892876 | 0.042662866 |
| FABP5 | -2.36753966 | 2.73E-05 |
| ADCY1 | 1.634148283 | 0.010830596 |
| MED30 | 0.691264842 | 0.028251857 |

| | | |
|---|---|---|
| PHKG1 | 2.03422638 | 0.000859576 |
| EN2 | 2.575480242 | 0.002418911 |
| SUN1 | 0.613037655 | 0.04378774 |
| GPR146 | 1.503589304 | 0.007679556 |
| NOS3 | -2.306295271 | 0.011350577 |
| CDK5 | -0.728122058 | 0.031604094 |
| TMUB1 | -0.727861133 | 0.012683986 |
| YWHAZ | -0.67804476 | 0.003105541 |
| BAALC | -2.836888286 | 1.37E-06 |
| CTHRC1 | -1.46733056 | 0.009018141 |
| TP53INP1 | 1.195215729 | 0.000571647 |
| FREM1 | 2.175048357 | 0.000680183 |
| GEM | 1.997759657 | 9.92E-06 |
| PDP1 | -0.660192206 | 0.012087596 |
| TMEM67 | 1.079569273 | 0.007321585 |
| CCDC171 | 1.136645257 | 0.010269181 |
| DIRAS2 | 4.217883982 | 2.54E-07 |
| MAMDC2 | 1.624201568 | 0.043315388 |
| CPA6 | 3.54986749 | 0.048111447 |
| C8orf34 | 2.471998212 | 0.039895032 |
| HGSNAT | 0.529933112 | 0.034063446 |
| RASEF | -4.500555779 | 0.03523686 |
| KIF27 | 1.43842146 | 0.000296427 |
| SVEP1 | 1.98899423 | 3.16E-05 |
| VEGFD | 7.057794235 | 1.93E-06 |
| CARD19 | -0.801207071 | 0.007544071 |
| ZNF367 | -3.394095771 | 1.36E-06 |
| NOL6 | -0.70828729 | 0.008842845 |
| VCP | -0.758061736 | 0.004817399 |
| MELK | -3.026601322 | 9.15E-07 |
| OTUD1 | 0.878903442 | 0.009830847 |
| ARHGAP12 | 0.721643492 | 0.005810857 |
| TSHR | 2.082888822 | 0.000271702 |
| CFL2 | -0.940035849 | 0.015772617 |
| ZCCHC24 | 0.633693618 | 0.028819129 |
| GJB2 | 4.519563997 | 0.042630086 |
| SKA3 | -5.227660298 | 7.66E-12 |
| DDIAS | -3.351045678 | 4.45E-05 |
| LRR1 | -0.863821179 | 0.040036746 |
| DEPP1 | 2.075252702 | 1.20E-05 |
| BEND7 | 0.894296135 | 0.00045103 |
| VSTM4 | 2.101907334 | 1.61E-09 |

| | | |
|---|---|---|
| COMTD1 | -1.478150463 | 0.000939822 |
| PMPCA | -0.658722509 | 0.010382991 |
| AK8 | 1.252947715 | 0.015995815 |
| SLC39A2 | -4.502201859 | 0.049681205 |
| ZNF219 | 1.03838963 | 0.002450513 |
| SALL2 | 0.912460036 | 0.009617913 |
| E2F7 | -2.633761688 | 0.000168209 |
| ARHGAP42 | 0.94118763 | 0.015579511 |
| OTOGL | -1.260033694 | 0.015245811 |
| PSMC3 | -0.897099205 | 0.005856772 |
| SMCO2 | -2.569571024 | 0.030493556 |
| CLMN | 2.860662414 | 1.25E-09 |
| PTER | 0.899894088 | 0.008128918 |
| CACNB2 | 1.947178984 | 0.045391132 |
| HACD1 | -2.143589088 | 7.43E-07 |
| ABTB2 | -1.383552175 | 0.000793386 |
| LIPC | 1.410869036 | 0.0206843 |
| TCP11L2 | 0.901091384 | 0.013795706 |
| RAB8B | 0.879111517 | 0.025453517 |
| IKBIP | -0.857126802 | 0.000920656 |
| AVPR1A | 9.148382917 | 3.93E-05 |
| CKB | -2.794036509 | 6.07E-07 |
| HPS6 | -0.677231155 | 0.010842966 |
| GABRB3 | 2.710770184 | 1.35E-05 |
| CCT2 | -0.620396517 | 0.026268224 |
| MYRFL | 5.045352153 | 0.00210666 |
| BORCS7 | 0.653359612 | 0.045100075 |
| TMEM100 | 1.60092814 | 0.032271259 |
| SMPD1 | -1.094434678 | 0.001681822 |
| ILK | -0.86753091 | 0.005403023 |
| NETO1 | 3.111629045 | 0.001272735 |
| CYB5R2 | -1.032158532 | 0.001918384 |
| SERPINB7 | -3.168190666 | 0.005578802 |
| SERPINB8 | -0.658590174 | 0.016634896 |
| ZMAT1 | 1.654070643 | 0.001267566 |
| TRIM66 | 0.817385059 | 0.008160862 |
| CENPN | -1.893757527 | 0.000129734 |
| ATMIN | 0.555195805 | 0.024749279 |
| TMX3 | 0.653803116 | 0.029628313 |
| MFAP4 | 1.382563288 | 0.000359048 |
| ATF7IP2 | 2.483002252 | 0.001355913 |
| MMP10 | -5.09844336 | 1.48E-07 |

| | | |
|---|---|---|
| GOLM2 | 0.77765816 | 0.009924521 |
| NNMT | 0.913448611 | 0.00060133 |
| YPEL4 | 1.273063295 | 0.019517445 |
| PPIB | -0.679540177 | 0.032944139 |
| LDHC | -4.702265888 | 0.009818736 |
| PCLAF | -4.890778511 | 1.73E-09 |
| KIF7 | -0.920187347 | 0.013739088 |
| PEX11A | 0.572160129 | 0.039591805 |
| ANPEP | -0.921528542 | 0.016431846 |
| NAV2 | -1.819162174 | 0.000124559 |
| GLYATL1 | -2.54986719 | 0.019852773 |
| PLK1 | -4.769310614 | 6.85E-13 |
| NAB2 | -0.959948961 | 0.002308805 |
| ATP23 | 0.934341505 | 0.001233716 |
| ELFN2 | -1.899105075 | 0.018823509 |
| STX3 | -0.63947761 | 0.038916361 |
| MTMR10 | -0.934972444 | 0.01165536 |
| SCG5 | -1.715057965 | 0.040295357 |
| GREM1 | -1.806803124 | 1.31E-05 |
| DIS3L | 0.600503574 | 0.015654323 |
| MAP1A | -0.975909423 | 0.00094873 |
| AKTIP | 0.525192265 | 0.044465078 |
| PHB1 | -0.75315856 | 0.010557383 |
| SLC27A4 | -0.650719784 | 0.030264006 |
| PRRX2 | -1.442940532 | 0.001106303 |
| TBC1D2B | 0.694546256 | 0.005168852 |
| KATNAL2 | 0.863836866 | 0.019026858 |
| IGF2 | 1.599023375 | 0.009126226 |
| DUS2 | -0.799313772 | 0.003026903 |
| STIM1 | 0.649434077 | 0.021617792 |
| TRIM68 | 0.593510282 | 0.030976666 |
| ZNF23 | 1.181804822 | 0.047702183 |
| TPM4 | -1.045266569 | 0.000153705 |
| CDT1 | -3.391502436 | 1.63E-07 |
| CACNB3 | -0.596573603 | 0.044200276 |
| TUBA1A | -1.13209736 | 0.000239552 |
| TUBA1C | -1.843239187 | 1.52E-06 |
| ZNF610 | 1.042742663 | 0.009336276 |
| AXL | -1.086020134 | 0.001016143 |
| ZNF526 | -0.828007612 | 0.004613002 |
| YIF1B | -1.408600034 | 2.67E-05 |
| DAPK3 | -1.049546126 | 0.000689596 |

| | | |
|---|---|---|
| CHAF1A | -1.358263451 | 0.015743548 |
| HDGFL2 | -0.507856846 | 0.044017415 |
| SEMA6B | 1.874349472 | 0.001106669 |
| NXN | -1.170185459 | 0.000976521 |
| GPT | 2.708752989 | 0.043109964 |
| CYB5D2 | 0.656216481 | 0.028342883 |
| ZNF83 | 0.698056162 | 0.049217401 |
| KRT80 | -1.115465726 | 0.036605098 |
| ANGPTL4 | 2.977649452 | 4.03E-09 |
| CD320 | -1.359936594 | 0.000233036 |
| SPRYD3 | -1.104710278 | 0.000333825 |
| IGFBP6 | -2.891787073 | 2.47E-06 |
| ZNF558 | 0.738717605 | 0.010450115 |
| CDK2AP2 | -0.713843444 | 0.030309388 |
| TK1 | -5.582760136 | 4.93E-17 |
| TMEM68 | 0.537824589 | 0.038115206 |
| SOST | -7.076133485 | 9.88E-07 |
| ABCA3 | 2.674180253 | 1.65E-06 |
| BEST1 | 1.420671744 | 0.005725478 |
| FTH1 | 1.117539283 | 0.000435115 |
| TRANK1 | 0.949214278 | 0.009505371 |
| FADD | -0.617082271 | 0.013367401 |
| SAC3D1 | -1.008251474 | 0.025737129 |
| BATF2 | 2.225193242 | 0.031639366 |
| CCDC88B | -1.45286242 | 0.004465275 |
| SCARA3 | -2.862551948 | 3.85E-13 |
| PBK | -4.752374607 | 1.75E-09 |
| SCARA5 | 5.906954464 | 0.004854713 |
| COPS6 | -0.822602571 | 0.018347616 |
| NUDT16L1 | -0.863528215 | 0.013316264 |
| RBPJ | 0.536395271 | 0.021952446 |
| LMBRD1 | 0.726455935 | 0.002798746 |
| GLYCTK | -1.38368649 | 0.001037906 |
| GNG4 | -3.264589687 | 0.000240908 |
| NT5DC2 | -1.007779175 | 0.000862266 |
| PDHB | -0.630799145 | 0.041525686 |
| PCMTD1 | 0.995274325 | 0.004170857 |
| ACOX2 | 0.892378519 | 0.005216102 |
| IRF2 | 0.661607052 | 0.012702182 |
| DEGS2 | 3.42224318 | 1.05E-05 |
| MFSD2A | -1.472205127 | 0.036739148 |
| DTYMK | -1.685624306 | 0.000196602 |

| | | |
|---|---|---|
| MLKL | -0.859353793 | 0.022054556 |
| CMAHP | 1.814734725 | 2.20E-07 |
| RHOH | 4.377291418 | 0.041309923 |
| STIP1 | -0.585274855 | 0.030643875 |
| REEP4 | -0.782615004 | 0.04284215 |
| TNXB | 2.654241202 | 0.000272252 |
| FEN1 | -1.719181501 | 0.001543654 |
| SERINC2 | -1.633862422 | 0.009289256 |
| COL3A1 | 0.840084927 | 0.040998151 |
| SLC20A2 | -0.645865546 | 0.034873933 |
| STAT3 | 0.575306243 | 0.0231364 |
| SLC16A4 | -0.842890132 | 0.029588498 |
| IL7R | -3.872679203 | 1.26E-31 |
| TMEM208 | -1.100954679 | 0.004209195 |
| NPNT | 2.96349094 | 0.005048926 |
| SHOX2 | 1.03321414 | 8.71E-05 |
| NSG1 | -1.9382703 | 0.005675505 |
| FSTL5 | 4.894076667 | 0.014603338 |
| ATOH8 | -2.652882276 | 1.20E-06 |
| ENHO | -2.452758896 | 0.026013966 |
| ZNF608 | 1.614844579 | 0.008487227 |
| LETM1 | -0.56688835 | 0.019037513 |
| CEP120 | 0.798600646 | 0.032875357 |
| STXBP6 | -0.782692478 | 0.044456322 |
| TM4SF20 | 2.618640643 | 0.005612483 |
| GRM5 | 4.089297506 | 0.030033604 |
| JMJD7-PLA2G4B | 1.22211221 | 0.04444319 |
| IRS1 | 0.650446699 | 0.037561825 |
| PARM1 | -3.881701703 | 0.014539669 |
| FAM110B | 1.327113199 | 1.17E-07 |
| ODAD2 | 0.939010874 | 0.004314639 |
| ZNF354A | 0.622110807 | 0.023062095 |
| ATF5 | 0.943342362 | 0.023721286 |
| MN1 | -0.801906456 | 0.016116231 |
| APEX2 | -0.536973514 | 0.04990397 |
| RAB3B | -2.365074855 | 3.64E-10 |
| RSPO1 | 2.507787128 | 4.11E-07 |
| SH3TC2 | -2.92113092 | 4.56E-08 |
| ADRB2 | -3.787068594 | 0.020443829 |
| GPRIN1 | -1.29419648 | 0.003318622 |
| HSPB3 | -1.894833404 | 0.0375191 |
| P2RY12 | 5.506944754 | 0.000528173 |

| | | |
|---|---|---|
| SLC33A1 | 0.751399694 | 0.017989097 |
| DTWD2 | 1.21469191 | 0.001118087 |
| CLIC3 | -2.055634824 | 1.17E-06 |
| CKAP2L | -5.161157527 | 8.56E-05 |
| APLF | 0.689262538 | 0.014712167 |
| BUB1 | -4.826198967 | 3.26E-10 |
| CHRNA5 | -1.69470609 | 0.046334886 |
| CENPX | -1.674163818 | 4.91E-08 |
| ASPSCR1 | -0.708374351 | 0.02643936 |
| FASN | -1.444031253 | 1.41E-06 |
| MT1E | -0.756582299 | 0.032313176 |
| RAC3 | -0.806422826 | 0.026715791 |
| NLGN1 | 0.940623298 | 0.035924673 |
| TAPT1 | 0.559873162 | 0.028717536 |
| TAS2R1 | 2.478334372 | 0.029527977 |
| LINGO1 | 1.728473657 | 0.024133649 |
| ROBO1 | 0.76070236 | 0.048144405 |
| AVEN | -0.931404005 | 0.001106718 |
| CALML6 | 1.463228317 | 0.014337933 |
| PUSL1 | -0.747439445 | 0.013293364 |
| TIGD4 | 4.17189854 | 0.003716516 |
| ALCAM | -1.413153903 | 0.001393133 |
| TRAPPC1 | -0.718091165 | 0.038029123 |
| ZPLD1 | 3.522160176 | 0.003261223 |
| SERPINA9 | -5.885285561 | 0.001463486 |
| FOXD4 | 1.5470027 | 0.014558353 |
| HNRNPA3 | -0.634328879 | 0.014643454 |
| VGLL2 | 6.699483313 | 0.001206186 |
| ADRA1B | -1.70073835 | 0.002050357 |
| ADPRM | 0.730321952 | 0.022834717 |
| GON7 | -0.834121887 | 0.015115121 |
| FAXDC2 | 1.09315165 | 0.000956143 |
| CDK1 | -3.455119825 | 1.15E-06 |
| SETMAR | 0.578688055 | 0.045726929 |
| SMAD1 | 0.657084527 | 0.032827564 |
| TCAF2 | 0.940657706 | 0.019576849 |
| TMEM182 | 1.53687451 | 0.002001273 |
| HARS1 | -0.754439124 | 0.00136217 |
| CD14 | 2.083554141 | 0.004219695 |
| NPAS2 | 1.049890281 | 0.004607038 |
| LONRF2 | 6.077009811 | 2.68E-05 |
| ELOVL6 | -0.892776265 | 0.001575716 |

| | | |
|---|---|---|
| SMAGP | -1.110050469 | 0.00894106 |
| EMB | -2.714988167 | 0.000806459 |
| ACYP2 | 0.621802182 | 0.046125313 |
| CAVIN4 | -1.213784426 | 0.027301205 |
| CDCA4 | -1.426656554 | 0.000274521 |
| GPR27 | 1.433605393 | 0.018169859 |
| TMEM43 | 0.617111781 | 0.011085209 |
| TPT1-AS1 | 0.715061063 | 0.029334713 |
| TANC2 | -0.781046071 | 0.005439553 |
| CAVIN3 | -1.682374854 | 2.37E-05 |
| HAS2 | -1.043153504 | 0.009126915 |
| PDGFD | 2.452195598 | 4.30E-11 |
| PLAC1 | 3.058020965 | 0.033893855 |
| S1PR1 | -0.895993145 | 0.016825124 |
| PYGO1 | 0.783769535 | 0.044123633 |
| XKR6 | 3.358578586 | 1.76E-05 |
| INSR | 0.76843965 | 0.003910392 |
| PRKCE | 0.910840081 | 0.002466161 |
| MORN4 | 0.837037649 | 0.023298828 |
| JUNB | 1.011800314 | 0.04917324 |
| SHCBP1 | -5.067460861 | 5.39E-11 |
| NPTX1 | -2.447221347 | 0.001659443 |
| GAA | 1.259441645 | 0.00034607 |
| ZDHHC16 | -0.656951409 | 0.035237308 |
| ESCO2 | -3.710343469 | 7.93E-06 |
| KRT15 | -4.3001901 | 0.026629974 |
| APLN | 3.173959763 | 1.36E-05 |
| PDE7B | 1.26471216 | 0.004555817 |
| HOPX | -5.251645182 | 0.021850992 |
| LRRC8C | -1.374009415 | 0.014770417 |
| ETFDH | 0.589964045 | 0.018968558 |
| RXFP1 | 1.984136593 | 0.032326146 |
| PTGER4 | 1.985886727 | 9.19E-07 |
| CXXC5 | -0.736269424 | 0.041848176 |
| RGS19 | -0.667311906 | 0.044407828 |
| VAT1L | -4.27626219 | 0.016901075 |
| GATM | 4.180247966 | 0.018244753 |
| CTPS1 | -1.027413815 | 0.008521358 |
| KNDC1 | 2.215019968 | 0.010072574 |
| ZNF540 | 1.037094952 | 0.049256488 |
| FBXL14 | 0.845771091 | 0.004160737 |
| ZNF570 | 0.766179611 | 0.041354099 |

| | | |
|---|---|---|
| MLLT3 | 0.919871624 | 0.007429235 |
| FAM90A1 | 2.522868332 | 0.03459016 |
| RRM2 | -4.6098056 | 5.93E-05 |
| PRNP | 1.087277195 | 0.000575899 |
| KLF17 | -5.008223124 | 0.01190836 |
| FBXW10 | 4.161182276 | 0.001299344 |
| DRC3 | 1.119547214 | 0.005784114 |
| JMJD1C | 0.877925477 | 0.018084293 |
| SYNPO | 1.059399902 | 0.008708945 |
| ZNF556 | 4.211321018 | 0.029232145 |
| THOP1 | -0.946969804 | 0.004439965 |
| KLF11 | 1.602905743 | 1.25E-06 |
| KRCC1 | 0.664352483 | 0.006979334 |
| CYCS | -0.566618898 | 0.01988647 |
| CCL11 | 5.796970521 | 0.003125441 |
| FRMD3 | -2.304591729 | 0.002432645 |
| MRPL13 | -0.600357952 | 0.033138686 |
| PRL | 2.39046816 | 0.014699455 |
| ID4 | 1.114087187 | 0.015577118 |
| MACROD2 | 0.793539916 | 0.018374909 |
| CERS6 | 1.17777317 | 0.000543942 |
| CSDC2 | -3.090248986 | 0.000121844 |
| RCAN2 | 2.632361876 | 5.53E-07 |
| IL16 | 3.153761387 | 1.82E-14 |
| GNB2 | -0.710286666 | 0.022979184 |
| PRSS27 | 1.394982531 | 0.000983782 |
| GTPBP2 | 0.755023744 | 0.031657641 |
| FIBP | -0.827636792 | 0.009422384 |
| PPP1CA | -0.897316822 | 0.005119079 |
| SNTG2 | -2.257841764 | 0.024668041 |
| MRPL52 | -0.876908986 | 0.010572708 |
| SMPDL3A | 1.042578662 | 0.017349155 |
| EFEMP2 | -0.551246626 | 0.034559209 |
| ZMAT3 | 0.975624981 | 3.29E-05 |
| ZFAND4 | 0.863864113 | 0.010683713 |
| CORO1B | -1.055740376 | 0.007721919 |
| MUS81 | -0.70883384 | 0.008267192 |
| CFL1 | -1.549661139 | 1.09E-06 |
| ZNG1A | 0.866118297 | 0.025347407 |
| HOXC5 | -1.258188754 | 0.011106522 |
| CYP7B1 | 2.81174316 | 1.34E-05 |
| RARG | -1.023405229 | 0.002242173 |

| | | |
|---|---|---|
| DHCR7 | -1.297559962 | 0.018705842 |
| LVRN | 2.2355683 | 0.002833863 |
| LCLAT1 | 0.729083277 | 0.034462903 |
| EVC2 | 1.260292469 | 1.17E-05 |
| ZNF680 | 0.740841266 | 0.030645734 |
| HPSE | -1.440363729 | 0.011571753 |
| COQ2 | -0.822052219 | 0.013533542 |
| ESRRA | -0.79084851 | 0.00631519 |
| PARP14 | 1.056197898 | 5.05E-05 |
| PARP15 | -2.370736576 | 0.029023244 |
| ABLIM3 | -0.900214292 | 0.0401639 |
| SLC2A14 | -5.193394583 | 0.037385688 |
| SNCG | -4.36666118 | 1.14E-05 |
| STOX2 | 3.064941014 | 4.56E-07 |
| MAP3K11 | -0.809293552 | 0.004724986 |
| OLR1 | 1.627020619 | 0.002206379 |
| EHBP1L1 | -0.907880765 | 0.00790244 |
| THAP2 | 1.014767599 | 0.004260329 |
| PPP1R14B | -1.375116068 | 0.000102339 |
| ZNRD2 | -0.567129939 | 0.032413194 |
| VEGFB | 0.876032366 | 0.006698727 |
| MST1 | 1.413541612 | 0.001433605 |
| GMPPB | -0.689700106 | 0.029372753 |
| CSPG4 | -1.509442959 | 0.035278489 |
| SNX33 | 0.517021262 | 0.047896949 |
| SULT1B1 | 4.11508411 | 0.000117898 |
| LRFN4 | -1.447272284 | 1.22E-05 |
| HSPB7 | -2.021246076 | 0.000704541 |
| CD7 | 4.492344564 | 0.000503386 |
| NET1 | -1.268325334 | 0.001131031 |
| CBX2 | -1.172957372 | 0.047017301 |
| ATP5MK | -0.569502202 | 0.046137135 |
| CEP19 | 0.846538392 | 0.009531528 |
| CD34 | -1.909845194 | 0.026499627 |
| CTSF | 1.043242276 | 0.000147069 |
| UQCC4 | -0.689283283 | 0.028119169 |
| TLR1 | 3.94868014 | 7.62E-07 |
| FAM174A | 0.618495784 | 0.024167618 |
| DIRC1 | 8.449948389 | 2.03E-08 |
| CHRNA9 | -3.320691307 | 0.00264799 |
| KCNJ5-AS1 | -1.59279543 | 0.023835843 |
| EXO1 | -3.045489153 | 5.70E-05 |

| | | |
|---|---|---|
| LIG4 | 0.87860416 | 0.007921232 |
| ATP2A2 | -0.925433704 | 0.031062705 |
| ZWILCH | -1.515845025 | 0.002798293 |
| CNTNAP2 | 2.970895929 | 0.034157306 |
| MYO1H | 6.784785264 | 0.00041433 |
| IL20RB | 1.294968364 | 0.01830823 |
| NPAS4 | -2.34291472 | 0.000243607 |
| CMKLR1 | 3.164213788 | 1.20E-05 |
| KY | -2.290170034 | 0.016505266 |
| IQCK | 0.865038975 | 0.031169607 |
| SLCO2A1 | -1.597261278 | 0.000769643 |
| BRSK2 | 1.911425392 | 0.028066009 |
| B4GAT1 | 1.011709651 | 0.000392364 |
| LEP | 4.618836072 | 0.003113142 |
| RESF1 | 0.784212631 | 0.027073344 |
| BRMS1 | -0.77049799 | 0.006736668 |
| HRAS | -0.631209527 | 0.033235455 |
| RIN1 | -1.369782602 | 0.000628695 |
| PDZK1 | -2.694729853 | 3.22E-06 |
| YIF1A | -0.745813532 | 0.018126362 |
| CNIH2 | -1.361915669 | 0.034992303 |
| RAB1B | -0.821831201 | 0.014139889 |
| MICOS13 | -0.782389541 | 0.016473162 |
| ASPHD1 | -1.393035069 | 0.000807457 |
| P2RY14 | 4.106533329 | 0.039147713 |
| CHST2 | 1.228755476 | 0.0189491 |
| ZDHHC14 | 0.82825416 | 0.03451804 |
| UBE2C | -5.255759046 | 2.41E-12 |
| DES | -2.916719446 | 1.20E-05 |
| PACS1 | -0.696018837 | 0.004594269 |
| SH3BP5L | -0.96997931 | 0.001098245 |
| YPEL2 | 1.027540445 | 0.003524107 |
| PSMD2 | -0.818233113 | 0.007876882 |
| PPM1E | 2.282181965 | 0.003064893 |
| CSRP2 | -1.216901567 | 0.01706006 |
| DDIT3 | 1.338279636 | 4.28E-05 |
| ARHGAP1 | -0.843938919 | 0.007325346 |
| MED16 | -0.63491764 | 0.049153803 |
| CHST1 | 2.48779639 | 0.017264121 |
| CATSPER1 | -4.397421141 | 0.000557105 |
| CCNE2 | -3.22018895 | 9.06E-07 |
| BANF1 | -0.695632589 | 0.036344168 |

| | | |
|---|---|---|
| NRIP3 | -0.970084111 | 0.02470547 |
| SCUBE2 | 1.184330358 | 0.010070003 |
| ZNF25 | 0.704432887 | 0.028695779 |
| SART1 | -0.54009474 | 0.046896977 |
| ALG10B | 1.1994238 | 0.015453967 |
| DRAP1 | -1.260674202 | 0.000400137 |
| UCP2 | -3.515904806 | 6.22E-06 |
| C11orf68 | -0.810462768 | 0.021814304 |
| MRPL48 | -0.583331986 | 0.049578868 |
| P2RY2 | -3.800306782 | 0.012655113 |
| FOSL1 | -1.674600063 | 4.16E-05 |
| CCDC85B | -1.378727292 | 0.001465868 |
| RMI2 | -1.602302388 | 0.002275875 |
| TOM1L2 | -0.481205533 | 0.033563036 |
| GPR156 | -2.439690834 | 7.00E-05 |
| MTLN | -0.751841933 | 0.03544953 |
| LINC02915 | -2.95058542 | 0.000169985 |
| AURKAIP1 | -0.969372562 | 0.003443107 |
| PRIMA1 | 2.772611803 | 0.000462198 |
| BAIAP2 | -1.258499245 | 0.005894019 |
| DOK7 | -6.272568348 | 6.21E-05 |
| LRRN1 | 3.205387065 | 0.010773376 |
| UNC119B | 0.493146627 | 0.044050102 |
| TUBB6 | -1.297162839 | 4.15E-06 |
| MBLAC2 | 0.798461661 | 0.023369174 |
| SSNA1 | -0.68674463 | 0.025837096 |
| MC5R | 2.590198571 | 0.009164279 |
| SPHK1 | -1.628904268 | 9.96E-06 |
| BNIP3 | 0.948607087 | 0.019036383 |
| ATAD5 | -1.952027811 | 0.011218587 |
| ZNF404 | 1.12529241 | 0.000111553 |
| RTTN | -0.835918534 | 0.03071514 |
| RPP38-DT | -2.532985452 | 0.023357586 |
| ACBD7 | -1.760678338 | 0.017437724 |
| ZNF135 | 0.579855299 | 0.03845937 |
| COX8A | -1.07242785 | 0.00320906 |
| RNPEP | -0.723702993 | 0.012308831 |
| CLEC14A | -2.386366177 | 0.000493499 |
| LPCAT4 | -0.812648197 | 0.00484788 |
| SLCO3A1 | 0.63872018 | 0.043178026 |
| DIRAS1 | -1.031688162 | 0.003736528 |
| PHLDB3 | 0.764193891 | 0.030439262 |

| | | |
|---|---|---|
| KBTBD11 | 1.232819626 | 0.00279064 |
| LMNB2 | -1.826933701 | 5.86E-05 |
| RNF152 | 1.671809235 | 2.76E-06 |
| FOXL1 | -1.391248832 | 0.004034284 |
| FOXC2 | -2.916047502 | 0.002002003 |
| BDNF | -1.521073423 | 0.024202271 |
| CCDC121 | 1.419808582 | 0.000563157 |
| BOK | -0.751051333 | 0.016315952 |
| ZNF843 | 0.984948576 | 0.016676185 |
| CDK5R1 | -0.841248219 | 0.033851963 |
| NCKAP5 | 2.925036951 | 1.12E-08 |
| LRRC37A3 | 2.076032707 | 1.06E-10 |
| FKBP9P1 | 1.341253689 | 0.015308675 |
| VSIG10 | -0.799409262 | 0.015391929 |
| WSB2 | -0.56508197 | 0.031766018 |
| TYMS | -2.173108528 | 5.90E-05 |
| TCEANC | 0.999614389 | 0.010880266 |
| TCIM | 2.352270608 | 0.001038119 |
| MAMSTR | -1.554035849 | 0.001773668 |
| GCNT4 | 0.957019861 | 0.028433013 |
| FIBIN | 2.491274785 | 6.04E-15 |
| DEAF1 | 0.901315299 | 0.00418248 |
| FBXO46 | -0.618737649 | 0.022742764 |
| SLC38A9 | 0.56919116 | 0.049617732 |
| POLE | -1.014829214 | 0.037849149 |
| RHOG | -0.747328434 | 0.023229191 |
| ZBTB34 | 0.789306895 | 0.032231424 |
| FBXO39 | 2.183966455 | 0.047870257 |
| DLGAP1-AS1 | 1.029063733 | 0.013989969 |
| LRRN4CL | 2.864942863 | 0.026961499 |
| HIC1 | -0.958559716 | 0.004720608 |
| NINJ2-AS1 | 0.9933096 | 0.000977589 |
| SAMD9L | 1.107582506 | 0.001891284 |
| ZFAS1 | 0.859910014 | 0.00830244 |
| TGIF1 | 0.688571886 | 0.004047584 |
| NIM1K | 1.193117497 | 0.021471915 |
| ERICH5 | 2.03508465 | 0.021019953 |
| CAVIN1 | -1.047712266 | 8.90E-05 |
| SLC25A22 | -0.769432844 | 0.020580837 |
| RABEP2 | -0.690437732 | 0.016461624 |
| ATOX1 | -0.946335763 | 0.008605611 |
| TBL1XR1 | 0.574559928 | 0.019785863 |

| | | |
|---|---|---|
| HASPIN | -3.025140765 | 0.000154694 |
| GBA1 | -0.644969933 | 0.021983943 |
| CD163L1 | -1.654952562 | 0.005394207 |
| CRACR2B | -2.681740954 | 0.034917646 |
| NAALADL2 | 0.822547239 | 0.001118866 |
| CD151 | -0.830711659 | 0.017013585 |
| POLR2L | -1.065773777 | 0.018648645 |
| FLII | -0.594968668 | 0.011663238 |
| HNRNPA0 | 0.496798126 | 0.040111822 |
| FAM87B | -2.689713564 | 0.003282558 |
| TENM3-AS1 | -1.301639485 | 0.048753779 |
| ZNF620 | 1.460411477 | 0.024655442 |
| UBE2N | -0.782945737 | 0.004647864 |
| RIC8A | -0.556958737 | 0.027284963 |
| DPY19L2 | 1.312823063 | 0.005591987 |
| NDUFAF3 | -0.724580477 | 0.047536983 |
| GRAMD1C | -1.885196761 | 0.000317316 |
| PPP1R42 | 2.851052009 | 0.047627357 |
| ZNF114 | -3.393948119 | 0.003342241 |
| ZC3H12D | -2.091443334 | 0.003047976 |
| PLEC | -1.276593203 | 1.31E-05 |
| GEN1 | -1.386445487 | 0.000899127 |
| PLEKHM3 | 0.724757577 | 0.031825584 |
| DNAJC22 | -2.074334867 | 0.001980381 |
| MAF | 2.18403444 | 5.16E-05 |
| GTPBP6 | -1.018403134 | 0.000813576 |
| ERN1 | 1.654957867 | 2.53E-05 |
| DHFR2 | 1.025500842 | 0.001390248 |
| RPP25 | -1.329767919 | 0.001416116 |
| THBD | -3.074112597 | 1.52E-05 |
| ZHX2 | 0.705708435 | 0.032658888 |
| CPNE7 | -3.733432792 | 1.73E-12 |
| MSC | 1.220162557 | 0.001316446 |
| HYI | -1.087454152 | 0.000890035 |
| AURKB | -5.371239567 | 1.70E-12 |
| DPM3 | -1.011392278 | 0.017286721 |
| FARSA | -0.563894644 | 0.027353623 |
| CALR | -1.016382655 | 0.009213268 |
| LINC00311 | -2.801979438 | 0.008279383 |
| MAGED1 | -1.124819979 | 0.000344183 |
| GADD45GIP1 | -0.811233223 | 0.009518773 |
| NSUN7 | 1.773142429 | 0.004346388 |

| | | |
|---|---|---|
| GATA2 | -1.666377476 | 0.00084016 |
| EGR3 | 1.648235266 | 0.007547681 |
| CATSPERE | 1.084152169 | 0.042116297 |
| ZBTB42 | -1.322977063 | 0.000162693 |
| LACC1 | 0.852180602 | 0.024392911 |
| RPRML | 3.153803615 | 0.042395235 |
| PCED1B | -1.128229999 | 0.018369227 |
| PIPOX | 2.171362992 | 0.016469067 |
| CDH5 | 2.094617158 | 0.00333706 |
| PCBP1-AS1 | 0.919396133 | 0.002808563 |
| MYADM | -0.791009215 | 0.021669879 |
| MROH1 | -0.6595859 | 0.030675511 |
| AKAP5 | -2.166776212 | 0.009032728 |
| PUF60 | -0.68909805 | 0.0085287 |
| SSC5D | -1.51134223 | 0.005100784 |
| DCTPP1 | -0.758021137 | 0.037444653 |
| PPP1R14BP3 | -1.436322047 | 0.013887055 |
| FAHD1 | -0.65746602 | 0.022895659 |
| TDRP | 1.873165767 | 0.001175142 |
| FZD2 | -0.689685042 | 0.033663692 |
| MCFD2 | 0.548526025 | 0.02324506 |
| GAS1 | 0.694636221 | 0.043679133 |
| ZNF571 | 0.893635474 | 0.00741704 |
| GLIPR1L2 | 2.689035687 | 2.20E-05 |
| RNF182 | -1.296006208 | 0.001789956 |
| H2AC6 | 0.984618728 | 0.000752483 |
| EIF2S3B | 2.852551457 | 0.014298058 |
| SLC47A2 | 6.086671486 | 0.046249509 |
| S1PR5 | -2.753879787 | 0.027228858 |
| ZFP3 | 0.64921899 | 0.020278714 |
| BHLHE22 | 6.839276189 | 2.10E-07 |
| PDIA3P1 | -0.805265295 | 0.022372528 |
| GREM2 | -2.095186147 | 2.58E-06 |
| CAPS2 | 1.773911576 | 2.19E-06 |
| OXTR | -2.826518641 | 0.012927902 |
| FAM83H | 1.082071276 | 0.003869581 |
| GPR137C | -2.050648996 | 0.003357058 |
| TRAPPC5 | 1.0992004 | 0.0107591 |
| FCRL6 | 3.772166874 | 0.016026255 |
| CHRM2 | -3.545988245 | 0.007162066 |
| NPM1 | -0.58743479 | 0.040780596 |
| TMEM102 | -1.248117822 | 0.004835845 |

| | | |
|---|---|---|
| ZNF678 | 0.952810002 | 0.048934573 |
| TMEM45A | 0.766744323 | 0.00571859 |
| RNF135 | 0.625443627 | 0.012231109 |
| FANCB | -2.106996209 | 0.001033673 |
| TNFSF15 | 7.042407418 | 2.56E-05 |
| ZFP41 | 0.65392862 | 0.017248793 |
| PHLDA2 | -2.24948892 | 5.46E-05 |
| ATG9B | -2.399013203 | 0.033130761 |
| ZNF875 | 0.523011835 | 0.025133943 |
| YIPF6 | 0.53812613 | 0.045947581 |
| DIPK2A | 1.514003454 | 0.000538427 |
| MACIR | -0.882352504 | 0.03485119 |
| AMIGO1 | 1.1610304 | 0.002205947 |
| GPR3 | -2.06353286 | 0.008504696 |
| SIAH2 | 0.844071765 | 0.003284717 |
| COPG1 | -0.710587668 | 0.01317949 |
| SLC9A9 | 1.987452041 | 0.001205105 |
| RELL1 | 0.780002033 | 0.03659384 |
| CLDN7 | -2.969234789 | 0.003298121 |
| TDRKH | -1.964681067 | 0.003471006 |
| MRPL41 | -0.987336388 | 0.003672671 |
| RGMA | 1.101488761 | 0.017541463 |
| LDOC1 | -1.114558726 | 0.008331344 |
| EXT1 | -0.807056318 | 0.002022961 |
| HHIPL1 | -1.032989504 | 0.010569232 |
| C1S | 1.7428744 | 1.15E-05 |
| CACNB4 | 2.554517373 | 0.000318959 |
| PGBD4 | 0.729014942 | 0.017296093 |
| TSHZ2 | 1.096766078 | 0.0004331 |
| FES | -0.954567604 | 0.016008081 |
| LIMK2 | -1.008327309 | 0.000770336 |
| ADI1 | -0.798234271 | 0.041768323 |
| SATB1 | 1.071268448 | 0.008242627 |
| TSPAN10 | -1.703034747 | 0.012625928 |
| SKA2 | -0.783120145 | 0.024205248 |
| ANXA2 | -1.223266035 | 1.27E-05 |
| PAPPA | 2.068624375 | 6.57E-08 |
| CRIP2 | -1.974193136 | 2.85E-09 |
| EWSR1 | -0.739113347 | 0.001862 |
| GJC1 | -1.443941353 | 0.000564389 |
| PYCR1 | -0.734036297 | 0.024963483 |
| SLC8A1 | -1.409219216 | 0.004093803 |

| | | |
|---|---|---|
| ABAT | -1.124111705 | 0.00904161 |
| GPC6 | 1.071563212 | 0.029253166 |
| ARHGEF37 | 1.829395461 | 0.000139023 |
| SMDT1 | -1.04089617 | 0.013676909 |
| RUVBL2 | -1.105640358 | 0.002772292 |
| PLGLB1 | 2.166027858 | 0.004137806 |
| FHL3 | -0.750890559 | 0.013618934 |
| RIPK4 | -1.874457628 | 0.004775831 |
| GRIN2A | -4.171439253 | 5.18E-08 |
| TREX2 | 1.098563023 | 0.043709851 |
| MX2 | 1.349376754 | 0.022300926 |
| COA5 | 0.485128499 | 0.049892806 |
| PCBP3 | 1.989497774 | 0.003945257 |
| TNFAIP8L3 | -1.376948771 | 0.020372393 |
| FBXL7 | 0.816094609 | 0.029983718 |
| ZNF438 | 0.687766306 | 0.015084775 |
| PSG9 | -1.894530439 | 0.049663184 |
| CMKLR2 | 1.371726138 | 0.000451625 |
| ALYREF | -1.081932041 | 0.000555838 |
| RFLNB | -1.164812205 | 0.001068592 |
| EFHC2 | 2.604432639 | 0.028897807 |
| NOG | -1.69942783 | 0.001312224 |
| TBL3 | -0.722963272 | 0.013511431 |
| ACP7 | -3.028238368 | 0.027272773 |
| TRAIP | -1.339191253 | 0.017632077 |
| B3GALT5 | -2.943130637 | 0.003799909 |
| ZNF703 | -1.402093934 | 0.001669573 |
| IQGAP3 | -4.991946667 | 5.27E-13 |
| ARSI | -1.942608864 | 9.93E-07 |
| PRKX | 0.633028115 | 0.030106025 |
| SMTN | -1.25451066 | 0.000103525 |
| NPB | -2.246760182 | 0.000483863 |
| ACTG1 | -1.51174007 | 2.33E-07 |
| TBX1 | 2.27988969 | 0.002389665 |
| ADRA2C | -1.610993353 | 0.021840011 |
| CRELD2 | -1.055315893 | 0.002793111 |
| SCFD2 | -1.906135623 | 1.52E-09 |
| GPR173 | -0.887585075 | 0.03343959 |
| TSPYL2 | 0.670248493 | 0.043762031 |
| PGP | -0.775022413 | 0.014537749 |
| PCDH9 | -1.103707133 | 0.041995159 |
| ACOT1 | -1.368927883 | 0.006852167 |

| | | |
|---|---|---|
| OAF | -0.901304564 | 0.027664217 |
| H2AC20 | -3.099425459 | 0.006122934 |
| POU6F1 | 0.826014122 | 0.007433485 |
| TSSC4 | -0.709005641 | 0.015201391 |
| PRKD1 | -1.456211175 | 0.000303966 |
| ZDHHC23 | -1.688956629 | 0.000260002 |
| SRPK3 | -2.425219156 | 0.000177763 |
| SLIT3 | 1.729568207 | 5.34E-06 |
| EFNA5 | 1.507585006 | 0.018253876 |
| PKP3 | -2.013083744 | 0.000281026 |
| CSF1 | 1.076432782 | 0.000594289 |
| KNTC1 | -1.317111999 | 0.014605068 |
| TXNRD2 | -0.747372027 | 0.014950379 |
| POU3F2 | -2.283692777 | 0.033340128 |
| CEND1 | -2.021753211 | 0.000569256 |
| C6orf58 | 1.805067961 | 0.008164082 |
| SOCS3 | 1.347762944 | 0.009351042 |
| XPOT | 0.84354317 | 0.019552069 |
| STING1 | -0.708692275 | 0.048101987 |
| CDCA2 | -3.644024869 | 8.38E-07 |
| OSBP2 | 1.941866854 | 0.029230369 |
| TMED9 | -0.835607578 | 0.005624828 |
| DRD1 | 2.092651997 | 0.014980823 |
| BTBD6 | -0.705932803 | 0.016215443 |
| RBM43 | 0.95438335 | 0.000264919 |
| ZFP90 | 0.641823315 | 0.0169139 |
| FAM227A | 1.263010038 | 0.025366406 |
| NOC4L | -0.564190361 | 0.034786373 |
| TMEM121 | 0.780611152 | 0.009485535 |
| TMEM106A | -1.430274893 | 2.03E-05 |
| ROBO2 | 4.135217136 | 0.000216451 |
| MAFF | 0.642009265 | 0.03046418 |
| CIB1 | -0.635914808 | 0.040763548 |
| SLC24A3 | 3.812148843 | 1.87E-07 |
| MANEAL | 1.188140787 | 0.00582529 |
| PRSS57 | -3.540810063 | 0.03986256 |
| PPIL6 | 1.675410121 | 0.000640811 |
| NOTUM | -6.153452066 | 6.00E-05 |
| GALNT17 | 7.447171156 | 1.00E-05 |
| CCDC137 | -0.796532797 | 0.00063055 |
| C12orf56 | -3.762759081 | 0.000197814 |
| TMEM105 | 4.093954575 | 0.013401424 |

| | | |
|---|---|---|
| GAS2L1 | -0.892207521 | 0.011049558 |
| TEDC1 | -2.013072558 | 0.000390478 |
| HS6ST3 | 2.794976919 | 0.005557946 |
| HGS | -0.842442344 | 0.001498674 |
| TNFAIP8L1 | -1.930330801 | 0.001627199 |
| FAM174B | 1.675256496 | 2.76E-06 |
| PARPBP | -1.422366507 | 0.014097747 |
| MUC1 | 0.85003933 | 0.014419653 |
| FAM131C | -2.347821282 | 0.000102382 |
| PDE6G | -2.556162802 | 0.025517366 |
| LSAMP | 2.264242275 | 9.75E-07 |
| AHNAK2 | -0.938493982 | 0.009432911 |
| OLFML2A | 1.954821174 | 0.003465764 |
| INKA1 | -1.052093815 | 0.048155153 |
| NDUFA4L2 | 2.053916079 | 0.001301833 |
| MYBL1 | -3.412470042 | 2.98E-07 |
| C11orf87 | 3.022828408 | 9.16E-26 |
| IFIT1 | 0.904180782 | 0.042156899 |
| SLC52A2 | -0.836289722 | 0.004416042 |
| PCYT2 | -1.652722932 | 7.26E-06 |
| GNB1L | -1.239902584 | 0.001933316 |
| TRIM69 | 1.02701665 | 0.000482385 |
| KLHDC8B | -1.040616892 | 0.000306863 |
| PTCH1 | 0.936023244 | 0.020366989 |
| SHOX | 5.288153389 | 0.003774297 |
| CCIN | -1.961599899 | 0.000292509 |
| SDHAP3 | 3.150535385 | 0.038370525 |
| ZNF566 | 0.694924128 | 0.044915999 |
| MATN1-AS1 | 1.470112134 | 0.003426993 |
| GSAP | 1.001880467 | 0.027116428 |
| CYP2R1 | 1.073362766 | 0.000946684 |
| ANKRD46 | 0.972583001 | 0.005069988 |
| WWOX | 1.228979112 | 6.64E-05 |
| BCL9L | -0.497874379 | 0.036245247 |
| KIF18B | -5.424869829 | 3.79E-11 |
| SAPCD2 | -5.893963617 | 0.000491998 |
| GPAT2 | -1.884176998 | 0.032150356 |
| MST1P2 | 2.19050752 | 5.06E-05 |
| THBS2 | 1.556554482 | 0.001762088 |
| CCDC30 | 1.327052522 | 0.001466755 |
| GLDN | 4.280636877 | 0.017995697 |
| GNG2 | -1.439360816 | 3.15E-06 |

| | | |
|---|---|---|
| INSIG1 | -1.088729276 | 0.01761871 |
| ZNF396 | 2.077840205 | 6.90E-05 |
| NF2 | -0.73026098 | 0.007217497 |
| SMIM29 | -0.95904026 | 0.002205519 |
| UBE2H | 0.522567445 | 0.043069495 |
| HPDL | -2.242632862 | 0.001760088 |
| KTN1-AS1 | 1.216407307 | 0.004744482 |
| KIF24 | -3.020871628 | 2.13E-05 |
| C17orf58 | 1.611348081 | 7.23E-06 |
| BCDIN3D | 0.601899525 | 0.033071761 |
| HYAL3 | -1.452536233 | 0.003510317 |
| MAPT | 2.410876905 | 0.000386009 |
| ERCC6L | -4.889083637 | 8.39E-09 |
| ZNF395 | 1.173432537 | 0.000271675 |
| TMEM232 | 1.605842284 | 0.002033997 |
| EFCAB6 | 1.026503535 | 0.017245337 |
| PTRH1 | -1.141637537 | 0.003469181 |
| PLCD1 | -1.005420566 | 0.009609347 |
| MITF | 2.016661524 | 0.002234329 |
| SHTN1 | -1.390041245 | 0.001919404 |
| ZNF546 | 0.796256187 | 0.017832997 |
| GCNT1 | -0.570601187 | 0.040304599 |
| TAF9B | -0.711777772 | 0.029658857 |
| PCDHB13 | 1.528538682 | 0.01574753 |
| PLEKHN1 | -3.705797675 | 5.03E-10 |
| TET3 | 1.484069228 | 7.57E-06 |
| SAMD11 | -3.34077208 | 3.38E-07 |
| TMSB4XP8 | -0.89567481 | 0.024426169 |
| TCEA1 | 1.155456624 | 0.00173993 |
| FANCA | -3.051702104 | 1.84E-05 |
| ADH1A | 5.562780875 | 0.007608676 |
| SEMA4D | 1.873292872 | 0.006042313 |
| MCRS1 | -0.745712969 | 0.006953507 |
| PEAR1 | -1.636669094 | 1.19E-06 |
| PABIR1 | 0.601064227 | 0.011718804 |
| PALM3 | -2.076960334 | 0.043009657 |
| DNER | -3.486530988 | 6.62E-05 |
| KLHL17 | -0.785660444 | 0.019413054 |
| TPRG1 | 3.587920399 | 0.002994774 |
| ARL4C | 1.443825572 | 0.000350531 |
| WNT7B | -5.346207126 | 0.006589626 |
| MAPK12 | -0.617040412 | 0.033553547 |

| | | |
|---|---|---|
| ZC3H6 | 1.16039919 | 0.000188141 |
| NCR3LG1 | -1.305666884 | 0.014456558 |
| POTEE | -1.788810427 | 0.013826046 |
| TUBB4B | -1.719882707 | 1.73E-05 |
| ZNF383 | 0.706814297 | 0.016815221 |
| HES4 | 1.135765629 | 0.021050852 |
| PLSCR1 | 0.780241358 | 0.003600081 |
| FOCAD | -0.760848162 | 0.003243522 |
| JAKMIP3 | 1.848259228 | 4.74E-05 |
| CLEC2A | 1.783391539 | 0.001814024 |
| IER5L | -1.44175167 | 1.38E-05 |
| H2AX | -1.513485561 | 0.00044496 |
| LCTL | -1.988957754 | 0.00353124 |
| FAM83G | -1.399378358 | 0.001054894 |
| FBLL1 | 2.021986878 | 0.016986242 |
| KRTAP1-1 | -4.010834273 | 0.001266985 |
| FAM72B | -1.314206559 | 0.01952671 |
| NANOS1 | 3.923805429 | 1.98E-10 |
| GOLGA8M | 1.98062766 | 0.000654728 |
| SAXO2 | 1.948077431 | 0.003858327 |
| H1-9P | -1.802155393 | 0.041602307 |
| PARVB | -0.757476121 | 0.041143024 |
| ZBED10P | -1.408509532 | 0.03792389 |
| ZNF548 | 0.543121599 | 0.041686003 |
| NHLRC3 | 1.089512607 | 0.003232967 |
| LINC00910 | 1.008146108 | 0.013803501 |
| LRRK2 | 1.13731476 | 0.002441034 |
| INSYN2A | -1.844260475 | 0.048725903 |
| FAM120AOS | 0.556249564 | 0.020908649 |
| UTS2B | 1.484128663 | 0.009119743 |
| NOC2L | -0.794033827 | 0.007556005 |
| NELFB | -0.670474503 | 0.011643888 |
| KCTD21 | -0.602752705 | 0.044706818 |
| SBSN | -2.987064453 | 1.14E-08 |
| CGB5 | -5.821990752 | 0.026545758 |
| RELN | -4.704615099 | 4.61E-06 |
| FAM111B | -4.068952645 | 1.29E-07 |
| APOD | 2.939579302 | 2.15E-05 |
| SF3B3 | -0.72059048 | 0.014698502 |
| NKAPL | -1.03774995 | 0.013065475 |
| JPT1 | -1.692872636 | 2.16E-06 |
| ZNF527 | 0.724554145 | 0.030941634 |

| | | |
|---|---|---|
| PCDH18 | -0.642466593 | 0.024576157 |
| BTBD8 | 0.811324386 | 0.025411009 |
| MAOA | 1.511730505 | 0.007047961 |
| C15orf61 | 1.137664152 | 0.000158717 |
| FHIT | 1.899298946 | 0.000593995 |
| RRP7A | -0.691895997 | 0.014165728 |
| FAM180A | -1.569624654 | 0.003133085 |
| HMGB1 | -0.766175665 | 0.005045438 |
| SPATA41 | 3.360000207 | 0.00431106 |
| IL1RAP | 1.473936971 | 0.000183886 |
| TDRD7 | -0.673832218 | 0.020940724 |
| AKR1C3 | 1.929212774 | 0.000331049 |
| WDSUB1 | 0.87428148 | 0.045615742 |
| COLCA1 | 3.201067846 | 0.024253265 |
| ACADSB | 0.6024485 | 0.030175871 |
| HRCT1 | -1.733543817 | 0.000549946 |
| GREB1 | 5.388691398 | 0.004222909 |
| SRGAP3 | 1.026069637 | 0.022317708 |
| TUBB | -0.942225478 | 0.000646937 |
| PPIA | -0.884797169 | 0.014536875 |
| ZNF493 | 0.974242405 | 0.030101775 |
| ZBTB44 | 0.860191265 | 0.019345496 |
| CGB7 | -2.932675281 | 0.024804946 |
| SRGAP2B | 0.90835209 | 0.026563804 |
| ARMCX4 | -0.881343287 | 0.002873813 |
| RFX8 | -1.452140939 | 0.002378819 |
| MYO18A | -0.511202976 | 0.030387534 |
| BORCS6 | -0.66329456 | 0.028568123 |
| MAN2A2 | 0.852552724 | 0.001167448 |
| SULF2 | 1.826323233 | 4.44E-14 |
| LAMA2 | 1.065354045 | 0.012062729 |
| XRCC2 | -1.67167479 | 0.032907811 |
| MMP1 | -1.922036228 | 4.11E-05 |
| ADH1B | 7.146855003 | 6.31E-11 |
| ZNF136 | 0.768513139 | 0.008601907 |
| TOMM7 | 0.703163284 | 0.005813431 |
| DAPK1 | 1.49565413 | 3.84E-06 |
| SNHG17 | 0.613088115 | 0.039567676 |
| CD47 | 0.825815727 | 0.003402364 |
| TLE1 | -1.157989292 | 0.019166044 |
| H2AC11 | -1.868169677 | 0.044176192 |
| ADA | -1.075524203 | 0.005111378 |

| | | |
|---|---|---|
| LAMB3 | -1.304439674 | 0.010598143 |
| KPNA5 | 0.944721296 | 0.023382647 |
| PDLIM7 | -1.154294326 | 0.004818908 |
| FLNA | -1.387678766 | 5.42E-05 |
| TMEM26 | -2.695346013 | 0.003830984 |
| SCOC-AS1 | 1.636422792 | 0.017046883 |
| CASP4 | 0.858612404 | 0.018930492 |
| AP2A1 | -0.532031329 | 0.031674354 |
| SERTAD1 | -1.163554059 | 0.000907858 |
| ANXA6 | -1.19426518 | 0.000242141 |
| SIGLEC15 | -4.517701098 | 2.24E-08 |
| MAFG | 0.628554845 | 0.041250439 |
| ARRDC1 | -0.678416659 | 0.02098424 |
| IGF2R | 0.784348013 | 0.033816755 |
| GAL3ST4 | -1.119585632 | 0.046470935 |
| SLC6A17 | -1.782872863 | 0.014057849 |
| SRC | -0.692896758 | 0.029247678 |
| PCNX3 | -0.478190822 | 0.035992867 |
| ABCB8 | -0.696481345 | 0.010984025 |
| SND1 | -0.538563365 | 0.046208278 |
| NEK5 | 2.29317561 | 0.015356014 |
| MIRLET7BHG | -1.169423882 | 0.000968215 |
| SERPINA1 | -2.925617782 | 0.00092499 |
| KANK2 | -0.591992271 | 0.032400467 |
| BLM | -3.001036681 | 0.000630369 |
| HMGA2-AS1 | -2.345026174 | 0.000736552 |
| SVIL | 1.245685715 | 0.011146519 |
| PELI1 | 1.178441596 | 0.01593942 |
| UAP1L1 | -0.779472221 | 0.019583435 |
| SLC22A5 | 0.745330403 | 0.035290616 |
| DACT3 | -0.964142343 | 0.000350801 |
| ADARB1 | 0.948626068 | 0.028379885 |
| HTT | -0.760554952 | 0.002271979 |
| DCHS2 | 3.560312754 | 4.04E-08 |
| HNRNPAB | -0.592567718 | 0.047498734 |
| COL13A1 | -2.403806608 | 7.27E-07 |
| PCDHB11 | 2.181236695 | 0.010209283 |
| SLC2A10 | 0.916550755 | 0.011977362 |
| IRF1-AS1 | 1.32156939 | 0.000767305 |
| SIPA1L1 | -0.885974129 | 0.008208721 |
| SSPOP | 1.755638502 | 0.026331243 |
| ELANE | 2.698939918 | 9.40E-07 |

| | | |
|---|---|---|
| ZNF624 | 0.884380406 | 0.020265204 |
| BCO2 | 1.61742301 | 0.011679442 |
| KCNMB2 | 3.765176839 | 0.001633862 |
| ENTPD6 | -0.689497242 | 0.029006934 |
| ENPP1 | -2.596399879 | 3.08E-09 |
| SERPINB2 | -2.142479714 | 0.001734485 |
| CFD | 3.06431788 | 4.84E-08 |
| MAP1LC3C | 3.663048369 | 1.19E-07 |
| KLHDC1 | 1.402710951 | 5.84E-05 |
| ATAD3A | -1.466425713 | 2.90E-06 |
| ZNF181 | 0.931386454 | 0.000578304 |
| GPAA1 | -0.751723336 | 0.017468329 |
| MYO1C | -0.86725868 | 0.001244169 |
| H2BC12 | 0.749677975 | 0.033239044 |
| TEAD4 | -0.76762611 | 0.009701808 |
| ERO1A | 0.697559006 | 0.016118093 |
| ZNF311 | 1.134149583 | 0.005919921 |
| S100A6 | -0.989482886 | 0.037861245 |
| MPZL1 | 1.208707797 | 7.13E-08 |
| MBP | 2.035036674 | 0.00176808 |
| C1orf122 | -0.634849956 | 0.048301679 |
| ZNF667 | 1.095766584 | 0.000817462 |
| ZBTB14 | 0.69057174 | 0.021186596 |
| ZNF248 | 0.77701519 | 0.006770687 |
| TOR4A | -1.194150666 | 0.009232884 |
| ZNF544 | -0.665704345 | 0.020701603 |
| TMEM229B | 2.420292986 | 2.10E-05 |
| NPIPB6 | 2.984476221 | 0.015894615 |
| FKBP1C | -1.468558816 | 0.018435165 |
| TMEM116 | 0.990880315 | 0.013634273 |
| ZKSCAN8 | 0.762860302 | 0.035226505 |
| HYLS1 | -1.016103836 | 0.035650753 |
| GET3 | -0.717775587 | 0.042537305 |
| SPRED2 | 0.980443283 | 0.002105293 |
| NTRK1 | 2.120023687 | 0.018841138 |
| ZNF69 | -1.65752439 | 5.63E-07 |
| ZNF568 | 0.683206728 | 0.02038065 |
| TPM2 | -1.22386224 | 0.000327751 |
| SH3BGRL2 | 2.01927965 | 5.10E-09 |
| ARC | -1.978821235 | 0.041004343 |
| PPIAP22 | -1.055943689 | 0.01197909 |
| CCDC69 | 1.973380199 | 0.000324569 |

| | | |
|---|---|---|
| CALM1 | -0.687786411 | 0.006352593 |
| TAFA2 | 1.39365201 | 0.015696229 |
| ANKRD13B | 0.733457932 | 0.018999755 |
| SMOC1 | 5.197828669 | 0.004805944 |
| MSRB1 | -0.969198335 | 0.011566351 |
| SMURF1 | -0.622056329 | 0.035140294 |
| SLC5A3 | 0.933382511 | 0.033614104 |
| EGFL6 | 3.615143312 | 1.12E-06 |
| APCDD1L | -2.545607015 | 0.000351931 |
| RASSF9 | 1.405760265 | 0.00556831 |
| GRIN3A | -2.957345563 | 0.000319431 |
| ZNF521 | 1.140785089 | 0.023283004 |
| PNP | -1.822408001 | 0.008843414 |
| ARHGAP11A | -2.540716849 | 2.74E-06 |
| HMGN2 | -0.944902297 | 0.007940677 |
| FICD | 0.679578023 | 0.045621123 |
| ITPRIPL1 | -1.895398827 | 0.015299291 |
| RASGEF1A | 2.197658233 | 2.92E-05 |
| APRT | -0.724507144 | 0.039933166 |
| GPRASP1 | 0.799871348 | 0.006851595 |
| ZFP2 | 1.308074997 | 0.004675577 |
| KIFBP | -0.490606502 | 0.039213933 |
| ARMCX6 | -0.523745939 | 0.039967472 |
| TDRKH-AS1 | -2.063241815 | 0.045039734 |
| INF2 | -0.894555557 | 0.00382604 |
| IQANK1 | 2.261860498 | 0.003508756 |
| EFCAB2 | 1.111034321 | 0.003188299 |
| STUM | 3.11672652 | 0.014501775 |
| SERTAD4-AS1 | 0.900202991 | 0.023089706 |
| LINC02901 | -3.324015078 | 0.03903283 |
| RAET1G | -1.29579075 | 0.023095911 |
| SAMD5 | 3.059635953 | 0.014733531 |
| CENPW | -2.353422863 | 0.000512933 |
| PLPP4 | -1.47788196 | 2.04E-07 |
| SNHG5 | 1.232971015 | 0.034583327 |
| PCMTD2 | 0.904232932 | 0.001961515 |
| ZYG11A | -2.772416453 | 0.000915327 |
| LINC01270 | 0.98697392 | 0.022678071 |
| LINC00963 | 0.667851116 | 0.011556556 |
| MAFB | 1.039333088 | 0.046771814 |
| C2orf72 | 3.495189311 | 0.025482785 |
| NHSL2 | -0.840616892 | 0.024848189 |

| | | |
|---|---|---|
| ASAH2B | 0.854014696 | 0.010478977 |
| AGAP9 | -0.870235774 | 0.037166698 |
| SYT15 | -3.07360641 | 0.000489432 |
| ZDBF2 | 0.985059631 | 0.012227434 |
| HNRNPCP2 | -1.115344364 | 0.024176243 |
| HLA-DMA | -1.876659362 | 8.45E-05 |
| PSMB8-AS1 | 1.289038613 | 0.032005225 |
| SPIN3 | 0.730152856 | 0.011585451 |
| COL15A1 | 2.902930436 | 3.97E-06 |
| NOTCH4 | -1.463707953 | 0.016045679 |
| PJVK | 1.438726611 | 0.001113294 |
| ERICH2 | 2.074115824 | 5.06E-07 |
| HSPA1A | -0.812278644 | 0.016511515 |
| HSPA1L | 0.898418576 | 0.006691994 |
| VARS1 | -1.029167196 | 0.000495362 |
| BAG6 | -0.519237213 | 0.030943816 |
| MICA | -0.984289738 | 0.003423447 |
| DHX16 | -0.504467847 | 0.036197582 |
| ACOXL-AS1 | 3.382107441 | 5.75E-06 |
| POLR1HASP | 0.883401697 | 0.031487265 |
| HLA-F | 1.718575272 | 0.010779782 |
| C5orf60 | 5.465536827 | 0.00636574 |
| RPL26L1-AS1 | -1.508669133 | 0.003815168 |
| INSYN2B | -1.124485798 | 0.009072688 |
| ODAD4 | 1.287463184 | 0.003470241 |
| DCTN1 | -0.703019634 | 0.004911254 |
| TCTN1 | 0.790545449 | 0.020770859 |
| ATP6V0E2-AS1 | 0.943471943 | 0.026548089 |
| PSG5 | -1.532051275 | 0.004684962 |
| SPIRE2 | 1.239503843 | 0.018097101 |
| LINC01121 | 6.03968535 | 0.000218065 |
| TMEM240 | 0.640553451 | 0.040622261 |
| CFAP96 | 1.702946425 | 0.037596028 |
| TRIQK | 0.692978358 | 0.022778271 |
| TMEM170B | 1.831337687 | 0.000661465 |
| MUC12 | -2.048992306 | 0.020682511 |
| ADGRG1 | -2.709963861 | 9.67E-05 |
| TECPR1 | -0.681551347 | 0.029680452 |
| MT1A | -2.182290275 | 0.02090919 |
| INSYN1 | 0.864175682 | 0.029195724 |
| MT1M | -1.482825798 | 0.014740914 |
| KRT81 | -4.506902532 | 4.28E-08 |

| | | |
|---|---|---|
| ATP6AP1L | 1.495927264 | 0.000208798 |
| CCDC85C | -1.896100281 | 0.000728466 |
| TMSB4X | -0.79022946 | 0.047013304 |
| SMIM11 | 1.522090853 | 0.000308188 |
| C21orf62-AS1 | 1.697856626 | 0.004397323 |
| HSP90AB2P | -2.79939701 | 0.036041243 |
| NYNRIN | -1.065211858 | 0.015022717 |
| DOK6 | 1.265374859 | 0.000322421 |
| JPT2 | -1.379658772 | 7.66E-05 |
| GOLGA8O | 2.809762751 | 0.032100722 |
| ATP10A | -0.581283808 | 0.04815694 |
| COL6A6 | -2.275017409 | 0.003901623 |
| CFAP44 | 0.919019129 | 0.043508346 |
| VGLL3 | 0.987868093 | 0.003952179 |
| SLC48A1 | 1.123788382 | 0.000422976 |
| SNORD17 | -2.13440062 | 0.021365964 |
| LINC01089 | 0.90418054 | 0.030594337 |
| RNF208 | 0.915025526 | 0.033913226 |
| CGB8 | -5.525788918 | 0.004959453 |
| CRIP1 | -2.266457631 | 0.006484272 |
| TRIM59 | -1.300725992 | 7.09E-06 |
| MLLT11 | -1.538551196 | 0.001038723 |
| ASIC3 | 1.568266225 | 0.004028068 |
| NRAS | -0.688558113 | 0.019944301 |
| HNRNPA3P6 | -1.447239814 | 0.042937813 |
| LTC4S | -3.071288352 | 2.75E-08 |
| MXD3 | -2.006610379 | 0.000677007 |
| HAUS7 | 1.553411371 | 0.002299493 |
| ANXA2P1 | -1.528962683 | 0.003575284 |
| RPLP1P6 | 1.513690597 | 0.013890764 |
| DNAJC9 | -1.175074615 | 0.006137336 |
| LEPROT | 0.69598737 | 0.016354306 |
| GPSM3 | -1.107143512 | 0.002313879 |
| NCKIPSD | 0.716392253 | 0.01989074 |
| ATF6B | -0.803900066 | 0.00703327 |
| S1PR3 | -1.722148865 | 1.71E-05 |
| RPL17P50 | 1.536866644 | 7.19E-05 |
| HMGB1P10 | -1.324152661 | 0.015560566 |
| CLIC1 | -1.544431766 | 1.23E-05 |
| ACTBP2 | -2.02695736 | 1.96E-05 |
| LINC01521 | 0.975241932 | 0.043892838 |
| LTB4R | 1.514834291 | 0.000181906 |

| | | |
|---|---|---|
| LTB4R2 | 1.282525554 | 0.020932295 |
| MRPL23 | -0.989994309 | 0.014218186 |
| CPNE1 | -0.780221249 | 0.014236008 |
| PAXIP1-AS2 | 0.918163096 | 0.011478838 |
| GCC2-AS1 | 1.701739701 | 0.006343657 |
| SMIM30 | -0.802082607 | 0.007068335 |
| POU2AF3 | 3.28673064 | 0.000175563 |
| NEURL1B | -4.294778203 | 0.001658397 |
| TUBAP2 | -2.920442058 | 2.09E-05 |
| NUTM2D | 1.36314694 | 0.000543404 |
| EML6 | 1.499333205 | 0.015661293 |
| C10orf105 | 5.932871438 | 0.018002232 |
| ARHGEF33 | 1.377656014 | 0.006202823 |
| DDX12P | -1.739478005 | 0.014532367 |
| LINC02981 | 1.67733287 | 0.039887346 |
| LINC00243 | 3.706773568 | 0.013603119 |
| ALOX12-AS1 | 0.845079706 | 0.046767923 |
| UBXN2B | 0.73139955 | 0.045803604 |
| BASP1-AS1 | 2.956834252 | 2.27E-05 |
| UBE2QL1 | 2.059790649 | 0.019758068 |
| LINC02449 | 1.42920909 | 0.012281906 |
| LINC02982 | 1.674115076 | 0.049657657 |
| DHRS4-AS1 | 1.094446226 | 8.47E-05 |
| FAM66B | 0.833519875 | 0.013201865 |
| MYL5 | 0.926650665 | 0.004622331 |
| MIR99AHG | 1.34547408 | 0.035649799 |
| LINC01139 | 6.235046512 | 0.00073275 |
| TTC34 | 1.795267426 | 0.020880545 |
| SP9 | -1.936674821 | 0.03185823 |
| FTH1P8 | 0.895789516 | 0.048549954 |
| HNRNPA1P12 | -3.389045603 | 0.044636727 |
| RPS18P9 | 1.076939894 | 0.002825565 |
| KRTAP1-5 | -3.07453539 | 0.013919176 |
| PLXNA4 | 3.220686269 | 4.65E-17 |
| HMSD | -4.423632968 | 0.000527719 |
| SLC12A8 | -0.874143503 | 0.008732557 |
| APOL6 | 1.255954035 | 0.000449966 |
| TTC4P1 | 5.332989998 | 0.018825802 |
| EXOC3-AS1 | -0.819435165 | 0.029138956 |
| FAM185A | 0.880785943 | 0.003289767 |
| LINC01615 | 1.617464039 | 0.026311927 |
| CSNK1A1P1 | -3.27260029 | 0.000434071 |

| | | |
|---|---|---|
| TMSB4XP4 | -1.31023053 | 0.024521152 |
| BCL10-AS1 | 2.299297047 | 0.034046875 |
| LINC01422 | 1.230981243 | 0.037981878 |
| CCDC18-AS1 | 1.190482136 | 0.012861917 |
| PRR15-DT | 2.999720039 | 0.001058091 |
| EEF1A1P1 | -1.804540416 | 0.017368578 |
| MIR181A2HG | 1.542417607 | 0.047008488 |
| EPB41L4A-AS1 | 0.947520921 | 0.000735669 |
| SLC44A3-AS1 | 1.744830955 | 0.002165123 |
| PPM1F-AS1 | -1.14207895 | 0.003499463 |
| BAZ2B-AS1 | 1.022313598 | 0.005861146 |
| DNAJC27-AS1 | 1.137621021 | 0.045702679 |
| PITX1-AS1 | -1.863262196 | 0.047102858 |
| HAGLR | -1.336262166 | 0.001235965 |
| HNRNPA1L3 | -1.117350073 | 0.007469732 |
| ZMIZ1-AS1 | 1.645800467 | 0.008484257 |
| RTCA-AS1 | 1.133906985 | 0.019291849 |
| TUBB8P6 | 4.843797324 | 0.018987822 |
| IMPDH1P8 | -5.483466757 | 0.002851483 |
| LINC00240 | 2.626403447 | 0.002572194 |
| EIPR1-IT1 | 4.145090631 | 0.024101554 |
| LINC00863 | 0.707479727 | 0.032433284 |
| TMEM233 | -6.892708405 | 5.68E-06 |
| RPS26P58 | -2.002931884 | 0.046732722 |
| RANP4 | 1.991154452 | 0.030492125 |
| SLC9A3-AS1 | 1.212129839 | 0.001657425 |
| PDK1-AS1 | 1.795370629 | 0.000374265 |
| MIR137HG | -1.463748662 | 0.001109568 |
| LINC00705 | -4.817827825 | 0.018098227 |
| SFTA1P | 3.310076232 | 0.000399947 |
| RFPL1S | 2.388851835 | 0.000427369 |
| JPX | 0.780701599 | 0.008231214 |
| NUTM2B-AS1 | 0.630687955 | 0.031330357 |
| GBP1P1 | 6.001343056 | 0.000663825 |
| MTND2P28 | 0.775611564 | 0.042423906 |
| MEG8 | -1.358492731 | 0.000605101 |
| TMEM30A-DT | 1.234074238 | 0.03140633 |
| TSBP1-AS1 | -2.844637654 | 0.002272983 |
| S1PR1-DT | -1.95523405 | 0.012445222 |
| FAM88F | 5.096025735 | 0.010517667 |
| FGF13-AS1 | 1.878451015 | 0.039125561 |
| LINC00623 | 1.651608069 | 0.00012604 |

| | | |
|---|---|---|
| TEX22 | 1.171167636 | 0.01238401 |
| SGMS1-AS1 | 1.044442607 | 0.001316037 |
| ZKSCAN8P1 | 1.3932403 | 0.032390745 |
| CROCC2 | 2.721902276 | 0.021322044 |
| THBS2-AS1 | 1.01150815 | 0.015700381 |
| LINC02542 | 1.973177517 | 0.027872394 |
| LINC00323 | 2.7646651 | 0.025307256 |
| UPK1A-AS1 | 1.775494931 | 0.002891685 |
| PAPPA-AS2 | -4.490278537 | 0.038540144 |
| FAM66C | 1.460935932 | 5.13E-06 |
| LINC02554 | 5.265482068 | 0.013224595 |
| PPP1CB-DT | 1.824337329 | 0.008031542 |
| LINC01359 | 3.138199666 | 0.014370158 |
| DANCR | -1.188883122 | 0.000632013 |
| COX6A1P2 | -1.168157334 | 0.019697151 |
| LINC00511 | -2.980002685 | 1.90E-06 |
| MUC12-AS1 | -5.886450121 | 1.44E-06 |
| WDR46 | -0.598816673 | 0.021882701 |
| LINC00629 | 1.825965765 | 0.016950201 |
| RPS28P7 | -1.002421324 | 0.020416714 |
| LMCD1-AS1 | 1.30650084 | 0.036788479 |
| WDR35-DT | 4.271605378 | 0.003933481 |
| COPS8-DT | 1.489973043 | 0.000650666 |
| KLLN | 1.425724351 | 0.00415085 |
| RBM26-AS1 | 1.111438866 | 0.035183099 |
| TP73-AS1 | 0.735687959 | 0.012511546 |
| KIF9-AS1 | 1.082328445 | 0.003967539 |
| LTB | -4.726995011 | 0.033428738 |
| LINC03013 | 5.797721306 | 0.000333363 |
| LINC01341 | 1.339794674 | 0.029284795 |
| MELTF-AS1 | 0.717778619 | 0.033657873 |
| LINC00578 | 1.178671304 | 0.002435465 |
| MT-ATP8 | 0.930046971 | 0.003504307 |
| PCAT6 | 0.958712705 | 0.031633868 |
| RRAGC-DT | 1.043140287 | 0.024370166 |
| SDAD1P1 | 1.278814144 | 0.006209292 |
| MIR34AHG | 1.206432366 | 0.012335974 |
| LYPLAL1-AS1 | 5.439422268 | 0.00026662 |
| SNHG26 | 1.177950414 | 0.001549085 |
| PROB1 | -1.151945638 | 0.000252558 |
| DHFR | -1.218854215 | 0.013393549 |
| WEE2-AS1 | 0.809209417 | 0.044970421 |

| | | |
|---|---|---|
| PCMTD1-DT | 1.083926463 | 0.002645862 |
| ORMDL1P1 | 4.997465784 | 0.014033831 |
| LINC01165 | 4.07392497 | 0.001392105 |
| EIF4A1P10 | -1.092354831 | 0.0286437 |
| RUNX3-AS1 | 5.186662234 | 0.012112669 |
| LINC00452 | 1.695378732 | 0.041368474 |
| KCNQ1-AS1 | 4.399509593 | 0.043626587 |
| ACTG1P11 | -2.843893606 | 0.035373172 |
| PATL2 | 2.370025982 | 0.018958088 |
| LINC01204 | -2.330815633 | 0.000615846 |
| PDE4DIPP1 | 5.373567067 | 0.006891002 |
| KHDC1-AS1 | 0.989873627 | 0.002784516 |
| ATXN1-AS1 | 1.054682393 | 0.024034765 |
| ALMS1-IT1 | -4.070729239 | 0.0007654 |
| TMSB4XP6 | -1.535605012 | 0.001579613 |
| CNOT6LP1 | 2.038333449 | 0.005375027 |
| HMGN2P3 | -1.480806214 | 0.033642583 |
| ACTG1P23 | -2.421481484 | 0.022970402 |
| EMC1-AS1 | 1.22117924 | 0.010361231 |
| PSMG3-AS1 | 1.156753712 | 0.000530241 |
| MKX-AS1 | 2.269461274 | 0.001644916 |
| KLHL7-DT | 2.630943361 | 0.026841175 |
| FBXW4P1 | -2.391568745 | 0.011764766 |
| RPL10P3 | -1.695800176 | 0.046859115 |
| LINC02541 | 1.678206895 | 0.019186402 |
| CDC20P1 | 1.699163733 | 0.020965011 |
| MIR4422HG | 3.355128993 | 0.043041449 |
| HAR1B | 2.709339875 | 0.000632957 |
| KLF3-AS1 | 1.54727373 | 0.000161126 |
| YBX1P2 | -1.762691336 | 0.017038585 |
| SYT15-AS1 | 2.307262389 | 0.002245461 |
| LINC02884 | 1.82341611 | 0.005688647 |
| MANCR | -5.349445567 | 8.33E-05 |
| IFT122P3 | 2.03207827 | 0.006532515 |
| MAP4K3-DT | 1.261890326 | 0.00015674 |
| CLIC1P1 | -2.091811612 | 0.048781646 |
| WARS2-AS1 | 0.867127508 | 0.003141388 |
| LINC01750 | 1.809023493 | 0.002990628 |
| FAM27C | 1.404870988 | 0.036446361 |
| FAM225A | -2.970618063 | 0.00124462 |
| LINC02015 | 2.825831 | 0.001886742 |
| DIRC3 | 3.109399366 | 2.21E-05 |

| | | |
|---|---|---|
| LINC00899 | 1.207349471 | 0.007405385 |
| PRH1 | 2.403410607 | 0.034036657 |
| TRAF3IP2-AS1 | 1.014798084 | 0.000631007 |
| DARS1-AS1 | 1.507385162 | 5.99E-05 |
| ANXA2P2 | -1.585934208 | 1.95E-07 |
| CAP1P2 | -2.014234382 | 0.014615239 |
| TMEM253 | 2.655874437 | 1.37E-06 |
| LINC01031 | 1.713392614 | 0.033889116 |
| NPM1P13 | 4.093708129 | 0.008596864 |
| FAM131B-AS2 | -0.709355582 | 0.037159128 |
| LINC01705 | -5.13446818 | 2.94E-09 |
| STEAP1B-AS1 | 6.078629279 | 0.000150489 |
| ITGA6-AS1 | 2.970163461 | 0.000320041 |
| COL4A2-AS1 | -3.497159525 | 0.02819921 |
| RPS2P32 | -1.388569275 | 0.04179511 |
| LYRM9 | 1.00811528 | 0.017412016 |
| SEC1P | 2.928645039 | 0.013790397 |
| LINC00702 | -1.320267092 | 0.000545142 |
| MPDU1-AS1 | 0.889996018 | 0.0100575 |
| GPX1 | -0.775291755 | 0.023777021 |
| RASA4DP | -3.683967156 | 0.000518155 |
| PFN1P1 | -1.430519736 | 0.016623252 |
| CNIH3-AS2 | -3.158682218 | 0.002321705 |
| LINC00841 | 1.833943951 | 0.008510623 |
| LINC01719 | 0.867137183 | 0.047632736 |
| TMEM238 | -1.650974519 | 0.040808295 |
| LINC00460 | -4.813685527 | 0.017991391 |
| PANTR1 | -2.80812152 | 0.042685508 |
| H2BC15 | 2.138675995 | 0.000489394 |
| SHROOM3-AS1 | 1.34362702 | 0.048633075 |
| LINC01503 | 1.157641156 | 0.001182587 |
| RPL10P9 | 1.75221543 | 0.027177046 |
| UBE2SP1 | -2.192592107 | 0.001330173 |
| ZNF879 | 0.795010869 | 0.012128592 |
| ERICH2-DT | 1.975448977 | 0.009963668 |
| LINC01426 | 1.629767515 | 0.006438307 |
| ZNF37BP | 0.9551389 | 0.006511693 |
| PINLYP | -1.14404982 | 0.044878824 |
| LNCTAM34A | 1.138767126 | 0.009323976 |
| SNRK-AS1 | -1.560791254 | 0.026088678 |
| VDAC1P3 | -3.928281595 | 0.04906405 |
| NPIPB2 | 2.092728655 | 0.02079592 |

| | | |
|---|---|---|
| GAS5 | 0.732244509 | 0.016649286 |
| EIF5AP4 | -2.293444323 | 0.019995176 |
| AARSD1P1 | 3.948077848 | 0.033193732 |
| LINC02966 | -3.607255745 | 0.035143923 |
| ID2-AS1 | 1.111829027 | 0.002089123 |
| ZSCAN31 | 1.226437285 | 0.012813984 |
| C12orf75 | -1.812092873 | 0.000167522 |
| HGH1 | -0.644667325 | 0.031816018 |
| SSR4P1 | 0.728392488 | 0.041273023 |
| RPS2P7 | -2.092447661 | 0.048332101 |
| L3MBTL2-AS1 | 2.290746865 | 0.000391789 |
| AGAP1-IT1 | -3.354743813 | 0.0141983 |
| MSC-AS1 | 0.994996526 | 0.000972147 |
| EPM2A-DT | 1.811715901 | 1.38E-08 |
| KIAA0040 | 1.418070613 | 0.039468832 |
| OLMALINC | 1.159661759 | 0.005691864 |
| ARHGEF7-AS2 | 3.351155388 | 0.011147296 |
| TTC28-AS1 | 0.850211901 | 0.013394655 |
| LINC01936 | 2.597280416 | 6.64E-06 |
| COX10-DT | 0.919172526 | 0.010217637 |
| GUSBP5 | -1.744848055 | 0.012264406 |
| LINC03099 | -1.479795333 | 0.013673461 |
| MFF-DT | 1.169847506 | 0.001299872 |
| PKMP1 | -1.908191766 | 0.037025621 |
| RNF217-AS1 | 1.225877614 | 0.003065782 |
| ZNF853 | 2.969100623 | 8.27E-07 |
| INTS6-AS1 | 1.19517359 | 0.0343145 |
| ZBTB40-IT1 | 3.20132607 | 0.046945466 |
| FTH1P11 | 1.03938443 | 0.037055601 |
| TTN-AS1 | 0.691721182 | 0.014400277 |
| LINC01522 | -4.836806681 | 0.049219275 |
| UNC5B-AS1 | 2.707194756 | 0.017796255 |
| SHISA9 | 4.078463698 | 7.06E-15 |
| LINC00857 | -1.439823259 | 0.000240131 |
| KIFC1 | -5.305762261 | 2.67E-14 |
| CDK6-AS1 | -2.000375345 | 0.014555004 |
| LINC00649 | 1.803002213 | 0.039550077 |
| LINC02918 | 2.232628475 | 0.036268212 |
| LINC01679 | 1.10546786 | 0.025927351 |
| ERRFI1-DT | 5.456440671 | 0.003188224 |
| TXNDC5 | -1.21654641 | 0.000543018 |
| OR1J4 | 2.357311284 | 0.001994127 |

| | | |
|---|---|---|
| NME1 | -0.984719552 | 0.00695648 |
| ACAD11 | 1.171407757 | 0.028153539 |
| RPL13P5 | -1.165989196 | 0.011998536 |
| L1TD1 | -6.10517923 | 3.00E-06 |
| AQP1 | 3.909333108 | 1.01E-08 |
| PNMA2 | 1.9524156 | 2.83E-09 |
| LINC02066 | 5.862416014 | 0.000915737 |
| CD302 | 1.794462771 | 7.47E-08 |
| PTPRG-AS1 | -1.31798267 | 0.023330003 |
| ARPC4 | -1.28289839 | 0.000858504 |
| INMT | 3.927199876 | 0.005432399 |
| ADAMTS9-AS2 | 2.332433471 | 2.62E-08 |
| ARPC1A | -0.606354385 | 0.030085042 |
| CTDNEP1P1 | -1.586335722 | 0.044715098 |
| PWP2 | 2.805781802 | 0.000580651 |
| SNHG3 | -0.87027976 | 0.004400754 |
| LASTR | -5.698304663 | 0.000379169 |
| C22orf39 | 0.636978127 | 0.023718867 |
| PEG10 | -1.543926838 | 3.93E-05 |
| PCDHGC4 | -1.839289883 | 0.014461242 |
| HLA-DMB | -4.807285701 | 0.001274107 |
| AP5Z1 | -0.628567691 | 0.018953541 |
| PCDHAC2 | 3.24879894 | 6.89E-07 |
| TICAM2 | -1.15972245 | 0.01132683 |
| C4orf48 | -0.845459405 | 0.027315298 |
| CFAP57 | 2.555606952 | 0.000504718 |
| RPLP0P2 | 2.354491056 | 0.01758544 |
| GSTA1 | -4.27361906 | 0.031372168 |
| TMEM141 | -0.986315659 | 0.008683355 |
| TMEM225B | 2.372619753 | 0.003705474 |
| GATA2-AS1 | -1.389520414 | 0.017895626 |
| SCARF2 | -1.11523029 | 0.000128412 |
| KCNK15-AS1 | 1.874786581 | 0.010949949 |
| PTCHD4 | 1.164892956 | 0.045612078 |
| C4A | 1.937646233 | 7.88E-05 |
| MSANTD2-AS1 | 1.574726573 | 0.002952247 |
| NEAT1 | 1.053253324 | 0.01411412 |
| BDNF-AS | 0.870480907 | 0.016984337 |
| ZNF585B | 0.727310895 | 0.010454025 |
| LINC02202 | 1.716806146 | 3.06E-06 |
| CEBPA | 2.454453594 | 0.000346026 |
| SNHG6 | 0.677589246 | 0.019262684 |

| | | |
|---|---|---|
| NUDT16L2P | -1.643693425 | 0.020049245 |
| LINC01550 | 4.230629705 | 2.93E-05 |
| UBR5-DT | 1.437439375 | 0.035498542 |
| SBF2-AS1 | 1.324157531 | 0.000119494 |
| EXTL3-AS1 | 1.995016501 | 0.003784558 |
| LINC00968 | 2.481500901 | 6.28E-06 |
| CIBAR1-DT | 3.274455249 | 0.015896902 |
| RASSF8-AS1 | 0.677759204 | 0.035361244 |
| H2AJ | -0.98053165 | 0.008708185 |
| UBAP1L | 1.330436111 | 0.015877944 |
| BRPF3-AS1 | 1.601652192 | 0.004635709 |
| SOCS2-AS1 | -1.030751851 | 0.011075357 |
| MIR210HG | 1.259756133 | 0.010554378 |
| LINC01252 | 2.247770226 | 0.006369141 |
| FBXO38-DT | 2.249146233 | 0.000271502 |
| CKMT2-AS1 | 1.075587144 | 0.005153653 |
| TWF2 | -0.763964357 | 0.013759782 |
| SEC24B-AS1 | 0.9727154 | 0.027879007 |
| LINC00926 | 1.225433802 | 0.025692717 |
| NNT-AS1 | 0.897296603 | 0.004511663 |
| ADH1C | 2.455039207 | 0.016703379 |
| LUCAT1 | 2.779936497 | 1.52E-07 |
| APELA | 4.109227882 | 0.001670143 |
| LETR1 | 5.621066798 | 0.017239883 |
| LINC02485 | 4.801436403 | 0.007796801 |
| SRP14-DT | 0.826254743 | 0.046707665 |
| LINC01085 | -5.156253391 | 1.26E-05 |
| COPB2-DT | 1.750578548 | 7.51E-05 |
| FAM151B-DT | 0.624043983 | 0.030392594 |
| ZNF436-AS1 | 1.025727692 | 0.027922588 |
| TICAM2-AS1 | 1.216265498 | 0.026983646 |
| LINC02057 | -4.719809885 | 0.02180046 |
| LINC02223 | 2.481434597 | 0.036677169 |
| TRGV7 | -3.902611374 | 0.007104813 |
| GJD2-DT | -3.543642165 | 1.36E-05 |
| LINC01018 | 2.763410922 | 0.030794988 |
| YJEFN3 | 1.601851997 | 0.01950098 |
| AGA-DT | 1.968891974 | 0.031322231 |
| ATP6V1E2 | 0.667180744 | 0.020321124 |
| SELENOP | 2.630824225 | 0.02305422 |
| GMDS-DT | 1.234465268 | 0.001533132 |
| RMEL3 | 3.593501942 | 0.016598882 |

| | | |
|---|---|---|
| LINC02600 | -2.963142375 | 0.003460707 |
| RIPK2-DT | 0.699921405 | 0.02433094 |
| MRPS30-DT | 2.256428907 | 0.00029214 |
| TMEM92-AS1 | 2.075301398 | 0.012291443 |
| ZNF345 | 1.053319798 | 0.030498691 |
| LINC02384 | 2.159216427 | 0.000771174 |
| HSPD1P11 | 2.069418374 | 0.026760789 |
| LINC02615 | 1.246643788 | 0.038421882 |
| LINC01094 | 1.458071534 | 0.002689032 |
| MTA1-DT | 0.90581443 | 0.004217539 |
| LINC01605 | -2.181610921 | 0.001962925 |
| STAU2-AS1 | 5.587196791 | 0.00112974 |
| PCDHGB6 | 1.384649478 | 0.002190607 |
| MAILR | 1.073075618 | 0.010559147 |
| TRNP1 | -1.740092411 | 9.16E-13 |
| EIF5AL1 | -1.543058198 | 0.013335275 |
| OTUD6B-AS1 | 0.620280553 | 0.035992054 |
| LNCOC1 | 2.752612432 | 0.000106348 |
| KBTBD11-AS1 | 3.177212173 | 0.000158759 |
| PCDHGA10 | 1.569437645 | 4.08E-05 |
| PCDHGB2 | 1.580337136 | 4.12E-05 |
| CLDN23 | 2.262531464 | 0.011406808 |
| ZNF260 | 0.732918188 | 0.037737559 |
| LYN | 0.988927772 | 0.027241678 |
| BHLHE22-AS1 | 7.369139734 | 8.45E-07 |
| PCDHGB7 | 1.007782042 | 0.001410312 |
| LINC03018 | 5.869955483 | 0.001065252 |
| PCDHGB1 | 1.313261353 | 0.002320164 |
| TPM4P3 | -1.023084152 | 0.004375622 |
| C4orf46P3 | 3.536296927 | 0.035161716 |
| MIR130AHG | 1.599803287 | 3.82E-05 |
| CAPN1-AS1 | -1.081246682 | 0.009147546 |
| FPGT | 0.674377623 | 0.045907441 |
| TMEM9B-AS1 | 1.144613959 | 0.015526823 |
| DPP3 | -0.995591312 | 0.001274517 |
| PGAM1P8 | -1.293539408 | 0.010654115 |
| TMEM123-DT | 2.951239852 | 0.039385536 |
| POLR2M | 0.696455207 | 0.021566887 |
| TRIL | 2.296363382 | 0.048084976 |
| PRECSIT | -1.793121518 | 0.000658309 |
| MRPL40P1 | -4.602433863 | 0.030269544 |
| PAPPA-AS1 | 2.259102902 | 0.010761346 |

| | | |
|---|---|---|
| CTSO | 1.63962394 | 0.000187606 |
| SMIM3 | 1.409760781 | 2.25E-05 |
| SUPT16HP1 | 1.200718662 | 0.041817337 |
| RPL41P5 | -1.38645537 | 0.025115605 |
| LINC02985 | 2.121898138 | 0.040703911 |
| CLEC12B | 2.529830959 | 0.044648684 |
| PLBD1-AS1 | 3.759601535 | 0.000118457 |
| LCIIAR | -2.360212706 | 0.017220694 |
| PPP1R14B-AS1 | -2.162205463 | 1.90E-07 |
| TMPO-AS1 | -1.86719539 | 0.009966604 |
| LNCOG | -1.538572826 | 0.024892388 |
| LINC02298 | 1.249883353 | 0.018084509 |
| C12orf75-AS1 | -2.941069361 | 8.95E-05 |
| INAFM1 | -0.758151825 | 0.009025499 |
| PRANCR | 1.516957843 | 0.000363323 |
| TUBA1B-AS1 | -2.598607423 | 5.65E-05 |
| LINC00640 | 3.879420434 | 2.80E-11 |
| DIO3OS | 4.611938887 | 9.70E-05 |
| LINC01397 | 2.320447337 | 0.029756901 |
| LINC00638 | 1.124976075 | 0.011028516 |
| AP1G2-AS1 | 1.882160333 | 0.000518611 |
| FOXN3-AS1 | -1.001549077 | 0.034937613 |
| LINC02279 | 2.641615496 | 0.002115343 |
| FPGT-TNNI3K | 1.825543589 | 0.049094227 |
| MIDEAS-AS1 | 2.602807369 | 4.29E-06 |
| LINC00517 | -3.036049226 | 0.007328888 |
| SIPA1L1-AS1 | -1.224498491 | 0.00338112 |
| ZNF710-AS1 | 1.289774934 | 0.00634376 |
| KIF23-AS1 | -1.530912369 | 0.000102419 |
| POU2F2-AS2 | 3.179413355 | 0.009658139 |
| ARNT2-DT | 2.821630512 | 0.002003311 |
| IQCH-AS1 | 1.952832503 | 1.69E-11 |
| GREM1-AS1 | -1.539990119 | 9.63E-06 |
| HMGB1P6 | -1.284782651 | 9.49E-05 |
| SLC22A31 | 2.903111247 | 0.022681125 |
| ZNRD2-DT | 1.5691915 | 0.035979001 |
| SNHG19 | -1.046344333 | 0.042491992 |
| SCX | -1.442245944 | 0.009077825 |
| LMF1-AS1 | -1.325543698 | 0.011019782 |
| ATP2A1-AS1 | -1.46417696 | 0.027207464 |
| INSYN1-AS1 | 3.500726312 | 0.03284275 |
| FAM157C | -4.613324263 | 0.003747148 |

| | | |
|---|---|---|
| ARHGAP23P1 | -2.926387954 | 0.030251054 |
| CCPG1 | 1.511153649 | 8.48E-06 |
| LINC01572 | -1.411028179 | 0.012106915 |
| LINC02129 | -4.805343462 | 0.019317217 |
| ZNNT1 | 1.858508653 | 0.000129385 |
| LINC02473 | 3.372233001 | 0.003549096 |
| LINC03100 | 2.930252506 | 0.036752196 |
| BOP1 | -1.224461922 | 0.000105789 |
| ATP2C2-AS1 | -2.082493282 | 0.000235342 |
| MANEA-DT | 2.762585227 | 0.018440705 |
| PAN3-AS1 | 1.432700734 | 0.001153375 |
| TBILA | -1.665871146 | 0.044040314 |
| LINC01686 | 2.088175866 | 0.012159153 |
| EEF1A1P38 | -1.1796652 | 0.011913263 |
| LINC00165 | 4.416296873 | 0.01119843 |
| FOXF2-DT | -2.427568906 | 0.015576185 |
| LINC00922 | 2.641856961 | 0.029514805 |
| ZFPM1-AS1 | 1.710254195 | 0.022803393 |
| LINC00662 | 0.824108465 | 0.00429344 |
| MMP25-AS1 | 1.093576241 | 0.008089317 |
| DLGAP1-AS2 | 1.171883507 | 0.016813671 |
| LINC02861 | 1.328727267 | 0.004856424 |
| MMP12 | -5.958333249 | 0.000772074 |
| LINC01569 | 1.283710716 | 0.013342326 |
| PCDHGA4 | 1.358873641 | 0.015375083 |
| MRPL12 | -0.936049816 | 0.033543633 |
| CCNQ | -0.796021719 | 0.010591789 |
| AKAP1-DT | 1.167285023 | 0.048187083 |
| TAPT1-AS1 | 1.7860846 | 0.000369678 |
| RN7SL689P | 1.286452305 | 0.005149396 |
| IKBKE | -1.13924495 | 0.000502053 |
| SNORD3A | -2.477452046 | 0.029454248 |
| SNRPGP2 | -1.151800125 | 0.016602408 |
| MYH4 | 3.251065189 | 0.045057994 |
| TTC39C-AS1 | 3.463002438 | 0.003265178 |
| ANXA8 | -1.803654601 | 0.037517307 |
| CLASP1-AS1 | 1.820508502 | 0.033567182 |
| RNF115 | -0.72723422 | 0.005711061 |
| MIR4664 | 2.155809808 | 0.016418711 |
| RARA-AS1 | -0.824859694 | 0.028192075 |
| FSBP | 0.972509879 | 0.019285725 |
| TXNIP | 2.386273664 | 1.25E-11 |

| | | |
|---|---|---|
| LINC00683 | 1.953290884 | 0.010162751 |
| LINC01910 | -4.890870238 | 0.002874974 |
| LINC01443 | 2.271048676 | 0.006020463 |
| MYO15B | 1.636840974 | 0.000177426 |
| SOCS3-DT | 2.824014281 | 0.015788739 |
| LINC01532 | 5.445310691 | 0.002361793 |
| ILF3-DT | -0.811624264 | 0.016196623 |
| ZNF566-AS1 | 1.000079749 | 0.016006598 |
| SNHG30 | -0.91521278 | 0.018044176 |
| LINC01415 | -1.379299751 | 0.004480212 |
| LINC00906 | 3.417127229 | 0.001876754 |
| KDSR-DT | 1.587860933 | 0.002850223 |
| SETBP1-DT | 2.408193978 | 1.53E-05 |
| ZNF582-DT | 1.185038861 | 0.021674562 |
| BRCA1P1 | 3.895903703 | 0.003026123 |
| RPLP0P12 | 2.61598866 | 0.000419387 |
| ARHGEF18-AS1 | 3.947339956 | 0.002755284 |
| MZF1-AS1 | 0.923717993 | 0.008523076 |
| LINC00664 | -2.96455783 | 5.00E-06 |
| LINC01081 | 3.021285825 | 0.015581588 |
| MAN1B1-DT | -0.918583894 | 0.035902606 |
| TRABD2B | 3.319126478 | 0.000101043 |
| IKBKG | -0.602151463 | 0.035024855 |
| RAB11B-AS1 | 0.920951209 | 0.0069351 |
| C10orf95-AS1 | 1.110521209 | 0.017667155 |
| KCNQ1OT1 | -1.07456977 | 0.027482001 |
| SNHG8 | 0.783910821 | 0.044194243 |
| LEISA1 | -2.625682396 | 0.000632698 |
| RASL10B | 1.577404737 | 0.001228532 |
| CTAGE15 | -4.342369077 | 0.041579274 |
| MMP28 | 4.94833325 | 0.015169919 |
| MILR1 | -1.775950962 | 0.003981307 |
| ATP2B1-AS1 | 1.355917987 | 0.000209552 |
| TAGAP-AS1 | 1.4932861 | 0.004019832 |
| ANKRD34A | -1.797071887 | 4.69E-05 |
| CASC15 | 2.346719921 | 5.54E-05 |
| FAM193B-DT | 4.182377611 | 0.000216282 |
| CPVL-AS2 | 2.318531608 | 0.037051262 |
| ZNF595 | 0.955539992 | 0.002989013 |
| CHASERR | 0.665235581 | 0.01162902 |
| MYHAS | 1.387936622 | 0.008510976 |
| LINC02035 | 1.553970628 | 1.28E-05 |

| | | |
|---|---|---|
| PACERR | 2.383915497 | 0.02180582 |
| H3-7 | -1.429732492 | 0.007435631 |
| DGCR11 | -1.016600622 | 0.009382943 |
| LINC01138 | 1.43679294 | 0.001183474 |
| ZNF658 | 0.845477388 | 0.018738956 |
| ZSCAN2-AS1 | 1.088916722 | 0.023492047 |
| LINC02340 | 4.399509593 | 0.043626587 |
| XKR5 | -2.653023937 | 0.044508275 |
| ARHGAP23 | -1.025683991 | 0.005693026 |
| UHRF1 | -3.133362759 | 9.72E-09 |
| PIK3R6 | 4.218783015 | 0.044804963 |
| RMRP | -2.05110773 | 0.02603304 |
| TYW1B | 2.330660642 | 0.000566129 |
| PIGW | -0.8970737 | 0.00755852 |
| RAB35-AS1 | -0.801871416 | 0.02313365 |
| CEBPB-AS1 | 1.687799914 | 0.000476182 |
| SYT15B | -3.21607443 | 0.042560287 |
| PSMB3 | -0.675884041 | 0.040392368 |
| SRD5A2 | 3.117411172 | 0.022904984 |
| CISD3 | -0.715994932 | 0.030786051 |
| LINC02139 | 2.808487895 | 0.029973815 |
| MYO19 | -0.848309921 | 0.010371019 |
| CHMP1B2P | 3.328756118 | 0.010654824 |
| TMEM191B | -3.448091529 | 0.041239974 |
| MRM1 | -0.799030618 | 0.015807954 |
| GOLGA6L10 | 1.84504982 | 0.001865143 |
| OR2S2 | -2.572825268 | 0.046862314 |
| CCDC163 | 2.114280565 | 9.46E-07 |
| LINC01943 | 1.379763462 | 0.016002304 |
| LINC01232 | 0.925867768 | 0.032241777 |
| EIF1B-AS1 | 1.071681479 | 0.003987164 |
| LINC01127 | 7.297309547 | 1.33E-07 |
| SNHG4 | -0.881275665 | 0.036463679 |
| FRG1DP | 1.442955188 | 0.011363906 |
| FRG1EP | 1.625039618 | 0.000936898 |
| FRG1GP | 1.655750931 | 0.000775913 |
| IQCJ-SCHIP1 | -1.336442625 | 0.005292646 |
| VSIG10L2 | 3.299004877 | 0.030047048 |
| SCO2 | -0.663989241 | 0.019943397 |
| LINC02795 | 1.968672126 | 0.025840763 |
| HULC | 1.472806331 | 0.0003232 |
| TCF12-DT | 0.90518209 | 0.034662694 |

| | | |
|---|---|---|
| LINC02988 | -5.010113155 | 0.021996456 |
| MITA1 | 2.382687168 | 3.44E-05 |
| KCNQ5-DT | -2.969770151 | 0.030493172 |
| COPG2IT1 | 2.370470757 | 0.011540338 |
| ACAD9-DT | 2.154333126 | 0.020358423 |
| ACTL10 | -1.46519831 | 0.001938349 |
| PANO1 | -1.186769794 | 0.007129292 |
| RPSA2 | -2.166737694 | 0.001038323 |
| LINC01512 | 3.395619151 | 0.041931335 |
| GOLGA2P5 | 2.433162996 | 0.01404294 |
| POM121L9P | 3.393806004 | 0.019443407 |
| RRN3P3 | -0.760878544 | 0.031704073 |
| CASTOR3P | 0.802079906 | 0.017847894 |
| AMZ2P1 | 1.661057897 | 0.000327945 |
| CASP4LP | 4.617030824 | 0.005728689 |
| SOD2 | 1.627271889 | 1.71E-06 |
