## Supplementary Table 9 for "Biallelic variants in *ARHGAP19* cause a motor-predominant neuropathy with asymmetry and conduction slowing"

**Supplementary Table 9.** Complete list of clustered enriched pathways including both up-regulated and down-regulated genes.

| ID | Term_Description | Fold_Enrichr | occurrence | support | lowest_p | highest_p | Up_regulated | Down_regulated | Cluster | Status |
|---|---|---|---|---|---|---|---|---|---|---|
| hsa04110 | Cell cycle | 4.50906916 | 10 | 0.35555914 | 7.65E-29 | 1.24E-28 | MDM2, SKP2, TGFB2, TFDP2 | MCM2, MCM4, MCM5, ORC6, ORC1, | 1 | Representative |
| hsa04218 | Cellular senescence | 1.87792676 | 10 | 0.09702797 | 2.85E-14 | 8.67E-10 | MDM2, TGFB2 | CCND1, CCND3, CCNE2, E2F1, MAP2 | 1 | Member |
| hsa04115 | p53 signaling pathway | 3.52852555 | 10 | 0.09413502 | 4.00E-14 | 1.99E-07 | RRM2B, CD82, ZMAT3, MDM2, SESN3, | CHEK1, CCNE2, RRM2, TP53I3, GTSE1 | 1 | Member |
| hsa04114 | Oocyte meiosis | 2.21632213 | 10 | 0.11300469 | 3.58E-11 | 4.42E-11 | ADCY2 | CDK1, MAD2L1, CCNB2, PKMYT1, AN | 1 | Member |
| hsa05203 | Viral carcinogenesis | 1.56018501 | 10 | 0.07521127 | 9.84E-11 | 4.33E-07 | CREB5, MDM2, SKP2, IL6ST, C3 | CCND1, CCND3, CCNA2, CCNE2, YW | 1 | Member |
| hsa04914 | Progesterone-mediated oocyte ma | 2.26010133 | 10 | 0.08392019 | 1.46E-10 | 1.76E-10 | ADCY2 | CCNB1, CDK1, ANAPC15, MAD2L1, BU | 1 | Member |
| hsa05169 | Epstein-Barr virus infection | 1.14274787 | 10 | 0.02599156 | 2.18E-08 | 0.0004517 | SKP2, CD40, MDM2, BCL2L11 | HLA-DMA, HLA-DMB, MAP2K3, VIM, C | 1 | Member |
| hsa05166 | Human T-cell leukemia virus 1 infe | 1.96144509 | 10 | 0.06480186 | 5.24E-07 | 2.11E-06 | IL2RB, ADCY2, CREB5, TGFB2, IL15, IL | TLN1, TSPO, TGFB1, ANAPC15, CCNB | 1 | Member |
| hsa04068 | FoxO signaling pathway | 2.28144822 | 10 | 0.09073237 | 3.08E-05 | 3.56E-05 | PRKAA2, INSR, SKP2, MDM2, BCL2L1 | CCNB1, CCNB2, CCND1, CDKN2D, PI | 1 | Member |
| hsa04814 | Motor proteins | 3.63230572 | 10 | 0.17374245 | 6.76E-15 | 1.05E-11 | DYNC2LI1, KIF27, MYO1H, MYO15B, | DYNC1I1, DCTN1, CAPZB, TUBA1B, TU | 2 | Representative |
| hsa04820 | Cytoskeleton in muscle cells | 3.27419107 | 10 | 0.12076923 | 8.45E-15 | 6.01E-11 | SGCA, ATP1A2, COL5A3, THBS2, NID2 | ITGA6, ITGA2, ITGA10, DSP, DES, VIM, | 3 | Representative |
| hsa05410 | Hypertrophic cardiomyopathy | 3.06728038 | 10 | 0.02693878 | 0.00017434 | 0.00017434 | CACNB4, PRKAA2, TGFB2, DTNA, SGC | SLC8A1, ITGA6, ITGA2, ITGA10, LMNA, | 3 | Member |
| hsa05412 | Arrhythmogenic right ventricular ca | 2.94262742 | 10 | 0.0232929 | 0.00195829 | 0.00262327 | CACNB4, LEF1, TCF7L2, GJA1, DTNA, | SLC8A1, LMNA, DSP, ITGA6, ITGA2, IT | 3 | Member |
| hsa05414 | Dilated cardiomyopathy | 2.87557536 | 10 | 0.01961538 | 0.00296627 | 0.00821797 | CACNB4, ADCY2, TGFB2, DTNA, SGC | SLC8A1, ITGA6, ITGA2, ITGA10, LMNA, | 3 | Member |
| hsa04014 | Ras signaling pathway | 1.7570223 | 10 | 0.04040816 | 1.41E-11 | 8.09E-10 | SHC2, CSF1, EFNA3, FGF9, HGF, VEG | FGF5, FGFR3, MET, RAC2, RIN1, ETS2, | 4 | Representative |
| hsa04010 | MAPK signaling pathway | 1.59601312 | 10 | 0.08852148 | 1.34E-10 | 6.19E-09 | DUSP6, MAPT, INSR, KIT, NTRK2, CAC | STMN1, ERBB3, FGFR3, MET, RPS6KA | 4 | Member |
| hsa04015 | Rap1 signaling pathway | 2.37211802 | 10 | 0.08195804 | 9.97E-09 | 4.47E-07 | CSF1, EFNA3, FGF9, HGF, VEGFA, PD | FGF5, FGFR3, MET, RAC2, MAP2K3, E | 4 | Member |
| hsa04020 | Calcium signaling pathway | 2.13850034 | 10 | 0.03000898 | 0.00015169 | 0.01735352 | ITPKB, NTRK2, P2RX7, MYLK4, PHKG1, | SLC8A1, ERBB3, FGFR3, MET, GRIN3A | 4 | Member |
| hsa04810 | Regulation of actin cytoskeleton | 2.42153714 | 10 | 0.07192308 | 7.43E-11 | 7.27E-09 | MYL5, MYLK4, INSRR, FGF9, PDGFD, | ARPC4, ARPC1B, ACTR3, ARPC2, PFN | 5 | Representative |
| hsa04530 | Tight junction | 2.40550047 | 10 | 0.08558685 | 1.33E-09 | 5.83E-08 | PARD6A, MARVELD2, PRKAA2, JAM2, | PPP2R1A, ACTN4, ACTB, ACTG1, VAS | 5 | Member |
| hsa05131 | Shigellosis | 1.96749893 | 10 | 0.03474849 | 1.81E-07 | 1.03E-05 | IL1R1, TLR4, PRKCE, CD14, MAP1LC3 | UBE2N, PYCARD, ARPC4, ARPC1B, AC | 5 | Member |
| hsa05130 | Pathogenic Escherichia coli infecti | 1.9428612 | 10 | 0.0290566 | 2.09E-07 | 1.48E-05 | LPAR4, IL1R1, TLR4, MYO1H, TNFSF1 | TUBA1B, TUBA1A, TUBA1C, TUBB4B, | 5 | Member |
| hsa05132 | Salmonella infection | 1.89796155 | 10 | 0.06205128 | 3.36E-07 | 1.79E-05 | LEF1, TCF7L2, TNFSF10, MYL5, TLR4, | MAP2K3, PYCARD, TCF7, ACTB, ACTG | 5 | Member |
| hsa04510 | Focal adhesion | 2.57776535 | 10 | 0.07270088 | 3.73E-07 | 8.05E-06 | MYLK4, MYL5, LAMC1, THBS2, TNXB, | ITGA6, ITGA2, ITGA10, FLNA, MYLK2, | 5 | Member |
| hsa05100 | Bacterial invasion of epithelial cell | 2.60224887 | 10 | 0.05135531 | 5.74E-06 | 7.10E-06 | GAB1, SHC2 | ARPC4, ARPC1B, ACTR3, ARPC2, ACT | 5 | Member |
| hsa04670 | Leukocyte transendothelial migrati | 2.15780538 | 10 | 0.03367347 | 7.68E-06 | 7.68E-06 | CDH5, JAM2, MYL5, VCAM1 | BCAR1, ACTB, ACTG1, MYL9, NCF2, R | 5 | Member |
| hsa04145 | Phagosome | 2.33315988 | 10 | 0.01955414 | 0.00016781 | 0.0193419 | C1R, OLR1, CD14, TLR4, C3, THBS2 | ITGA2, ACTB, ACTG1, NCF2, DYNC1I1, | 5 | Member |
| hsa04520 | Adherens junction | 2.5112237 | 10 | 0.03821811 | 0.0015062 | 0.00228886 | LEF1, TCF7L2, INSR, CDH5, MYL5 | TCF7, MET, RAC2, ACTB, ACTG1, VCL, | 5 | Member |
| hsa05135 | Yersinia infection | 1.51103918 | 10 | 0.01307692 | 0.00169284 | 0.01691375 | TLR4, CCL2 | ARPC4, ARPC1B, ACTR3, ARPC2, BCA | 5 | Member |
| hsa05205 | Proteoglycans in cancer | 1.98820944 | 10 | 0.1262704 | 4.35E-09 | 1.56E-07 | GAB1, ESR1, MDM2, PLAU, TLR4, VEG | ANK1, FLNA, ACTB, ACTG1, CCND1, T | 6 | Representative |
| hsa01521 | EGFR tyrosine kinase inhibitor resis | 2.07560327 | 10 | 0.07028826 | 1.73E-06 | 2.27E-06 | GAB1, SHC2, HGF, BCL2L11, VEGFA, | ERBB3, MET, FGFR3, AXL | 6 | Member |
| hsa05206 | MicroRNAs in cancer | 1.91173985 | 10 | 0.09870983 | 2.04E-06 | 5.87E-06 | VEGFA, ZEB1, PIM1, TGFB2, SPRY2, B | CDC25A, CDC25C, FSCN1, TPM1, RA | 6 | Member |
| hsa05211 | Renal cell carcinoma | 1.64039613 | 10 | 0.02615385 | 0.00116869 | 0.00144103 | TGFB2, VEGFA, EPAS1, GAB1, HGF | TGFB1, MET | 6 | Member |
| hsa04933 | AGE-RAGE signaling pathway in dia | 2.23526506 | 10 | 0.00701763 | 0.02048596 | 0.02546015 | NOX4, TGFB2, VCAM1, PIM1, VEGFA, | EDN1, THBD, CCND1, DIAPH1, PLCB3 | 6 | Member |
| hsa05212 | Pancreatic cancer | 1.66048261 | 3 | 0.00666667 | 0.0480844 | 0.0480844 | PLD1, VEGFA, TGFB2 | RAC2, E2F1, TGFB1, RAD51, CCND1 | 6 | Member |
| hsa04540 | Gap junction | 2.74877189 | 10 | 0.03696542 | 2.10E-08 | 0.00024992 | GJA1, MAP2K5, ADCY2, PDGFD | GJD2, CDK1, TUBB4B, TUBB2A, TUBB | 7 | Representative |
| hsa05016 | Huntington disease | 1.06998153 | 2 | 0.00606227 | 0.00039098 | 0.02833155 | NDUFA4L2, SOD2, CREB5, PPARG, ER | COX8A, HTT, PLCB3, AP2M1, HIP1, DC | 7 | Member |
| hsa05020 | Prion disease | 1.07389908 | 2 | 0.00891941 | 0.0021492 | 0.00295053 | PRNP, LAMC1, NDUFA4L2, CREB5 | COX8A, GRIN3A, GRIN2A, GRIN2D, TU | 7 | Member |
| hsa05012 | Parkinson disease | 1.02408619 | 4 | 0.00638984 | 0.00710006 | 0.02244382 | LRRK2, NDUFA4L2, ERN1, MAPT, NFE | COX8A, GNAS, CALM3, TUBA1B, TUBA | 7 | Member |
| hsa05034 | Alcoholism | 2.01395168 | 10 | 0.00670163 | 4.63E-08 | 0.00460286 | MAOB, CREB5, NTRK2, SHC2 | GRIN3A, GRIN2A, GRIN2D, GNAS, HD | 8 | Representative |
| hsa05032 | Morphine addiction | 1.3560608 | 9 | 0.00641026 | 3.00E-05 | 3.00E-05 | ADCY2, GABRB3 | GNG4, GNG11, GNG2, ADORA1, GNA | 8 | Member |
| hsa04713 | Circadian entrainment | 1.99776814 | 9 | 0.00641026 | 4.15E-05 | 4.15E-05 | GRIA1, ADCY2, PER2 | GRIN2A, GRIN2D, CALM3, PLCB3, GN | 8 | Member |
| hsa04726 | Serotonergic synapse | 1.16901793 | 9 | 0.00641026 | 6.34E-05 | 6.34E-05 | MAOB, GABRB3 | GNAS, PLCB3, GNG4, GNG11, GNG2 | 8 | Member |
| hsa04724 | Glutamatergic synapse | 1.83378541 | 9 | 0.00641026 | 0.00014757 | 0.00014757 | GRIA1, SLC1A3, ADCY2, SLC1A7, PLD | GRIN3A, GRIN2A, GRIN2D, GNAS, PLC | 8 | Member |
| hsa05163 | Human cytomegalovirus infection | 1.38721223 | 10 | 0.01892763 | 0.00015627 | 0.00436138 | VEGFA, CCL2, ADCY2, PTGER4, IL1R1, | GNG4, GNG11, GNG2, CALM3, PLCB3 | 8 | Member |
| hsa04728 | Dopaminergic synapse | 1.38975175 | 9 | 0.00641026 | 0.00025542 | 0.00025542 | MAOB, CREB5, GRIA1 | GNAS, PLCB3, GNG4, GNG11, GNG2, | 8 | Member |
| hsa04926 | Relaxin signaling pathway | 1.4901767 | 9 | 0.00641026 | 0.00027824 | 0.00027824 | CREB5, VEGFA, ADCY2, SHC2 | GNG4, GNG11, GNG2, EDN1, GNAS, T | 8 | Member |
| hsa04727 | GABAergic synapse | 1.37438595 | 9 | 0.00641026 | 0.00150765 | 0.00150765 | ADCY2, GABRB3, GABARAPL1 | GNG4, GNG11, GNG2, SLC38A5 | 8 | Member |
| hsa05170 | Human immunodeficiency virus 1 i | 1.31337608 | 9 | 0.00641026 | 0.00212977 | 0.00212977 | TLR4 | GNG4, GNG11, GNG2, CALM3, CDK1, | 8 | Member |
| hsa04725 | Cholinergic synapse | 1.01366671 | 9 | 0.00641026 | 0.00274727 | 0.00274727 | ADCY2, CREB5 | PLCB3, GNG4, GNG11, GNG2 | 8 | Member |
| hsa04935 | Growth hormone synthesis, secreti | 1.22208417 | 10 | 0.00653846 | 0.01024231 | 0.01389948 | ADCY2, GHR, CREB5, SHC2 | PLCB3, GNAS, BCAR1, SST, MAP2K3 | 8 | Member |
| hsa04723 | Retrograde endocannabinoid signa | 0.95273593 | 9 | 0.00641026 | 0.01093741 | 0.01093741 | GABRB3, GRIA1, ADCY2, NDUFA4L2 | PLCB3, GNG4, GNG11, GNG2 | 8 | Member |
| hsa04750 | Inflammatory mediator regulation | 2.03096664 | 6 | 0.00673469 | 0.0318108 | 0.03401746 | PTGER4, ASIC2, ASIC3, IL1R1, IL1RAP, | BDKRB1, PTGER2, PLCB3, GNAS, CAL | 8 | Member |
| hsa04062 | Chemokine signaling pathway | 1.25251921 | 9 | 0.00641026 | 0.04347252 | 0.04347252 | CXCL16, CCL2, CCL7, CCL8, CCL11, | CCL26, PLCB3, GNG4, GNG11, GNG2, | 8 | Member |
| hsa03460 | Fanconi anemia pathway | 2.96514752 | 10 | 0.03672507 | 2.19E-07 | 1.47E-05 | | FANCA, FANCB, FANCD2, FANCI, BLM | 9 | Representative |
| hsa03440 | Homologous recombination | 1.9634085 | 10 | 0.01904781 | 0.000237 | 0.01577412 | | BLM, RAD51, EME1, RAD54L, BRIP1 | 9 | Member |
| hsa04350 | TGF-beta signaling pathway | 2.32467566 | 10 | 0.01910906 | 7.56E-07 | 2.55E-05 | ACVR2A, INHBB, TGFB2, BMP4, CHRD | SMAD7, ID1, ID3, PPP2R1A, TGFB1, N | 10 | Representative |
| hsa04550 | Signaling pathways regulating pluri | 1.55274137 | 10 | 0.00653846 | 0.01819186 | 0.02506284 | WNT4, WNT3, WNT9B, FZD3, LIF, IL6S | TCF7, TBX3, FGFR3, ID1, ID3 | 10 | Member |
| hsa05219 | Bladder cancer | 3.44113173 | 10 | 0.06208078 | 1.18E-06 | 4.10E-05 | VEGFA, DAPK1, MDM2 | MMP1, RASSF1, DAPK3, E2F1, FGFR3, | 11 | Representative |
| hsa05222 | Small cell lung cancer | 1.75050878 | 10 | 0.00689786 | 4.90E-06 | 0.00064696 | FHIT, LAMC1, SKP2 | CCNE2, CCND1, ITGA6, ITGA2, RARB, | 11 | Member |
| hsa05223 | Non-small cell lung cancer | 1.54097818 | 10 | 0.00653846 | 5.98E-05 | 6.71E-05 | FHIT, HGF | CCND1, E2F1, RARB, RASSF1, MET | 11 | Member |
| hsa01522 | Endocrine resistance | 1.30599756 | 10 | 0.01277968 | 0.00030142 | 0.00495188 | ESR1, SHC2, ADCY2, JAG1, MDM2 | GNAS, CCND1, E2F1 | 11 | Member |
| hsa05207 | Chemical carcinogenesis - recepto | 1.62294511 | 10 | 0.04040816 | 0.0006282 | 0.00071746 | ESR1, ADCY2, CREB5, VEGFA, EPHX1, | CHRNA9, E2F1, CCND3, BIRC5, GNAS | 11 | Member |
| hsa05218 | Melanoma | 2.07560327 | 10 | 0.01307692 | 0.00072987 | 0.00377805 | MITF, FGF9, HGF, PDGFD, FGF17, MD | CCND1, MET, FGF5, E2F1 | 11 | Member |
| hsa05220 | Chronic myeloid leukemia | 1.21076857 | 10 | 0.01307692 | 0.00319831 | 0.00351788 | TGFB2, MDM2, SHC2 | TGFB1, E2F1, CCND1 | 11 | Member |
| hsa05214 | Glioma | 1.03780163 | 9 | 0.00666667 | 0.00377805 | 0.00412823 | SHC2, MDM2 | CALM3, CCND1, E2F1 | 11 | Member |
| hsa05162 | Measles | 1.04610405 | 5 | 0.00632911 | 0.01691375 | 0.01691375 | TLR4, IFIH1, IL2RB, MX1 | MSN, CCND1, CCND3, CCNE2, IKBKE | 11 | Member |
| hsa05160 | Hepatitis C | 0.7112207 | 5 | 0.00632911 | 0.02854306 | 0.02854306 | MX1 | IKBKE, PPP2R1A, YWHAH, YWHAZ, E2 | 11 | Member |
| hsa04917 | Prolactin signaling pathway | 0.88055896 | 2 | 0.00670584 | 0.04035625 | 0.04416952 | SHC2, ESR1 | PRLR, CCND1 | 11 | Member |
| hsa03082 | ATP-dependent chromatin remode | 1.0255922 | 10 | 0.01307692 | 1.65E-06 | 2.71E-06 | PHF10 | DPF1, ACTB, ACTG1, BCL7C, RUVBL2 | 12 | Representative |
| hsa05416 | Viral myocarditis | 2.59450408 | 10 | 0.01403526 | 0.00105748 | 0.00116664 | CD40, SGCA, DTNA | CCND1, HLA-DMA, HLA-DMB, ACTB, | 12 | Member |
| hsa03030 | DNA replication | 2.56398051 | 10 | 0.01904935 | 8.85E-06 | 1.05E-05 | | LIG1, FEN1, MCM5, MCM2, MCM4, PO | 13 | Representative |
| hsa03430 | Mismatch repair | 1.26341068 | 10 | 0.00653846 | 0.00954655 | 0.01055477 | | LIG1, EXO1 | 13 | Member |
| hsa04390 | Hippo signaling pathway | 2.22781417 | 10 | 0.06897864 | 9.53E-06 | 1.28E-05 | PARD6A, TGFB2, WNT4, WNT3, WNT9 | WWC1, RASSF1, BIRC5, YWHAH, YWH | 14 | Representative |
| hsa05225 | Hepatocellular carcinoma | 1.94339924 | 10 | 0.03248945 | 6.56E-05 | 0.00010646 | SHC2, LEF1, TCF7L2, FZD3, WNT4, W | CCND1, TCF7, E2F1, TGFB1, MET, GST | 14 | Member |
| hsa05215 | Prostate cancer | 2.07560327 | 10 | 0.02615385 | 8.98E-05 | 0.00026698 | LEF1, TCF7L2, MDM2, INSRR, CREB5, | CCND1, TCF7, E2F1, CCNE2, MMP3 | 14 | Member |
| hsa04936 | Alcoholic liver disease | 1.81615286 | 10 | 0.01346939 | 0.00648204 | 0.00894184 | TLR4, CD14, PRKAA2, NOX4, LEF1, TC | IKBKE, CPT1A, FASN, CCND1, TCF7, M | 14 | Member |
| hsa05216 | Thyroid cancer | 2.13665042 | 10 | 0.00673469 | 0.00677319 | 0.02703259 | PPARG, LEF1, TCF7L2 | CCND1, TCF7 | 14 | Member |
| hsa05226 | Gastric cancer | 2.06088268 | 10 | 0.02214286 | 0.02211549 | 0.03316842 | SHC2, LEF1, TCF7L2, FZD3, WNT4, W | CCND1, TCF7, TGFB1, MET, CCNE2, E | 14 | Member |
| hsa05224 | Breast cancer | 1.66048261 | 10 | 0.00653846 | 0.02356055 | 0.03242851 | ESR1, SHC2, JAG1, LEF1, TCF7L2, FZD | CCND1, TCF7, E2F1, FGF5 | 14 | Member |
| hsa04934 | Cushing syndrome | 1.66048261 | 10 | 0.00653846 | 0.02356055 | 0.03242851 | CREB5, ADCY2, AGTR1, FZD3, LEF1, T | PLCB3, GNAS, TCF7, CCND1, E2F1, C | 14 | Member |
| hsa05221 | Acute myeloid leukemia | 2.66369086 | 1 | 0.00571429 | 0.02475397 | 0.02475397 | RUNX1T1, LEF1, TCF7L2, PIM1, KIT, PE | CCNA2, CCND1, TCF7 | 14 | Member |
| hsa04310 | Wnt signaling pathway | 1.8275752 | 6 | 0.00689786 | 0.03856492 | 0.0478014 | FZD3, LEF1, TCF7L2, DKK2, WNT4, W | PLCB3, RAC2, CCND1, CCND3, FOSL | 14 | Member |
| hsa05167 | Kaposi sarcoma-associated herpe | 1.31337608 | 9 | 0.00641026 | 0.04642875 | 0.04642875 | IL6ST, C3, MAP1LC3C, VEGFA, CLEC2 | IKBKE, E2F1, GNG4, GNG11, GNG2, C | 14 | Member |
| hsa04060 | Cytokine-cytokine receptor interac | 2.51056424 | 10 | 0.02615385 | 1.08E-05 | 1.08E-05 | ACVR2A, IL2RB, IL6ST, IL18R1, IL1RA | TGFB1, RELT, PRLR, IL7R, IL21R, IL11, | 15 | Representative |
| hsa04630 | JAK-STAT signaling pathway | 1.9069605 | 10 | 0.01313184 | 0.00028044 | 0.003367 | IL2RB, IL15RA, IL7, IL15, TSLP, GHR, L | IL7R, IL21R, PRLR, IL13RA2, IL27, IL11 | 15 | Member |
| hsa04144 | Endocytosis | 1.38060489 | 10 | 0.01910906 | 3.45E-05 | 0.0007991 | MDM2, PARD6A, RAB11FIP1, PLD1, A | HGS, AP2M1, FGFR3, RAB11FIP5, PSD | 16 | Representative |
| hsa04666 | Fc gamma R-mediated phagocytos | 2.50503842 | 10 | 0.02539708 | 0.00096604 | 0.00418474 | PRKCE, PLD1, ASAP3 | RAC2, ARPC4, ARPC1B, ACTR3, ARPC | 16 | Member |
| hsa05418 | Fluid shear stress and atheroscler | 2.31146727 | 10 | 0.05264454 | 5.13E-05 | 0.00069729 | VCAM1, CCL2, NFE2L2, BMP4, ACVR2 | NCF2, RAC2, GSTT2B, THBD, EDN1, A | 17 | Representative |
| hsa05230 | Central carbon metabolism in can | 1.13509554 | 10 | 0.00690476 | 5.74E-05 | 7.18E-05 | KIT | PKM, FGFR3, MET, PFKP | 18 | Representative |
| hsa01230 | Biosynthesis of amino acids | 1.63709553 | 4 | 0.00670163 | 0.01108314 | 0.04491267 | ASS1 | PKM, PFKP, PRPS1, BCAT2, GOT2, NA | 18 | Member |
| hsa04210 | Apoptosis | 1.90547185 | 10 | 0.04 | 0.00011876 | 0.00135009 | TNFSF10, ERN1, BCL2L11, CTSO, CTS | BIRC5, CAPN1, CAPN2, LMNA, LMNB1 | 19 | Representative |
| hsa04141 | Protein processing in endoplasmic | 1.1941827 | 10 | 0.00689786 | 0.0295178 | 0.03908162 | MAN1A1, MAN1C1, ERN1, HERPUD1, | CAPN1, CAPN2, VCP, DERL3, TXNDC5 | 19 | Member |
| hsa05202 | Transcriptional misregulation in ca | 1.60387525 | 10 | 0.0301085 | 0.00025554 | 0.00184897 | IL2RB, PPARG, PLAU, ZEB1, RUNX1T1, | MET, MMP3, CCNA2 | 20 | Representative |
| hsa05144 | Malaria | 3.37888904 | 10 | 0.01961538 | 0.00029435 | 0.00392366 | HGF, VCAM1, TLR4, CCL2, CD40, THB | SDC1, MET, TGFB1 | 21 | Representative |
| hsa04659 | Th17 cell differentiation | 1.43853692 | 10 | 0.01299578 | 0.00070451 | 0.01108345 | IFNGR1, IL2RB, IL1R1, IL1RAP, IL6ST | HLA-DMA, HLA-DMB, IL21R, TGFB1, IL | 21 | Member |
| hsa05142 | Chagas disease | 1.34807222 | 10 | 0.00653846 | 0.00517203 | 0.00552809 | TLR4, C3, IFNGR1, TGFB2, CCL2 | PLCB3, GNAS, TGFB1, PPP2R1A | 21 | Member |
| hsa04380 | Osteoclast differentiation | 1.40605383 | 10 | 0.01317215 | 0.01032937 | 0.01803513 | CSF1, IL1R1, IFNGR1, FOSL2, MITF, P | FOSL1, ACP5, FHL2, TGFB1, NCF2 | 21 | Member |
| hsa05145 | Toxoplasmosis | 1.51346072 | 10 | 0.01307692 | 0.0252436 | 0.03361386 | LAMC1, TLR4, IFNGR1, CD40, TGFB2 | MAP2K3, HLA-DMA, HLA-DMB, ITGA6, | 21 | Member |
| hsa05321 | Inflammatory bowel disease | 2.25453458 | 2 | 0.00653846 | 0.02742937 | 0.02742937 | TGFB2, IFNGR1, MAF, TLR4, IL18R1 | TGFB1, HLA-DMA, HLA-DMB, IL21R | 21 | Member |
| hsa05146 | Amoebiasis | 1.52939188 | 6 | 0.00673469 | 0.0345532 | 0.0369475 | LAMC1, IL1R1, TLR4, CD14, TGFB2 | GNAS, PLCB3, TGFB1, VCL, ACTN4 | 21 | Member |
| hsa05140 | Leishmaniasis | 1.92298538 | 2 | 0.00653846 | 0.04411014 | 0.04411014 | C3, TLR4, TGFB2, IFNGR1 | HLA-DMA, HLA-DMB, TGFB1, EEF1A2, | 21 | Member |
| hsa04722 | Neurotrophin signaling pathway | 1.1887546 | 10 | 0.01307692 | 0.00035743 | 0.00054159 | NTRK2, SHC2, GAB1, MAP2K5, SH2B2 | ARHGDIA, ARHGDIB, MAGED1, CALM | 22 | Representative |
| hsa04962 | Vasopressin-regulated water reabs | 2.36522233 | 7 | 0.00666667 | 0.04031409 | 0.0477322 | CREB5, DYNC2LI1 | GNAS, ARHGDIA, ARHGDIB, DCTN1, D | 22 | Member |
| hsa04371 | Apelin signaling pathway | 2.34342304 | 10 | 0.01277968 | 0.00037111 | 0.03649293 | PRKAA2, MYLK4, PRKCE, ADCY2, MAP | APLNR, SPHK1, GNG4, GNG11, GNG2, | 23 | Representative |
| hsa04919 | Thyroid hormone signaling pathwa | 1.78429053 | 10 | 0.04255533 | 0.00064043 | 0.00072407 | ESR1, RCAN2, ATP1A2, MED12L, MDM | PLCB3, ATP1B3, ACTB, ACTG1, SLC16 | 23 | Member |
| hsa04971 | Gastric acid secretion | 2.76747102 | 10 | 0.01346939 | 0.0009607 | 0.0180253 | ADCY2, ATP1A2, MYLK4 | PLCB3, GNAS, ATP1B3, EZR, SST, ACT | 23 | Member |
| hsa04961 | Endocrine and other factor-regulat | 2.61526012 | 6 | 0.01261842 | 0.0051434 | 0.0391114 | PTH1R, ATP2B1, ESR1, ATP1A2 | SLC8A1, AP2M1, PLCB3, GNAS, ATP1 | 23 | Member |
| hsa04022 | cGMP-PKG signaling pathway | 1.97676502 | 1 | 0.00666667 | 0.00879267 | 0.00879267 | KCNMB2, KCNMB4, ADRA2A, AGTR1, | MYL9, ADRA1B, PLCB3, VASP, SLC8A | 23 | Member |
| hsa04261 | Adrenergic signaling in cardiomyoc | 1.95168665 | 2 | 0.00635365 | 0.04031409 | 0.0417871 | CACNB4, ATP1A2, ADCY2, CREB5, AG | SLC8A1, ACTC1, TPM1, TPM2, TPM4, | 23 | Member |
| hsa04921 | Oxytocin signaling pathway | 1.58913375 | 3 | 0.00641026 | 0.04345684 | 0.04892867 | MYLK4, ADCY2, CACNB4, PRKAA2, M | MYLK2, CALM3, GNAS, PLCB3, MYL9, | 23 | Member |
| hsa04148 | Efferocytosis | 1.31166595 | 10 | 0.00653846 | 0.00055261 | 0.00067902 | CD47, PPARG, PTGER4, XKR6 | AXL, BCAR1, TYRO3, CPT1A, PTGER2, | 24 | Representative |
| hsa04930 | Type II diabetes mellitus | 1.14704391 | 10 | 0.00653846 | 0.00062015 | 0.00077505 | INSR, PRKCE | PKM | 25 | Representative |
| hsa04066 | HIF-1 signaling pathway | 0.88954426 | 10 | 0.00653846 | 0.03165012 | 0.03898325 | VEGFA, INSR, TLR4, IFNGR1 | PFKP, EDN1 | 25 | Member |
| hsa04923 | Regulation of lipolysis in adipocyte | 1.42443361 | 1 | 0.00628931 | 0.04756745 | 0.04756745 | ADCY2, INSR, TSHR | GNAS, ADORA1 | 25 | Member |
| hsa04137 | Mitophagy - animal | 0.97949817 | 8 | 0.00653846 | 0.00207109 | 0.00314872 | GABARAPL1, MAP1LC3C, MITF | E2F1, VCP, TOMM40 | 26 | Representative |
| hsa00330 | Arginine and proline metabolism | 2.72422929 | 9 | 0.00641026 | 0.00239029 | 0.03556939 | ALDH3A2, MAOB, P4HA1 | AGMAT, SMS, CKB, ALDH4A1, P4HA3, | 27 | Representative |
| hsa04360 | Axon guidance | 2.30758245 | 9 | 0.01273885 | 0.00308034 | 0.03175924 | PARD6A, SLIT3, SLIT2, UNC5B, ROBO | RAC2, NTNG1, EPHB1, SEMA7A, MET, | 28 | Representative |
| hsa04064 | NF-kappa B signaling pathway | 1.48594325 | 10 | 0.00653846 | 0.00424918 | 0.00582772 | IL1R1, TNFSF13B, CD40, TLR4, CD14, | CARD10 | 29 | Representative |
| hsa05417 | Lipid and atherosclerosis | 1.43778768 | 10 | 0.00653846 | 0.00485205 | 0.00717487 | OLR1, CCL2, VCAM1, TLR4, CD14, ER | PLCB3, MMP1, MAP2K3, PYCARD, NC | 29 | Member |
| hsa05134 | Legionellosis | 1.74350674 | 10 | 0.00653846 | 0.0248558 | 0.03145839 | C3, TLR4, CD14 | PYCARD, VCP, EEF1A2 | 29 | Member |
| hsa04621 | NOD-like receptor signaling pathw | 1.01797103 | 1 | 0.00680272 | 0.04095608 | 0.04095608 | P2RX7, TLR4, ANTXR2, GBP2, TXNIP, G | PYCARD, IKBKE, PLCB3 | 29 | Member |
| hsa04714 | Thermogenesis | 0.88954426 | 10 | 0.00653846 | 0.00535942 | 0.00792164 | CREB5, ADCY2, PPARG, PRKAA2, ND | GNAS, ACTB, ACTG1, DPF1, CPT1A, M | 30 | Representative |
| hsa04152 | AMPK signaling pathway | 1.55670245 | 10 | 0.01418511 | 0.01829868 | 0.02408147 | PRKAA2, INSR, PPARG, LEPR, LEP, CR | CPT1A, PFKP, FASN, PPP2R1A, CCND | 30 | Member |
| hsa04211 | Longevity regulating pathway | 1.41748516 | 1 | 0.00571429 | 0.02373882 | 0.02373882 | ADCY2, PRKAA2, INSR, CREB5, SOD2, | SESN3, SESN2, PPARG | 30 | Member |
| hsa04932 | Non-alcoholic fatty liver disease | 1.22782165 | 9 | 0.00641026 | 0.03213594 | 0.03429199 | MLXIPL, PRKAA2, LEP, BCL2L11, ERN1 | TGFB1, MAP3K11, COX8A | 30 | Member |
| hsa04610 | Complement and coagulation casc | 2.48059903 | 10 | 0.00653846 | 0.00647098 | 0.00708277 | F13A1, PLAU, TFPI, C3, C4A, C1R, C1 | PROC, SERPINB2, BDKRB1, THBD, SE | 31 | Representative |
| hsa05322 | Systemic lupus erythematosus | 3.30209611 | 10 | 0.00653846 | 0.02854933 | 0.03054708 | ELANE, C4A, C3, C1S, C1R, CD40 | HLA-DMA, HLA-DMB, GRIN2A, ACTN4 | 31 | Member |
| hsa04216 | Ferroptosis | 2.542614 | 9 | 0.01265823 | 0.0097946 | 0.01267124 | GCLC, SLC40A1, FTH1, FTL, SLC39A8, | MAP1LC3C, PRNP | 32 | Representative |
| hsa04142 | Lysosome | 1.25843663 | 10 | 0.00653846 | 0.00995281 | 0.0129121 | CTSO, CTSF, GAA, IDUA, CLN5 | TCIRG1, SMPD1, PPT1, AP1B1, ACP5, | 33 | Representative |
| hsa04640 | Hematopoietic cell lineage | 2.09866552 | 8 | 0.00689786 | 0.01022797 | 0.01217003 | CD14, KIT, IL1R1, CD7, CSF1, IL7 | ITGA6, ITGA2, CD9, IL7R, HLA-DMA, H | 34 | Representative |
| hsa04120 | Ubiquitin mediated proteolysis | 0.69186776 | 10 | 0.01277968 | 0.01099812 | 0.01241098 | SKP2, MDM2 | SAE1, UBE2N, UBE2S, ANAPC15 | 35 | Representative |
| hsa04512 | ECM-receptor interaction | 2.24857021 | 8 | 0.00673469 | 0.01612172 | 0.01996694 | LAMC1, TNXB, THBS2, CD47, FREM1 | COL6A6, SPP1, ITGA6, HMMR, SDC1, | 36 | Representative |
| hsa01232 | Nucleotide metabolism | 2.13095269 | 2 | 0.00668122 | 0.01653858 | 0.04551139 | AK4, AK3, RRM2B | AK5, ENPP1, RRM2, IMPDH1, DTYMK, | 37 | Representative |
| hsa04130 | SNARE interactions in vesicular tra | 1.36211464 | 10 | 0.00653846 | 0.03199628 | 0.04018647 | STX17, VAMP4 | YKT6 | 38 | Representative |
| hsa05033 | Nicotine addiction | 2.01794762 | 8 | 0.00673469 | 0.03950566 | 0.04356643 | GABRB3, GRIA1 | GRIN3A, GRIN2A, GRIN2D | 39 | Representative |
| hsa00565 | Ether lipid metabolism | 1.86272088 | 1 | 0.00680272 | 0.04568886 | 0.04568886 | PLD1, GDPD1 | PAFAH1B3, LPCAT2, LPCAT4 | 40 | Representative |
